# Systemic treatment options for metastatic castration resistant prostate cancer: A living systematic review

**DOI:** 10.1101/2025.04.15.25325837

**Authors:** Syed Arsalan Ahmed Naqvi, Muhammad Umair Anjum, Arifa Bibi, Muhammad Ali Khan, Kaneez Zahra Rubab Khakwani, Huan He, Manal Imran, Syeda Zainab Kazmi, Ammad Raina, Ewan K. Cobran, R. Bryan Rumble, Thomas K. Oliver, Neeraj Agarwal, Yousef Zakharia, Mary-Ellen Taplin, Oliver Sartor, Parminder Singh, Jacob J. Orme, Daniel S. Childs, Rahul A. Parikh, Rohan Garje, Mohammad Hassan Murad, Alan H. Bryce, Irbaz Bin Riaz

**Affiliations:** Division of Hematology and Oncology, Department of Medicine, Mayo Clinic, Phoenix, Arizona, United States; Department of Internal Medicine, University of Oklahoma, Oklahoma City, Oklahoma, United States; Department of Internal Medicine, The University of Arizona, Tucson, Arizona, United States; Department of Biomedical Informatics and Data Science, Yale University, New Haven, Connecticut, United States; Department of Internal Medicine, Dow University of Health Sciences, Karachi, Pakistan; Department of Internal Medicine, Canyon Vista Medical Center, Midwestern University, Sierra Vista, Arizona, United States; Division of Epidemiology, Department of Quantitative Health Sciences, Mayo Clinic, Scottsdale, Arizona, United States; American Society of Clinical Oncology, Alexandria, Virginia, United States; Division of Medical Oncology, Department of Internal Medicine, Huntsman Cancer Institute (NCI-CCC), University of Utah, Salt Lake City, Utah, United States; Dana-Farber Cancer Institute, Harvard Medical School, Boston, Massachusetts, United States; Department of Oncology, Mayo Clinic, Rochester, Minnesota, United States; Division of Hematology and Oncology, University of Kansas Medical Center, Kansas City, Kansas, United States; Miami Cancer Institute, Baptist Health South Florida, Miami, Florida, United States; Evidence-Based Practice Center, Mayo Clinic, Rochester, Minnesota, United States; Department of Medical Oncology and Developmental Therapeutics, City of Hope Cancer Center, Goodyear, Arizona, United States

## Abstract

**Background:** Optimal treatment selection for metastatic castration resistant prostate cancer (mCRPC) remains challenging due to evolving standards of care in castration sensitive setting.

**Purpose:** To synthesize and appraise evidence on systemic therapy for mCRPC patients stratified by prior therapy and *HRR* alterations informing a clinical practice guideline.

**Data Sources:** MEDLINE and EMBASE (inception to 5 March 2025) using living search.

**Study Selection:** Randomized clinical trials assessing systemic therapy in mCRPC.

**Data Extraction:** Primary outcomes assessed were progression free survival (PFS) and overall survival (OS).

**Data Synthesis:** This report of the living systematic review (LSR) includes 143 trials with 17,523 patients (59 phase III/IV trials, 8,941 patients; 84 phase II, 8,582 patients). In the setting of prior androgen deprivation therapy (ADT) alone or ADT+docetaxel, treatment benefit was observed with poly (ADP-ribose) polymerase inhibitors (PARPi) in combination with androgen receptor pathway inhibitors (ARPI) for *BRCA*+ subgroup. In the setting of prior ADT+ARPI or ADT+ARPI+docetaxel, treatment benefit was observed with PARPi monotherapy for *BRCA*+ subgroup. Treatment benefit with PARPi may be observed for select non-*BRCA* homologous recombination repair (*HRR*) alterations (*CDK12*, *PALB2*). Treatment benefit was observed with abiraterone, enzalutamide, cabazitaxel, docetaxel (if no prior docetaxel), and Lu^177^ (if PSMA+) for patients without *HRR* alterations.

**Limitations:** Study-level data and indirectness in evidence.

**Conclusion:** Findings from the current LSR suggest that optimal treatment for mCRPC should be individualized based on prior therapy and *HRR* alterations. Current evidence favors PARPi alone (ARPI exposed) or in combination with ARPI (ARPI naïve) for patients with *BRCA* alterations, while ARPI alone, chemotherapy, and Lu^177^ remain potential options for patients without *HRR* alterations.

**Registration:** https://osf.io/46tjm

**Primary Funding Source:** NIH U24 grant (U24CA265879-01-1).

## Introduction

Metastatic castration resistant prostate cancer (mCRPC) is a lethal disease with a median survival of 25.6 months ^1^. It has been an area of active investigation with hundreds of trials conducted over the last decade. Several agents such as androgen receptor pathway inhibitors (ARPI), novel chemotherapeutics, poly (ADP-ribose) polymerase (PARP) inhibitors, and radiopharmaceutical therapies have been approved. While this pace of drug approvals is a blessing, it also becomes challenging to reconcile evidence from trials over the last decades for current clinical practice due to factors such as evolving standard of care in metastatic hormone sensitive prostate cancer (mHSPC), heterogeneous inclusion criteria related to prior lines of treatment, control arms that do not always reflect clinical practice, and the need to consider clinically relevant subgroups defined by homologous recombination repair (*HRR*) pathway alterations, and prostate-specific membrane antigen (PSMA) expression.

Here, we have developed a living systematic review (LSR) to support the rapidly evolving clinical practice guidelines for the management of mCRPC. The goal of this systematic review is to summarize evidence from all randomized clinical trials and present it with emphasis on the following key points (1) evidence for each drug class and drug type as a quick resource for evidence repository in mCRPC (2) evidence by receipt of previous treatment to contextualize the evidence in relevance to clinical practice (3) and evidence stratified by different alterations in the *HRR* pathway. This LSR will be continuously updated as new evidence is published, and the updates will be hosted on a companion interactive website (**<u>living website link</u>**). By facilitating ongoing updates and interactive components, this review seeks to provide a comprehensive and dynamic resource that adapts to the evolving landscape of mCRPC management.

## Methods

This LSR was conducted using the living interactive evidence (LIvE) synthesis framework ^2-4^ and is reported in accordance with the Preferred Reporting Items for Systematic reviews and Meta-Analyses (PRISMA) ^5^ (**Supplement Methods 1**). This study was registered in the Open Science Framework (https://osf.io/46tjm).

### Data Sources and Search

MEDLINE and EMBASE were comprehensively searched using a structured search strategy (**Supplement Methods 2**). Subsequently, a “living” auto search has been created with weekly updates to identify new evidence as it becomes available. The cutoff date for this report from our LSR is March 5^th^, 2025.

### Study Selection

Full-text articles of phase II, III or IV randomized clinical trials assessing systemic therapy in mCRPC were included. Trials before 1990, purely phase I trials, articles in non-English language, and non-randomized studies were excluded.

Study selection was conducted by two independent reviewers (SAAN and UA) with discrepancies resolved by the senior reviewer (IBR).

### Data Extraction and Quality Assessment

The extracted data included trial characteristics, baseline population characteristics, outcome results in the overall population and in clinically relevant subgroups. Two reviewers (SAAN and UA) independently extracted data with machine-facilitated annotations and examined risk of bias using the Cochrane Risk of Bias tool version 2 ^6^. Discrepancies were resolved by consensus and input from a third reviewer (IBR).

Patient important outcomes included progression free survival (PFS) and overall survival (OS). In instances where multiple PFS definitions were reported, radiographic PFS (rPFS) was used (**Supplement Tables 1-2)**. Time to disease progression (TTP) or composite endpoint of PFS (defined as composite of radiographic disease progression, PSA progression and/or clinical progression) were used if PFS was not reported.

### Data synthesis

The evidence was collated and stratified by (i) treatment class; (ii) treatment agent; (iii) receipt of previous treatment and (iv) *HRR* alteration status.

Eligible prior therapies included androgen deprivation therapy (ADT) with or without first-generation anti-androgens; ADT+ARPI; ADT+docetaxel; and ADT+ARPI+docetaxel. Five clinically relevant categories were defined according to the following criteria: (a) trials in which patients only received prior ADT or <25% patients received prior ARPI or docetaxel were classified into the “prior ADT” subgroup; (b) trials in which all patients received prior ADT and >75% patients received prior ARPI were classified into the “prior ADT+ARPI” subgroup; (c) trials in which all patients received prior ADT and >75% patients received prior docetaxel were classified into the “prior ADT+docetaxel” subgroup; (d) trials in which all patients received prior ADT and >75% patients received both prior ARPI and docetaxel were classified into the “prior ADT+ARPI+docetaxel” subgroup; (e) trials in which all patients received prior ADT and >25% but <75% patients received prior ARPI or docetaxel were classified into the “heterogeneous prior therapy” subgroup. Trials were included if they met the pre-specified criteria or reported survival data by prior therapy subgroup. These criteria were finalized after consensus of a panel of oncologists.

The relative effect estimates along with their 95% confidence intervals (CI) pushed from the analysis module to the *Tabulator* module are translated into intervention risk, and absolute risk differences using relative estimates and assumed baseline event risk.

The absolute risk difference per 1000 patients using relative risk (RR) is calculated as:

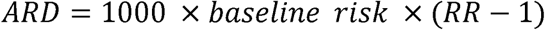

The absolute risk difference per 1000 patients using hazard ratio (HR) is calculated as:

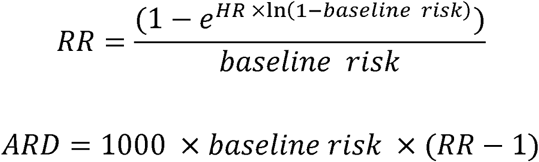

## Results

### Baseline characteristics

As of March 5^th^ 2025, 143 trials (186 references) ^7-192^ with 17,523 patients were included **(Figure 1)**. Of these, 84 (59%) were phase II randomized, and 59 (41%) were phase III/IV randomized clinical trials. A total of 8,582 and 8,941 patients were included in the 84 phase II ^99-105,107-115,117-137,139-147,149-158,160-185,190,192^, and 59 phase III/IV ^7-9,11,13,15-20,26,29-38,42,45-56,58,60,65,71-78,80,84,86,87,89-91,93-95,97,186,191^ trials, respectively. The median age ranged from 68 to 71 years (interquartile range) across the phase II trials, and 69 to 71 years (interquartile range) across the phase III/IV trials. Out of the 59 included phase III/IV trials, chemotherapy as monotherapy was assessed in 11 trials, ARPI monotherapy in seven trials, immunotherapy in four trials, the combination of two ARPIs in three trials, PARPi with ARPI in three trials, and radiopharmaceutical/radioligand monotherapy was assessed in two trials.

**Figure 1.**
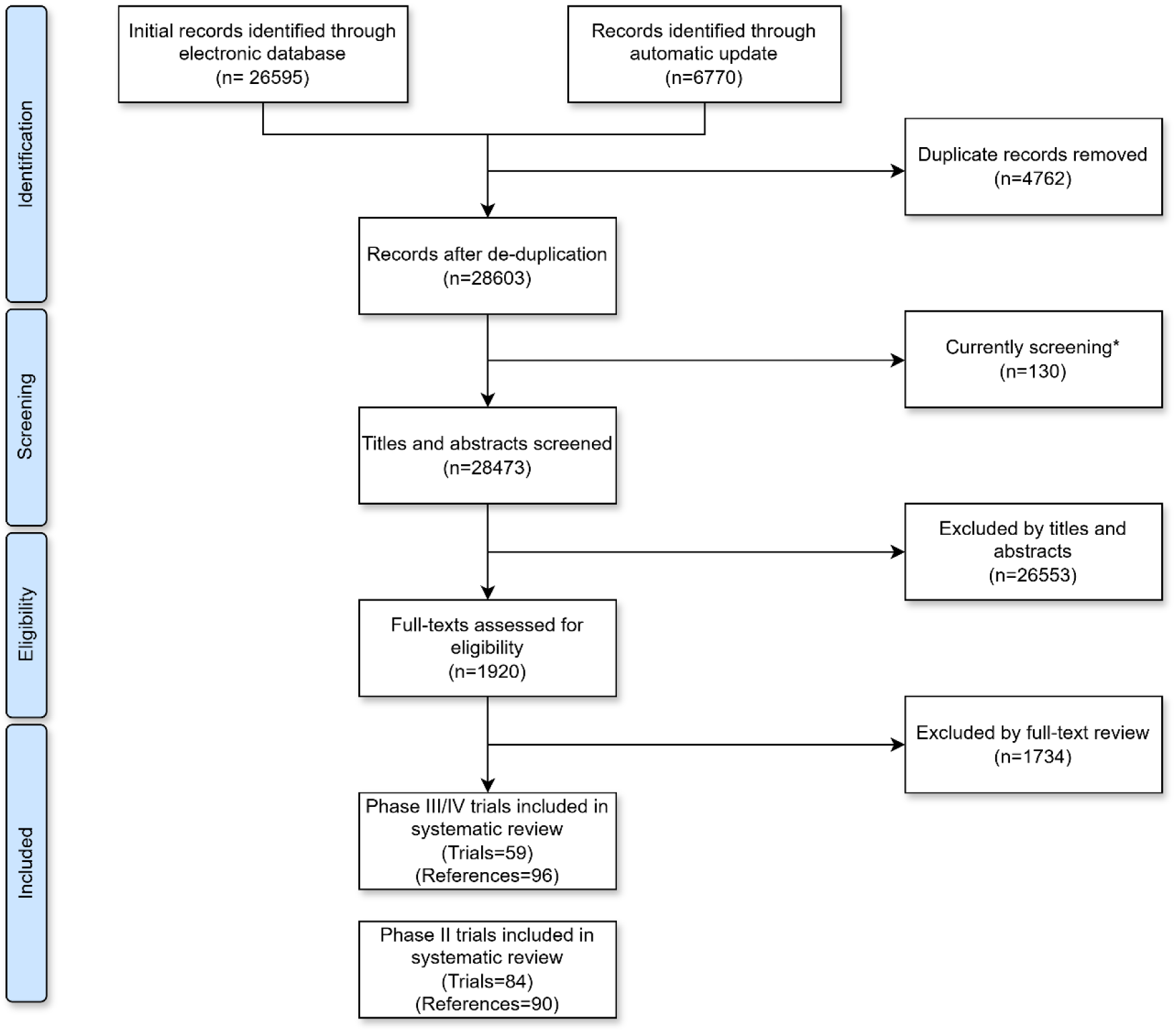
PRISMA flowchart outlining the study selection process.

In terms of risk of bias (**Supplement Figures 1-2**), some concerns for the assessment of PFS were present in most phase II and a few phase III/IV trials due to their open-label nature. However, the risk of bias across most phase II and III/IV trials assessing OS was low.

Additional characteristics and distribution by race/ethnicity are provided in **Tables 1-6**, **Supplement Tables 3-12**, and **Supplement Results**.

**Table 1.**
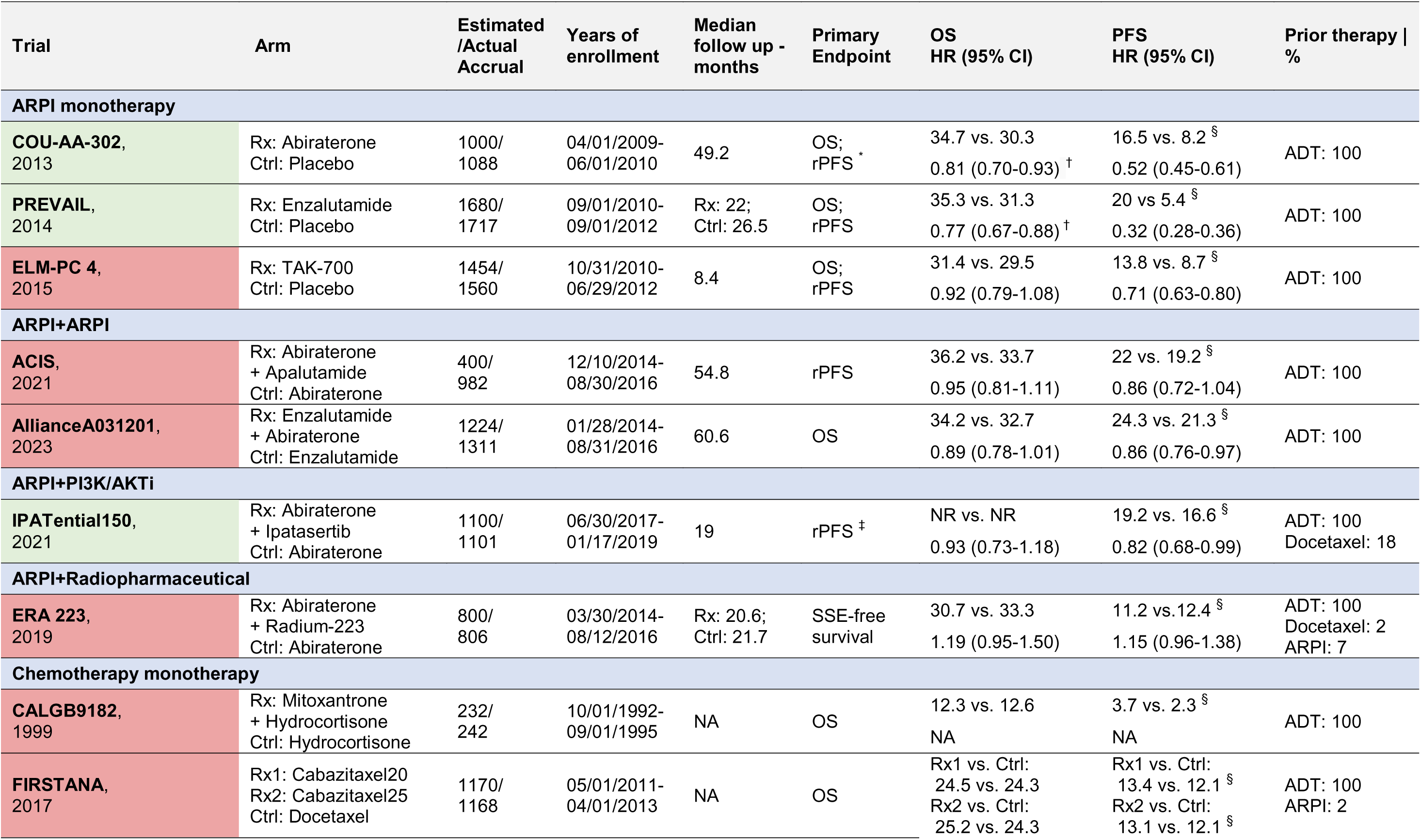
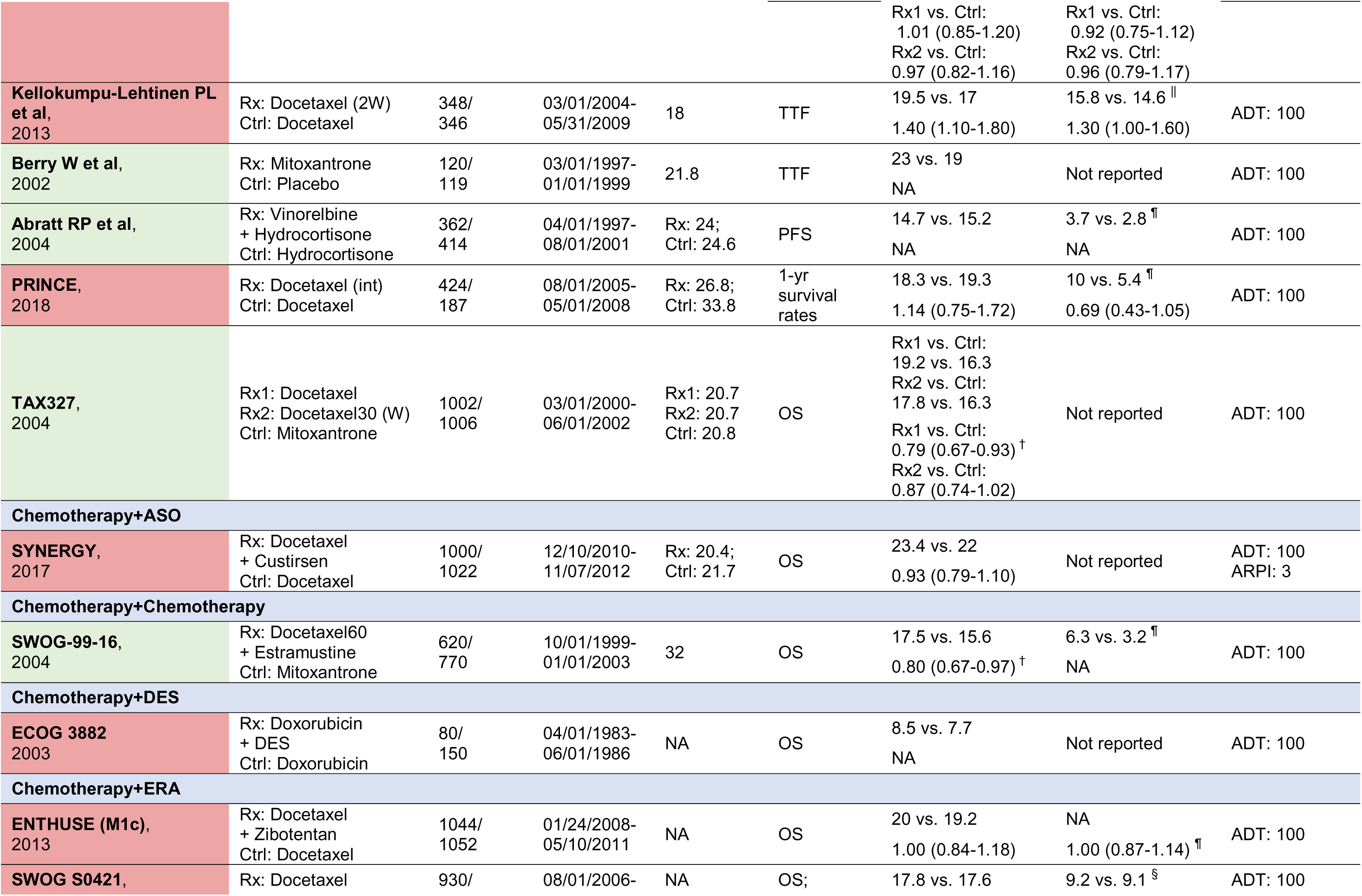
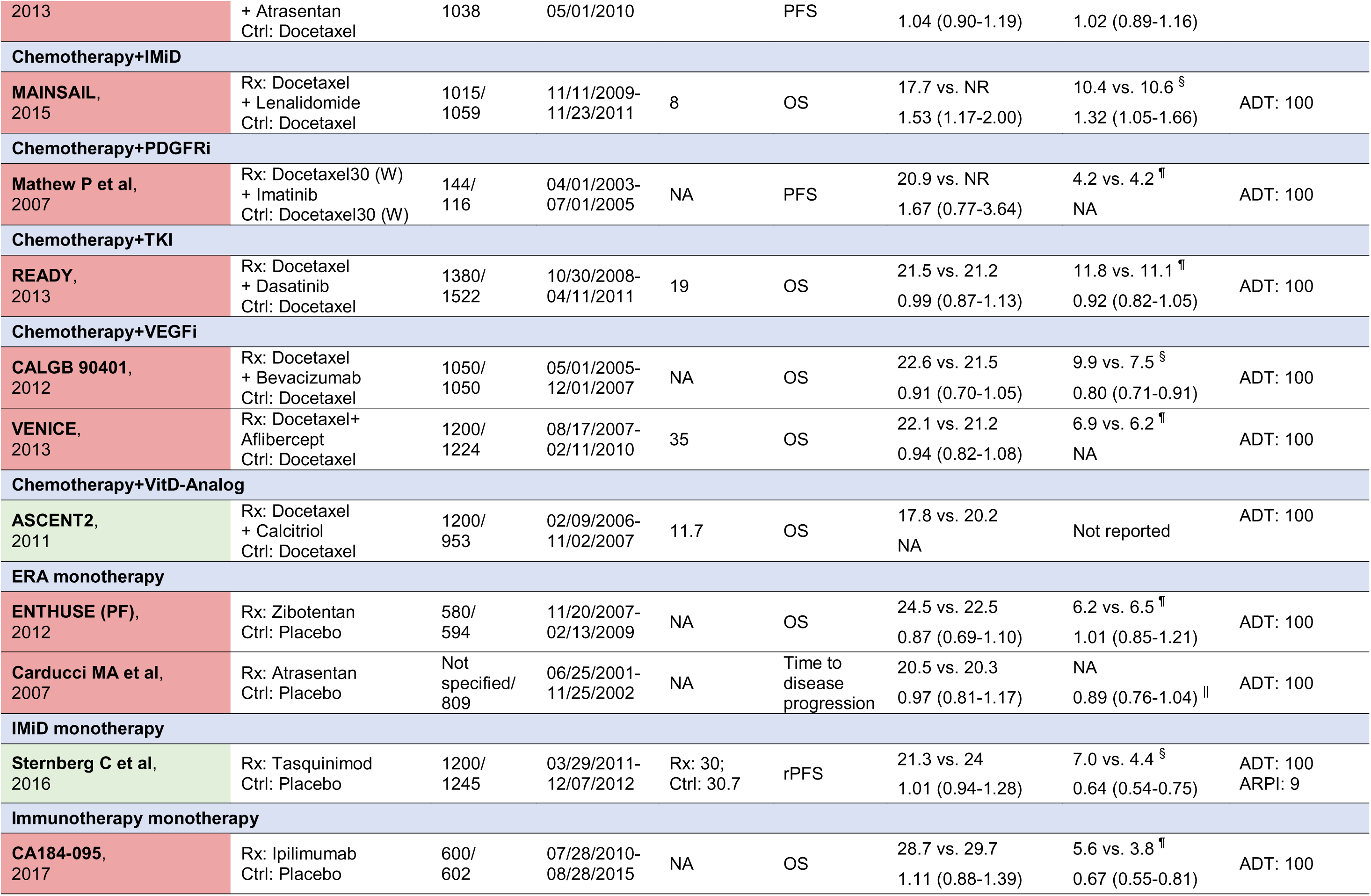
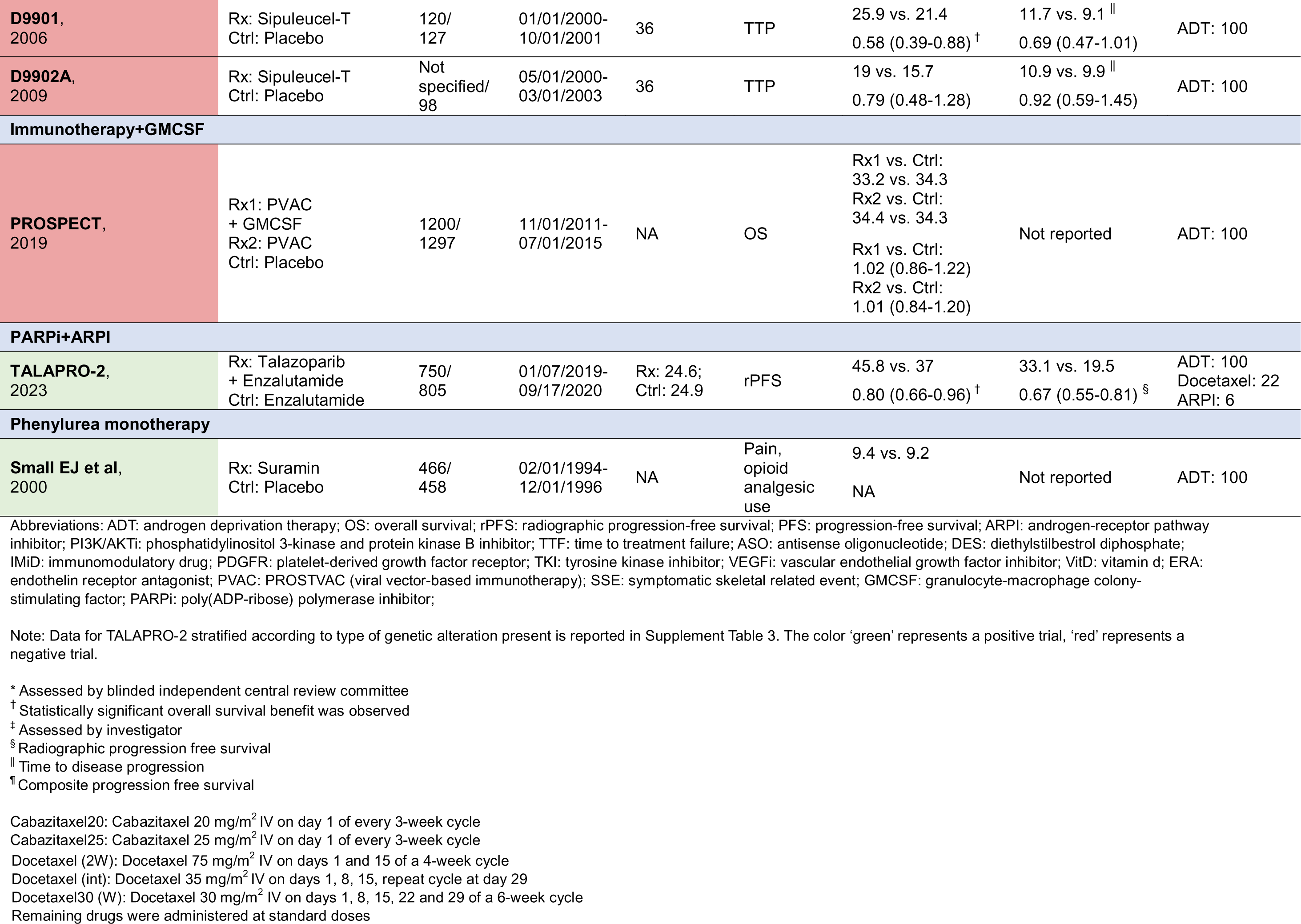
Summary of characteristics and results of included phase III trials in which patients received prior ADT only.

Heterogeneity in reporting of outcomes and subgroups in the included trials is outlined in **Supplement Results**, and **Supplement Tables 13-16**.

## Results for survival outcomes

Interactive results are available on the living website (**<u>living website link</u>**). Results can be conditionally filtered by treatment class, treatment type (combination vs. monotherapy), treatment agent, control, and prior therapy.

Here, we report the results for PFS and OS at the level of each phase III/IV trial organized by receipt of eligible prior therapy (**Tables 1-6** and **Supplement Table 17**). Additional results from phase II trials are provided in **Supplement Tables 6 and 18**.

### (1) Prior ADT with and without first-generation anti-androgens

A total of 29 phase III trials ^7,11,16-18,26,27,29,30,32-34,36,37,45-47,52-54,57,58,65,69,73,76,77,80,89,93,94,188,189^ reporting PFS and 38 ^7,11,13,15-18,28-30,32-34,36,37,45-49,51-54,57,58,69,72,73,75-78,80,85,89,93,94,188,189^ reporting OS were considered eligible for prior ADT subgroup.

#### a. Monotherapy

In terms of **ARPI monotherapy**, abiraterone acetate (COU-AA-302) was associated with a statistically significant improvement in rPFS ^27^ (0.52; 0.45-0.61) and OS ^28^ (HR: 0.81; 95% CI: 0.70-0.93) compared to placebo. Enzalutamide (PREVAIL) ^57^ was associated with a statistically significant improvement in rPFS (0.32; 0.28-0.36) and OS (0.77; 0.67-0.88) compared to placebo.

In terms of **chemotherapy monotherapy**, docetaxel with prednisone (TAX327) ^85^ was associated with a statistically significant improvement in OS (0.79; 0.67-0.93) compared to mitoxantrone with prednisone.

In terms of **single-agent immunotherapy**, sipuleucel-T was associated with a statistically significant improvement in OS compared to placebo in D9901 ^29^ (0.58; 0.39-0.88) but not in D9902A ^93^ (0.79; 0.48-1.28) and IMPACT ^36^ (0.79; 0.59-1.03).

In terms of **radiopharmaceutical monotherapy**, radium-223 (ALSYMPCA) ^13^ was associated with a statistically significant improvement in OS compared to placebo (0.69; 0.52-0.92) in docetaxel-naïve patients.

#### b. Combination therapy

In terms of **combination of two ARPIs**, enzalutamide+abiraterone (Alliance A031201) ^11^ compared to enzalutamide alone was associated with a statistically significant improvement in rPFS (0.86; 0.76-0.97) but not in OS (0.89; 0.78-1.01).

In terms of **combination of ARPI with PARPi**, olaparib+abiraterone (PROpel) ^69^ was associated with a statistically significant improvement in rPFS (0.62; 0.49-0.79) but not in OS (0.85; 0.67-1.07) compared to abiraterone alone in overall population. Talazoparib combined with enzalutamide (TALAPRO-2) ^189^ was associated with a statistically significant improvement in both rPFS (0.67; 0.55-0.81) and OS (0.80; 0.66-0.96) compared to enzalutamide alone.

In patients with *HRR* alterations, olaparib+abiraterone (PROpel) ^65,69^ was associated with a statistically significant improvement in both rPFS (0.45; 0.31-0.65) and OS (0.66; 0.45-0.95). Talazoparib combined with enzalutamide (TALAPRO-2) ^188^ was associated with a statistically significant improvement in both rPFS (0.47; 0.36-0.61) and OS (0.62; 0.48-0.81) compared to enzalutamide alone in patients with *HRR* alterations.

In patients with *BRCA* alterations, olaparib+abiraterone (PROpel) ^65,69^ was associated with a statistically significant improvement in both rPFS (0.18; 0.09-0.34) and OS (0.29; 0.14-0.56) compared to abiraterone alone. Talazoparib combined with enzalutamide (TALAPRO-2) ^81,188^ was associated with a statistically significant improvement in both rPFS (0.20; 0.11-0.36) and OS (0.50; 0.32-0.78) compared to enzalutamide alone in patients with *BRCA* alterations.

In patients with non-*BRCA* alterations, olaparib+abiraterone (PROpel) ^65,69^ was associated with a statistically significant improvement in rPFS (0.72; 0.58-0.90) but not in OS (0.91; 0.73-1.13) compared to abiraterone alone. Talazoparib combined with enzalutamide (TALAPRO-2) ^81,188^ was associated with a statistically significant improvement in rPFS (0.71; 0.52-0.96) but not in OS (0.73; 0.52-1.02) compared to enzalutamide alone.

Results for other mono-therapeutic agents and combination therapies in patients who received prior ADT are available in **Tables 1** and **6**. Summary of findings with certainty of evidence is outlined in **Supplement Tables 19** and **24**.

### (2) Prior ADT+ARPI

A total of eight ^35,42,55,60,86,97,186,191^ phase III/IV trials reporting PFS and five ^35,61,86,97,191^ reporting OS were considered eligible for prior ADT+ARPI subgroup.

#### a. Monotherapy

In terms of **PARPi monotherapy**, analysis for the overall population (cohort A+B) showed that olaparib (PROfound) ^60,61^ was associated with a statistically significant improvement in rPFS (0.49; 0.38-0.63) but not in OS (0.79; 0.61-1.03) compared to enzalutamide/abiraterone.

In patients with *BRCA* alterations, olaparib (PROfound) ^64^ was associated with a statistically significant improvement in both rPFS (0.22; 0.15-0.32) and OS (0.63; 0.42-0.95) compared to enzalutamide/abiraterone. However, subgroup analysis for patients who had received prior ARPI showed that olaparib was not associated with a statistically significant improvement in both rPFS (0.77; 0.50-1.22) and OS (1.12; 0.69-1.85) compared to enzalutamide/abiraterone. Likewise, analysis for overall population (*BRCA* and/or *ATM* alterations) showed that rucaparib (TRITON-3) ^86^ was not associated with a statistically significant improvement in OS (0.94; 0.72-1.23) when compared to enzalutamide/abiraterone/docetaxel. However, rucaparib was associated with a statistically significant improvement in rPFS when compared to enzalutamide/abiraterone/docetaxel (0.61; 0.47-0.80), enzalutamide/abiraterone (0.47; 0.34-0.66) and docetaxel (0.64; 0.46-0.88) in overall population (*BRCA* and/or *ATM* alterations). Results for patients with only *BRCA* alterations were also consistent.

In terms of **radioligand monotherapy**, ^177^Lu-PSMA-617 (PSMAfore) ^97^ was associated with a statistically significant improvement in rPFS (0.49; 0.39-0.61) but not in OS (0.98; 0.75-1.28) compared to enzalutamide/abiraterone.

#### b. Combination therapy

In terms of **addition of chemotherapy to ARPI continuation**, docetaxel+enzalutamide continuation after progression (PRESIDE) ^55^ was associated with a statistically significant improvement in cPFS (0.72; 0.53-0.96) compared to enzalutamide continuation alone.

In terms of **combination of ARPI with immunotherapy**, enzalutamide+atezolizumab (IMbassador250) ^35^ was not associated with statistically significant rPFS improvement (0.98; 0.75-1.27) but was associated with statistically significant OS harm (1.58; 1.13-2.20) compared to enzalutamide alone.

Results for other mono-therapeutic agents and combination therapies in patients who received prior ADT+ARPI are available in **Tables 2** and **6**. Summary of findings with certainty of evidence is outlined in **Supplement Tables 20 and 24.**

**Table 2.**
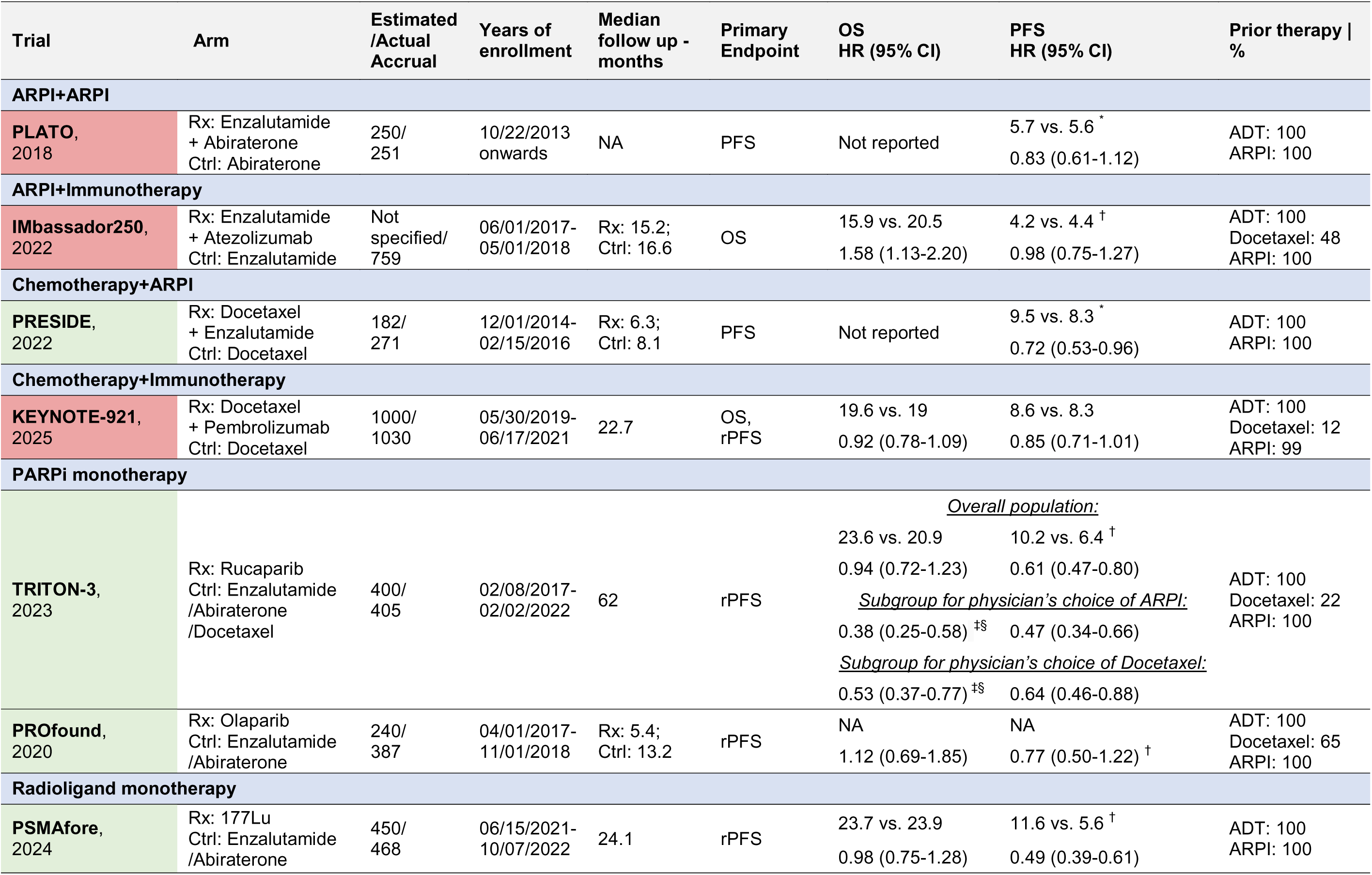
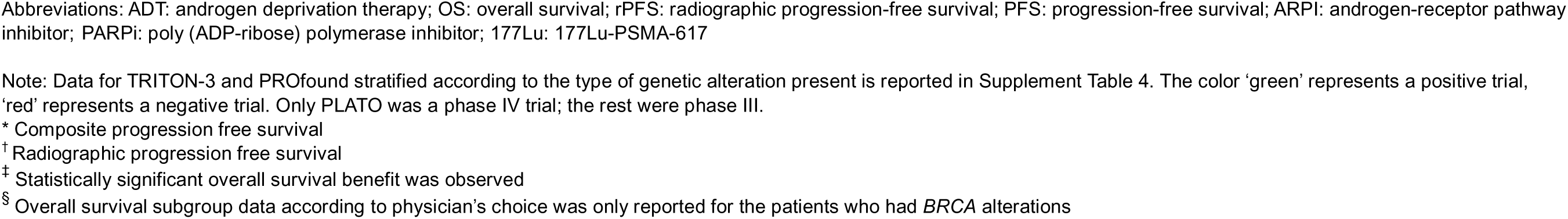
Summary of characteristics and results of included phase III trials in which patients received prior ADT and ARPI.

**Table 3.**
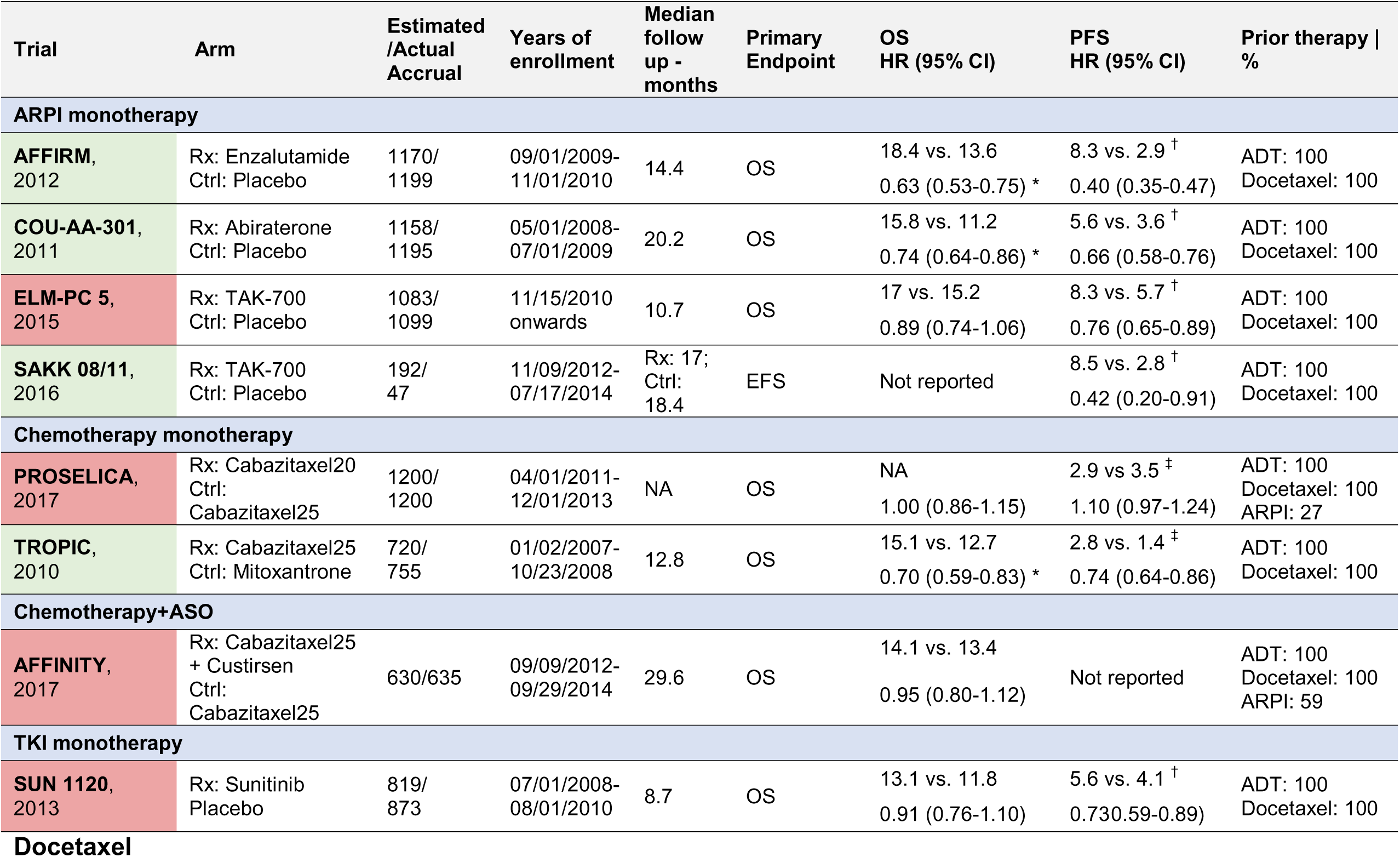
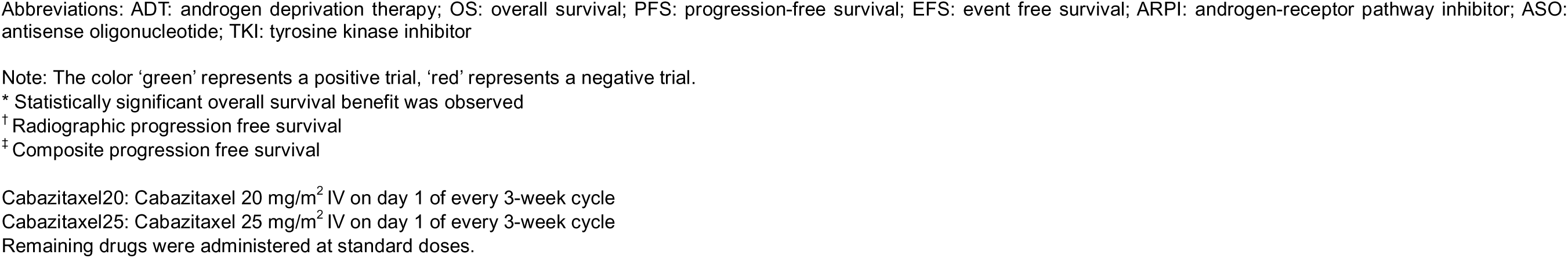
Summary of characteristics and results of included phase III trials in which patients received prior ADT and Docetaxel.

| Trial | Arm | Estimated /Actual Accrual | Years of enrollment | Median follow up - months | Primary Endpoint | OS HR (95% CI) | PFS HR (95% CI) | Prior therapy \| % |
| --- | --- | --- | --- | --- | --- | --- | --- | --- |
| **ARPI monotherapy** |  |  |  |  |  |  |  |  |
| **AFFIRM, 2012** | Rx: Enzalutamide<br>Ctrl: Placebo | 1170/<br>1199 | 09/01/2009-11/01/2010 | 14.4 | OS | 18.4 vs. 13.6<br>0.63 (0.53-0.75) * | 8.3 vs. 2.9<sup>†</sup><br>0.40 (0.35-0.47) | ADT: 100<br>Docetaxel: 100 |
| **COU-AA-301, 2011** | Rx: Abiraterone<br>Ctrl: Placebo | 1158/<br>1195 | 05/01/2008-07/01/2009 | 20.2 | OS | 15.8 vs. 11.2<br>0.74 (0.64-0.86) * | 5.6 vs. 3.6<sup>†</sup><br>0.66 (0.58-0.76) | ADT: 100<br>Docetaxel: 100 |
| **ELM-PC 5, 2015** | Rx: TAK-700<br>Ctrl: Placebo | 1083/<br>1099 | 11/15/2010<br>onwards | 10.7 | OS | 17 vs. 15.2<br>0.89 (0.74-1.06) | 8.3 vs. 5.7<sup>†</sup><br>0.76 (0.65-0.89) | ADT: 100<br>Docetaxel: 100 |
| **SAKK 08/11, 2016** | Rx: TAK-700<br>Ctrl: Placebo | 192/<br>47 | 11/09/2012-07/17/2014 | Rx: 17;<br>Ctrl: 18.4 | EFS | Not reported | 8.5 vs. 2.8<sup>†</sup><br>0.42 (0.20-0.91) | ADT: 100<br>Docetaxel: 100 |
| **Chemotherapy monotherapy** |  |  |  |  |  |  |  |  |
| **PROSELICA, 2017** | Rx: Cabazitaxel20<br>Ctrl: Cabazitaxel25 | 1200/<br>1200 | 04/01/2011-12/01/2013 | NA | OS | NA<br>1.00 (0.86-1.15) | 2.9 vs 3.5<sup>‡</sup><br>1.10 (0.97-1.24) | ADT: 100<br>Docetaxel: 100<br>ARPI: 27 |
| **TROPIC, 2010** | Rx: Cabazitaxel25<br>Ctrl: Mitoxantrone | 720/<br>755 | 01/02/2007-10/23/2008 | 12.8 | OS | 15.1 vs. 12.7<br>0.70 (0.59-0.83) * | 2.8 vs. 1.4<sup>‡</sup><br>0.74 (0.64-0.86) | ADT: 100<br>Docetaxel: 100 |
| **Chemotherapy+ASO** |  |  |  |  |  |  |  |  |
| **AFFINITY, 2017** | Rx: Cabazitaxel25<br>+ Custirsen<br>Ctrl: Cabazitaxel25 | 630/635 | 09/09/2012-09/29/2014 | 29.6 | OS | 14.1 vs. 13.4<br>0.95 (0.80-1.12) | Not reported | ADT: 100<br>Docetaxel: 100<br>ARPI: 59 |
| **TKI monotherapy** |  |  |  |  |  |  |  |  |
| **SUN 1120, 2013** | Rx: Sunitinib<br>Placebo | 819/<br>873 | 07/01/2008-08/01/2010 | 8.7 | OS | 13.1 vs. 11.8<br>0.91 (0.76-1.10) | 5.6 vs. 4.1<sup>†</sup><br>0.730.59-0.89) | ADT: 100<br>Docetaxel: 100 |

### (3) Prior ADT+docetaxel

A total of 9 phase III trials ^9,23,31,42,50,69,71,74,87^ reporting PFS and 11 ^8,9,13,23,31,36,50,69,71,75,87^ reporting OS were considered eligible for prior ADT+docetaxel subgroup.

#### a. Monotherapy

In terms of **ARPI monotherapy**, abiraterone (COU-AA-301) ^23^ was associated with a statistically significant improvement in both rPFS (0.66; 0.58-0.76) and OS (0.74; 0.64-0.86) compared to placebo. Enzalutamide (AFFIRM) ^9^ was associated with a statistically significant improvement in both rPFS (0.40; 0.35-0.47) and OS (0.63; 0.53-0.75) compared to placebo.

In terms of **chemotherapy monotherapy**, cabazitaxel 25 mg/m² (TROPIC) ^87^ was associated with a statistically significant improvement in both cPFS (0.74; 0.64-0.86) and OS (0.70; 0.59-0.83) compared to mitoxantrone.

In terms of **radiopharmaceutical monotherapy**, radium-223 (ALSYMPCA) ^13^ was associated with a statistically significant improvement in OS (0.70; 0.56-0.88) compared to placebo in patients who received prior docetaxel.

#### b. Combination therapy

In terms of **combinations of ARPI with PARPi,** niraparib combined with abiraterone (MAGNITUDE) ^42^ was not associated with a statistically significant improvement in rPFS (0.89; 0.48-1.66) compared to abiraterone alone in overall population. In patients with *HRR* alterations, niraparib + abiraterone was associated with a statistically significant improvement in both rPFS (0.76; 0.60-0.97) and OS (0.70; 0.49-0.99) after adjusting for cross-over. In patients with *BRCA* alterations, niraparib + abiraterone (MAGNITUDE) ^42,43^ was associated with a statistically significant improvement in both rPFS (0.55; 0.39-0.78) and OS (0.54; 0.33-0.90) after adjusting for cross-over, compared to abiraterone alone.

In patients with non-*BRCA* alterations, niraparib+abiraterone (MAGNITUDE) ^42^ was not associated with a statistically significant improvement in rPFS (0.99; 0.68-1.45) compared to abiraterone alone. OS was not reported.

Results for other mono-therapeutic agents and combination therapies in patients who received prior ADT+docetaxel are available in **Tables 3** and **6**. Summary of findings with certainty of evidence is outlined in **Supplement Tables 21 and 24**.

### (4) Prior ADT+ARPI+docetaxel

A total of four phase III/IV trials reporting PFS ^19,38,91,95^ and OS ^19,38,91,95^ were considered eligible for prior ADT+ARPI+docetaxel subgroup. These trials included patients who had progressed on/previously received both ARPI and docetaxel separately but not in combination with intent of triplet therapy in mHSPC setting.

In terms of **chemotherapy monotherapy**, cabazitaxel 25 mg/m² (CARD) ^95^ was associated with a statistically significant improvement in both rPFS (0.54; 0.40-0.73) and OS (0.64; 0.46-0.89) compared to enzalutamide/abiraterone.

In terms of **radioligand therapy**,^177^Lu-PSMA-617 added to standard of care (VISION) in PSMA-positive patients ^91^ was associated with a statistically significant improvement in both rPFS (0.40; 0.29-0.57) and OS (0.62; 0.52-0.74) compared to standard of care.

In terms of **TKI monotherapy**, cabozantinib (COMET-1) ^19^ was associated with a statistically significant improvement in rPFS (0.48; 0.40-0.57) but not in OS (0.90; 0.76-1.06) compared to prednisone.

Results for other combination therapies in patients who received prior ADT+ARPI+docetaxel are available in **Table 4**. Summary of findings with certainty of evidence is outlined in **Supplement Table 22** and **24.**

**Table 4.** Summary of characteristics and results of included phase III trials in which patients received prior ADT, ARPI and Docetaxel.

| Trial | Arm | Estimated /Actual Accrual | Years of enrollment | Median follow up - months | Primary Endpoint | OS HR (95% CI) | PFS HR (95% CI) | Prior therapy \| % |
| --- | --- | --- | --- | --- | --- | --- | --- | --- |
| **Chemotherapy monotherapy** |  |  |  |  |  |  |  |  |
| **CARD, 2019** | Rx: Cabazitaxel25<br>Ctrl: Enzalutamide /Abiraterone | 234/<br>255 | 11/01/2015-11/01/2018 | 9.2 | rPFS | 13.6 vs. 11<br>0.64 (0.46-0.89) * | 8 vs. 3.7<sup>†</sup><br>0.54 (0.40-0.73) | ADT: 100<br>Docetaxel: 100<br>ARPI: 99.6 |
| **PARPi+Immunotherapy** |  |  |  |  |  |  |  |  |
| **KEYLYNK-010, 2023** | Rx: Olaparib<br>+ Pembrolizumab<br>Ctrl: Enzalutamide /Abiraterone | 780/<br>793 | 05/30/2019-07/16/2021 | 18.7 | OS;<br>rPFS | 15.8 vs. 14.6<br>0.94 (0.77-1.14) | 4.6 vs. 4.2<sup>†</sup><br>0.96 (0.79-1.16) | ADT: 100<br>Docetaxel: 98<br>ARPI: 100 |
| **Radioligand therapy+Standard of Care** |  |  |  |  |  |  |  |  |
| **VISION, 2021** | Rx: 177Lu + SOC<br>Ctrl: SOC | 814/<br>831 | 06/04/2018-10/23/2019 | 20.9 | OS;<br>PFS | 15.3 vs. 11.3<br>0.62 (0.52-0.74) * | 8.7 vs. 3.4<sup>†</sup><br>0.40 (0.29-0.57) | ADT: 100<br>Docetaxel: 97<br>ARPI: 100 |
| **TKI monotherapy** |  |  |  |  |  |  |  |  |
| **COMET-1, 2016** | Rx: Cabozantinib<br>Ctrl: Prednisone | Not<br>specified/<br>1028 | 07/02/2012-11/14/2014 | NA | OS | 11 vs. 9.8<br>0.90 (0.76-1.06) | 5.6 vs. 2.8<sup>†</sup><br>0.48 (0.40-0.57) | ADT: 100<br>Docetaxel: 100<br>ARPI: 92 (Abi); 25 (Enza) |

Abbreviations: ADT: androgen deprivation therapy; OS: overall survival; PFS: progression-free survival; rPFS: radiographic progression-free survival; ARPI: androgen-receptor pathway inhibitor; PARPi: poly (ADP-ribose) polymerase inhibitor; TKI: tyrosine kinase inhibitor; 177Lu: 177Lu-PSMA-617; SOC: standard of care; Abi: abiraterone; Enza: enzalutamide
Note: The color 'green' represents a positive trial, 'red' represents a negative trial. Only CARD was a phase IV trial; the rest were phase III.
Cabazitaxel25: Cabazitaxel 25 mg/m<sup>2</sup> IV on day 1 of every 3-week cycle
\* Statistically significant overall survival benefit was observed
<sup>†</sup> Radiographic progression free survival

Results for mono-therapeutic agents and combination therapies in patients who received heterogeneous prior therapy are available in **Tables 5** and **6**. Summary of findings with certainty of evidence is outlined in **Supplement Tables 23-24.**

**Table 5.**
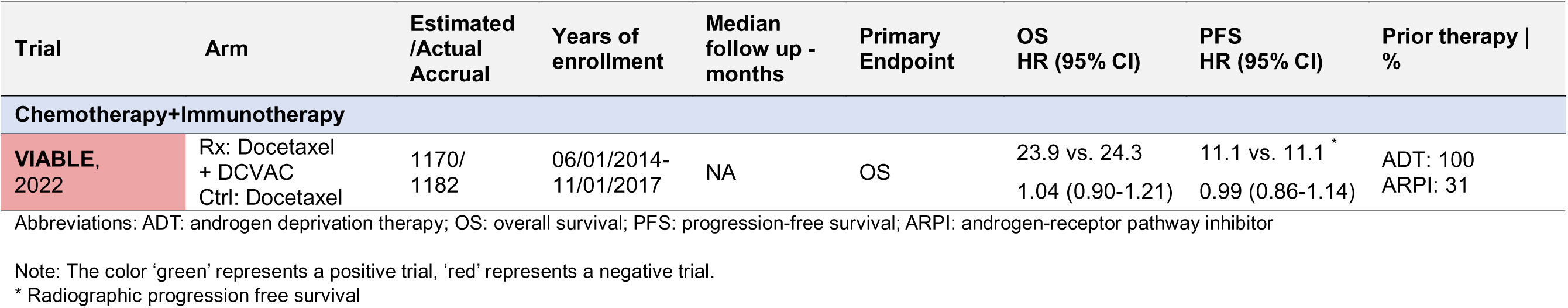
Summary of characteristics and results of included phase III trials in which patients received heterogeneous prior therapy.

## Discussion

This report from the living, interactive systematic review presents comprehensively synthesized and critically appraised relative and absolute effects of mCRPC systemic treatment options by prior therapy and relevant biomarkers using data from 143 randomized trials. It serves as the first systematic resource designed to continuously adapt as new data emerges, ensuring relevance for clinical practice and providing clinicians with a synthesized and appraised, evidence-based framework to support data-driven management strategies for mCRPC (**Table 7**). Detailed results are hosted on an interactive website (**<u>living website link</u>**).

In patients **with prior ADT and *HRR* alterations**, the combination of PARPi and ARPIs have emerged as new options. However, it is important to emphasize that different *HRR* alterations are not equivalent in eliciting a response to PARPi^193^. Abiraterone+olaparib ^65^, enzalutamide+talazoparib ^80^, and niraparib+abiraterone ^43^ demonstrated rPFS benefit over ARPI alone, with a greater benefit in *BRCA 1/2* subgroup. Meta-analysis adjusting for subsequent life-prolonging therapies and cross-over in the MAGNITUDE trial ^43^ showed consistent benefit in *BRCA1/2* group **(Supplement Table 25)**. For non-*BRCA HRR* alterations, meta-analysis pooling evidence from the PROpel, TALAPRO-2 (cohort 2), and MAGNITUDE trials demonstrated an rPFS benefit in *CDK12* subgroup, and a potential signal of benefit in *PALB2* subgroup. However, no survival benefit was observed in *ATM* or *CHEK2* subgroups (**Supplement Tables 25**). These findings are consistent with the recent FDA pooled analysis ^194^. Despite these analyses, the sample size for non-*BRCA HRR* genes was too small for a meaningful comparison. It is also important to consider the results from the BRCAAway trial ^183^ which showed improved rPFS with concurrent use of olaparib and abiraterone compared to either of drugs used alone or in sequence. Taken together, these findings support the combined use of PARPi and ARPI in patients with *BRCA1/2*, *CDK12*, or *PALB2* gene alterations who have previously received ADT alone (**Table 7**).

In **patients with prior ADT and no *HRR* alterations**, ARPI like abiraterone acetate (COU-AA-302) ^27,28^ or enzalutamide (PREVAIL) ^57^ may be considered due to rPFS and OS benefit. Docetaxel ^85^ can also be considered in progressive disease after ADT while cabazitaxel should be relegated to post-docetaxel setting considering results from the FIRSTANA trial ^34^ which showed no survival advantage over docetaxel in chemo-naïve setting.

In patients **with prior ARPI and *HRR* alterations** particularly in *BRCA1/2* genes, adding a PARPi (olaparib/rucaparib) may be an effective option. The PROfound trial ^60^ reported an rPFS benefit with olaparib in overall population. However, only 3.4% of the cohort had received ARPIs in pre-mCRPC setting, highlighting a gap in data for upfront use. Likewise, the TRITON2 ^195^ and TRITON3 ^86^ trials showed a consistent effect with rucaparib in patients with *HRR* genes alterations, particularly in *BRCA1/2* genes. Although no OS benefit was seen in TRITON3 trial, likely due to extensive cross-over and receipt of subsequent life-prolonging therapies (**Supplement Table 26**), these data solidify rucaparib’s role for *BRCA*-altered mCRPC, especially following ARPI failure. However, in *ATM*-altered cases, PARPi monotherapy did not show any survival benefit (**Supplement Table 27**). Given low representation of patients with prior ARPI in TALAPRO-2 ^80^, PROPEL ^65^, and MAGNITUDE ^43^ trials, the benefit with PARPi and ARPI is not generalizable to patients with prior exposure to ARPI. Also, considering the established cross-resistance between sequential ARPIs, ARPI switching after failure is not preferred.

In patients with **prior ARPI and no *HRR* alterations,** docetaxel remains an established first-line mCRPC therapy as it was the preferred subsequent therapy in trials assessing ARPI+ADT in mHSPC (**Supplement Table 28**). Several studies have explored strategies continuing ARPI at progression while adding docetaxel or ^177^Lu-PSMA-617 but none has showed definitive OS benefit. For example, PRESIDE ^55^ trial suggests modest rPFS benefit with the addition of docetaxel, while continuing enzalutamide beyond progression, although OS data were not reported. The phase II ABIDO-SOGUG ^153^ trial, did not meet the rPFS endpoint with the addition of docetaxel to abiraterone beyond progression. Moreover, emerging data from the phase III trial PSMAfore ^97^ demonstrated an rPFS benefit with ^177^Lu-PSMA-617 following ARPI failure in PSMA+ mCRPC patients compared to switching to another ARPI; however, with no OS benefit likely due to high cross-over. Likewise, recent phase II ENZA-p trial ^184^ also showed that ^177^Lu-PSMA-617 with enzalutamide improved PSA-PFS compared to enzalutamide alone in PSMA+ patients with high-risk features, though OS data are still pending. Despite advancements, limited data for ARPI-pretreated patients make optimizing post-ARPI treatment challenging, highlighting the need for randomized trials to define better options for these patients.

In patients **with prior docetaxel and *HRR* alterations** particularly in the *BRCA1/2* genes, PARPi with ARPI may be preferred. However, only 179 patients (22%) in PROpel, 179 (22%) in TALAPRO-2, and 85 (20%) in MAGNITUDE received prior docetaxel in mHSPC (**Supplement Table 11**). Subgroup data for prior docetaxel from PROpel (olaparib + abiraterone), and TALAPRO-2 (talazoparib + enzalutamide) showed that the PARPi with ARPI improved rPFS compared to ARPI alone (**Tables 1 and6**). However, the MAGNITUDE trial (niraparib + abiraterone) did not demonstrate an rPFS benefit in this patient population.

**Table 6.**
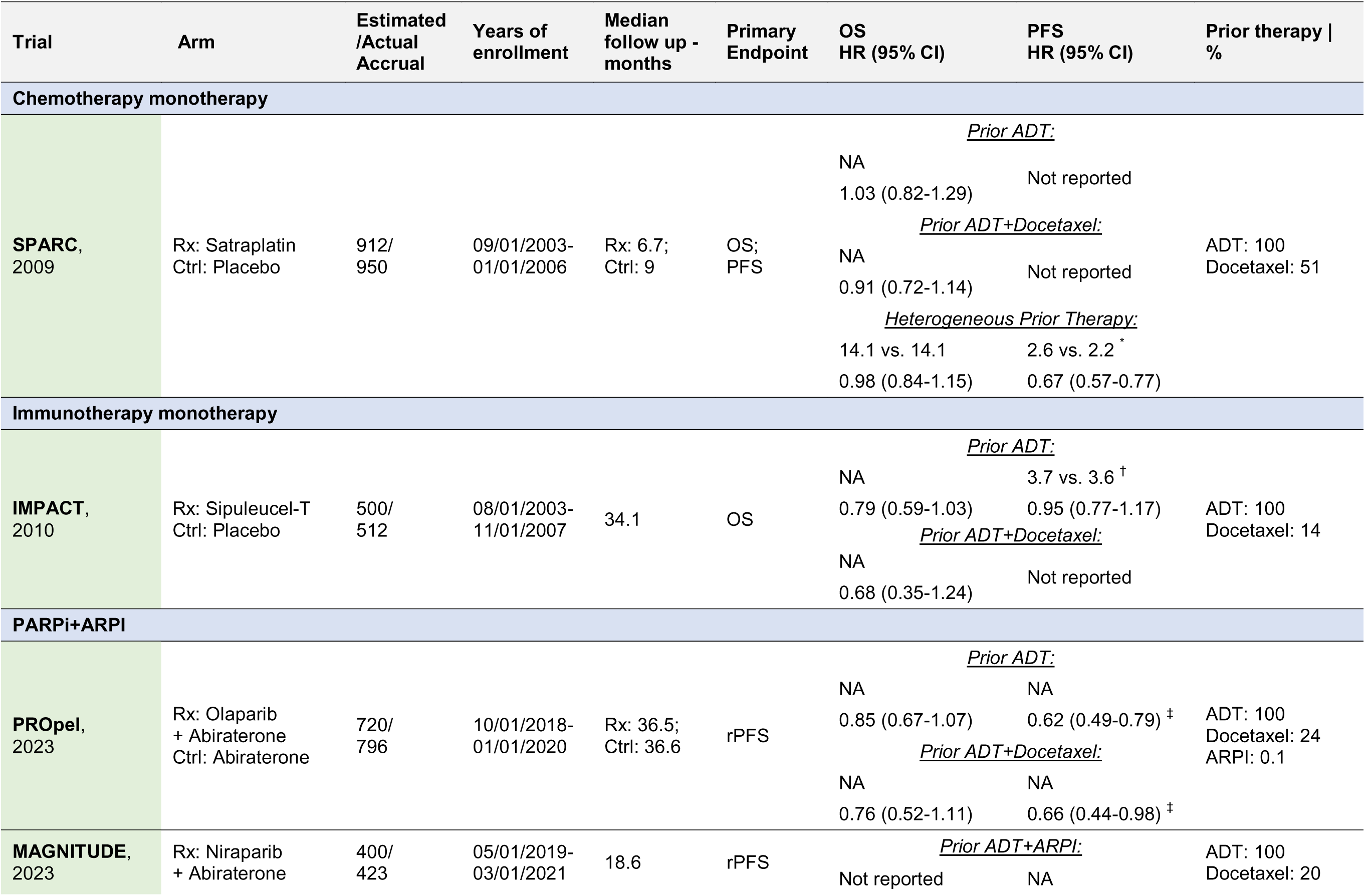
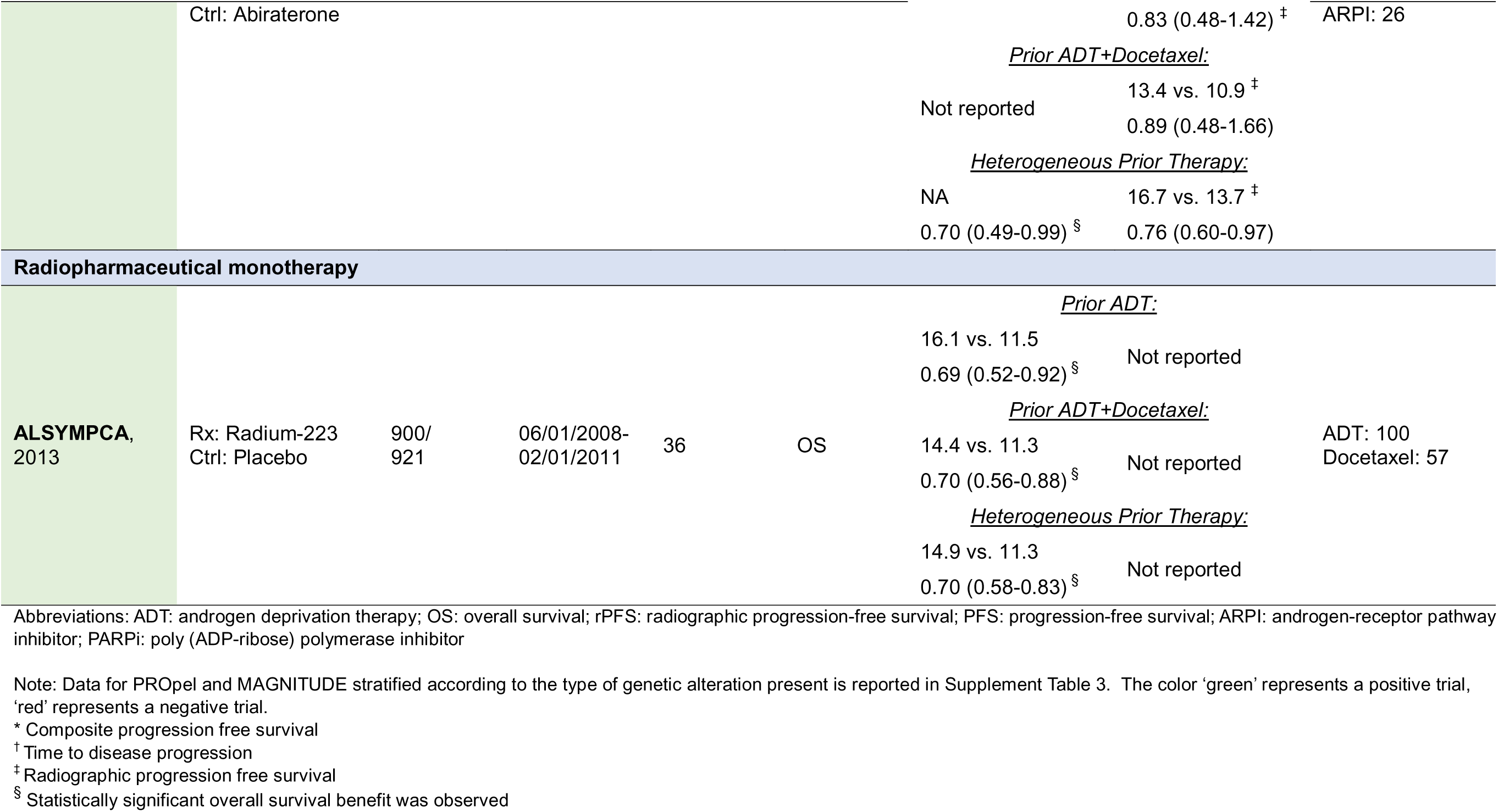
Summary of characteristics and results of included phase III trials that reported data for multiple subgroups.

**Table 7.** Summary of evidence.

| Prior ADT only |  |  | Prior ADT + ARPI |  |  | Prior ADT + Docetaxel |  |  | Prior ADT + ARPI + Docetaxel |  |  |
| --- | --- | --- | --- | --- | --- | --- | --- | --- | --- | --- | --- |
| Treatment | Trial | Evidence Category | Treatment | Trial | Evidence Category | Treatment | Trial | Evidence Category | Treatment | Trial | Evidence Category |
| HRR positive |  |  |  |  |  |  |  |  |  |  |  |
| BRCA positive |  |  |  |  |  |  |  |  |  |  |  |
| PARPi + ARPI | MAGNITUDE PROpel TALAPRO-2 | Category B | PARPi | PROfound TRITON-3 | Category B | PARPi + ARPI | MAGNITUDE PROpel TALAPRO-2 | Category B | PARPi | PROfound TRITON-3 | Category B |
| BRCA negative * |  |  |  |  |  |  |  |  |  |  |  |
| PARPi + ARPI | MAGNITUDE PROpel TALAPRO-2 | Category B | PARPi | PROfound | Category B | PARPi + ARPI | MAGNITUDE PROpel TALAPRO-2 | Category B | PARPi | PROfound | Category B |
| HRR negative |  |  |  |  |  |  |  |  |  |  |  |
| Abiraterone +Prednisone | COU-AA-302 | Category A | Docetaxel | TAX327 | Category B | Abiraterone +Prednisone | COU-AA-301 | Category A | Cabazitaxel | CARD | Category B |
| Enzalutamide | PREVAIL | Category A | Cabazitaxel † | FIRSTANA | Category B | Enzalutamide | AFFIRM | Category A |  |  |  |
| Docetaxel | TAX327 | Category A |  |  |  | Cabazitaxel | TROPIC | Category A |  |  |  |
| Cabazitaxel † | FIRSTANA | Category B |  |  |  |  |  |  |  |  |  |
| PSMA+ |  |  |  |  |  |  |  |  |  |  |  |
| - | - | - | - | - | - | - | - | - | 177LuPSMA 617 | VISION | Category B |
| Bone only, symptomatic |  |  |  |  |  |  |  |  |  |  |  |
| Radium-223 | ALSYMPCA | Category B | Radium-223 | ALSYMPCA | Category B | Radium-223 | ALSYMPCA | Category B | Radium-223 | ALSYMPCA | Category B |
| Indolent disease (slow rising PSA, asymptomatic/low volume disease) |  |  |  |  |  |  |  |  |  |  |  |
| Sipuleucel-T | D9901 D9902A IMPACT | Category B | Sipuleucel-T | D9901 D9902A IMPACT | Category B | Sipuleucel-T | D9901 D9902A IMPACT | Category B | Sipuleucel-T | D9901 D9902A IMPACT | Category B |
| MSI-high/dMMR |  |  |  |  |  |  |  |  |  |  |  |
| Pembrolizumab | KEYNOTE-158 | Category C | Pembrolizumab | KEYNOTE-158 | Category C | Pembrolizumab | KEYNOTE-158 | Category C | Pembrolizumab | KEYNOTE-158 | Category C |
| Oligometastatic disease/ progression |  |  |  |  |  |  |  |  |  |  |  |

| Radiotherapy/<br>Surgery | ARTO | Category<br>C | Radiotherapy/<br>Surgery | ARTO | Category<br>C | Radiotherapy/<br>Surgery | ARTO | Category<br>C | Radiotherapy/<br>Surgery | ARTO | Category<br>C |
| --- | --- | --- | --- | --- | --- | --- | --- | --- | --- | --- | --- |
| Disease Progression despite all above options |  |  |  |  |  |  |  |  |  |  |  |
| - | - | - | - | - | - | - | - | - | Cabazitaxel<br>+Carboplatin | MDACC<br>Study | Category<br>C |
| - | - | - | - | - | - | - | - | - | Carboplatin | - | Category<br>C |
| - | - | - | - | - | - | - | - | - | Best supportive/<br>palliative care | - | Category<br>C |
| **Category A:** Direct evidence from a randomized phase III clinical trial available<br>**Category B:** Consensus of panel of oncologists with extrapolated evidence from available randomized phase III clinical trials<br>**Category C:** Consensus of panel of oncologists without existing phase III evidence |  |  |  |  |  |  |  |  |  |  |  |

Abbreviations: ADT: androgen deprivation therapy; PARPi: poly (ADP-ribose) polymerase inhibitor; ARPI: androgen-receptor pathway inhibitor; MSI-high: microsatellite instability-high; dMMR: deficient mismatch repair
Note: ARTO<sup>199</sup> and KEYNOTE-158<sup>200</sup> were phase II trials that were not included in this systematic review because they did not meet the prespecified inclusion criteria. However, due to the lack of available trials for mCRPC patients with MSI-high/dMMR or oligometastatic disease progression, these trials were used to inform the recommendations based on the consensus of a panel of oncologists.
\* The greatest benefit in patients with non-*BRCA* *HRR* alterations were observed in those having alterations in *CDK12* and *PALB2* genes with limited benefit observed in *ATM* and *CHEK2* genes. PARPi + ARPI options include Niraparib + Abiraterone, Olaparib + Abiraterone, and Talazoparib + Enzalutamide. PARPi monotherapy options include Olaparib, and Rucaparib
† Cabazitaxel is an alternative option for docetaxel in patients who have experienced an allergic reaction or toxicity like peripheral neuropathy

In patients **with prior docetaxel and no *HRR* alterations**, abiraterone acetate (COU-AA-301) ^20^ and enzalutamide (AFFIRM) ^9^ showed survival benefits and may be considered. An alternative option is cabazitaxel, as supported by the phase III TROPIC trial ^87^, which demonstrated improved survival compared to mitoxantrone in this patient population.

In patients **with both ARPI and docetaxel**, there is a lack of direct evidence for management of mCRPC. There are no trials specifically reporting outcomes in patients who have received ‘true’ triplet therapy as a single, combined approach in hormone-sensitive disease. Instead, the existing data mainly pertain to patients with mCRPC who have progressed sequentially on an ARPI and a taxane or vice versa, rather than in combination. **In patients with *HRR* alterations**, especially in the *BRCA1/2* genes, PARPi (olaparib/rucaparib) may be preferred given significant rPFS benefits in this population, as seen in the PROfound ^60-64^ and TRITON3 ^86^ trials. **In patients without *HRR* alterations,** ^177^Lu-PSMA-617 offers survival benefits, as per results from the VISION trial ^91^. Likewise, increased PSA response, without any survival benefit, was observed with ^177^Lu-PSMA-617 compared to cabazitaxel in TheraP trial ^158,159^. Hence, cabazitaxel remains a suitable option for patients who can tolerate it and is superior to a second ARPI after sub-optimal response to the first. The CARD trial ^95^ showed a survival advantage with cabazitaxel over switching ARPI after progression with the caveat that the patients were required to have a sub-optimal response to the initial ARPI.

It is important to consider additional disease characteristics such as minimally symptomatic or bone only disease at progression. Radium-223 ^13^ improved OS and delayed skeletal related events in symptomatic bone metastases without nodal or visceral involvement (ALSYMPCA) and was safe in sequence with ^177^Lu-PSMA-617 (RALU) ^196^. The PEACE-3 trial ^197^ suggests combining radium-223 with enzalutamide reduces progression, though longer follow-up is needed for OS data, potentially offering a new first line option with a bone-protective agent and ADT, for ARPI-naïve mCRPC patients with bone metastases. Sipuleucel-T ^29,36,93^ may be limited to asymptomatic or minimally symptomatic disease, and pembrolizumab may be considered in high tumor mutational burden, dMMR or MSI-H patients who have been heavily pre-treated with standard therapies. However, these trials predate ARPI use or excluded prior ARPI, limiting relevance to current practice, where most patients will likely have progressed on an ARPI.

There are several strengths of our work. This ‘living’ review is the first comprehensive synthesis providing critically appraised randomized evidence from 141 clinical trials. Findings are organized and accessible through an interactive online platform, allowing users to filter data by variables like prior therapy, treatment type, specific treatments, control arms, and trial phase. As evidence in this field continues to evolve rapidly, with multiple phase III trials currently underway **(Supplement Table 29)**, this review is designed to incorporate new data using the living, interactive evidence synthesis framework allowing timely updates, and ensuring that clinicians have access to the most current and comprehensive evidence available for managing mCRPC. While the discussion of using large language models for LSRs is beyond the scope of this review, emerging evidence suggests that human-artificial intelligence interaction can tremendously facilitate the process without compromising on accuracy ^198^.

There are a few limitations. Variations in the eligibility criteria of included trials limited quantitative synthesis of evidence by meta-analyses and consequently, outcomes for some therapies relied on data from single trials, leading to imprecision. There was inconsistent reporting of outcome and subgroup analyses across trials. Not all trials provided data on rPFS. While we assessed OS as a more definitive endpoint, survival may have been underestimated due to cross-over and subsequent life-prolonging therapies in certain trials (**Supplement Table 26**), which were not consistently adjusted across studies. Another limitation arises from the rapid evolution of prostate cancer treatment standards over the past decade. Even the latest mCRPC trials were conducted before the current treatment regimens for mHSPC were established. This creates challenges in applying these findings to contemporary settings, especially for patients who have undergone intensified upfront treatments. Hence, the lack of direct evidence for patients progressing on ARPI agents or triplet therapy in mHSPC lowers certainty of the evidence. Likewise, thresholds for categorizing patients into one of the five categories by receipt of prior therapy were finalized by consensus, which may be arbitrary and contributes to a degree of indirectness.

## Data Availability

All data produced in the present work are contained in the manuscript.

## Neeraj Agarwal (NA)

NA received honorarium before May 2021 and during his lifetime for consulting to Astellas, AstraZeneca, Aveo, Bayer, Bristol Myers Squibb, Calithera, Clovis, Eisai, Eli Lilly, EMD Serono, Exelixis, Foundation Medicine, Genentech, Gilead, Janssen, Merck, MEI Pharma, Nektar, Novartis, Pfizer, Pharmacyclics, and Seattle Genetics. He has also received research funding during his lifetime (to NA’s institution) from Arnivas, Astellas, AstraZeneca, Bavarian Nordic, Bayer, Bristol Meyers Squibb, Calithera, Celldex, Clovis, CRISPR Therapeutics, Eisai, Eli Lilly, EMD Serono, Exelixis, Genentech, Gilead, Glaxo Smith Kline, Immunomedics, Janssen, Lava, Medivation, Merck, Nektar, Neoleukin, New Link Genetics, Novartis, Oric, Pfizer, Prometheus, Rexahn, Roche, Sanofi, Seattle Genetics, Takeda, and Tracon.

## Yousef Zakharia (YZ)

YZ has received honoraria for data safety monitoring board membership from Janssen Research and Development. He has served as a consultant or advisor to Roche/Genentech, Eisai, Amgen, Castle Biosciences, Novartis, Exelixis, Pfizer, Cardinal Health, Bayer, Janssen, TTC Oncology, Clovis Oncology, EMD Serono, Seagen, Bristol Myers Squibb/Medarex, Myovant Sciences, Genzyme, Gilead Sciences, AstraZeneca and Array BioPharma. He has received research funding to his institution from Pfizer, Exelixis, and Eisai. His travel, accommodations, and expenses have been supported by Newlink Genetics.

## Mary Ellen Taplin (MET)

MET has served on the advisory boards for Astellas, Novartis, Lakena, Flare, Pfizer, J&J, and AstraZeneca.

## Oliver Sartor (OS)

OS has received grants/contracts from Advanced Accelerator Applications, Amgen, AstraZeneca, Bayer, In Vitae, Janssen, Lantheus, Merck, Novartis, Sanofi, and Point Biopharma. He has also received consulting fees from Advanced Accelerator Applications, Amgen, ART Bioscience, Astellas Pharma, AstraZeneca, Bayer, Clarity Pharmaceuticals, EMD Serono, Fusion Pharmaceuticals, Isotopen Technologien, Janssen, MacroGenics, Novartis, Pfizer, Point Biopharma, Ratio, Sanofi, Telix Pharmaceuticals, and TeneoBio. Additionally, he has participated on a data safety monitoring board/advisory board for Pfizer, Merck, Janssen, AAA, Novartis, and AstraZeneca; received support for attending meeting and/or travel from Bayer, Lantheus, and Sanofi; and has stock/stock options in AbbVie, Cardinal Health, Clarity Pharmaceuticals, Convergent, Eli Lilly, Abbot, Ratio, United Health Group, and Telix.

## Parminder Singh (PS)

PS has served on the advisory boards for Aveo Pharmaceuticals, Bayer Healthcare Pharmaceuticals, EMD Serono Inc, and Janssen Research & Development, LLC.

## Daniel S. Childs (DSC)

DSC has received honoraria from Targeted Oncology, IntrinsiQ, MJH Life Sciences, and the International Centers for Precision Oncology Foundation. He has served as a consultant or advisor to Janssen Biotech (institution) and Novartis (institution) and received research funding to his institution from Janssen Biotech. His travel, accommodations, and expenses have been supported by the Prostate Cancer Foundation.

## Rahul A. Parikh (RP)

RP has stock and other ownership interests in IBRX. He has also received a patent on DNA repair pathways in cancer but does not receive any royalty from this.

## Rohan Garje (RG)

RG has received research funding to his institution from Endocyte/Advanced Accelerator Applications, Pfizer, Amgen, Immunomedics, Xencor, Exelixis, and Janssen Oncology.

## Alan Haruo Bryce (AHB)

AHB has received grants from Janssen and funding to his institution from Janssen, AstraZeneca, and Gilead. Additionally, he has received personal fees from AstraZeneca, Merck, Bayer, Elsevier, Fallon Medica, Horizon CME, PRIME Education, MJH Life Sciences, and Novartis outside the submitted work. He also holds a patent for therapeutic targeting of cancer patients with NRG1 rearrangements.

