## Supplement for "Systemic treatment options for metastatic castration resistant prostate cancer: A living systematic review"

**Online-only supplementary material**

### **Supplement Figure 1.** Risk of bias across the phase II trials included in this systematic review

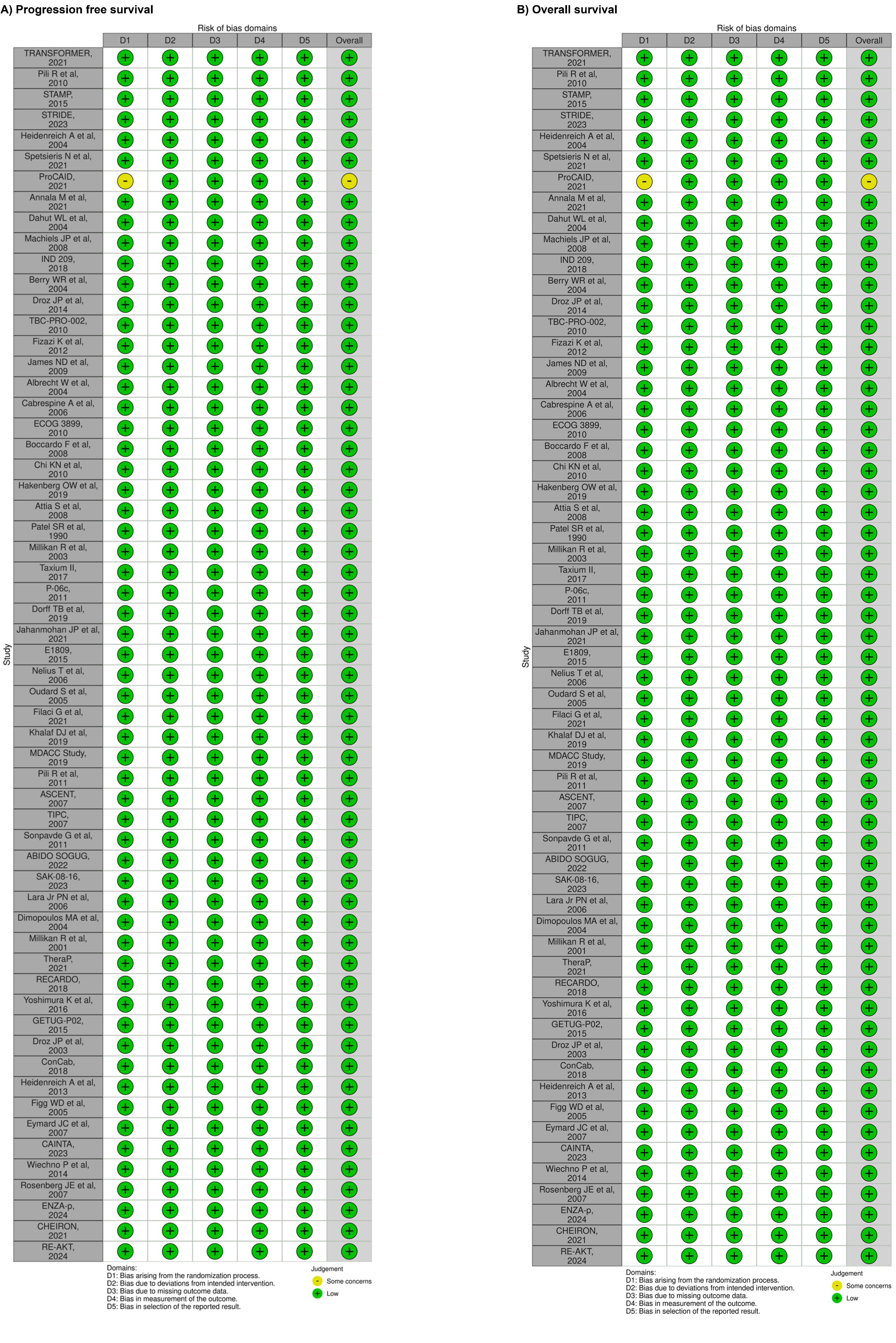

Note: Khalaf DJ et al (2019), Krainer M et al (2007), ABIDO SOGUG (2022), EORTC (2009), Berry WR et al (2004), Droz JP et al (2003), Eymard JC et al (2007), Horti J et al (2009), Pili R et al (2010), Stadler WM et al (2004), James ND et al (2009), Carducci MA et al (2003), and Bradley DA et al (2011) did not report progression-free survival, hence, time to tumor progression was assessed as a surrogate endpoint for risk of bias. Spetsieris N et al (2021) did not report progression-free survival, hence, time to treatment failure was assessed as a surrogate endpoint for risk of bias.

### **Supplement Figure 2.** Risk of bias across the phase III trials included in this systematic review

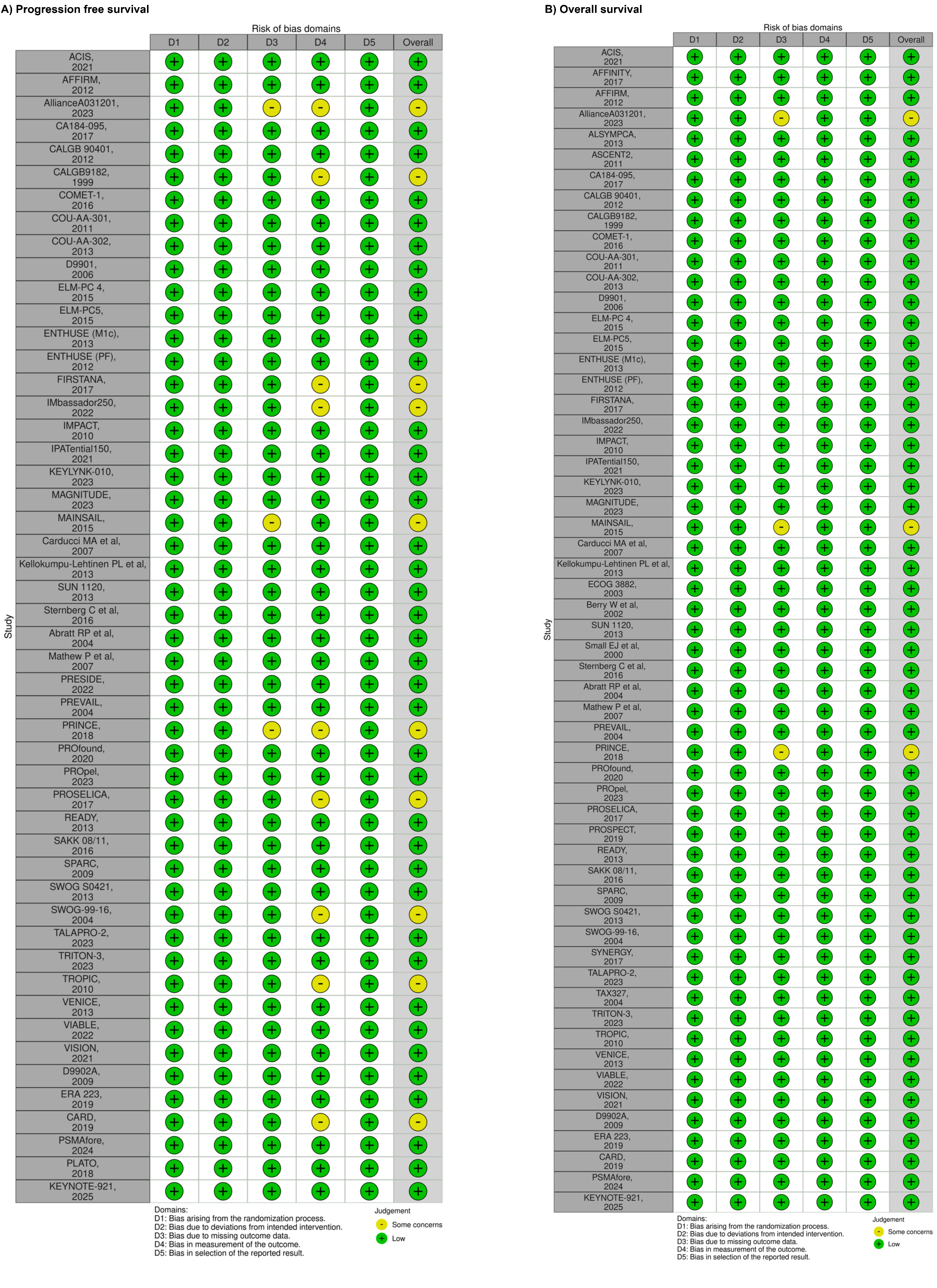

Note: CALGB9182 (1999), Kellokumpu-Lehtinen PL et al (2013), D9901 (2006), IMPACT (2010), and D9902A (2009) did not report progression-free survival, hence, time to tumor progression was assessed as a surrogate endpoint for risk of bias. PLATO and CARD are phase IV trials. The rest are phase III.

### **Supplement Table 1:** Outcome definitions across included phase III trials

| **Trial** | **Arm** | **Progression Free Survival (PFS)** | **Overall Survival (OS)** | **Time to tumor progression (TTP)** | **Radiographic Objective Response Rate (ORR)** | **PSA 50% Response** |
| --- | --- | --- | --- | --- | --- | --- |
| **Prior ADT only** | | | | | | |
| **ARPI monotherapy** | | | | | | |
| **COU-AA-302**, 2013 | Rx: Abiraterone Ctrl: Placebo | The time from randomization to the appearance of the first radiological evidence of progressive disease; defined as soft tissue lesions according to modified RECIST criteria or new bone lesions on bone scan according to PCWG2; or death from any cause. | The time from randomization to the date of death from any cause. | Not reported | The proportion of patients achieving a complete response or partial response according to RECIST criteria. | The proportion of patients with ≥50% decrease in PSA levels from baseline. |
| **PREVAIL**, 2014 | Rx: Enzalutamide Ctrl: Placebo | The time from the date of randomization to radiographic progression (soft tissue lesion progression according to RECIST v1.1, or bone lesion progression according to PCWG2 criteria) or death from any cause. | The time from randomization to the date of death from any cause. | Not reported | The proportion of patients achieving a complete response or partial response according to RECIST v1.1 criteria. | The proportion of patients with ≥50% decrease in the PSA value from the baseline, confirmed ≥3 weeks later. |
| **ELM-PC 4**, 2015 | Rx: TAK-700 Ctrl: Placebo | The time from the date of randomization to radiographic progression (soft tissue lesion progression according to RECIST v1.1, or bone lesion progression according to PCWG2 criteria) or death from any cause. | The time from randomization to the date of death from any cause. | Not reported | The proportion of patients achieving a complete response or partial response according to RECIST v1.1 and PCWG2 criteria, determined on 2 consecutive investigator assessments ≥4 weeks apart. | The proportion of patients with ≥50% decrease in PSA levels from baseline. |
| **ARPI+ARPI** | | | | | | |
| **ACIS**, 2021 | Rx: Abiraterone  + Apalutamide Ctrl: Abiraterone | The time from the date of randomization to radiographic progression (soft tissue lesion progression according to RECIST v1.1, or bone lesion progression according to PCWG2 criteria) death from any cause. | The time from randomization to the date of death from any cause. | Not reported | The proportion of patients achieving a complete response or partial response according to modified RECIST v1.1 criteria. | The proportion of patients with ≥50% decrease in PSA levels from baseline. |
| **AllianceA031201**, 2023 | Rx: Enzalutamide + Abiraterone Ctrl: Enzalutamide | Defined according to the PCWG2/3 criteria. | The time from study registration to the date of death from any cause. | Not reported | Not reported | The proportion of patients with ≥50% decrease in PSA levels from baseline. |
| **ARPI+PI3K/AKTi** | | | | | | |
| **IPATential150**, 2021 | Rx: Abiraterone + Ipatasertib Ctrl: Abiraterone | The time from randomization to the occurrence of radiographic disease progression defined according to PCWG3 criteria or death from any cause. | The time from randomization to the date of death from any cause. | Not reported | The proportion of patients achieving a complete response or partial response according to RECIST v1.1 and PCWG3 criteria. | The proportion of patients with ≥50% decrease in PSA levels from baseline. |
| **ARPI+Radiopharmaceutical** | | | | | | |
| **ERA 223**, 2019 | Rx: Abiraterone + Radium-223 Ctrl: Abiraterone | The time from randomization to radiologic progression or death from any cause. | The time from randomization to the date of death from any cause. | Not reported | Not reported | Not reported |
| **Chemotherapy monotherapy** | | | | | | |
| **CALGB9182**, 1999 | Rx: Mitoxantrone + Hydrocortisone Ctrl: Hydrocortisone | Not reported | The time from randomization to the date of death from any cause. | The time from randomization to disease progression (defined as worsening performance status of ≥ 1, appearance of ≥2 new lesions on bone scan, or an increase of serum PSA level ≥100% above baseline) or death. | Not reported | The proportion of patients with ≥50% decrease in PSA levels from baseline at a follow-up visit between 4 to 8 weeks. |
| **FIRSTANA**, 2017 | Rx1: Cabazitaxel20 Rx2: Cabazitaxel25 Ctrl: Docetaxel | The time from randomization to the occurrence of tumor progression (according to RECIST v1.1 criteria), PSA progression, pain progression or death from any cause. | The time from randomization to the date of death from any cause. | Not reported | The proportion of patients achieving a complete response or partial response according to RECIST v1.1 criteria. | The proportion of patients with ≥50% decrease in PSA levels from baseline for two successive evaluations at least 3 weeks apart, in patients with baseline PSA value of ≥10 ng/ml. |
| **Kellokumpu-Lehtinen PL et al**, 2013 | Rx: Docetaxel (2W) Ctrl: Docetaxel | Not reported | The time from randomization to the date of death from any cause. | The time from randomization to cancer progression or death due to any cause. | Not reported | The proportion of patients with ≥50% decrease in the PSA value from the baseline, confirmed ≥4 weeks later. |
| **Berry W et al**, 2002 | Rx: Mitoxantrone Ctrl: Placebo | Not reported | Reported but not defined | Not reported | Reported but not defined | The proportion of patients with a ≥50% decline lasting ≥2 months with stabilization or improvement of performance status for ≥2 weeks. |
| **Abratt RP et al**, 2004 | Rx: Vinorelbine + Hydrocortisone Ctrl: Hydrocortisone | The time from the date of randomization to progression (appearance of a new lesion or a ≥25% increase in an existing lesion, PSA progression, increase in pain intensity, or deterioration in Karnofsky performance status) or death from any cause. | Reported but not defined | Not reported | Evaluated according to WHO criteria. | The proportion of patients with ≥50% decrease in the PSA value maintained for ≥6 weeks. |
| **PRINCE**, 2018 | Rx: Docetaxel (int) Ctrl: Docetaxel | The time from the date of randomization to progression (PSA >4ng/ml with 50% increase compared to baseline over 2 consecutive visits, radiological progression per RECIST criteria, or symptomatic progression) or death from any cause. | Reported but not defined | Not reported | Not reported | Not reported |
| **TAX327**, 2004 | Rx1: Docetaxel Rx2: Docetaxel30 (W) Ctrl: Mitoxantrone | Not reported | Reported but not defined | Not reported | The proportion of patients achieving a complete response or partial response according to WHO criteria. | The proportion of patients with ≥50% decrease in the PSA value from the baseline, confirmed ≥3 weeks later. |
| **Chemotherapy+ASO** | | | | | | |
| **SYNERGY**, 2017 | Rx: Docetaxel + Custirsen Ctrl: Docetaxel | Not reported | The time from randomization to the date of death from any cause. | Not reported | Not reported | The proportion of patients with ≥50% decrease in PSA levels from baseline, based on PCWG2 criteria. |
| **Chemotherapy+Chemotherapy** | | | | | | |
| **SWOG-99-16**, 2004 | Rx: Docetaxel60 + Estramustine Ctrl: Mitoxantrone | The time from randomization to the occurrence of objective progression, PSA progression, or death from any cause. | The time from randomization to the date of death from any cause. | Not reported | The proportion of patients achieving a complete response or partial response according to WHO criteria. | The proportion of patients with ≥50% decrease in the PSA value from the baseline, confirmed ≥4 weeks later. |
| **Chemotherapy+DES** | | | | | | |
| **ECOG 3882** 2003 | Rx: Doxorubicin + DES Ctrl: Doxorubicin | Not reported | Reported but not defined | Not reported | Not reported | Not reported |
| **Chemotherapy+ERA** | | | | | | |
| **ENTHUSE (M1c)**, 2013 | Rx: Docetaxel + Zibotentan Ctrl: Docetaxel | The time from the date of randomization to clinical progression (≥4 new bony lesions on bone scan/CT/MRI, increased pain, skeletal-related event, new/progressed soft tissue disease according to modified RECIST criteria) or death from any cause. | The time from randomization to the date of death from any cause. | Not reported | Not reported | The proportion of patients with ≥50% decrease in PSA levels from baseline for two successive evaluations at least 2 weeks apart. |
| **SWOG S0421**, 2013 | Rx: Docetaxel + Atrasentan Ctrl: Docetaxel | The time from randomization to progression (increased pain/analgesia, soft tissue disease per RECIST, new lesions on bone scan) or death from any cause. | The time from randomization to the date of death from any cause. | Not reported | The proportion of patients achieving a complete response or partial response according to RECIST criteria. | The proportion of patients with ≥50% decrease in PSA levels from baseline. |
| **Chemotherapy+IMiD** | | | | | | |
| **MAINSAIL**, 2015 | Rx: Docetaxel + Lenalidomide Ctrl: Docetaxel | The time from randomization to disease progression assessed by investigators per RECIST v1.1 criteria, or death from any cause. | The time from randomization to the date of death from any cause. | Not reported | The proportion of patients achieving a complete response or partial response according to RECIST v1.1 criteria. | The proportion of patients with ≥50% decrease in PSA levels from baseline. |
| **Chemotherapy+PDGFRi** | | | | | | |
| **Mathew P et al**, 2007 | Rx: Docetaxel30 (W) + Imatinib Ctrl: Docetaxel30 (W) | The time from the date of randomization to progression or death from any cause. Progression was defined as the appearance of symptoms, clinical signs, radiographic evidence of disease progression, or the first of consecutive PSA increases of ≥25% over baseline or nadir. | Reported but not defined | Not reported | Not reported | The proportion of patients with ≥50% decrease in the PSA value maintained for ≥6 weeks. |
| **Chemotherapy+TKI** | | | | | | |
| **READY**, 2013 | Rx: Docetaxel + Dasatinib Ctrl: Docetaxel | The time from randomization to the progression or death due to any cause. Progression was defined as any skeletal-related event, tumor progression according to modified RECIST (accompanied with either PSA progression or investigator-defined clinical progression) or presence of ≥2 new lesions on bone scan confirmed ≥6 weeks later. | The time from randomization to the date of death from any cause. | Not reported | The proportion of patients achieving a complete response or partial response according to RECIST criteria. | Not reported |
| **Chemotherapy+VEGFi** | | | | | | |
| **CALGB 90401**, 2012 | Rx: Docetaxel + Bevacizumab Ctrl: Docetaxel | The time from randomization to disease progression (defined by using PSAWG1 criteria with the exception that more than 2 new bone lesions were required for bone progression on a bone scan) or death from any cause. | The time from randomization to the date of death from any cause. | Not reported | The proportion of patients achieving a complete response or partial response according to RECIST criteria. | The proportion of patients with ≥50% decrease in PSA levels from baseline for two successive evaluations at least 4 weeks apart. |
| **VENICE**, 2013 | Rx: Docetaxel+ Aflibercept Ctrl: Docetaxel | The time from randomization to tumor progression, PSA progression, occurrence of a skeletal-related event, pain progression, radiotherapy for cancer-related symptoms or death, whichever came first. | The time from randomization to the date of death from any cause. | Not reported | The proportion of patients achieving a complete response or partial response according to RECIST criteria. | The proportion of patients with ≥50% decrease in the PSA value from the baseline, confirmed ≥3 weeks later. |
| **Chemotherapy+VitD-Analog** | | | | | | |
| **ASCENT2**, 2011 | Rx: Docetaxel + Calcitriol Ctrl: Docetaxel | Not reported | The time from randomization to the date of death from any cause. | Not reported | Not reported | Not reported |
| **ERA monotherapy** | | | | | | |
| **ENTHUSE (PF)**, 2012 | Rx: Zibotentan Ctrl: Placebo | The time from the date of randomization to clinical progression (≥4 new bony lesions confirmed on bone scan, increased pain, skeletal-related event, objective progression of visceral/nodal disease according to modified RECIST criteria) or death from any cause. | The time from randomization to the date of death from any cause. | Not reported | Not reported | Not reported |
| **Carducci MA et al**, 2007 | Rx: Atrasentan Ctrl: Placebo | Not reported | Reported but not defined | The time from randomization to the disease progression; defined as first occurrence of either ≥2 new bone lesions seen on bone scan, ≥1 new/progressed lesion (according to modified RECIST criteria), metastatic pain, skeletal-related event or need for new intervention. | Not reported | Not reported |
| **IMiD monotherapy** | | | | | | |
| **Sternberg C et al**, 2016 | Rx: Tasquinimod Ctrl: Placebo | The time from the date of randomization to radiographic progression (soft tissue lesion progression according to RECIST v1.1, bone lesion progression according to PCWG2 criteria, or radiographically confirmed spinal cord compression/fracture due to malignant progression) or death from any cause. | The time from randomization to the date of death from any cause. | Reported but not defined | Not reported | Not reported |
| **Immunotherapy monotherapy** | | | | | | |
| **CA184-095**, 2017 | Rx: Ipilimumab Ctrl: Placebo | The time from randomization to the earliest date of confirmed PSA progression, radiologic progression, clinical deterioration, or death from any cause (as determined by the investigator). | The time from randomization to the date of death from any cause. | Not reported | Not reported | The proportion of patients with ≥50% decrease in PSA levels from baseline that was confirmed by a second PSA value ≥6 weeks later. |
| **D9901**, 2006 | Rx: Sipuleucel-T  Ctrl: Placebo | Not reported | Reported but not defined | The time from randomization to disease progression seen on radiographic imaging, new cancer-related pain or clinical events such as spinal cord compression, nerve root compression, or pathologic fracture. | Not reported | Not reported |
| **D9902A**, 2009 | Rx: Sipuleucel-T  Ctrl: Placebo | Not reported | The time from randomization to the date of death from any cause. | The time from randomization to disease progression seen on radiographic imaging, new cancer-related pain or clinical events such as spinal cord compression, nerve root compression, or pathologic fracture. | Not reported | Not reported |
| **Immunotherapy+GMCSF** | | | | | | |
| **PROSPECT**, 2019 | Rx1: PVAC + GMCSF Rx2: PVAC Ctrl: Placebo | Not reported | The time from randomization to the date of death from any cause. | Not reported | The proportion of patients achieving a complete response or partial response according to RECIST v1.1 criteria. | Not reported |
| **PARPi+ARPI** | | | | | | |
| **TALAPRO-2**, 2023 | Rx: Talazoparib + Enzalutamide Ctrl: Enzalutamide | The time from randomization to radiographic progression assessed by blinded independent central review per RECIST 1.1 (soft tissue) and PCWG-3 criteria (bone) or death from any cause, whichever occurs first. | The time from randomization to the date of death from any cause. | Not reported | The proportion of patients with measurable soft tissue disease at baseline who achieve an objective response assessed by blinded independent central review per RECIST 1.1 criteria. | The proportion of patients with ≥50% decrease in PSA levels from baseline. |
| **Phenylurea monotherapy** | | | | | | |
| **Small EJ et al**, 2000 | Rx: Suramin Ctrl: Placebo | Not reported | Reported but not defined | Not reported | Not reported | The proportion of patients with ≥50% decrease in the PSA value maintained for ≥4 weeks. |
| **Prior ADT+ARPI** | | | | | | |
| **ARPI+ARPI** | | | | | | |
| **PLATO**, 2018 | Rx: Enzalutamide + Abiraterone Ctrl: Abiraterone | The time from randomization to first evidence of radiographic progression (PCWG2 criteria for bone lesions, RECIST v1.1 for soft tissue lesions), unequivocal clinical progression (new onset prostate cancer pain requiring opiates, deterioration of ECOG performance status ≥3 due to prostate cancer, or initiation of cytotoxic chemotherapy, radiation, or surgical intervention), or death from any cause. | Not reported | Not reported | The proportion of patients achieving a complete response or partial response according to RECIST v1.1 criteria. | The proportion of patients with ≥50% decrease in the PSA value from the baseline, confirmed ≥3 weeks later. |
| **ARPI+Immunotherapy** | | | | | | |
| **IMbassador250**, 2022 | Rx: Enzalutamide + Atezolizumab Ctrl: Enzalutamide | The time from randomization to soft tissue lesion progression as defined per PCWG3 modified RECIST v1.1 criteria or ≥2 new bone lesions confirmed by a second bone scan ≥6 weeks later, or death from any cause. | The time from randomization to the date of death from any cause. | Not reported | The proportion of patients achieving a complete response or partial response on 2 consecutive occasions ≥6 weeks apart, as determined by the PCWG3 criteria. | The proportion of patients with ≥50% decrease in PSA levels from baseline, confirmed 3 weeks later. |
| **Chemotherapy+ARPI** | | | | | | |
| **PRESIDE**, 2022 | Rx: Docetaxel  + Enzalutamide Ctrl: Docetaxel | The time from the date of randomization to progression or death within 112 days of treatment discontinuation without objective evidence of radiographic progression. Progression could be either radiographic (bone/soft tissue) or clinical (new onset of cancer pain requiring opiate analgesia, deterioration to an ECOG PS ≥3; or initiation of subsequent lines of cytotoxic chemotherapy, radiotherapy, or surgical intervention). | Not reported | Not reported | The proportion of patients achieving a complete response or partial response according to RECIST v1.1 criteria. | The proportion of patients with ≥50% decrease in PSA levels from baseline. |
| **Chemotherapy+Immunotherapy** | | | | | | |
| **KEYNOTE-921**, 2025 | Rx: Docetaxel  + Pembrolizumab Ctrl: Docetaxel | The time from randomization to  radiographic disease progression by BICR per PCWG3-modified RECIST 1.1 or death due to any cause, whichever occurred first. | The time from randomization to the date of death from any cause. | Not reported | The proportion of patients achieving a complete response or partial response according to PCWG3-modified RECIST v1.1 criteria by BICR. | The proportion of patients with ≥50% decrease in PSA levels from baseline, confirmed 3 weeks later. |
| **PARPi monotherapy** | | | | | | |
| **TRITON-3**, 2023 | Rx: Rucaparib Ctrl: Enzalutamide /Abiraterone /Docetaxel | The time from randomization to radiographic progression assessed by central independent radiology review per RECIST 1.1 (soft tissue) and PCWG-3 criteria (bone) or death from any cause, whichever occurs first. | The time from randomization to the date of death from any cause. | Not reported | The proportion of patients with measurable disease at baseline who achieve a complete or partial tumor response using radiographic response assessed by RECIST 1.1 (soft tissue) and no progression in bone per PCWG3 criteria. | The proportion of patients with ≥50% decrease in the PSA value from the baseline, confirmed ≥3 weeks later. |
| **PROfound**, 2020 | Rx: Olaparib Ctrl: Enzalutamide /Abiraterone | The time from randomization to radiographic progression assessed by blinded independent central review per RECIST 1.1 (soft tissue) and PCWG-3 criteria (bone) or death from any cause, whichever occurs first. | The time from randomization to the date of death from any cause. | Not reported | The proportion of patients with a confirmed partial response or complete response per RECIST 1.1 criteria by independent central review and no evidence of bone progression per PCWG3 criteria. | The proportion of patients with ≥50% decrease in the PSA value from the baseline, confirmed ≥3 weeks later. |
| **Radioligand monotherapy** | | | | | | |
| **PSMAfore**, 2024 | Rx: 177Lu Ctrl: Enzalutamide /Abiraterone | The time from randomization to first documented radiographic disease progression according to PCWG3-modified RECIST v1.1 assessed by BICR, or death. | The time from randomization to the date of death from any cause. | Not reported | The proportion of patients achieving a complete response or partial response according to PCWG3-modified RECIST v1.1 criteria, as assessed by BICR. | The proportion of patients with ≥50% decrease in the PSA value from the baseline, confirmed by a second assessment within 4 weeks. |
| **Prior ADT+Docetaxel** | | | | | | |
| **ARPI monotherapy** | | | | | | |
| **AFFIRM**, 2012 | Rx: Enzalutamide Ctrl: Placebo | The time from randomization to the appearance of the first radiological evidence of progressive disease; defined by RECIST v1.1 for soft tissue disease, or the appearance of ≥2 new bone lesions on bone scan, as per the PCWG2 guidelines; or death from any cause. | The time from randomization to the date of death from any cause. | Not reported | Not reported | The proportion of patients with ≥50% decrease in PSA levels from baseline. |
| **COU-AA-301**, 2011 | Rx: Abiraterone Ctrl: Placebo | The time from randomization to the appearance of the first radiological evidence of progressive disease or death. Progressively increasing soft tissue lesions according to modified RECIST criteria (a baseline lymph node of ≥2.0 cm considered to be a target lesion) or progression according to bone scans showing ≥2 new lesions not consistent with tumor flare. | The time from randomization to the date of death from any cause. | Not reported | Not reported | The proportion of patients with ≥50% decrease in the PSA value from the baseline, confirmed ≥4 weeks later. |
| **ELM-PC 5**, 2015 | Rx: TAK-700 Ctrl: Placebo | The time from the date of randomization to radiographic progression (soft tissue lesion progression according to RECIST v1.1, or bone lesion progression according to PCWG2 criteria) or death from any cause. | The time from randomization to the date of death from any cause. | Not reported | The proportion of patients achieving a complete response or partial response according to RECIST v1.1 criteria. | The proportion of patients with ≥50% decrease in PSA levels from baseline. |
| **SAKK 08/11**, 2016 | Rx: TAK-700 Ctrl: Placebo | The time from the date of randomization to radiographic progression (soft tissue lesion progression according to modified RECIST v1.1, or bone lesion progression according to PCWG2 criteria) or death from any cause. | The time from randomization to the date of death from any cause. | Not reported | Not reported | The proportion of patients with ≥50% decrease in PSA levels from baseline. |
| **Chemotherapy monotherapy** | | | | | | |
| **PROSELICA**, 2017 | Rx: Cabazitaxel20 Ctrl: Cabazitaxel25 | The time from randomization to the first occurrence of radiographic progression (per RECIST v1.1), PSA progression, pain progression, or death from any cause. | The time from randomization to the date of death from any cause. | The time from randomization to the first radiologic evidence of progression, per RECIST 1.1. | The proportion of patients achieving a complete response or partial response according to RECIST v1.1 criteria. | The proportion of patients with ≥50% decrease in the PSA value maintained for ≥3 weeks. |
| **TROPIC**, 2010 | Rx: Cabazitaxel25 Ctrl: Mitoxantrone | The time from randomization to disease progression (PSA, tumor or pain progression) or death from any cause. | The time from randomization to the date of death from any cause. | The time from randomization to the evidence of progressive disease as measured by RECIST criteria. | The proportion of patients achieving a complete response or partial response according to RECIST criteria. | A ≥ 50% reduction in serum PSA, determined only for patients with a serum PSA ≥ 20ng/mL at baseline, confirmed ≥ 3 weeks later. |
| **Chemotherapy+ASO** | | | | | | |
| **AFFINITY**, 2017 | Rx: Cabazitaxel25  + Custirsen Ctrl: Cabazitaxel25 | Not reported | The time from randomization to the date of death from any cause. | Not reported | Not reported | The proportion of patients with a ≥50% decline (minimum of 5 ng/mL) in PSA from baseline, maintained for ≥ 3 weeks, without evidence of either objective progressive disease or PSA progression. |
| **TKI monotherapy** | | | | | | |
| **SUN 1120**, 2013 | Rx: Sunitinib Placebo | The time from randomization to objective radiographic progression or death from any cause. | The time from randomization to the date of death from any cause. | Not reported | The proportion of patients achieving a complete response or partial response according to RECIST criteria. | Not reported |
| **Prior ADT+Docetaxel+ARPI** | | | | | | |
| **Chemotherapy monotherapy** | | | | | | |
| **CARD**, 2019 | Rx: Cabazitaxel25 Ctrl: Enzalutamide /Abiraterone | The time from the date of randomization to radiographic progression (soft tissue lesion progression according to RECIST version 1.1, or bone lesion progression according to PCWG2 criteria) or death. | The time from randomization to the date of death from any cause. | Not reported | The proportion of patients achieving a complete response or partial response according to RECIST v1.1 criteria. | The proportion of patients with ≥50% decrease in the PSA value from the baseline, confirmed ≥3 weeks later. |
| **PARPi+Immunotherapy** | | | | | | |
| **KEYLYNK-010**, 2023 | Rx: Olaparib + Pembrolizumab Ctrl: Enzalutamide /Abiraterone | The time from randomization to the occurrence of radiographic disease progression defined according to PCWG-modified RECIST v1.1 criteria assessed by blinded independent central review or death. | The time from randomization to the date of death from any cause. | Not reported | The proportion of patients achieving a complete response or partial response according to PCWG-modified RECIST v1.1 criteria assessed by blinded independent central review. | The proportion of patients with ≥50% decrease in PSA levels from baseline. |
| **Radioligand therapy+Standard of Care** | | | | | | |
| **VISION**, 2021 | Rx: 177Lu + SOC Ctrl: SOC | The time from randomization to independently centrally reviewed disease progression (per PCWG3 criteria) or death. | The time from randomization to the date of death from any cause. | Not reported | The proportion of patients with measurable soft tissue disease at baseline who achieve an objective response assessed by blinded independent central review per RECIST 1.1 criteria. | Not reported |
| **TKI monotherapy** | | | | | | |
| **COMET-1**, 2016 | Rx: Cabozantinib Ctrl: Prednisone | The time from randomization to the first radiographic progression per modified RECIST v1.1 (bone and/or soft tissue) or death from any cause. | The time from randomization to the date of death from any cause. | Not reported | Not reported | The proportion of patients with ≥50% decrease in PSA levels from baseline. |
| **Heterogeneous Prior Therapy** | | | | | | |
| **Chemotherapy+Immunotherapy** | | | | | | |
| **VIABLE**, 2022 | Rx: Docetaxel + DCVAC Ctrl: Docetaxel | The time from randomization to the occurrence of objective evidence of radiographic progression or death due to any cause. | The time from randomization to the date of death from any cause. | Not reported | Not reported | Not reported |
| **Multiple subgroups (ADT/ADT+ARPI/ADT+Docetaxel/ADT+ARPI+Docetaxel/Heterogeneous prior therapy)** | | | | | | |
| **Chemotherapy monotherapy** | | | | | | |
| **SPARC**, 2009 | Rx: Satraplatin Ctrl: Placebo | The time from randomization to the occurrence of tumor progression, skeletal-related events, symptomatic progression, or death from any cause. | The time from randomization to the date of death from any cause. | Not reported | The proportion of patients achieving a complete response or partial response according to RECIST criteria. | The proportion of patients with ≥50% decrease in the PSA value from the baseline, confirmed ≥4 weeks later. |
| **Immunotherapy monotherapy** | | | | | | |
| **IMPACT**, 2010 | Rx: Sipuleucel-T  Ctrl: Placebo | Not reported | The time from randomization to the date of death from any cause. | The time from randomization to objective disease progression; defined as either an increase of ≥50% in the sum of the products of diameters for index lesions, new appearance/unequivocal progression of non-index lesions, ≥2 new lesions on bone scan, a new pathologic fracture or spinal cord compression. | Not reported | Not reported |
| **PARPi+ARPI** | | | | | | |
| **PROpel**, 2023 | Rx: Olaparib + Abiraterone Ctrl: Abiraterone | The time from randomization to radiographic progression assessed by investigator per RECIST 1.1 (soft tissue) and PCWG-3 criteria (bone) or death from any cause, whichever occurs first. | The time from randomization to the date of death from any cause. | Not reported | The proportion of patients with measurable disease at baseline who achieve a complete or partial tumor response using radiographic response assessed by RECIST 1.1 (soft tissue) and PCWG-3 criteria (bone). | The proportion of patients with ≥50% decrease in the PSA value from the baseline, confirmed ≥3 weeks later. |
| **MAGNITUDE**, 2023 | Rx: Niraparib + Abiraterone Ctrl: Abiraterone | The time from randomization to radiographic progression assessed by blinded independent central review per RECIST v1.1 (soft tissue) and PCWG-3 criteria (bone) or death from any cause, whichever occurs first. | The time from randomization to the date of death from any cause. | Not reported | The proportion of patients achieving a complete response or partial response according to RECIST v1.1 criteria with no evidence of bone progression according to the PCWG3 criteria, as assessed by central review. | Not reported |
| **Radiopharmaceutical monotherapy** | | | | | | |
| **ALSYMPCA**, 2013 | Rx: Radium-223 Ctrl: Placebo | Not reported | The time from randomization to the date of death from any cause. | Not reported | Not reported | Not reported |

Abbreviations: ADT: androgen deprivation therapy; ARPI: androgen-receptor pathway inhibitor; RECIST: response evaluation criteria in solid tumors; PCWG: prostate cancer working group; PSA: prostate-specific antigen; PI3K/AKTi: phosphatidylinositol 3-kinase and protein kinase B inhibitor; WHO: world health organization; PSAWG: prostate-specific antigen working group; ASO: antisense oligonucleotide; DES: diethylstilbestrol diphosphate; ERA: endothelin receptor antagonist; IMiD: immunomodulatory drug; PDGFRi: platelet-derived growth factor receptor inhibitor; TKI: tyrosine kinase inhibitor; VEGFi: vascular endothelial growth factor inhibitor; VitD: vitamin d; PVAC: PROSTVAC (viral vector-based immunotherapy); GMCSF: granulocyte-macrophage colony-stimulating factor; ECOG PS: eastern cooperative oncology group performance status; BICR: blinded independent central review; 177Lu: 177Lu-PSMA-617; PARPi: poly(ADP-ribose) polymerase inhibitor; SOC: standard of care

Note: PLATO and CARD were phase IV trials. All remaining trials were phase III. The color ‘green’ represents a positive trial, ‘red’ represents a negative trial.

Cabazitaxel20: Cabazitaxel 20 mg/m^2^ IV on day 1 of every 3-week cycle

Cabazitaxel25: Cabazitaxel 25 mg/m^2^ IV on day 1 of every 3-week cycle

Docetaxel (2W): Docetaxel 75 mg/m^2^ IV on days 1 and 15 of a 4-week cycle

Docetaxel (int): Docetaxel 35 mg/m^2^ IV on days 1, 8, 15, repeat cycle at day 29

Docetaxel30 (W): Docetaxel 30 mg/m^2^ IV on days 1, 8, 15, 22 and 29 of a 6-week cycle

Remaining drugs were administered at standard doses.

### **Supplement Table 2:** Outcome definitions across included phase II trials

| **Trial** | **Arm** | **Progression Free Survival (PFS)** | **Overall Survival (OS)** | **Time to tumor progression (TTP)** | **Radiographic Objective Response Rate (ORR)** | **PSA 50% Response** |
| --- | --- | --- | --- | --- | --- | --- |
| **Prior ADT only** | | | | | | |
| **Antiandrogen+Chemotherapy** | | | | | | |
| **Takahashi M et al**, 2013 | Rx: Antiandrogen + Tegafur-Uracil Ctrl: Antiandrogen | Not reported | Not reported | Not reported | Not reported | The proportion of patients with ≥50% decrease in PSA levels from baseline. |
| **Antifungal monotherapy** | | | | | | |
| **Antonarakis ES et al**, 2013 | Rx: Itraconazole600 Ctrl: Itraconazole200 | The time from randomization to clinical progression (worsening disease-related symptoms, new complications), radiographic progression (per RECIST criteria, or ≥2 new bone lesions on bone scan), or death from any cause. | Not reported | Not reported | The proportion of patients achieving a complete response or partial response according to RECIST criteria. | The proportion of patients with ≥50% decrease in the PSA value from the baseline, maintained for ≥4 weeks. |
| **Antifungal+Bisphosphonate** | | | | | | |
| **Figg WD et al,** 2005 | Rx: Ketoconazole + Alendronate Ctrl: Ketoconazole | The time from study enrollment to date of progression, death or last follow-up. | The time from study enrollment to the date of death or loss to follow-up. | Not reported | The proportion of patients achieving a complete response or partial response according to RECIST criteria. | The proportion of patients with ≥50% decrease in the PSA value from the baseline, confirmed ≥3 weeks later. |
| **Antifungal+Chemotherapy** | | | | | | |
| **Millikan R et al**, 2001 | Rx1: Ketoconazole + Doxorubicin Rx2: Ketoconazole | Not reported | Reported but not defined | Reported but not defined | Not reported | The proportion of patients with ≥50% decrease in PSA levels from baseline. |
| **ARPI monotherapy** | | | | | | |
| **TERRAIN**, 2016 | Rx: Enzalutamide Ctrl: Bicalutamide | The time from randomization to progression (radiographic progression determined by independent central review per RECIST v1.1, skeletal-related event, or initiation of new antineoplastic therapy) or death from any cause. | Not reported | Not reported | The proportion of patients achieving a complete response or partial response according to RECIST v1.1 criteria. | The proportion of patients with ≥50% decrease in PSA levels from baseline. |
| **ARPI+ARPI** | | | | | | |
| **Khalaf DJ et al**, 2019 | Rx1: Abiraterone followed by Enzalutamide Rx2: Enzalutamide followed by Abiraterone | Not reported | The time from treatment initiation to the date of death or loss to follow-up. | The time from treatment initiation to confirmed PSA progression, radiographic progression (PCWG2 criteria), clinical progression, or death from prostate cancer, whichever occurred first. | Not reported | Not reported |
| **ARPI+Immunotherapy** | | | | | | |
| **STAMP**, 2015 | Rx1: Abiraterone + cSipuleucel-T Rx2: Abiraterone + sSipuleucel-T | Not reported | Reported but not defined | Not reported | Not reported | The proportion of patients with ≥50% decrease in the PSA value from the baseline. |
| **STRIDE**, 2023 | Rx1: Enzalutamide + cSipuleucel-T  Rx2: Enzalutamide + sSipuleucel-T | Not reported | Reported but not defined | Not reported | Not reported | Not reported |
| **ARPI+TKI** | | | | | | |
| **Dorff TB et al**, 2019 | Rx: Abiraterone  + Dasatinib Ctrl: Abiraterone | The time from treatment initiation to the first evidence of disease progression (per RECIST v1.1) or death from any cause. | The time from treatment initiation to the date of death or loss to follow-up. | Not reported | The proportion of patients achieving a complete response or partial response according to RECIST v1.1 criteria. | Not reported |
| **ASO monotherapy** | | | | | | |
| **Yu EY et al**, 2018 | Rx: Apatorsen Ctrl: Prednisone | The time from randomization to PSA and radiographic progression (per PCWG2 and RECIST v1.1 criteria),disease-related global or severe deterioration of health status requiring discontinuation of treatment without evidence of progression, the need for palliative radiation therapy, or death from any cause. | Not reported | The time from randomization to PSA and radiographic progression (per PCWG2 and RECIST v1.1 criteria),disease-related global or severe deterioration of health status requiring discontinuation of treatment without evidence of progression, or the need for palliative radiation therapy. | The proportion of patients achieving a complete response or partial response according to RECIST v1.1 criteria. | The proportion of patients with ≥50% decrease in PSA levels from baseline. |
| **Tolcher AW et al,**  2002 | Rx: ISIS 3521 Ctrl: ISIS 5132 | Not reported | Not reported | Not reported | Not reported | The proportion of patients with ≥50% decrease in the PSA value from the baseline, confirmed ≥4 weeks later. |
| **Chemotherapy monotherapy** | | | | | | |
| **Heidenreich A et al**, 2004 | Rx1: L-Doxorubicin25  Rx2: L-Doxorubicin50 | Not reported | The time from treatment initiation to the date of death or loss to follow-up. | The time from treatment initiation to first sign of disease progression (≥50% PSA increase, worsening performance status, or intensification of malignancy-related symptoms) or death. | Not reported | The proportion of patients with ≥50% decrease in the PSA value from the baseline, confirmed ≥4 weeks later. |
| **TIPC**, 2007 | Rx: Docetaxel30 (W) Ctrl: Prednisolone | The time from randomization to PSA progression, subjective progression (using PSPA score), change of systemic antineoplastic therapy, or death from any cause. | Reported but not defined | Not reported | Not reported | The proportion of patients with ≥50% decrease in the PSA value from the baseline, confirmed ≥4 weeks later. |
| **Krainer M et al**, 2007 | Rx: Docetaxel25 (W) Ctrl: Vinorelbine | Not reported | Not reported | The time from treatment initiation to disease progression, defined as a confirmed PSA increase of 50% (minimum increase of 1.0 ng/ml), progression in measurable disease or the appearance of new lesions. | Not reported | The proportion of patients with ≥50% decrease in PSA levels from baseline. |
| **Chemotherapy+Antifungal** | | | | | | |
| **Millikan R et al,** 2003 | Rx: Vin/Estra + Keto/Doxorubicin Ctrl: Paclitaxel + Estramustine + Etoposide | Not reported | Reported but not defined | Not reported | The proportion of patients achieving a complete response or partial response according to WHO criteria. | The proportion of patients with ≥50% decrease in the PSA value maintained for ≥8 weeks. |
| **Chemotherapy+ARPI** | | | | | | |
| **CHEIRON**, 2021 | Rx: Docetaxel + Enzalutamide Ctrl: Docetaxel | The time from randomization to first evidence of radiographic progression (PCWG2 criteria for bone lesions, RECIST v1.1 for soft tissue lesions), PSA progression or death from any cause. | The time from randomization to the date of death from any cause. | Not reported | The proportion of patients achieving a complete response or partial response according to RECIST criteria. | The proportion of patients with ≥50% decrease in PSA levels from baseline. |
| **Chemotherapy+ASO** | | | | | | |
| **Chi KN et al,** 2010 | Rx: Docetaxel + Custirsen Ctrl: Docetaxel | The time from randomization to progression or death from any cause. | The time from randomization to the date of death from any cause. | Not reported | The proportion of patients achieving a complete response or partial response according to RECIST criteria. | The proportion of patients with ≥50% decrease in PSA levels from baseline. |
| **Wiechno P et al**, 2014 | Rx: Docetaxel + LY2181308 Ctrl: Docetaxel | The time from randomization to disease progression (clinical symptoms, radiographic progression as per RECIST criteria, ≥2 new lesions on bone scan) or death from any cause. | Reported but not defined | Not reported | Not reported | The proportion of patients with ≥50% decrease in PSA levels from baseline. |
| **Chemotherapy+Bcl-2 inhibitor** | | | | | | |
| **EORTC**, 2009 | Rx: Docetaxel  + Oblimersen Ctrl: Docetaxel | Not reported | Not reported | The time from randomization to PSA progression, objective progression, or death due to progression. | The proportion of patients achieving a complete response or partial response according to RECIST criteria. | The proportion of patients with ≥50% decrease in the PSA value from the baseline, maintained for ≥4 weeks. |
| **Sonpavde G et al**, 2011 | Rx: Docetaxel + AT-101 Ctrl: Docetaxel | The time from randomization to soft tissue disease progression (per RECIST), bone scan progression (appearance of ≥2 new bone lesions), skeletal event, or death from any cause. | The time from randomization to the date of death from any cause. | Not reported | The proportion of patients achieving a complete response or partial response according to RECIST criteria. | The proportion of patients with ≥50% decrease in PSA levels from baseline. |
| **Chemotherapy+Chemotherapy** | | | | | | |
| **Machiels JP et al**, 2008 | Rx: Docetaxel35 (W) + Estramustine Ctrl: Docetaxel35 (W) | The time to randomization to PSA progression, tumor progression or death from any cause. | The time from randomization to the date of last follow-up or death from any cause. | Not reported | The proportion of patients achieving a complete response or partial response according to RECIST criteria. | The proportion of patients with ≥50% decrease in the PSA value from the baseline, confirmed ≥4 weeks later. |
| **Berry WR et al**, 2004 | Rx: Paclitaxel + Estramustine Ctrl: Paclitaxel | Not reported | The time from treatment initiation to the date of death or loss to follow-up. | Time from treatment initiation to the first sign of progressive disease or death from any cause. | Not reported | The proportion of patients with ≥50% decrease in the PSA value from the baseline, confirmed ≥4 weeks later. |
| **Albrecht W et al**, 2004 | Rx: Estramustine + Vinblastine Ctrl: Estramustine | Not reported | Reported but not defined | Not reported | Not reported | The proportion of patients with ≥50% decrease in the PSA value from the baseline, confirmed ≥4 weeks later. |
| **Cabrespine A et al**, 2006 | Rx: Paclitaxel + Carboplatin Ctrl: Mitoxantrone | Not reported | The time from study enrollment to the date of death or loss to follow-up. | Not reported | The proportion of patients achieving a complete response or partial response according to WHO criteria. | The proportion of patients with ≥50% decrease in the PSA value from the baseline, confirmed ≥4 weeks later. |
| **Nelius T et al**, 2006 | Rx1: Docetaxel70 + Estramustine840 Rx2: Docetaxel70 + Estramustine420 | Not reported | The time from treatment initiation to the date of death or loss to follow-up. | Not reported | Not reported | The proportion of patients with ≥50% decrease in PSA levels from baseline. |
| **Oudard S et al**, 2005 | Rx1: Docetaxel70 + Estramustine Rx2: Docetaxel35  (D2,D9 Q3W) + Estramustine Ctrl: Mitoxantrone | Not reported | The time from study enrollment to the date of death or loss to follow-up. | Not reported | The proportion of patients achieving a complete response or partial response according to WHO criteria. | The proportion of patients with ≥50% decrease in the PSA value from the baseline, confirmed ≥3 weeks later. |
| **Droz JP et al**, 2003 | Rx1: Oxaliplatin + 5-Fluorouracil Rx2: Oxaliplatin | Not reported | The time from treatment initiation to the date of death. | The time from treatment initiation to the first objective evidence of tumor progression, last contact or start of further antitumor therapy. | Not reported | The proportion of patients with ≥50% decrease in PSA levels from baseline. |
| **Galsky MD et al**, 2005 | Rx1: Ixabepilone + Estramustine Rx2: Ixabepilone | Not reported | Not reported | Not reported | The proportion of patients achieving a complete response or partial response according to RECIST criteria. | The proportion of patients with ≥50% decrease in PSA levels from baseline, confirmed by 3 successive evaluations ≥2 weeks apart. |
| **Eymard JC et al**, 2007 | Rx: Docetaxel70 + Estramustine Ctrl: Docetaxel | Not reported | Reported but not defined | Reported but not defined | The proportion of patients achieving a complete response or partial response according to WHO criteria. | The proportion of patients with ≥50% decrease in the PSA value from the baseline, confirmed ≥3 weeks later. |
| **Chemotherapy+Curcuminoid** | | | | | | |
| **Jahanmohan JP et al**, 2021 | Rx: Docetaxel + Curcumin Ctrl: Docetaxel | Not reported | The time from randomization to the date of death from any cause. | The time from randomization to the first objective evidence of progression, defined as either a PSA increase of ≥25% with an absolute increase of 2 ng/mL confirmed at two consecutive visits at least two weeks apart, radiographic progression according to RECIST v1.1 criteria, or documentation of two or more new bone lesions on a bone scan. | Not reported | The proportion of patients with ≥50% decrease in PSA levels from baseline. |
| **Chemotherapy+Immunotherapy** | | | | | | |
| **Dahut WL et al**, 2004 | Rx: Docetaxel30 (W) ^a^ + Thalidomide Ctrl: Docetaxel30 (W) ^a^ | The time from randomization to progression or death from any cause. Progression was defined as >25% size increase of all soft tissue masses and/or presence of new lesions, need for radiation, ≥2 consecutive PSA increases ≥50% for patients with PSA response, or ≥25% for patients without PSA response. | The time from randomization to the date of death from any cause. | Not reported | The proportion of patients achieving a complete response or partial response according to RECIST criteria. | The proportion of patients with ≥50% decrease in the PSA value from the baseline, confirmed ≥4 weeks later. |
| **ECOG 3899**, 2010 | Rx1: 13-cis Retinoic acid + Interferon-alpha2b + Paclitaxel Rx2: Vinorelbine + Mitoxantrone + Estramustine | The time from study enrollment to progression (measurable disease, bone or PSA), death or loss to follow-up. | The time from study enrollment to the date of death or loss to follow-up. | Not reported | Not reported | The proportion of patients with ≥50% decrease in the PSA value from the baseline, confirmed ≥1 week later. |
| **E1809**, 2015 | Rx: Docetaxel + PVAC-VF Ctrl: Docetaxel | Not reported | The time from randomization to the date of death from any cause. | The time from the date of treatment initiation to radiographic progression. | The proportion of patients achieving a complete response or partial response according to RECIST v1.1 criteria. | The proportion of patients with ≥50% decrease in PSA levels from baseline. |
| **de Bono et al**, 2014 | Rx: Docetaxel + Figitumumab Ctrl: Docetaxel | The time from randomization to first event of disease progression, which was defined as one or more of the following: confirmed PSA progression; 2 new bone lesions; progressive disease according to RECIST; increased pain requiring one or more of the following: narcotics for >2 weeks, radiation therapy, doubling the corticosteroid dose, radionuclide therapy, or palliative chemotherapy; intervention for any prostate cancer-related events (e.g., radiation, surgery); new symptoms related to tumor growth; or death because of any cause. | Not reported | Not reported | Not reported | The proportion of patients with ≥50% decrease in the PSA value from the baseline, confirmed ≥3 weeks later. |
| **Heidenreich A et al,**  2013 | Rx: Docetaxel  + Intetumumab Ctrl: Docetaxel | The time from randomization to progression (radiographic or clinical), or death from any cause. | Reported but not defined | Not reported | Not reported | The proportion of patients with ≥50% decrease in the PSA value from the baseline, confirmed ≥6 weeks later. |
| **Chemotherapy+Radiopharmaceutical** | | | | | | |
| **Taxium II**, 2017 | Rx: Docetaxel  + Rhenium-188-HEDP Ctrl: Docetaxel | The time from randomization to progression (PSA/radiographic/clinical) or death from any cause. | Reported but not defined | Not reported | The proportion of patients achieving a complete response or partial response according to RECIST v1.1 criteria. | The proportion of patients with ≥50% decrease in PSA levels from baseline. |
| **Chemotherapy+TKI** | | | | | | |
| **Horti J et al**, 2009 | Rx: Docetaxel + Vandetanib Ctrl: Docetaxel | Not reported | Not reported | Time from randomization to objective disease progression or death from any cause. | The proportion of patients achieving a complete response or partial response according to RECIST criteria. | The proportion of patients with ≥50% decrease in the PSA value from the baseline, confirmed 2-4 weeks later. |
| **Chemotherapy+Vascular disrupting agent** | | | | | | |
| **Pili R et al,**  2010 | Rx: Docetaxel + Vadimezan Ctrl: Docetaxel | Not reported | The time from treatment initiation to the date of death from any cause. | The time from treatment initiation to first objective documentation of progression. | The proportion of patients achieving a complete response or partial response according to RECIST criteria. | The proportion of patients with ≥50% decrease in the PSA value from the baseline, confirmed ≥4 weeks later. |
| **Chemotherapy+VitD-Analog** | | | | | | |
| **Attia S et al**, 2008 | Rx: Docetaxel35 (W) ^b^ + Doxercalciferol Ctrl: Docetaxel35 (W) ^b^ | Reported but not defined | Reported but not defined | Not reported | The proportion of patients achieving a complete response or partial response according to WHO criteria. | The proportion of patients with ≥50% decrease in the PSA value from the baseline, confirmed ≥4 weeks later. |
| **ASCENT**, 2007 | Rx: Docetaxel36 (W) + Calcitriol Ctrl: Docetaxel36 (W) | The time from randomization to either radiographic progression (per RECIST), skeletal-related event, or death from any cause. | Reported but not defined | Not reported | The proportion of patients achieving a complete response or partial response according to RECIST criteria. | The proportion of patients with ≥50% decrease in the PSA value from the baseline, confirmed ≥4 weeks later. |
| **Corticosteroid monotherapy** | | | | | | |
| **Venkitaraman R et al**, 2015 | Rx: Dexamethasone Ctrl: Prednisolone | Not reported | Not reported | Not reported | The proportion of patients achieving a complete response or partial response according to RECIST criteria. | The proportion of patients with ≥50% decrease in the PSA value from the baseline, confirmed ≥4 weeks later. |
| **Corticosteroid+Immunotherapy** | | | | | | |
| **Yoshimura K et al**, 2016 | Rx: Dexamethasone + PPV Ctrl: Dexamethasone | Not reported | The time from treatment initiation to the date of death. | Not reported | Not reported | The proportion of patients with ≥50% decrease in PSA levels from baseline at 12 weeks. |
| **Corticosteroid+VEGFi** | | | | | | |
| **Stadler WM et al**, 2004 | Rx: Dexamethasone + SU5416 Ctrl: Dexamethasone | Not reported | Not reported | The time from randomization to a single rise in PSA >25% over the nadir, new bone lesion or evidence of progressive disease (per RECIST criteria). | Not reported | Not reported |
| **ERA monotherapy** | | | | | | |
| **James ND et al**, 2009 | Rx1: Zibotentan15  Rx2: Zibotentan10 Ctrl: Placebo | Not reported | The time from randomization to the date of death from any cause. | The time from randomization to first occurrence of clinical progression (symptoms requiring chemotherapy, radiotherapy, surgery, or initiation of new therapy), cancer pain (requiring opiates), radiographic progression of soft tissue (as per RECIST) or death from any cause. | The proportion of patients achieving a complete response or partial response according to RECIST criteria. | Not reported |
| **Carducci MA et al**, 2003 | Rx1: Atrasentan2.5 Rx2: Atrasentan10 Ctrl: Placebo | Not reported | Not reported | The time from randomization to the development of new bone/ soft tissue lesions, requirement of palliative treatment with an opioid analgesic for new disease-related pain, the occurrence of new disease-related symptoms requiring intervention, clinical events (determined as progression by investigator), or death from any cause. | Not reported | Not reported |
| **IMiD monotherapy** | | | | | | |
| **Pili R et al**, 2011 | Rx: Tasquinimod Ctrl: Placebo | The time from randomization to the first occurrence of either symptomatic progression, radiologic progression (per RECIST or PCWG2 guidelines), or death from any cause. | Reported but not defined | Not reported | The proportion of patients achieving a complete response or partial response according to RECIST criteria. | The proportion of patients with ≥50% decrease in PSA levels from baseline. |
| **Immunotherapy monotherapy** | | | | | | |
| **PERSEUS**, 2016 | Rx1: Abituzumab750 + SOC Rx2: Abituzumab1500 + SOC Ctrl: SOC | The time from randomization to first documented radiographic progression (appearance of 2 new bone lesions on scintigraphy, confirmed after 6 weeks if asymptomatic or mildly symptomatic; soft-tissue lesion progression according to RECIST 1.0 modified according to PCWG2; or presence of skeletal events defined as cord compression or fracture) or death from any cause within 12 weeks following the last tumor assessment. | The time from randomization to the date of death from any cause. | The time from the date of randomization to the date of objective radiographic progression. | The proportion of patients achieving a complete response or partial response, as per RECIST criteria modified according to PCWG2. | The proportion of patients with ≥50% decrease in the PSA value from the baseline, confirmed ≥3 weeks later. |
| **Figg WD et al**, 2001 | Rx: ThalidomideHD Ctrl: Thalidomide | Not reported | Not reported | Not reported | The proportion of patients achieving a complete response or partial response according to standard objective criteria. Standard objective criteria were used to assess soft tissue lesion changes. Disappearance of > 50% of the number of metastatic lesions on bone scan was also considered a partial response. | The proportion of patients with ≥50% decrease in PSA levels from baseline. |
| **Immunotherapy+Chemotherapy** | | | | | | |
| **Arlen PM et al,** 2006 | Rx: Vaccine + GMCSF + Docetaxel Ctrl: Vaccine + GMCSF | The time from randomization to progression (radiographic/PSA) or death from any cause. | Not reported | Not reported | Not reported | The proportion of patients with ≥50% decrease in PSA levels from baseline. |
| **Immunotherapy+GMCSF** | | | | | | |
| **TBC-PRO-002**, 2010 | Rx: PVAC+GMCSF Ctrl: Placebo | The time from randomization to the identification of ≥2 new sites of bone metastasis on bone scan, >20% increase in lymph node metastasis (according to RECIST), clinical signs/ symptoms of progression (as per investigator) or death from any cause. | Reported but not defined | Not reported | The proportion of patients achieving a complete response or partial response according to RECIST criteria. | Not reported |
| **Integrin inhibitor monotherapy** | | | | | | |
| **Bradley DA et al,** 2011 | Rx: Cilengitide500 Ctrl: Cilengitide2000 | Not reported | Not reported | The time from treatment initiation to progression (measurable disease per RECIST criteria, development of ≥2 new lesions on bone scan, pain progression requiring opioid therapy and/or radionuclide/radiation treatment, or death from any cause. | The proportion of patients achieving a complete response or partial response according to RECIST criteria. | The proportion of patients with ≥50% decrease in the PSA value from the baseline, confirmed ≥3 weeks later. |
| **Matrix Metalloproteinase Inhibitor monotherapy** | | | | | | |
| **Lara Jr PN et al**, 2006 | Rx: BMS-275291 [1200mg] Ctrl: BMS-275291 [2400mg] | Not reported | Reported but not defined | Not reported | Not reported | Not reported |
| **Progestin monotherapy** | | | | | | |
| **Patel SR et al**, 1990 | Rx: Megestrol Acetate Ctrl: Dexamethasone | Not reported | Reported but not defined | Not reported | The proportion of patients achieving a complete response or partial response according to WHO criteria. | Not reported |
| **Somatostatin+Corticosteroid** | | | | | | |
| **Dimopoulos MA et al**, 2004 | Rx: Lanreotide + Dexamethasone Ctrl: Estramustine + Etoposide | Not reported | The time from study enrollment to the date of death. | The time from study enrollment to disease progression or death. | The proportion of patients achieving a complete response or partial response according to WHO criteria. | The proportion of patients with ≥50% decrease in PSA levels from baseline. |
| **TKI monotherapy** | | | | | | |
| **Boccardo F et al**, 2008 | Rx: Gefitinib Ctrl: Placebo | Reported but not defined | Reported but not defined | Not reported | The proportion of patients achieving a complete response or partial response according to WHO criteria. | The proportion of patients with ≥50% decrease in the PSA value from the baseline, confirmed ≥4 weeks later. |
| **TKI+1st gen AA** | | | | | | |
| **Azad AA et al**, 2014 | Rx: Vandetanib + Bicalutamide Ctrl: Bicalutamide | Not reported | Not reported | Not reported | The proportion of patients achieving a complete response or partial response according to RECIST criteria. | The proportion of patients with ≥50% decrease in the PSA value from the baseline, confirmed ≥4 weeks later. |
| **Sridhar SS et al**, 2015 | Rx1: Pazopanib + Bicalutamide Rx2: Pazopanib | The time from treatment initiation to disease progression or death from any cause. | Not reported | Not reported | The proportion of patients achieving a complete response or partial response according to RECIST criteria. | The proportion of patients with ≥50% decrease in the PSA value from the baseline, confirmed ≥3 weeks later. |
| **Prior ADT+ARPI** | | | | | | |
| **ARPI+TKI** | | | | | | |
| **Spetsieris N et al**, 2021 | Rx: Abiraterone + Sunitinib Ctrl: Abiraterone + Dasatinib | Not reported | Reported but not defined | Not reported | Not reported | Not reported |
| **Bipolar Androgen Therapy** | | | | | | |
| **TRANSFORMER**, 2021 | Rx: BAT Ctrl: Enzalutamide | The time from the date of randomization to earliest sign of radiographic progression (soft tissue lesion progression according to RECIST v1.1, or bone lesion progression according to PCWG2 criteria), development of symptoms, initiation of another anticancer treatment or death from any cause. | The time from randomization to the date of death from any cause. | Not reported | The proportion of patients achieving a complete response or partial response according to RECIST and PCWG2 criteria. | The proportion of patients with ≥50% decrease in PSA levels from baseline, according to PCWG2 criteria. |
| **Chemotherapy+ARPI** | | | | | | |
| **ABIDO SOGUG**, 2022 | Rx: Docetaxel + Abiraterone Ctrl: Docetaxel | The time from randomization to radiographic progression (per PCWG2 and modified RECIST criteria) or death from any cause. | The time from randomization to the date of death from any cause. | Not reported | The proportion of patients achieving a complete response or partial response according to RECIST criteria. | The proportion of patients with ≥50% decrease in PSA levels from baseline. |
| **Chemotherapy+Chemotherapy** | | | | | | |
| **MDACC Study**, 2019 | Rx: Cabazitaxel25 + Carboplatin Ctrl: Cabazitaxel25 | The time from treatment initiation to disease progression or death from any cause. Disease progression was defined as progression of measurable disease by RECIST criteria, ≥2 new areas by bone scan attributable to prostate cancer (rather than flare) or new or increasing size of lytic lesions by CT scan/MRI, need for palliative radiotherapy, surgery/kyphoplasty to any neoplastic bone lesion, cancer-associated clinical deterioration, or receipt of any additional prostate cancer-specific therapy as prescribed by the treating physician. | The time from study enrollment to the date of death. | Not reported | Not reported | The proportion of patients with ≥50% decrease in PSA levels from baseline. |
| **Prior ADT+Docetaxel** | | | | | | |
| **ASO+Chemotherapy** | | | | | | |
| **P-06c,**  2011 | Rx1: Custirsen + Docetaxel Rx2: Custirsen  + Mitoxantrone | The time from treatment initiation to the first documentation of disease progression (radiographic progression per RECIST, pain progression or deterioration of performance status) or death from any cause. | The time from treatment initiation to the date of death from any cause. | Not reported | The proportion of patients achieving a complete response or partial response according to RECIST criteria. | The proportion of patients with ≥50% decrease in PSA levels from baseline, confirmed on ≥2 consecutive measurements 4-6 weeks apart. |
| **Aurora kinase inhibitor monotherapy** | | | | | | |
| **Meulenbeld HJ et al**, 2012 | Rx1: Danusertib330 Rx2: Danusertib500 | Reported but not defined | Not reported | Not reported | The proportion of patients achieving a complete response or partial response according to RECIST criteria. | The proportion of patients with ≥50% decrease in the PSA value from the baseline, confirmed ≥4 weeks later. |
| **Chemotherapy monotherapy** | | | | | | |
| **GETUG-P02,**  2015 | Rx1: Etoposide Rx2: Vinorelbine Rx3: Mitoxantrone | Reported but not defined | Reported but not defined | Not reported | The proportion of patients achieving a complete response or partial response according to RECIST criteria. | The proportion of patients with ≥50% decrease in the PSA value from the baseline, confirmed ≥3 weeks later. |
| **CAINTA**, 2023 | Rx: Cabazitaxel ID Ctrl: Cabazitaxel25 | The time from study enrollment to disease progression (per RECIST v1.1 criteria) or death from any cause. | Reported but not defined | Not reported | The proportion of patients achieving a complete response or partial response according to RECIST v1.1 criteria. | The proportion of patients with ≥50% decrease in the PSA value from the baseline in patients with a baseline PSA ≥20 mg/L by week 12. |
| **Rosenberg JE et al**, 2007 | Rx: Ixabepilone Ctrl: Mitoxantrone | Not reported | Reported but not defined | Not reported | The proportion of patients achieving a complete response or partial response according to RECIST criteria. | The proportion of patients with ≥50% decrease in PSA levels from baseline, confirmed on 2 consecutive measurements. |
| **Chemotherapy+Chemotherapy** | | | | | | |
| **RECARDO**, 2018 | Rx: Docetaxel + Carboplatin Ctrl: Docetaxel | The time from treatment initiation to disease progression, death or the off-study date. Disease progression was defined as a (confirmed) PSA increase over the baseline value (50% increase for patients with, an 25% increase for patients without an initial PSA response since the start of the treatment, with a minimum rise of 5 ng/ml) and/or progression on imaging according to the RECIST criteria. | The time from treatment initiation to the date of death. | Not reported | The proportion of patients achieving a complete response or partial response according to RECIST criteria. | The proportion of patients with ≥50% decrease in the PSA value from the baseline, confirmed ≥4 weeks later. |
| **Chemotherapy+Immunotherapy** | | | | | | |
| **Fizazi K et al**, 2012 | Rx: Mitoxantrone + Siltuximab Ctrl: Mitoxantrone | The time from the date of randomization to progressive disease (soft tissue progression per RECIST criteria, ≥3 new bone lesions confirmed by a second bone scan or clinical deterioration due to pain requiring palliative intervention, skeletal-related events, or other disease-related events requiring intervention) or death from any cause. | The time from randomization to the date of death from any cause. | Not reported | Not reported | The proportion of patients with ≥50% decrease in the PSA value from the baseline, confirmed ≥3 weeks later. |
| **Hakenberg OW et al**, 2019 | Rx: Mitoxantrone  + Olaratumab Ctrl: Mitoxantrone | The time from randomization to the first occurrence of radiographic progression (per RECIST v1.1), progression on bone scan, clinical progression, or death from any cause. | The time from randomization to the date of death from any cause. | Not reported | The proportion of patients achieving a complete response or partial response according to RECIST v1.1 criteria. | The proportion of patients with ≥50% decrease in the PSA value from the baseline, confirmed ≥3 weeks later. |
| **Immunotherapy monotherapy** | | | | | | |
| **Filaci G et al**, 2021 | Rx1: GX301 vaccine (8D) Rx2: GX301 vaccine (4D) Rx3: GX301 vaccine (2D) | Reported but not defined | Reported but not defined | Not reported | Not reported | Not reported |
| **TKI monotherapy** | | | | | | |
| **Droz JP et al**, 2014 | Rx: Nintedanib150 Ctrl: Nintedanib250 | The time from treatment initiation to the first occurrence of tumor progression (increase in mean PPI score for pain, analgesic score, analgesic radiotherapy, or radiologic progression with new/progressive bone lesions) or death from any cause. | Reported but not defined | Not reported | The proportion of patients achieving a complete response or partial response according to RECIST criteria. | The proportion of patients with ≥50% decrease in the PSA value from the baseline, confirmed ≥4 weeks later. |
| **Prior ADT+Docetaxel+ARPI** | | | | | | |
| **ARPI monotherapy** | | | | | | |
| **SAK-08-16**, 2023 | Rx: Darolutamide Ctrl: Placebo | The time from randomization to radiographic progression (per PCWG3 and RECIST 1.1 criteria) or death from any cause. | The time from treatment initiation to the date of death from any cause. | Not reported | Not reported | The proportion of patients with ≥50% decrease in PSA levels from baseline. |
| **Chemotherapy monotherapy** | | | | | | |
| **ConCab**, 2018 | Rx: Cabazitaxel (W) Ctrl: Cabazitaxel25 | The time from randomization the first documentation of PSA progression, pain progression, radiologic progression (per RECIST v1.1 or PCWG2 criteria), or death from any cause. | The time from randomization to the date of death from any cause. | Not reported | The proportion of patients achieving a complete response or partial response according to RECIST v1.1 criteria. | The proportion of patients with ≥50% decrease in PSA levels from baseline, given that the baseline PSA is at least 10 ng/ml. |
| **Chemotherapy+AKTi** | | | | | | |
| **RE-AKT**, 2024 | Rx: Enzalutamide  + Capivasertib Ctrl: Enzalutamide | The time from randomization to first RECIST 1.1 progression, bone scan progression defined by PCWG2, or death from any cause. | The time from randomization to the date of death from any cause. | Not reported | The proportion of patients achieving a complete response or partial response according to RECIST v1.1 criteria. | The proportion of patients with ≥50% decrease in PSA levels from baseline. |
| **Radioligand monotherapy** | | | | | | |
| **TheraP**, 2021 | Rx: 177Lu Ctrl: Cabazitaxel20 | The time from randomization to first evidence of PSA progression (per PCWG3 criteria), radiographic progression (per RECIST 1.1 and PCWG3 criteria), commencement of non-protocol anticancer treatment, or death from any cause. | The time from study enrollment to the date of death or loss to follow-up. | Not reported | The proportion of patients achieving a complete response or partial response according to RECIST v1.1 criteria. | The proportion of patients with ≥50% decrease in PSA levels from baseline. |
| **Heterogeneous Prior Therapy** | | | | | | |
| **ARPI+Radioligand** | | | | | | |
| **ENZA-p**, 2024 | Rx: Enzalutamide + 177Lu Ctrl: Enzalutamide | The time from randomization to first evidence of radiographic progression (PCWG3 criteria for bone lesions, RECIST v1.1 for soft tissue lesions), or the date of last known follow-up without progression. | The time from randomization to the date of death from any cause or loss to follow-up. | Not reported | Not reported | The proportion of patients with ≥50% decrease in the PSA value from the baseline. |
| **Chemotherapy monotherapy** | | | | | | |
| **TAXYNERGY**, 2017 | Rx: Docetaxel Ctrl: Cabazitaxel25 | The time from randomization to the first occurrence of either radiographic (per RECIST v1.1 criteria), PSA, or clinical (skeletal-related events, increasing pain requiring increased narcotic analgesics, urinary obstruction) progression or death from any cause. | The time from randomization to the date of death from any cause. | Not reported | Not reported | The proportion of patients with ≥50% decrease in the PSA value from the baseline, confirmed ≥3 weeks later. |
| **Chemotherapy+Immunotherapy** | | | | | | |
| **IND 209**, 2018 | Rx: Docetaxel  + Reolysin Ctrl: Docetaxel | Not reported | The time from randomization to the date of last follow-up or death from any cause. | Not reported | The proportion of patients achieving a complete response or partial response according to RECIST criteria. | Not reported |
| **Kongsted P et al,**  2017 | Rx: Docetaxel  + DCVAC Ctrl: Docetaxel | The time from treatment initiation to clinical, radiographic and/or PSA progression or death. Progression was defined per PCWG2, with a modification that a confirmed ≥25% increase (≥2 ng/mL) above baseline or nadir PSA levels was required to fulfill the criterion for biochemical progression in patients with or without a PSA decline following four cycles of docetaxel. | Not reported | Not reported | The proportion of patients achieving a complete response or partial response according to RECIST v1.1 criteria. | The proportion of patients with ≥50% decrease in PSA levels from baseline. |
| **PARPi+ARPi** | | | | | | |
| **BRCAAway**, 2024 | Rx1: Abiraterone Rx2: Olaparib Rx3: Abiraterone  + Olaparib | The time from randomization to radiographic progression (as per RECIST v1.1, PCWG3), clinical assessment (disease-related symptoms requiring intervention) or death from any cause. | Not reported | Not reported | The proportion of patients achieving a complete response or partial response according to RECIST v1.1 criteria. | The proportion of patients with ≥50% decrease in the PSA value from the baseline, confirmed ≥4 weeks later. |
| **TKI monotherapy** | | | | | | |
| **Monk P et al**, 2018 | Rx: Tivantinib Ctrl: Placebo | The time from treatment initiation to the date of documented progression (per PCWG2 and RECIST v1.1 criteria), investigator-determined clinical deterioration, or death from any cause. | Not reported | Not reported | The proportion of patients achieving a complete response or partial response according to RECIST criteria. | Not reported |
| **Smith DC et al**, 2013 | Rx: Cabozantinib Ctrl: Placebo | The time from randomization to radiographic progression (per RECIST criteria) or death from any cause. | Not reported | Not reported | The proportion of patients achieving a complete response or partial response according to RECIST criteria. | Not reported |
| **Multiple subgroups (ADT/ADT+ARPI/ADT+Docetaxel/ADT+ARPI+Docetaxel/Heterogeneous prior therapy)** | | | | | | |
| **Chemotherapy monotherapy** | | | | | | |
| **Annala M et al**, 2021 | Rx: Cabazitaxel25 Ctrl: Enzalutamide /Abiraterone | The time from randomization to the first documented evidence of any progression (PSA, radiographic, or clinical) or death from any cause. | The time from randomization to the date of death from any cause. | The time from randomization to the first documented evidence of any progression (PSA, radiographic, or clinical). | Not reported | The proportion of patients with ≥50% decrease in PSA levels from baseline. |
| **Chemotherapy+AKTi** | | | | | | |
| **ProCAID**, 2021 | Rx: Docetaxel + Capivasertib Ctrl: Docetaxel | The time from the date of randomization to disease progression (PSA or bone metastases progression per PCWG2, soft tissue disease progression per RECIST v1.1, clinical progression, or commencement of new cancer-therapy) or death from any cause. | The time from randomization to the date of death from any cause. | Not reported | Not reported | Evaluated according to PCWG2 criteria. |

Abbreviations: ADT: androgen deprivation therapy; PSA: prostate-specific antigen; RECIST: response evaluation criteria in solid tumors; PCWG: prostate cancer working group; ARPI: androgen-receptor pathway inhibitor; cSipuleucel-T: concurrent sipuleucel-T; sSipuleucel-T: sequential sipuleucel-T; TKI: tyrosine kinase inhibitor; ASO: antisense oligonucleotide; Bcl-2: B-cell lymphoma 2 protein; PSPA: performance status/pain/analgesics score; WHO: world health organization; PVAC: PROSTVAC (viral vector-based immunotherapy); VEGFi: vascular endothelial growth factor inhibitor; VitD: vitamin d; IMiD: immunomodulatory drug; ERA: endothelin receptor antagonist; GMCSF: granulocyte-macrophage colony-stimulating factor; AA: antiandrogen; BAT: bipolar androgen therapy; PPI: present pain intensity score; 177Lu: 177Lu-PSMA-617; SOC: standard of care; PARPi: poly(ADP-ribose) polymerase inhibitor; AKTi: protein kinase B inhibitor; Vin/Estra: vinblastine/estramustine; keto: ketoconazole

The color ‘green’ represents a positive trial, ‘red’ represents a negative trial.

Itraconazole600: Itraconazole 600mg/day PO

Itraconazole200: Itraconazole 200mg/day PO

L-Doxorubicin25: Liposomal Doxorubicin (dissolved in 250 mL 5% glucose) 25 mg/m^2^ IV every 2 weeks for 12 consecutive cycles

L-Doxorubicin50: Liposomal Doxorubicin (dissolved in 250 mL 5% glucose) 50 mg/m^2^ IV every 4 weeks for 6 consecutive cycles

Docetaxel30 (W): Docetaxel 30 mg/m^2^ IV every week for 5 weeks, followed by 1 week rest (6-week cycle)

Docetaxel25 (W): Docetaxel 25 mg/m^2^ IV every week for 2 cycles (8-week cycle)

Docetaxel35 (W): Docetaxel 35 mg/m^2^ IV on days 2 and 9 every 3 weeks

Docetaxel70: Docetaxel 70 mg/m^2^ on day 2 every 3 weeks

Estramustine840: Estramustine 840 mg/day on days 1-5

Estramustine420: Estramustine 420 mg/day on days 1-3

Docetaxel35 (D2,D9 Q3W): Docetaxel 35 mg/m^2^ on day 2 and 9 every 3wks

Docetaxel30 (W) ^a^ : Docetaxel 30 mg/m^2^ IV every week for 3 weeks, followed by 1 week rest (4-week cycle)

Docetaxel35 (W) ^b^ : Docetaxel 35 mg/m^2^ IV on days 1, 8, and 15 (4-week cycle)

Docetaxel36 (W): Docetaxel 36 mg/m^2^ IV every week for 3 weeks (4-week cycle)

Zibotentan15: Zibotentan 15 mg PO OD

Zibotentan10: Zibotentan 10 mg PO OD

Atrasentan2.5: Atrasentan 2.5 mg PO OD

Atrasentan10: Atrasentan 10 mg PO OD

Abituzumab750: Abituzumab 750 mg every 3 weeks

Abituzumab1500: Abituzumab 1500 mg every 3 weeks

ThalidomideHD: Thalidomide initial dose of 200 mg/day with increments of 200 mg/day every 2 weeks to a maximum dose of 1200 mg

Cilengitide500: Cilengitide 500 mg IV twice weekly (six-week cycle)

Cilengitide2000: Cilengitide 2000 mg IV twice weekly (six-week cycle)

Cabazitaxel25: Cabazitaxel 25 mg/m^2^ IV every 3 weeks

Danusertib330: Danusertib 330 mg/m^2^ IV on days 1,8 and 15 every 4 weeks

Danusertib500: Danusertib 500 mg/m^2^ IV on days 1 and 15 every 4 weeks

Cabazitaxel ID: Cabazitaxel at an initial dose of 25 mg/m^2^ followed by dose adaptations in cycle 2 and beyond according to a prespecified dosing algorithm considering previous-cycle hematologic toxicity and cabazitaxel AUC (with a target AUC of 0.8–1.2 mg*hour/L)

GX301 vaccine (8D): GX301, 8 doses on days 1, 3, 5, 7, 14, 21, 35 and 63

GX301 vaccine (4D): GX301, 4 doses on days 1, 14, 35 and 63

GX301 vaccine (2D): GX301, 2 doses on days 1 and 63

Nintedanib150: Nintedanib 150 mg PO BID for 6 months

Nintedanib250: Nintedanib 250 mg PO BID for 6 months

Cabazitaxel (W): Cabazitaxel 25 mg/m^2^ IV weekly for 5 weeks (6-week cycle)

Cabazitaxel20: Cabazitaxel 20 mg/m^2^ IV every 3 weeks

Remaining drugs were administered at standard doses.

### **Supplement Table 3:** Distribution of race and ethnicity across the included phase III trials

| **Trial** | **Arm** | **Race \| N (%)** | | | | | **Ethnicity \| N (%)** | |
| --- | --- | --- | --- | --- | --- | --- | --- | --- |
|  |  | **Asian** | **American Indian or Alaskan Native** | **Native Hawaiian/ Pacific Islander** | **Black or African American** | **White** | **Hispanic/Latino** | **Non-Hispanic/Non-Latino** |
| **Prior ADT only** | | | | | | | | |
| **ARPI monotherapy** | | | | | | | | |
| **COU-AA-302**, 2013 | Rx: Abiraterone Ctrl: Placebo | NA | NA | NA | NA | NA | NA | NA |
| **PREVAIL**, 2014 | Rx: Enzalutamide Ctrl: Placebo | 167 (10) | 1 ( <0.1) | 2 (0.1) | 34 (2) | 1324 (77) | 38 (2) | 1527 (89) |
| **ELM-PC 4**, 2015 | Rx: TAK-700 Ctrl: Placebo | NA | NA | NA | NA | NA | NA | NA |
| **ARPI+ARPI** | | | | | | | | |
| **ACIS**, 2021 | Rx: Abiraterone  + Apalutamide Ctrl: Abiraterone | 111 (11) | 17 (2) | NA | 37 (4) | 738 (75) | 104 (11) | 833 (85) |
| **AllianceA031201**, 2023 | Rx: Enzalutamide + Abiraterone Ctrl: Enzalutamide | NA | NA | NA | 162 (12) | 1088 (83) | NA | NA |
| **ARPI+PI3K/AKTi** | | | | | | | | |
| **IPATential150**, 2021 | Rx: Abiraterone + Ipatasertib Ctrl: Abiraterone | 219 (20) | NA | NA | NA | 762 (69) | NA | NA |
| **ARPI+Radiopharmaceutical** | | | | | | | | |
| **ERA 223**, 2019 | Rx: Abiraterone + Radium-223 Ctrl: Abiraterone | 157 (19) | 2 (<1) | NA | 26 (3) | 569 (71) | NA | NA |
| **Chemotherapy monotherapy** | | | | | | | | |
| **CALGB9182**, 1999 | Rx: Mitoxantrone + Hydrocortisone Ctrl: Hydrocortisone | NA | NA | NA | NA | 219 (90) | NA | NA |
| **FIRSTANA**, 2017 | Rx1: Cabazitaxel20 Rx2: Cabazitaxel25 Ctrl: Docetaxel | 47 (4) | NA | NA | 23 (2) | 1088 (93) | NA | NA |
| **Kellokumpu-Lehtinen PL et al**, 2013 | Rx: Docetaxel (2W) Ctrl: Docetaxel | NA | NA | NA | NA | NA | NA | NA |
| **Berry W et al**, 2002 | Rx: Mitoxantrone Ctrl: Placebo | NA | NA | NA | 10 (8) | 105 (89) | 4 (3) | 115 (97) |
| **Abratt RP et al**, 2004 | Rx: Vinorelbine + Hydrocortisone Ctrl: Hydrocortisone | NA | NA | NA | NA | NA | NA | NA |
| **PRINCE**, 2018 | Rx: Docetaxel (int) Ctrl: Docetaxel | NA | NA | NA | NA | NA | NA | NA |
| **TAX327**, 2004 | Rx1: Docetaxel Rx2: Docetaxel30(W) Ctrl: Mitoxantrone | NA | NA | NA | NA | NA | NA | NA |
| **Chemotherapy+ASO** | | | | | | | | |
| **SYNERGY**, 2017 | Rx: Docetaxel + Custirsen Ctrl: Docetaxel | NA | NA | NA | NA | NA | NA | NA |
| **Chemotherapy+Chemotherapy** | | | | | | | | |
| **SWOG-99-16**, 2004 | Rx: Docetaxel60 + Estramustine Ctrl: Mitoxantrone | 1 (<1) | NA | NA | 104 (14) | 647 (84) | 50 (6) | 720 (94) |
| **Chemotherapy+DES** | | | | | | | | |
| **ECOG 3882** 2003 | Rx: Doxorubicin + DES Ctrl: Doxorubicin | NA | NA | NA | NA | 124 (83) | NA | NA |
| **Chemotherapy+ERA** | | | | | | | | |
| **ENTHUSE (M1c)**, 2013 | Rx: Docetaxel + Zibotentan Ctrl: Docetaxel | NA | NA | NA | NA | NA | NA | NA |
| **SWOG S0421**, 2013 | Rx: Docetaxel + Atrasentan Ctrl: Docetaxel | 20 (2) | NA | NA | 137 (14) | 806 (81) | 41 (4) | 953 (96) |
| **Chemotherapy+IMiD** | | | | | | | | |
| **MAINSAIL**, 2015 | Rx: Docetaxel + Lenalidomide Ctrl: Docetaxel | NA | NA | NA | NA | NA | NA | NA |
| **Chemotherapy+PDGFRi** | | | | | | | | |
| **Mathew P et al**, 2007 | Rx: Docetaxel30 (W) + Imatinib Ctrl: Docetaxel30 (W) | NA | NA | NA | 7 (6) | 102 (88) | 7 (6) | 109 (94) |
| **Chemotherapy+TKI** | | | | | | | | |
| **READY**, 2013 | Rx: Docetaxel + Dasatinib Ctrl: Docetaxel | NA | NA | NA | NA | 1301 (85) | NA | NA |
| **Chemotherapy+VEGFi** | | | | | | | | |
| **CALGB 90401**, 2012 | Rx: Docetaxel + Bevacizumab Ctrl: Docetaxel | NA | NA | NA | NA | 919 (88) | NA | NA |
| **VENICE**, 2013 | Rx: Docetaxel+ Aflibercept Ctrl: Docetaxel | NA | NA | NA | NA | 1112 (91) | NA | NA |
| **Chemotherapy+VitD-Analog** | | | | | | | | |
| **ASCENT2**, 2011 | Rx: Docetaxel + Calcitriol Ctrl: Docetaxel | NA | NA | NA | NA | NA | NA | NA |
| **ERA monotherapy** | | | | | | | | |
| **ENTHUSE (PF)**, 2012 | Rx: Zibotentan Ctrl: Placebo | 200 (34) | NA | NA | 11 (2) | 380 (64) | NA | NA |
| **Carducci MA et al**, 2007 | Rx: Atrasentan Ctrl: Placebo | NA | NA | NA | NA | NA | NA | NA |
| **IMiD monotherapy** | | | | | | | | |
| **Sternberg C et al**, 2016 | Rx: Tasquinimod Ctrl: Placebo | 73 (6) | NA | NA | 28 (2) | 1088 (87) | 139 (11) | 1106 (89) |
| **Immunotherapy monotherapy** | | | | | | | | |
| **CA184-095**, 2017 | Rx: Ipilimumab Ctrl: Placebo | 6 (1) | 3 (<1) | 1 (<1) | 31 (5) | 546 (91) | NA | NA |
| **D9901**, 2006 | Rx: Sipuleucel-T  Ctrl: Placebo | NA | NA | NA | NA | 115 (91) | NA | NA |
| **D9902A**, 2009 | Rx: Sipuleucel-T  Ctrl: Placebo | NA | NA | NA | NA | 90 (92) | NA | NA |
| **Immunotherapy+GMCSF** | | | | | | | | |
| **PROSPECT**, 2019 | Rx1: PVAC + GMCSF Rx2: PVAC Ctrl: Placebo | 20 (2) | NA | NA | 65 (5) | 1207 (93) | 43 (3) | 1254 (97) |
| **PARPi+ARPI** | | | | | | | | |
| **TALAPRO-2**, 2023 | Rx: Talazoparib + Enzalutamide Ctrl: Enzalutamide | 247 (30.7) | NA | NA | 16 (2) | 498 (61.9) | NA | NA |
| **Phenylurea monotherapy** | | | | | | | | |
| **Small EJ et al**, 2000 | Rx: Suramin Ctrl: Placebo | NA | NA | NA | 43 (9) | 415 (91) | NA | NA |
| **Prior ADT+ARPI** | | | | | | | | |
| **ARPI+ARPI** | | | | | | | | |
| **PLATO**, 2018 | Rx: Enzalutamide + Abiraterone Ctrl: Abiraterone | NA | NA | NA | NA | NA | NA | NA |
| **ARPI+Immunotherapy** | | | | | | | | |
| **IMbassador250**, 2022 | Rx: Enzalutamide + Atezolizumab Ctrl: Enzalutamide | NA | NA | NA | NA | NA | NA | NA |
| **Chemotherapy+ARPI** | | | | | | | | |
| **PRESIDE**, 2022 | Rx: Docetaxel  + Enzalutamide Ctrl: Docetaxel | NA | NA | NA | 3 (1) | 267 (99) | 1 (<1) | 271 (99.6) |
| **Chemotherapy+Immunotherapy** | | | | | | | | |
| **KEYNOTE-921**, 2025 | Rx: Docetaxel  + Pembrolizumab Ctrl: Docetaxel | 154 (15) | 14 (1) | 1 (<0.1) | 26 (3) | 800 (78) | NA | NA |
| **PARPi monotherapy** | | | | | | | | |
| **TRITON-3**,  2023 | Rx: Rucaparib Ctrl: Enzalutamide /Abiraterone /Docetaxel | 5 (1) | NA | NA | 14 (3) | 302 (75) | NA | NA |
| **PROfound**, 2020 | Rx: Olaparib Ctrl: Enzalutamide /Abiraterone | 62 (25) ^a^ | NA | NA | 3 (1.2) * | 164 (67) * | NA | NA |
| **Radioligand monotherapy** | | | | | | | | |
| **PSMAfore**, 2024 | Rx: 177Lu Ctrl: Enzalutamide /Abiraterone | 3 (<1%) | NA | NA | 12 (3) | 426 (91) | NA | NA |
| **Prior ADT+Docetaxel** | | | | | | | | |
| **ARPI monotherapy** | | | | | | | | |
| **AFFIRM**, 2012 | Rx: Enzalutamide Ctrl: Placebo | NA | NA | NA | NA | NA | NA | NA |
| **COU-AA-301**, 2011 | Rx: Abiraterone Ctrl: Placebo | NA | NA | NA | NA | NA | NA | NA |
| **ELM-PC 5**, 2015 | Rx: TAK-700 Ctrl: Placebo | 125 (11) | 5 (<1) | NA | 27 (2) | 925 (84) | NA | NA |
| **SAKK 08/11**, 2016 | Rx: TAK-700 Ctrl: Placebo | NA | NA | NA | NA | NA | NA | NA |
| **Chemotherapy monotherapy** | | | | | | | | |
| **PROSELICA**, 2017 | Rx: Cabazitaxel20 Ctrl: Cabazitaxel25 | 81 (7) | NA | NA | 25 (2) | 1054 (88) | NA | NA |
| **TROPIC**, 2010 | Rx: Cabazitaxel25 Ctrl: Mitoxantrone | 58 (8) | NA | NA | 40 (5) | 631 (84) | NA | NA |
| **Chemotherapy+ASO** | | | | | | | | |
| **AFFINITY**, 2017 | Rx: Cabazitaxel25  + Custirsen Ctrl: Cabazitaxel25 | 10 (2) | 1 (<1) | NA | 19 (3) | 474 (75) | 6 (1) | 629 (99) |
| **TKI monotherapy** | | | | | | | | |
| **SUN 1120**, 2013 | Rx: Sunitinib Placebo | NA | NA | NA | NA | NA | NA | NA |
| **Prior ADT+Docetaxel+ARPI** | | | | | | | | |
| **Chemotherapy monotherapy** | | | | | | | | |
| **CARD**, 2019 | Rx: Cabazitaxel25 Ctrl: Enzalutamide /Abiraterone | NA | NA | NA | NA | NA | NA | NA |
| **PARPi+Immunotherapy** | | | | | | | | |
| **KEYLYNK-010**, 2023 | Rx: Olaparib + Pembrolizumab Ctrl: Enzalutamide /Abiraterone | 161 (20) | 1 (<1) | 2 (<1) | 5 (<1) | 618 (78) | NA | NA |
| **Radioligand therapy+Standard of Care** | | | | | | | | |
| **VISION**, 2021 | Rx: 177Lu + SOC Ctrl: SOC | 20 (2) | NA | NA | 55 (7) | 721 (87) | NA | NA |
| **TKI monotherapy** | | | | | | | | |
| **COMET-1**, 2016 | Rx: Cabozantinib Ctrl: Prednisone | 2 (<1) | 1 (<0.1) | NA | 20 (2) | 785 (76) | NA | NA |
| **Heterogeneous Prior Therapy** | | | | | | | | |
| **Chemotherapy+Immunotherapy** | | | | | | | | |
| **VIABLE**, 2022 | Rx: Docetaxel + DCVAC Ctrl: Docetaxel | 9 (<1) | NA | 1 (<0.1) | 45 (4) | 1091 (92) | 28 (2) | 1151 (97) |
| **Multiple subgroups (ADT/ADT+ARPI/ADT+Docetaxel/ADT+ARPI+Docetaxel/Heterogeneous prior therapy)** | | | | | | | | |
| **Chemotherapy monotherapy** | | | | | | | | |
| **SPARC**, 2009 | Rx: Satraplatin Ctrl: Placebo | NA | NA | NA | NA | 841 (89) | NA | NA |
| **Immunotherapy monotherapy** | | | | | | | | |
| **IMPACT**, 2010 | Rx: Sipuleucel-T  Ctrl: Placebo | NA | NA | NA | 30 (5.9) | 461 (90) | NA | NA |
| **PARPi+ARPI** | | | | | | | | |
| **PROpel**, 2023 | Rx: Olaparib + Abiraterone Ctrl: Abiraterone | 138 (17) | NA | NA | 25 (3) | 557 (70) | NA | NA |
| **MAGNITUDE**, 2023 | Rx: Niraparib + Abiraterone Ctrl: Abiraterone | 70 (17) | 2 (<1) | NA | 5 (1.2) | 313 (74) | 51 (12.1) | 335 (79) |
| **Radiopharmaceutical monotherapy** | | | | | | | | |
| **ALSYMPCA**, 2013 | Rx: Radium-223 Ctrl: Placebo | NA | NA | NA | NA | 865 (94) | NA | NA |

Abbreviations: ADT: androgen deprivation therapy; ARPI: androgen-receptor pathway inhibitor; PI3K/AKTi: phosphatidylinositol 3-kinase and protein kinase B inhibitor; ASO: antisense oligonucleotide; DES: diethylstilbestrol diphosphate; ERA: endothelin receptor antagonist; IMiD: immunomodulatory drug; PDGFRi: platelet-derived growth factor receptor inhibitor; TKI: tyrosine kinase inhibitor; VEGFi: vascular endothelial growth factor inhibitor; VitD: vitamin d; PVAC: PROSTVAC (viral vector-based immunotherapy); GMCSF: granulocyte-macrophage colony-stimulating factor; PARPi: poly(ADP-ribose) polymerase inhibitor; 177Lu: 177Lu-PSMA-617; SOC: standard of care

Note: PLATO and CARD were phase IV trials. All remaining trials were phase III. The color ‘green’ represents a positive trial, ‘red’ represents a negative trial.

* PROfound trial only reported data for Cohort A instead of overall population

Cabazitaxel20: Cabazitaxel 20 mg/m^2^ IV on day 1 of every 3-week cycle

Cabazitaxel25: Cabazitaxel 25 mg/m^2^ IV on day 1 of every 3-week cycle

Docetaxel (2W): Docetaxel 75 mg/m^2^ IV on days 1 and 15 of a 4-week cycle

Docetaxel (int): Docetaxel 35 mg/m^2^ IV on days 1, 8, 15, repeat cycle at day 29

Docetaxel30 (W): Docetaxel 30 mg/m^2^ IV on days 1, 8, 15, 22 and 29 of a 6-week cycle

Remaining drugs were administered at standard doses.

### **Supplement Table 4:** Distribution of race and ethnicity across the included phase II trials

| **Trial** | **Arm** | **Race \| N (%)** | | | | | **Ethnicity \| N (%)** | |
| --- | --- | --- | --- | --- | --- | --- | --- | --- |
|  |  | **Asian** | **American Indian or Alaskan Native** | **Native Hawaiian/ Pacific Islander** | **Black or African American** | **White** | **Hispanic/**  **Latino** | **Non-Hispanic/Non-Latino** |
| **Prior ADT only** | | | | | | | | |
| **Antiandrogen+Chemotherapy** | | | | | | | | |
| **Takahashi M et al**, 2013 | Rx: Antiandrogen + Tegafur-Uracil Ctrl: Antiandrogen | NA | NA | NA | NA | NA | NA | NA |
| **Antifungal monotherapy** | | | | | | | | |
| **Antonarakis ES et al**, 2013 | Rx: Itraconazole600 Ctrl: Itraconazole200 | NA | NA | NA | NA | 31 (67) | NA | NA |
| **Antifungal+Bisphosphonate** | | | | | | | | |
| **Figg WD et al,** 2005 | Rx: Ketoconazole + Alendronate Ctrl: Ketoconazole | NA | NA | NA | NA | NA | NA | NA |
| **Antifungal+Chemotherapy** | | | | | | | | |
| **Millikan R et al**, 2001 | Rx1: Ketoconazole + Doxorubicin Rx2: Ketoconazole | NA | NA | NA | NA | NA | NA | NA |
| **ARPI monotherapy** | | | | | | | | |
| **TERRAIN**, 2016 | Rx: Enzalutamide Ctrl: Bicalutamide | 5 (1) | NA | 2 (<1) | 18 (5) | 348 (93) | 4 (1) | 371 (99) |
| **ARPI+ARPI** | | | | | | | | |
| **Khalaf DJ et al**, 2019 | Rx1: Abiraterone followed by Enzalutamide Rx2: Enzalutamide followed by Abiraterone | NA | NA | NA | NA | NA | NA | NA |
| **ARPI+Immunotherapy** | | | | | | | | |
| **STAMP**, 2015 | Rx1: Abiraterone + cSipuleucel-T Rx2: Abiraterone + sSipuleucel-T | 2 (3) | NA | NA | 5 (7) | 62 (90) | NA | NA |
| **STRIDE**, 2023 | Rx1: Enzalutamide + cSipuleucel-T  Rx2: Enzalutamide + sSipuleucel-T | NA | NA | NA | 2 (4) | 50 (96) | NA | NA |
| **ARPI+TKI** | | | | | | | | |
| **Dorff TB et al**, 2019 | Rx: Abiraterone  + Dasatinib Ctrl: Abiraterone | 3 (12) | NA | NA | 3 (12) | 11 (42) | 9 (35) | 17 (65) |
| **ASO monotherapy** | | | | | | | | |
| **Yu EY et al**, 2018 | Rx: Apatorsen Ctrl: Prednisone | NA | NA | NA | NA | 69 (93) | NA | NA |
| **Tolcher AW et al,** 2002 | Rx: ISIS 3521 Ctrl: ISIS 5132 | NA | NA | NA | NA | NA | NA | NA |
| **Chemotherapy monotherapy** | | | | | | | | |
| **Heidenreich A et al**, 2004 | Rx1: L-Doxorubicin25  Rx2: L-Doxorubicin50 | NA | NA | NA | NA | NA | NA | NA |
| **TIPC**, 2007 | Rx: Docetaxel30 (W) Ctrl: Prednisolone | NA | NA | NA | NA | NA | NA | NA |
| **Krainer M et al**, 2007 | Rx: Docetaxel25 (W) Ctrl: Vinorelbine | NA | NA | NA | NA | NA | NA | NA |
| **Chemotherapy+Antifungal** | | | | | | | | |
| **Millikan R et al,** 2003 | Rx: Vin/Estra + Keto/Doxorubicin Ctrl: Paclitaxel + Estramustine + Etoposide | NA | NA | NA | NA | NA | NA | NA |
| **Chemotherapy+ARPI** | | | | | | | | |
| **CHEIRON**, 2021 | Rx: Docetaxel + Enzalutamide Ctrl: Docetaxel | NA | NA | NA | NA | NA | NA | NA |
| **Chemotherapy+ASO** | | | | | | | | |
| **Chi KN et al,**  2010 | Rx: Docetaxel + Custirsen Ctrl: Docetaxel | NA | NA | NA | NA | NA | NA | NA |
| **Wiechno P et al**, 2014 | Rx: Docetaxel + LY2181308 Ctrl: Docetaxel | NA | NA | NA | NA | NA | NA | NA |
| **Chemotherapy+Bcl-2 inhibitor** | | | | | | | | |
| **EORTC**, 2009 | Rx: Docetaxel  + Oblimersen Ctrl: Docetaxel | NA | NA | NA | NA | NA | NA | NA |
| **Sonpavde G et al**, 2011 | Rx: Docetaxel + AT-101 Ctrl: Docetaxel | NA | NA | NA | 9 (4) | 205 (93) | NA | NA |
| **Chemotherapy+Chemotherapy** | | | | | | | | |
| **Machiels JP et al**, 2008 | Rx: Docetaxel35 (W) + Estramustine Ctrl: Docetaxel35 (W) | NA | NA | NA | NA | NA | NA | NA |
| **Berry WR et al**, 2004 | Rx: Paclitaxel + Estramustine Ctrl: Paclitaxel | NA | NA | NA | 20 (12) | 141 (87) | 1 (<1) | 162 (99) |
| **Albrecht W et al**, 2004 | Rx: Estramustine + Vinblastine Ctrl: Estramustine | NA | NA | NA | NA | NA | NA | NA |
| **Cabrespine A et al**, 2006 | Rx: Paclitaxel + Carboplatin Ctrl: Mitoxantrone | NA | NA | NA | NA | NA | NA | NA |
| **Nelius T et al**, 2006 | Rx1: Docetaxel70 + Estramustine840 Rx2: Docetaxel70 + Estramustine420 | NA | NA | NA | NA | NA | NA | NA |
| **Oudard S et al**, 2005 | Rx1: Docetaxel70 + Estramustine Rx2: Docetaxel35  (D2,D9 Q3W) + Estramustine Ctrl: Mitoxantrone | NA | NA | NA | NA | NA | NA | NA |
| **Droz JP et al**, 2003 | Rx1: Oxaliplatin + 5-Fluorouracil Rx2: Oxaliplatin | NA | NA | NA | NA | NA | NA | NA |
| **Galsky MD et al**, 2005 | Rx1: Ixabepilone + Estramustine Rx2: Ixabepilone | NA | NA | NA | NA | NA | NA | NA |
| **Eymard JC et al**, 2007 | Rx: Docetaxel70 + Estramustine Ctrl: Docetaxel | NA | NA | NA | NA | NA | NA | NA |
| **Chemotherapy+Curcuminoid** | | | | | | | | |
| **Jahanmohan JP et al**, 2021 | Rx: Docetaxel + Curcumin Ctrl: Docetaxel | NA | NA | NA | NA | NA | NA | NA |
| **Chemotherapy+Immunotherapy** | | | | | | | | |
| **Dahut WL et al**, 2004 | Rx: Docetaxel30 (W) ^a^ + Thalidomide Ctrl: Docetaxel30 (W) ^a^ | NA | NA | NA | NA | NA | NA | NA |
| **ECOG 3899**, 2010 | Rx1: 13-cis Retinoic acid + Interferon-alpha2b + Paclitaxel Rx2: Vinorelbine + Mitoxantrone + Estramustine | NA | NA | NA | 4 (6) | 58 (92) | 1 (2) | 62 (98) |
| **E1809**, 2015 | Rx: Docetaxel + PVAC-VF Ctrl: Docetaxel | NA | NA | NA | NA | 9 (90) | NA | NA |
| **de Bono et al**, 2014 | Rx: Docetaxel + Figitumumab Ctrl: Docetaxel | NA | NA | NA | 6 (3) | 191 (94) | NA | NA |
| **Heidenreich A et al,**  2013 | Rx: Docetaxel  + Intetumumab Ctrl: Docetaxel | 21 (16) | NA | NA | 3 (2) | 107 (82) | NA | NA |
| **Chemotherapy+Radiopharmaceutical** | | | | | | | | |
| **Taxium II**, 2017 | Rx: Docetaxel  + Rhenium-188-HEDP Ctrl: Docetaxel | NA | NA | NA | NA | NA | NA | NA |
| **Chemotherapy+TKI** | | | | | | | | |
| **Horti J et al**, 2009 | Rx: Docetaxel + Vandetanib Ctrl: Docetaxel | NA | NA | NA | 7 (8) | 72 (84) | NA | NA |
| **Chemotherapy+Vascular disrupting agent** | | | | | | | | |
| **Pili R et al,**  2010 | Rx: Docetaxel + Vadimezan Ctrl: Docetaxel | NA | NA | NA | NA | NA | NA | NA |
| **Chemotherapy+VitD-Analog** | | | | | | | | |
| **Attia S et al**, 2008 | Rx: Docetaxel35 (W) ^b^ + Doxercalciferol Ctrl: Docetaxel35 (W) ^b^ | NA | NA | NA | 2 (3) | 65 (93) | NA | NA |
| **ASCENT**, 2007 | Rx: Docetaxel36 (W) + Calcitriol Ctrl: Docetaxel36 (W) | NA | NA | NA | 29 (12) | 205 (82) | NA | NA |
| **Corticosteroid monotherapy** | | | | | | | | |
| **Venkitaraman R et al**, 2015 | Rx: Dexamethasone Ctrl: Prednisolone | NA | NA | NA | NA | NA | NA | NA |
| **Corticosteroid+Immunotherapy** | | | | | | | | |
| **Yoshimura K et al**, 2016 | Rx: Dexamethasone + PPV Ctrl: Dexamethasone | NA | NA | NA | NA | NA | NA | NA |
| **Corticosteroid+VEGFi** | | | | | | | | |
| **Stadler WM et al**, 2004 | Rx: Dexamethasone + SU5416 Ctrl: Dexamethasone | NA | NA | NA | 7 (21) | 26 (76) | 1 (3) | 33 (97) |
| **ERA monotherapy** | | | | | | | | |
| **James ND et al**, 2009 | Rx1: Zibotentan15  Rx2: Zibotentan10 Ctrl: Placebo | NA | NA | NA | NA | NA | NA | NA |
| **Carducci MA et al**, 2003 | Rx1: Atrasentan2.5 Rx2: Atrasentan10 Ctrl: Placebo | 2 (1) | NA | NA | 6 (2) | 278 (97) | 2 (1) | 287 (99) |
| **IMiD monotherapy** | | | | | | | | |
| **Pili R et al**, 2011 | Rx: Tasquinimod Ctrl: Placebo | 2 (<1) | NA | NA | 22 (11) | 173 (86) | 8 (4) | 193 (96) |
| **Immunotherapy monotherapy** | | | | | | | | |
| **PERSEUS**, 2016 | Rx1: Abituzumab750 + SOC Rx2: Abituzumab1500 + SOC Ctrl: SOC | 1 (<1) | NA | NA | 6 (3) | 171 (95) | NA | NA |
| **Figg WD et al**, 2001 | Rx: ThalidomideHD Ctrl: Thalidomide | NA | NA | NA | NA | NA | NA | NA |
| **Immunotherapy+Chemotherapy** | | | | | | | | |
| **Arlen PM et al,** 2006 | Rx: Vaccine + GMCSF + Docetaxel Ctrl: Vaccine + GMCSF | NA | NA | NA | NA | NA | NA | NA |
| **Immunotherapy+GMCSF** | | | | | | | | |
| **TBC-PRO-002**, 2010 | Rx: PVAC+GMCSF Ctrl: Placebo | NA | NA | NA | 14 (11) | 104 (85) | 2 (2) | 120 (98) |
| **Integrin inhibitor monotherapy** | | | | | | | | |
| **Bradley DA et al,** 2011 | Rx: Cilengitide500 Ctrl: Cilengitide2000 | 2 (5) | NA | NA | 4 (9) | 38 (86) | NA | NA |
| **Matrix Metalloproteinase Inhibitor monotherapy** | | | | | | | | |
| **Lara Jr PN et al**, 2006 | Rx: BMS-275291 [1200mg] Ctrl: BMS-275291 [2400mg] | NA | NA | NA | NA | NA | NA | NA |
| **Progestin monotherapy** | | | | | | | | |
| **Patel SR et al**, 1990 | Rx: Megestrol Acetate Ctrl: Dexamethasone | NA | NA | NA | NA | NA | NA | NA |
| **Somatostatin+Corticosteroid** | | | | | | | | |
| **Dimopoulos MA et al**, 2004 | Rx: Lanreotide + Dexamethasone Ctrl: Estramustine + Etoposide | NA | NA | NA | NA | NA | NA | NA |
| **TKI monotherapy** | | | | | | | | |
| **Boccardo F et al**, 2008 | Rx: Gefitinib Ctrl: Placebo | NA | NA | NA | NA | NA | NA | NA |
| **TKI+1st gen AA** | | | | | | | | |
| **Azad AA et al**, 2014 | Rx: Vandetanib + Bicalutamide Ctrl: Bicalutamide | NA | NA | NA | NA | NA | NA | NA |
| **Sridhar SS et al**, 2015 | Rx1: Pazopanib + Bicalutamide Rx2: Pazopanib | NA | NA | NA | NA | NA | NA | NA |
| **Prior ADT+ARPI** | | | | | | | | |
| **ARPI+TKI** | | | | | | | | |
| **Spetsieris N et al**, 2021 | Rx: Abiraterone + Sunitinib Ctrl: Abiraterone + Dasatinib | 3 (2) | NA | NA | 20 (11) | 146 (81) | 10 (6) | 169 (94) |
| **Bipolar Androgen Therapy** | | | | | | | | |
| **TRANSFORMER**, 2021 | Rx: BAT Ctrl: Enzalutamide | 5 (3) | 1 (<1) | NA | 14 (7) | 170 (87) | 6 (3) | 183 (94) |
| **Chemotherapy+ARPI** | | | | | | | | |
| **ABIDO SOGUG**, 2022 | Rx: Docetaxel + Abiraterone Ctrl: Docetaxel | NA | NA | NA | NA | NA | NA | NA |
| **Chemotherapy+Chemotherapy** | | | | | | | | |
| **MDACC Study**, 2019 | Rx: Cabazitaxel25 + Carboplatin Ctrl: Cabazitaxel25 | NA | NA | NA | 24 (15) | 121 (76) | NA | NA |
| **Prior ADT+Docetaxel** | | | | | | | | |
| **ASO+Chemotherapy** | | | | | | | | |
| **P-06c,**  2011 | Rx1: Custirsen + Docetaxel Rx2: Custirsen  + Mitoxantrone | NA | NA | NA | NA | NA | NA | NA |
| **Aurora kinase inhibitor monotherapy** | | | | | | | | |
| **Meulenbeld HJ et al**, 2012 | Rx1: Danusertib330 Rx2: Danusertib500 | NA | NA | NA | NA | NA | NA | NA |
| **Chemotherapy monotherapy** | | | | | | | | |
| **GETUG-P02,**  2015 | Rx1: Etoposide Rx2: Vinorelbine Rx3: Mitoxantrone | NA | NA | NA | NA | NA | NA | NA |
| **CAINTA**, 2023 | Rx: Cabazitaxel ID Ctrl: Cabazitaxel25 | NA | NA | NA | NA | NA | NA | NA |
| **Rosenberg JE et al**, 2007 | Rx: Ixabepilone Ctrl: Mitoxantrone | NA | NA | NA | NA | NA | NA | NA |
| **Chemotherapy+Chemotherapy** | | | | | | | | |
| **RECARDO**, 2018 | Rx: Docetaxel + Carboplatin Ctrl: Docetaxel | NA | NA | NA | NA | NA | NA | NA |
| **Chemotherapy+Immunotherapy** | | | | | | | | |
| **Fizazi K et al**, 2012 | Rx: Mitoxantrone + Siltuximab Ctrl: Mitoxantrone | NA | NA | NA | 6 (6) | 99 (93) | NA | NA |
| **Hakenberg OW et al**, 2019 | Rx: Mitoxantrone  + Olaratumab Ctrl: Mitoxantrone | NA | NA | NA | NA | 120 (99) | 5 (4) | 116 (96) |
| **Immunotherapy monotherapy** | | | | | | | | |
| **Filaci G et al**, 2021 | Rx1: GX301 vaccine (8D) Rx2: GX301 vaccine (4D) Rx3: GX301 vaccine (2D) | NA | NA | NA | NA | NA | NA | NA |
| **TKI monotherapy** | | | | | | | | |
| **Droz JP et al**, 2014 | Rx: Nintedanib150 Ctrl: Nintedanib250 | NA | NA | NA | NA | NA | NA | NA |
| **Prior ADT+Docetaxel+ARPI** | | | | | | | | |
| **ARPI monotherapy** | | | | | | | | |
| **SAK-08-16**, 2023 | Rx: Darolutamide Ctrl: Placebo | NA | NA | NA | NA | NA | NA | NA |
| **Chemotherapy monotherapy** | | | | | | | | |
| **ConCab**, 2018 | Rx: Cabazitaxel (W) Ctrl: Cabazitaxel25 | NA | NA | NA | NA | NA | NA | NA |
| **Chemotherapy+AKTi** | | | | | | | | |
| **RE-AKT**, 2024 | Rx: Enzalutamide  + Capivasertib Ctrl: Enzalutamide | NA | NA | NA | NA | 97 (97) | NA | NA |
| **Radioligand monotherapy** | | | | | | | | |
| **TheraP**, 2021 | Rx: 177Lu Ctrl: Cabazitaxel20 | NA | NA | NA | NA | NA | NA | NA |
| **Heterogeneous Prior Therapy** | | | | | | | | |
| **ARPI+Radioligand** | | | | | | | | |
| **ENZA-p**, 2024 | Rx: Enzalutamide + 177Lu Ctrl: Enzalutamide | NA | NA | NA | NA | NA | NA | NA |
| **Chemotherapy monotherapy** | | | | | | | | |
| **TAXYNERGY**, 2017 | Rx: Docetaxel Ctrl: Cabazitaxel25 | 1 (2) | NA | NA | 7 (11) | 55 (87) | NA | NA |
| **Chemotherapy+Immunotherapy** | | | | | | | | |
| **IND 209**, 2018 | Rx: Docetaxel  + Reolysin Ctrl: Docetaxel | NA | NA | NA | NA | NA | NA | NA |
| **Kongsted P et al,** 2017 | Rx: Docetaxel  + DCVAC Ctrl: Docetaxel | NA | NA | NA | NA | NA | NA | NA |
| **PARPi+ARPi** | | | | | | | | |
| **BRCAAway**, 2024 | Rx1: Abiraterone Rx2: Olaparib Rx3: Abiraterone  + Olaparib | NA | NA | NA | 6 (10) | 54 (89) | 1 (2) | 60 (98) |
| **TKI monotherapy** | | | | | | | | |
| **Monk P et al**, 2018 | Rx: Tivantinib Ctrl: Placebo | 1 (1) | NA | NA | 8 (10) | 69 (88) | NA | 75 (96) |
| **Smith DC et al**, 2013 | Rx: Cabozantinib Ctrl: Placebo | NA | NA | NA | NA | NA | NA | NA |
| **Multiple subgroups (ADT/ADT+ARPI/ADT+Docetaxel/ADT+ARPI+Docetaxel/Heterogeneous prior therapy)** | | | | | | | | |
| **Chemotherapy monotherapy** | | | | | | | | |
| **Annala M et al**, 2021 | Rx: Cabazitaxel25 Ctrl: Enzalutamide /Abiraterone | NA | NA | NA | NA | NA | NA | NA |
| **Chemotherapy+AKTi** | | | | | | | | |
| **ProCAID**, 2021 | Rx: Docetaxel + Capivasertib Ctrl: Docetaxel | NA | NA | NA | NA | NA | NA | NA |

Abbreviations: ADT: androgen deprivation therapy; ARPI: androgen-receptor pathway inhibitor; cSipuleucel-T: concurrent sipuleucel-T; sSipuleucel-T: sequential sipuleucel-T; TKI: tyrosine kinase inhibitor; ASO: antisense oligonucleotide; Bcl-2: B-cell lymphoma 2 protein; PVAC: PROSTVAC (viral vector-based immunotherapy); VitD: vitamin d; VEGFi: vascular endothelial growth factor inhibitor; ERA: endothelin receptor antagonist; IMiD: immunomodulatory drug; SOC: standard of care; GMCSF: granulocyte-macrophage colony-stimulating factor; AA: antiandrogen; BAT: bipolar androgen therapy; PARPi: poly(ADP-ribose) polymerase inhibitor; 177Lu: 177Lu-PSMA-617; AKTi: protein kinase B inhibitor; Vin/Estra: vinblastine/estramustine; keto: ketoconazole

The color ‘green represents a positive trial, ‘red’ represents a negative trial.

Itraconazole600: Itraconazole 600mg/day PO

Itraconazole200: Itraconazole 200mg/day PO

L-Doxorubicin25: Liposomal Doxorubicin (dissolved in 250 mL 5% glucose) 25 mg/m^2^ IV every 2 weeks for 12 consecutive cycles

L-Doxorubicin50: Liposomal Doxorubicin (dissolved in 250 mL 5% glucose) 50 mg/m^2^ IV every 4 weeks for 6 consecutive cycles

Docetaxel30 (W): Docetaxel 30 mg/m^2^ IV every week for 5 weeks, followed by 1 week rest (6-week cycle)

Docetaxel25 (W): Docetaxel 25 mg/m^2^ IV every week for 2 cycles (8-week cycle)

Docetaxel35 (W): Docetaxel 35 mg/m^2^ IV on days 2 and 9 every 3 weeks

Docetaxel70: Docetaxel 70 mg/m^2^ on day 2 every 3 weeks

Estramustine840: Estramustine 840 mg/day on days 1-5

Estramustine420: Estramustine 420 mg/day on days 1-3

Docetaxel35 (D2,D9 Q3W): Docetaxel 35 mg/m^2^ on day 2 and 9 every 3wks

Docetaxel30 (W) ^a^ : Docetaxel 30 mg/m^2^ IV every week for 3 weeks, followed by 1 week rest (4-week cycle)

Docetaxel35 (W) ^b^ : Docetaxel 35 mg/m^2^ IV on days 1, 8, and 15 (4-week cycle)

Docetaxel36 (W): Docetaxel 36 mg/m^2^ IV every week for 3 weeks (4-week cycle)

Zibotentan15: Zibotentan 15 mg PO OD

Zibotentan10: Zibotentan 10 mg PO OD

Atrasentan2.5: Atrasentan 2.5 mg PO OD

Atrasentan10: Atrasentan 10 mg PO OD

Abituzumab750: Abituzumab 750 mg every 3 weeks

Abituzumab1500: Abituzumab 1500 mg every 3 weeks

ThalidomideHD: Thalidomide initial dose of 200 mg/day with increments of 200 mg/day every 2 weeks to a maximum dose of 1200 mg

Cilengitide500: Cilengitide 500 mg IV twice weekly (six-week cycle)

Cilengitide2000: Cilengitide 2000 mg IV twice weekly (six-week cycle)

Cabazitaxel25: Cabazitaxel 25 mg/m^2^ IV every 3 weeks

Danusertib330: Danusertib 330 mg/m^2^ IV on days 1,8 and 15 every 4 weeks

Danusertib500: Danusertib 500 mg/m^2^ IV on days 1 and 15 every 4 weeks

Cabazitaxel ID: Cabazitaxel at an initial dose of 25 mg/m^2^ followed by dose adaptations in cycle 2 and beyond according to a prespecified dosing algorithm considering previous-cycle hematologic toxicity and cabazitaxel AUC (with a target AUC of 0.8–1.2 mg*hour/L)

GX301 vaccine (8D): GX301, 8 doses on days 1, 3, 5, 7, 14, 21, 35 and 63

GX301 vaccine (4D): GX301, 4 doses on days 1, 14, 35 and 63

GX301 vaccine (2D): GX301, 2 doses on days 1 and 63

Nintedanib150: Nintedanib 150 mg PO BID for 6 months

Nintedanib250: Nintedanib 250 mg PO BID for 6 months

Cabazitaxel (W): Cabazitaxel 25 mg/m^2^ IV weekly for 5 weeks (6-week cycle)

Cabazitaxel20: Cabazitaxel 20 mg/m^2^ IV every 3 weeks

Remaining drugs were administered at standard doses.

### **Supplement Table 5:** Data reported for race and ethnicity across included trials

| **Race/ Ethnicity** | **Phase II** | | **Phase III** | | **Total** | |
| --- | --- | --- | --- | --- | --- | --- |
|  | **Trials \| N** | **Patients \| N** | **Trials \| N** | **Patients \| N** | **Trials \| N** | **Patients \| N** |
| White | 30 | 3345 | 41 | 26538 | 71 | 29883 |
| Black/African American | 25 | 252 | 31 | 1088 | 56 | 1340 |
| Asian | 12 | 48 | 26 | 2166 | 38 | 2214 |
| American Indian/Alaskan Native | 1 | 1 | 10 | 47 | 11 | 48 |
| Native Hawaiian/Pacific Islander | 1 | 2 | 5 | 7 | 6 | 9 |
| Hispanic/Latino | 12 | 50 | 12 | 512 | 24 | 562 |
| Non-Hispanic/Non-Latino | 13 | 1848 | 12 | 9003 | 25 | 10851 |

### **Supplement Table 6:** Summary of characteristics of included phase II trials

A) **Prior ADT only**

| **Trial** | **Arm** | **Estimated /Actual Accrual** | **Years of enrollment** | **Median follow up - months** | **Primary Endpoint** | **OS HR (95% CI)** | **PFS HR (95% CI)** | **Prior therapy \| %** |
| --- | --- | --- | --- | --- | --- | --- | --- | --- |
| **Antiandrogen+Chemotherapy** | | | | | | | | |
| **Takahashi M et al**, 2013 | Rx: Antiandrogen + Tegafur-Uracil Ctrl: Antiandrogen | Not specified/ 52 | 02/01/2006- 11/01/2009 | NA | PSA50 response rate | Not reported | Not reported | ADT: 100 |
| **Antifungal monotherapy** | | | | | | | | |
| **Antonarakis ES et al**, 2013 | Rx: Itraconazole600 Ctrl: Itraconazole200 | Not specified/ 46 | NA | NA | PSA-PFS rate at 24 weeks | Not reported | 8.3 vs. 2.7 * | ADT: 100 |
|  |  |  |  |  |  |  | NA |  |
| **Antifungal+Bisphosphonate** | | | | | | | | |
| **Figg WD et al,** 2005 | Rx: Ketoconazole + Alendronate Ctrl: Ketoconazole | 72/ 72 | 03/01/1993 onwards | 23.9 | PSA50 response rate; ORR | 19 vs. NR | 4.6 vs. 3.8 * | ADT: 100 |
|  |  |  |  |  |  | NA | NA |  |
| **Antifungal+Chemotherapy** | | | | | | | | |
| **Millikan R et al**, 2001 | Rx1: Ketoconazole + Doxorubicin Rx2: Ketoconazole | 90/ 90 | 07/01/1995- 10/01/1996 | 36 | OS; Response rate | 12.5 vs. 12.5 | Not reported | ADT: 100 |
|  |  |  |  |  |  | NA |  |  |
| **ARPI monotherapy** | | | | | | | | |
| **TERRAIN**, 2016 | Rx: Enzalutamide Ctrl: Bicalutamide | 370/ 375 | 03/22/2011- 07/11/2013 | Rx: 20 Ctrl: 16.7 | PFS | Not reported | 15.7 vs. 5.8 * | ADT: 100 |
|  |  |  |  |  |  |  | 0.44 (0.34-0.57) |  |
| **ARPI+ARPI** | | | | | | | | |
| **Khalaf DJ et al**, 2019 | Rx1: Abiraterone followed by Enzalutamide Rx2: Enzalutamide followed by Abiraterone | 200/ 202 | 10/21/2014- 12/13/2016 | 30.7 | Time to second PSA progression; PSA30 response to second-line therapy | 28.8 vs. 24.7 | 7.9 vs. 7.3 ^†^ | ADT: 100 Docetaxel: 5 |
|  |  |  |  |  |  | 0.79 (0.54-1.16) | 0.95 (0.70-1.29) |  |
| **ARPI+Immunotherapy** | | | | | | | | |
| **STAMP**, 2015 | Rx1: Abiraterone + cSipuleucel-T Rx2: Abiraterone + sSipuleucel-T | 60/ 69 | 12/01/2011- 02/01/2013 | NA | Cumulative APC activation | 30 vs. 34.2 | Not reported | ADT: 100 |
|  |  |  |  |  |  | 1.0 (0.51-1.98) |  |  |
| **STRIDE**, 2023 | Rx1: Enzalutamide + cSipuleucel-T  Rx2: Enzalutamide + sSipuleucel-T | 100/ 52 | 10/11/2013- 07/17/2017 | Rx: 40.6 Ctrl: 40.2 | PA2024-specific T cell proliferation response | 34.7 vs. 32.5 | Not reported | ADT: 100 |
|  |  |  |  |  |  | 1.41 (0.68-2.92) |  |  |
| **ARPI+TKI** | | | | | | | | |
| **Dorff TB et al**, 2019 | Rx: Abiraterone  + Dasatinib Ctrl: Abiraterone | 96/ 26 | 09/01/2012- 04/01/2015 | 41.8 | PFS | 41.2 vs. 26.9 | 15.7 vs. 9 ^‡^ | ADT: 100 |
|  |  |  |  |  |  | NA | NA |  |
| **ASO monotherapy** | | | | | | | | |
| **Yu EY et al**, 2018 | Rx: Apatorsen Ctrl: Prednisone | 64/ 74 | 01/01/2011- 11/01/2013 | NA | Proportion of patients without disease progression at 12 weeks | Not reported | 17.9 vs. 13.9 | ADT: 100 |
|  |  |  |  |  |  |  | NA |  |
| **Tolcher AW et al,**  2002 | Rx: ISIS 3521 Ctrl: ISIS 5132 | Not specified/ 31 | NA | NA | PSA50 response rate; ORR; Treatment failure; Safety | Not reported | Not reported | ADT: 100 |
| **Chemotherapy monotherapy** | | | | | | | | |
| **Heidenreich A et al**, 2004 | Rx1: L-Doxorubicin25  Rx2: L-Doxorubicin50 | 50/ 48 | NA | 42 | PSA50 response rate | Not reported | Not reported | ADT: 100 |
| **TIPC**, 2007 | Rx: Docetaxel30 (W) Ctrl: Prednisolone | 182/ 134 | 10/01/2002- 12/01/2004 | NA | PSA50 response rate at 6 weeks | 27 vs. 18 | 11 vs. 4 * | ADT: 100 |
|  |  |  |  |  |  | NA | NA |  |
| **Krainer M et al**, 2007 | Rx: Docetaxel25 (W) Ctrl: Vinorelbine | Not specified/ 40 | NA | NA | Time to disease progression | Not reported | 14.5 vs. 4.4 ^†^ | ADT: 100 |
|  |  |  |  |  |  |  | NA |  |
| **Chemotherapy+Antifungal** | | | | | | | | |
| **Millikan R et al,** 2003 | Rx: Vin/Estra + Keto/Doxorubicin Ctrl: Paclitaxel + Estramustine + Etoposide | 92/ 75 | 01/01/1998- 05/01/2000 | NA | OS ORR PSA50 response rate | 23.4 vs. 16.9 | Not reported | ADT: 100 |
|  |  |  |  |  |  | NA |  |  |
| **Chemotherapy+ARPI** | | | | | | | | |
| **CHEIRON**, 2021 | Rx: Docetaxel + Enzalutamide Ctrl: Docetaxel | 232/ 246 | 09/24/2014- 11/10/2017 | 24.1 | Proportion of patients without disease progression at 6 months | 28.7 vs. 30.3 | 12.8 vs. 9.6 ^‡^ | ADT: 100 |
|  |  |  |  |  |  | 1.11 (0.79-1.56) | 0.62 (0.44-0.85) |  |
| **Chemotherapy+ASO** | | | | | | | | |
| **Chi KN et al,**  2010 | Rx: Docetaxel + Custirsen Ctrl: Docetaxel | 80/ 82 | 09/01/2005- 12/01/2006 | 35 | PSA50 response rate | 23.8 vs. 16.9 | 7.3 vs. 6.1 * | ADT: 100 |
|  |  |  |  |  |  | 0.61 (0.36-1.02) | 0.86 (0.54-1.38) |  |
| **Wiechno P et al**, 2014 | Rx: Docetaxel + LY2181308 Ctrl: Docetaxel | 150/ 154 | 04/01/2008- 10/01/2010 | NA | PFS | 27 vs. 29 | 8.6 vs. 9 * | ADT: 100 |
|  |  |  |  |  |  | NA | NA |  |
| **Chemotherapy+Bcl-2 inhibitor** | | | | | | | | |
| **EORTC**, 2009 | Rx: Docetaxel  + Oblimersen Ctrl: Docetaxel | 102/ 111 | 04/01/2004- 01/01/2006 | NA | PSA50 response rate; Toxicity | Not reported | 4.2 vs. 6.3 ^†^ | ADT: 100 |
|  |  |  |  |  |  |  | NA |  |
| **Sonpavde G et al**, 2011 | Rx: Docetaxel + AT-101 Ctrl: Docetaxel | 250/ 221 | 11/01/2007- 04/01/2009 | NA | OS | 18.1 vs. 17.8 | 11 vs. 10.3 * | ADT: 100 |
|  |  |  |  |  |  | 1.07 (0.72-1.55) | 0.88 (0.63-1.22) |  |
| **Chemotherapy+Chemotherapy** | | | | | | | | |
| **Machiels JP et al**, 2008 | Rx: Docetaxel35 (W) + Estramustine Ctrl: Docetaxel35 (W) | 136/ 150 | 02/01/2004- 08/01/2006 | 16 | PSA50 response rate | 19.3 vs. 21 | 6.3 vs. 6.6 * | ADT: 100 |
|  |  |  |  |  |  | NA | NA |  |
| **Berry WR et al**, 2004 | Rx: Paclitaxel + Estramustine Ctrl: Paclitaxel | 166/ 166 | 12/01/1998- 12/01/1999 | NA | Objective response | 16.1 vs. 13.1 | 5.5 vs. 4.3 ^†^ | ADT: 100 |
|  |  |  |  |  |  | NA | NA |  |
| **Albrecht W et al**, 2004 | Rx: Estramustine + Vinblastine Ctrl: Estramustine | 80/ 90 | 10/01/1995- 11/01/1996 | NA | PSA response rate | 10.1 vs. 11.7 | Not reported | ADT: 100 |
|  |  |  |  |  |  | NA |  |  |
| **Cabrespine A et al**, 2006 | Rx: Paclitaxel + Carboplatin Ctrl: Mitoxantrone | 40/ 40 | 03/01/2002- 12/01/2004 | Rx: 7.5 Ctrl: 6.3 | PSA50 response rate | 14.5 vs. 11.1 | Not reported | ADT: 100 |
|  |  |  |  |  |  | NA |  |  |
| **Nelius T et al**, 2006 | Rx1: Docetaxel70 + Estramustine840 Rx2: Docetaxel70 + Estramustine420 | 68/ 72 | 01/01/2002- 07/01/2005 | 14.5 | PSA50 response rate | 21 vs. 22 | Not reported | ADT: 100 |
|  |  |  |  |  |  | NA |  |  |
| **Oudard S et al**, 2005 | Rx1: Docetaxel70 + Estramustine Rx2: Docetaxel35 (D2,D9 Q3W) + Estramustine Ctrl: Mitoxantrone | 130/ 130 | 01/01/2000- 01/01/2002 | 12 | PSA50 response rate | Rx1 vs Rx2: 18.6 vs. 18.4 Rx1 vs Ctrl:  18.6 vs. 13.4 Rx2 vs Ctrl:  18.4 vs. 13.4 | Not reported | ADT: 100 |
|  |  |  |  |  |  | Rx1 vs Rx2: 1.43 (0.89-2.31) Rx1 vs Ctrl:  1.08 (0.66-1.76) Rx2 vs Ctrl:  0.75 (0.46-1.21) |  |  |
| **Droz JP et al**, 2003 | Rx1: Oxaliplatin + 5-Fluorouracil Rx2: Oxaliplatin | Not specified/ 54 | 02/01/1998- 08/01/1999 | 7.9 | Best overall response rate | 11.4 vs. 9.4 | 3.4 vs. 2.6 ^†^ | ADT: 100 |
|  |  |  |  |  |  | NA | NA |  |
| **Galsky MD et al**, 2005 | Rx1: Ixabepilone + Estramustine Rx2: Ixabepilone | Not specified/ 92 | 12/01/2001- 10/01/2003 | NA | PSA50 response rate | Not reported | Not reported | ADT: 100 |
| **Eymard JC et al**, 2007 | Rx: Docetaxel70 + Estramustine Ctrl: Docetaxel | 90/ 92 | 02/09/2001- 04/01/2003 | 17.7 | PSA50 response rate | 19.3 vs. 17.8 | 5.7 vs. 2.9 ^†^ | ADT: 100 |
|  |  |  |  |  |  | NA | NA |  |
| **Chemotherapy+Curcuminoid** | | | | | | | | |
| **Jahanmohan JP et al**, 2021 | Rx: Docetaxel + Curcumin Ctrl: Docetaxel | 100/ 50 | 06/01/2014- 07/01/2016 | NA | Time to disease progression | 15.8 vs. 19.8 | 3.7 vs. 5.3 * | ADT: 100 |
|  |  |  |  |  |  | NA | NA |  |
| **Chemotherapy+Immunotherapy** | | | | | | | | |
| **Dahut WL et al**, 2004 | Rx: Docetaxel30 (W) ^d^ + Thalidomide Ctrl: Docetaxel30 (W) ^d^ | 75/ 75 | 12/01/1999- 10/01/2001 | 26.4 | PFS; OS | 28.9 vs. 14.7 | 5.9 vs. 3.7 * | ADT: 100 |
|  |  |  |  |  |  | NA | NA |  |
| **ECOG 3899**, 2010 | Rx1: 13-cis Retinoic acid + Interferon-alpha2b + Paclitaxel Rx2: Vinorelbine + Mitoxantrone + Estramustine | 70/ 70 | 01/31/2001- 10/07/2003 | 59 | PSA50 response rate | 13.9 vs. 19.4 | 2.5 vs. 5.9 * | ADT: 100 |
|  |  |  |  |  |  | NA | NA |  |
| **E1809**, 2015 | Rx: Docetaxel + PVAC-VF Ctrl: Docetaxel | 135/ 10 | 12/01/2010- 03/01/2012 | 20.5 | OS | 20.8 vs. NR | Not reported | ADT: 100 |
|  |  |  |  |  |  | NA |  |  |
| **de Bono et al**, 2014 | Rx: Docetaxel + Figitumumab Ctrl: Docetaxel | 100/ 204 | 10/01/2006- 07/01/2009 | NA | PSA50 response rate | Not reported | 4.9 vs. 7.9 * | ADT: 100 |
|  |  |  |  |  |  |  | 1.44 (1.06-1.96) |  |
| **Heidenreich A et al,**  2013 | Rx: Docetaxel  + Intetumumab Ctrl: Docetaxel | 120/ 131 | 05/01/2007- 12/01/2009 | NA | PFS | 17.2 vs. 20.6 | 7.6 vs. 11 * | ADT: 100 |
|  |  |  |  |  |  | 1.47 (0.85-2.52) | 1.73 (1.11-2.69) |  |
| **Chemotherapy+Radiopharmaceutical** | | | | | | | | |
| **Taxium II**, 2017 | Rx: Docetaxel  + Rhenium-188-HEDP Ctrl: Docetaxel | 88/ 88 | 08/01/2012- 11/01/2014 | 18.4 | PFS | 23.7 vs. 21 | 9.8 vs. 8.6 * | ADT: 100 |
|  |  |  |  |  |  | NA | NA |  |
| **Chemotherapy+TKI** | | | | | | | | |
| **Horti J et al**, 2009 | Rx: Docetaxel + Vandetanib Ctrl: Docetaxel | 80/ 86 | 01/24/2006- 11/24/2006 | NA | PSA50 response rate | Not reported | 7.7 vs. 9.8 ^†^ | ADT: 100 |
|  |  |  |  |  |  |  | NA |  |
| **Chemotherapy+Vascular disrupting agent** | | | | | | | | |
| **Pili R et al,**  2010 | Rx: Docetaxel + Vadimezan Ctrl: Docetaxel | 66/ 71 | NA | 24 | Safety | 17 vs. 17.2 | 8.7 vs. 8.4 ^†^ | ADT: 100 |
|  |  |  |  |  |  | 0.80 (0.46-1.39) | 0.81 (0.39-1.70) |  |
| **Chemotherapy+VitD-Analog** | | | | | | | | |
| **Attia S et al**, 2008 | Rx: Docetaxel35 (W) ^e^ + Doxercalciferol Ctrl: Docetaxel35 (W) ^e^ | 66/ 70 | 10/01/2002- 07/01/2005 | 17.6 | PSA50 response rate | 17.8 vs. 16.4 | 6.2 vs. 6.2 * | ADT: 100 |
|  |  |  |  |  |  | NA | NA |  |
| **ASCENT**, 2007 | Rx: Docetaxel36 (W) + Calcitriol Ctrl: Docetaxel36 (W) | 232/ 250 | 09/01/2002- 01/01/2004 | 18.3 | PSA50 response rate | NR vs. 16.4 | Not reported | ADT: 100 |
|  |  |  |  |  |  | 0.67 (0.45-0.97) ^§^ |  |  |
| **Corticosteroid monotherapy** | | | | | | | | |
| **Venkitaraman R et al**, 2015 | Rx: Dexamethasone Ctrl: Prednisolone | 72/ 82 | 04/01/2006- 07/01/2010 | 11 | PSA50 response rate | Not reported | Not reported | ADT: 100 |
| **Corticosteroid+Immunotherapy** | | | | | | | | |
| **Yoshimura K et al**, 2016 | Rx: Dexamethasone + PPV Ctrl: Dexamethasone | 80/ 73 | 04/01/2008- 10/01/2013 | NA | PSA-PFS | 73.9 vs. 34.9 | Not reported | ADT: 100 |
|  |  |  |  |  |  | 0.41 (0.21-0.83) ^§^ |  |  |
| **Corticosteroid+VEGFi** | | | | | | | | |
| **Stadler WM et al**, 2004 | Rx: Dexamethasone + SU5416 Ctrl: Dexamethasone | 60/ 36 | 07/01/2000- 04/01/2002 | NA | Time to disease progression | Not reported | 2.3 vs. 1 ^†^ | ADT: 100 |
|  |  |  |  |  |  |  | NA |  |
| **ERA monotherapy** | | | | | | | | |
| **James ND et al**, 2009 | Rx1: Zibotentan15  Rx2: Zibotentan10 Ctrl: Placebo | 260/ 312 | 07/14/2004 onwards | Rx1: 21.8 Rx2: 22.7 Ctrl: 17.3 | Time to disease progression | Rx1 vs. Ctrl: 23.9 vs. 19.9 Rx2 vs. Ctrl: 23.5 vs. 19.9 | Rx1 vs. Ctrl: 3.8 vs. 3.7 ^†^ Rx2 vs. Ctrl: 4.6 vs. 3.7 ^†^ | ADT: 100 |
|  |  |  |  |  |  | Rx1 vs. Ctrl: 0.76 (0.61-0.94) ^§^ Rx2 vs. Ctrl: 0.83 (0.67-1.02) | Rx1 vs. Ctrl: 0.86 (0.72-1.04) Rx2 vs. Ctrl: 1.06 (0.89-1.27) |  |
| **Carducci MA et al**, 2003 | Rx1: Atrasentan2.5 Rx2: Atrasentan10 Ctrl: Placebo | Not specified/ 288 | 02/01/1998- 12/01/1998 | NA | Time to disease progression | Not reported | Rx1 vs. Ctrl: 5.9 vs. 4.5 ^‡^ Rx2 vs. Ctrl: 6 vs. 4.5 ^‡^ | ADT: 100 |
|  |  |  |  |  |  |  | NA |  |
| **IMiD monotherapy** | | | | | | | | |
| **Pili R et al**, 2011 | Rx: Tasquinimod Ctrl: Placebo | 200/ 206 | 12/01/2007- 06/01/2009 | 37 | PFS; Progression-free proportion at 6 months | 33.4 vs. 30.4 | 8.8 vs. 4.4 ^†^ | ADT: 100 |
|  |  |  |  |  |  | 0.87 (0.59-1.29) | 0.54 (0.36-0.82) |  |
| **Immunotherapy monotherapy** | | | | | | | | |
| **PERSEUS**, 2016 | Rx1: Abituzumab750 + SOC Rx2: Abituzumab1500 + SOC Ctrl: SOC | 165/ 180 | 04/01/2011- 12/01/2012 | NA | PFS | Not reported | Rx1 vs. Ctrl: 3.4 vs. 3.3 * Rx2 vs. Ctrl: 4.3 vs. 3.3 * | ADT: 100 |
|  |  |  |  |  |  |  | Rx1 vs. Ctrl: 0.89 (0.57-1.39) Rx2 vs. Ctrl: 0.81 (0.52-1.26) |  |
| **Figg WD et al**, 2001 | Rx: ThalidomideHD Ctrl: Thalidomide | Not specified/ 63 | NA | NA | ORR; PSA50 response rate | Not reported | Not reported | ADT: 100 |
| **Immunotherapy+Chemotherapy** | | | | | | | | |
| **Arlen PM et al,** 2006 | Rx: Vaccine + GMCSF + Docetaxel Ctrl: Vaccine + GMCSF | 28/ 28 | NA | NA | Immune response | Not reported | 3.2 vs. 1.8 * | ADT: 100 |
|  |  |  |  |  |  |  | NA |  |
| **Immunotherapy+GMCSF** | | | | | | | | |
| **TBC-PRO-002**, 2010 | Rx: PVAC+GMCSF Ctrl: Placebo | 120/ 125 | 11/01/2003- 07/01/2005 | 41.3 | PFS | 25.1 vs. 16.6 | 3.8 vs. 3.7 * | ADT: 100 |
|  |  |  |  |  |  | 0.56 (0.37-0.85) ^§^ | 0.88 (0.57-1.38) |  |
| **Integrin inhibitor monotherapy** | | | | | | | | |
| **Bradley DA et al,** 2011 | Rx: Cilengitide500 Ctrl: Cilengitide2000 | 106/ 44 | 01/17/2005- 01/24/2007 | NA | Objective clinical progression at 6 months | Not reported | 2.7 vs. 2.8 ^†^ | ADT: 100 |
|  |  |  |  |  |  |  | NA |  |
| **Matrix Metalloproteinase Inhibitor monotherapy** | | | | | | | | |
| **Lara Jr PN et al**, 2006 | Rx: BMS-275291 [1200mg] Ctrl: BMS-275291 [2400mg] | 68/ 80 | 05/01/2002- 07/01/2003 | 15 | PFS at 4 months | NR vs. 21 | Not reported | ADT: 100 |
|  |  |  |  |  |  | NA |  |  |
| **Progestin monotherapy** | | | | | | | | |
| **Patel SR et al**, 1990 | Rx: Megestrol Acetate Ctrl: Dexamethasone | Not specified/ 58 | 05/01/1985- 06/01/1986 | NA | OS | 8.8 vs. 8.1 | Not reported | ADT: 100 |
|  |  |  |  |  |  | NA |  |  |
| **Somatostatin+Corticosteroid** | | | | | | | | |
| **Dimopoulos MA et al**, 2004 | Rx: Lanreotide + Dexamethasone Ctrl: Estramustine + Etoposide | Not specified/ 40 | NA | NA | PSA50 response rate | 18 vs. 18.8 | 4 vs. 6 * | ADT: 100 |
|  |  |  |  |  |  | NA | NA |  |
| **TKI monotherapy** | | | | | | | | |
| **Boccardo F et al**, 2008 | Rx: Gefitinib Ctrl: Placebo | 74/ 82 | 10/01/2002- 11/01/2003 | 29 | PSA50 response rate | 26.5 vs. 20.5 | 4 vs. 4.5 * | ADT: 100 |
|  |  |  |  |  |  | 0.69 (0.39-1.23) | 0.74 (0.46-1.18) |  |
| **TKI+1st gen AA** | | | | | | | | |
| **Azad AA et al**, 2014 | Rx: Vandetanib + Bicalutamide Ctrl: Bicalutamide | Not specified/ 39 | 01/27/2009- 01/31/2011 | NA | PSA50 response rate | Not reported | Not reported | ADT: 100 |
| **Sridhar SS et al**, 2015 | Rx1: Pazopanib + Bicalutamide Rx2: Pazopanib | Not specified/ 23 | 09/01/2007- 03/01/2011 | NA | PSA50 response rate | Not reported | 7.3 vs. 7.3 * | ADT: 100 |
|  |  |  |  |  |  |  | NA |  |

Abbreviations: ADT: androgen deprivation therapy; OS: overall survival; PFS: progression-free survival; ORR: objective response rate; ARPI: androgen-receptor pathway inhibitor; APC: antigen-presenting cell; PA2024: prostatic acid phosphatase-sargramostim fusion protein; cSipuleucel-T: concurrent sipuleucel-T; sSipuleucel-T: sequential sipuleucel-T; TKI: tyrosine kinase inhibitor; ASO: antisense oligonucleotide; Vin/Estra: vinblastine/estramustine; Keto: ketoconazole; Bcl-2: B-cell lymphoma 2 protein; PVAC: PROSTVAC (viral vector-based immunotherapy); VitD: vitamin d; VEGFi: vascular endothelial growth factor inhibitor; ERA: endothelin receptor antagonist; IMiD: immunomodulatory drug; SOC: standard of care; GMCSF: granulocyte-macrophage colony-stimulating factor; AA: antiandrogen; PSA: prostate-specific antigen

Note: The color ‘green’ represents a positive trial, ‘red’ represents a negative trial.

* Composite progression free survival

^†^ Radiographic progression free survival

^‡^ Time to disease progression

^§^ Statistically significant overall survival benefit was observed

Itraconazole600: Itraconazole 600mg/day PO

Itraconazole200: Itraconazole 200mg/day PO

L-Doxorubicin25: Liposomal Doxorubicin (dissolved in 250 mL 5% glucose) 25 mg/m^2^ IV every 2 weeks for 12 consecutive cycles

L-Doxorubicin50: Liposomal Doxorubicin (dissolved in 250 mL 5% glucose) 50 mg/m^2^ IV every 4 weeks for 6 consecutive cycles

Docetaxel30 (W): Docetaxel 30 mg/m^2^ IV every week for 5 weeks, followed by 1 week rest (6-week cycle)

Docetaxel25 (W): Docetaxel 25 mg/m^2^ IV every week for 2 cycles (8-week cycle)

Docetaxel35 (W): Docetaxel 35 mg/m^2^ IV on days 2 and 9 every 3 weeks

Docetaxel70: Docetaxel 70 mg/m^2^ on day 2 every 3 weeks

Estramustine840: Estramustine 840 mg/day on days 1-5

Estramustine420: Estramustine 420 mg/day on days 1-3

Docetaxel35 (D2,D9 Q3W): Docetaxel 35 mg/m^2^ on day 2 and 9 every 3wks

Docetaxel30 (W) ^d^ : Docetaxel 30 mg/m^2^ IV every week for 3 weeks, followed by 1 week rest (4-week cycle)

Docetaxel35 (W) ^e^ : Docetaxel 35 mg/m^2^ IV on days 1, 8, and 15 (4-week cycle)

Docetaxel36 (W): Docetaxel 36 mg/m^2^ IV every week for 3 weeks (4-week cycle)

Zibotentan15: Zibotentan 15 mg PO OD

Zibotentan10: Zibotentan 10 mg PO OD

Atrasentan2.5: Atrasentan 2.5 mg PO OD

Atrasentan10: Atrasentan 10 mg PO OD

Abituzumab750: Abituzumab 750 mg every 3 weeks

Abituzumab1500: Abituzumab 1500 mg every 3 weeks

ThalidomideHD: Thalidomide initial dose of 200 mg/day with increments of 200 mg/day every 2 weeks to a maximum dose of 1200 mg

Cilengitide500: Cilengitide 500 mg IV twice weekly (six-week cycle)

Cilengitide2000: Cilengitide 2000 mg IV twice weekly (six-week cycle)

All other drugs were administered in standard doses.

B) **Prior ADT + ARPI**

| **Trial** | **Arm** | **Estimated /Actual Accrual** | **Years of enrollment** | **Median follow up - months** | **Primary Endpoint** | **OS HR (95% CI)** | **PFS HR (95% CI)** | **Prior therapy \| %** |
| --- | --- | --- | --- | --- | --- | --- | --- | --- |
| **ARPI+TKI** | | | | | | | | |
| **Spetsieris N et al**, 2021 | Rx: Abiraterone + Sunitinib Ctrl: Abiraterone + Dasatinib | 180/ 179 | 03/01/2011- 02/01/2015 | NA | TTF | 22.9 vs. 20.8 | 5.5 vs. 5.7 * | ADT: 100 Docetaxel: 25 ARPI: 100 |
|  |  |  |  |  |  | 1.04 (0.72-1.49) | 0.85 (0.59-1.22) |  |
| **Bipolar Androgen Therapy** | | | | | | | | |
| **TRANSFORMER**, 2021 | Rx: BAT Ctrl: Enzalutamide | 194/ 195 | 04/01/2015- 04/01/2018 | 31.9 | PFS | 32.9 vs. 29 | 6.1 vs. 8.3 ^†^ | ADT: 100 Docetaxel: 12 ARPI: 100 |
|  |  |  |  |  |  | 0.95 (0.66-1.39) | 1.24 (0.87-1.77) |  |
| **Chemotherapy+ARPI** | | | | | | | | |
| **ABIDO SOGUG**, 2022 | Rx: Docetaxel + Abiraterone Ctrl: Docetaxel | 160/ 148 | 02/01/2014- 07/01/2016 | Rx: 15 Ctrl: 11.4 | rPFS rate at 12 months | 17.4 vs. 16.9 | 8.8 vs. 9.8 ^‡^ | ADT: 100 ARPI: 100 |
|  |  |  |  |  |  | NA | NA |  |
| **Chemotherapy+Chemotherapy** | | | | | | | | |
| **MDACC Study**, 2019 | Rx: Cabazitaxel25 + Carboplatin Ctrl: Cabazitaxel25 | 160/ 160 | 08/17/2012- 05/11/2015 | 31 | PFS | 18.5 vs. 17.3 | NA | ADT ARPI |
|  |  |  |  |  |  | 0.89 (0.63-1.25) | 0.62 (0.42-0.90) * |  |

Abbreviations: ADT: androgen deprivation therapy; OS: overall survival; PFS: progression-free survival; ARPI: androgen-receptor pathway inhibitor; TKI: tyrosine kinase inhibitor; TTF: time to treatment failure; rPFS: radiographic progression-free survival; BAT: bipolar androgen therapy

Note: The color ‘green’ represents a positive trial, ‘red’ represents a negative trial.

* Time to treatment failure

^†^ Time to disease progression

^‡^ Radiographic progression free survival

Cabazitaxel25: Cabazitaxel 25 mg/m^2^ IV every 3 weeks

All other drugs were administered in standard doses.

C) **Prior ADT + Docetaxel**

| **Trial** | **Arm** | **Estimated /Actual Accrual** | **Years of enrollment** | **Median follow up - months** | **Primary Endpoint** | **OS HR (95% CI)** | **PFS HR (95% CI)** | **Prior therapy \| %** |
| --- | --- | --- | --- | --- | --- | --- | --- | --- |
| **ASO+Chemotherapy** | | | | | | | | |
| **P-06c,**  2011 | Rx1: Custirsen + Docetaxel Rx2: Custirsen  + Mitoxantrone | 40/ 45 | 07/01/2006- 04/01/2007 | 39 | Safety | 15.8 vs. 11.5 | 7.2 vs. 3.4 * | ADT: 100 Docetaxel: 100 |
|  |  |  |  |  |  | NA | NA |  |
| **Aurora kinase inhibitor monotherapy** | | | | | | | | |
| **Meulenbeld HJ et al**, 2012 | Rx1: Danusertib330 Rx2: Danusertib500 | 58/ 88 | 09/01/2007- 10/01/2009 | NA | PSA50 response rate | Not reported | 2.8 vs. 2.8 * | ADT: 100 Docetaxel: 100 |
|  |  |  |  |  |  |  | NA |  |
| **Chemotherapy monotherapy** | | | | | | | | |
| **GETUG-P02,**  2015 | Rx1: Etoposide Rx2: Vinorelbine Rx3: Mitoxantrone | 90/ 92 | 2006-2010 | 10.4 | Palliative beneﬁt rate | 8.4 vs. 12.6 vs. 10.6 | 2 vs. 4.1 vs. 3.9 ^†^ | ADT: 100 Docetaxel: 100 |
|  |  |  |  |  |  | NA | NA |  |
| **CAINTA**, 2023 | Rx: Cabazitaxel ID Ctrl: Cabazitaxel25 | 72/ 73 | 10/01/2014- 04/01/2018 | NA | Clinical feasibility rate | 16.2 vs. 7.3 | 9.5 vs. 4.4 ^†^ | ADT: 100 Docetaxel: 100 ARPI: 67 |
|  |  |  |  |  |  | NA | NA |  |
| **Rosenberg JE et al**, 2007 | Rx: Ixabepilone Ctrl: Mitoxantrone | 80/ 82 | 02/01/2003- 06/01/2005 | NA | PSA50 response rate | 10.4 vs. 9.8 | Not reported | ADT: 100 Docetaxel: 100 |
|  |  |  |  |  |  | NA |  |  |
| **Chemotherapy+Chemotherapy** | | | | | | | | |
| **RECARDO**, 2018 | Rx: Docetaxel + Carboplatin Ctrl: Docetaxel | 150/ 75 | 07/01/2009- 04/01/2013 | 18 | PFS | 18.9 vs. 18.5 | 11.7 vs. 12.7 * | ADT: 100 Docetaxel: 100 |
|  |  |  |  |  |  | NA | NA |  |
| **Chemotherapy+Immunotherapy** | | | | | | | | |
| **Fizazi K et al**, 2012 | Rx: Mitoxantrone + Siltuximab Ctrl: Mitoxantrone | 134/ 97 | 10/01/2006- 02/01/2008 | NA | PFS | 10.2 vs. 13 | 3.2 vs. 7.5 * | ADT: 100 Docetaxel: 100 |
|  |  |  |  |  |  | 1.45 (0.79-2.68) | 1.72 (1.01-2.93) |  |
| **Hakenberg OW et al**, 2019 | Rx: Mitoxantrone  + Olaratumab Ctrl: Mitoxantrone | Not specified/ 123 | 10/01/2010 onwards | NA | PFS | 14.2 vs. 12.8 | 2.3 vs. 2.4 * | ADT: 100 Docetaxel: 100 |
|  |  |  |  |  |  | 1.08 (0.72-1.61) | 1.29 (0.87-1.90) |  |
| **Immunotherapy monotherapy** | | | | | | | | |
| **Filaci G et al**, 2021 | Rx1: GX301 vaccine (8D) Rx2: GX301 vaccine (4D) Rx3: GX301 vaccine (2D) | 120/ 98 | 11/01/2014 onwards | 24 | Immunological response rate; Safety | 22.9 vs. NR vs. 21.5 | 4.9 vs. 5.7 vs. 4.2 * | ADT: 100 Docetaxel: 100 ARPI: 9 |
|  |  |  |  |  |  | NA | NA |  |
| **TKI monotherapy** | | | | | | | | |
| **Droz JP et al**, 2014 | Rx: Nintedanib150 Ctrl: Nintedanib250 | 58/ 81 | 11/01/2005- 01/01/2007 | NA | PSA20 response rate | 8.2 vs. NR | 2.4 vs. 2.5 * | ADT: 100 Docetaxel: 100 |
|  |  |  |  |  |  | NA | NA |  |

Abbreviations: ADT: androgen deprivation therapy; OS: overall survival; PFS: progression-free survival; ASO: antisense oligonucleotide; TKI: tyrosine kinase inhibitor

Note: The color ‘green’ represents a positive trial, ‘red’ represents a negative trial.

* Composite progression free survival

^†^ Time to disease progression

Danusertib330: Danusertib 330 mg/m^2^ IV on days 1,8 and 15 every 4 weeks

Danusertib500: Danusertib 500 mg/m^2^ IV on days 1 and 15 every 4 weeks

Cabazitaxel ID: Cabazitaxel at an initial dose of 25 mg/m^2^ followed by dose adaptations in cycle 2 and beyond according to a prespecified dosing algorithm considering previous-cycle hematologic toxicity and cabazitaxel AUC (with a target AUC of 0.8–1.2 mg*hour/L)

Cabazitaxel25: Cabazitaxel 25 mg/m^2^ IV every 3 weeks

GX301 vaccine (8D): GX301, 8 doses on days 1, 3, 5, 7, 14, 21, 35 and 63

GX301 vaccine (4D): GX301, 4 doses on days 1, 14, 35 and 63

GX301 vaccine (2D): GX301, 2 doses on days 1 and 63

Nintedanib150: Nintedanib 150 mg PO BID for 6 months

Nintedanib250: Nintedanib 250 mg PO BID for 6 months

All other drugs were administered in standard doses.

D) **Prior ADT + Docetaxel + ARPI**

| **Trial** | **Arm** | **Estimated /Actual Accrual** | **Years of enrollment** | **Median follow up - months** | **Primary Endpoint** | **OS HR (95% CI)** | **PFS HR (95% CI)** | **Prior therapy \| %** |
| --- | --- | --- | --- | --- | --- | --- | --- | --- |
| **ARPI monotherapy** | | | | | | | | |
| **SAK-08-16**, 2023 | Rx: Darolutamide Ctrl: Placebo | 88/ 92 | 04/20/2017- 11/19/2020 | 18 | rPFS at 12 weeks | 24 vs. 21.3 | 5.5 vs. 4.5 * | ADT: 100 Docetaxel: 91 ARPI: 100 |
|  |  |  |  |  |  | 0.62 (0.30-1.26) | 0.54 (0.32-0.91) |  |
| **Chemotherapy monotherapy** | | | | | | | | |
| **ConCab**, 2018 | Rx: Cabazitaxel (W) Ctrl: Cabazitaxel25 | 100/ 101 | 04/19/2012- 10/21/2015 | 21 | Dose intensity | 15.6 vs. 14.6 | 6.4 vs. 6 ^†^ | ADT: 100 Docetaxel: 100 ARPI: 84 |
|  |  |  |  |  |  | NA | 0.73 (0.47-1.13) |  |
| **Chemotherapy+AKTi** | | | | | | | | |
| **RE-AKT**, 2024 | Rx: Enzalutamide  + Capivasertib Ctrl: Enzalutamide | 136/  100 | 06/07/2016-06/09/2019 | 43 | ORR;  PSA50 response rate;  CTC count conversion | 13.9 vs. 11 | 5.6 vs. 3.5 ^‡^ | ADT: 100 Docetaxel: 100 ARPI: 100 |
|  |  |  |  |  |  | 0.76 (0.51-1.15) | 0.78 (0.47-1.30) |  |
| **Radioligand monotherapy** | | | | | | | | |
| **TheraP**, 2021 | Rx: 177Lu Ctrl: Cabazitaxel20 | 200/ 200 | 02/06/2018- 09/03/2019 | 35.7 | PSA50 response rate | 16.4 vs. 19.4 | NA | ADT: 100 Docetaxel: 100 ARPI: 91 |
|  |  |  |  |  |  | 0.97 (0.70-1.35) | 0.64 (0.46-0.88) * |  |

Abbreviations: ADT: androgen deprivation therapy; OS: overall survival; PFS: progression-free survival; ARPI: androgen-receptor pathway inhibitor; rPFS: radiographic progression-free survival; AKTi: protein kinase B inhibitor; 177Lu: 177Lu-PSMA-617

Note: The color ‘green’ represents a positive trial, ‘red’ represents a negative trial.

* Time to disease progression

^†^ Composite progression free survival

^‡^ Radiographic progression free survival

Cabazitaxel (W): Cabazitaxel 25 mg/m^2^ IV weekly for 5 weeks (6-week cycle)

Cabazitaxel25: Cabazitaxel 25 mg/m^2^ IV every 3 weeks

Cabazitaxel20: Cabazitaxel 20 mg/m^2^ IV every 3 weeks

Remaining drugs were administered at standard doses.

E) **Heterogeneous Prior Therapy**

| **Trial** | **Arm** | **Estimated /Actual Accrual** | **Years of enrollment** | **Median follow up - months** | **Primary Endpoint** | **OS HR (95% CI)** | **PFS HR (95% CI)** | **Prior therapy \| %** |
| --- | --- | --- | --- | --- | --- | --- | --- | --- |
| **ARPI+Radioligand** | | | | | | | | |
| **ENZA-p**, 2024 | Rx: Enzalutamide + 177Lu Ctrl: Enzalutamide | 160/ 162 | 08/17/2020- 07/26/2022 | 20 | PSA-PFS | 34 vs. 26 | NA | ADT: 100 Docetaxel: 55 ARPI: 13 |
|  |  |  |  |  |  | 0.55 (0.36-0.84) * | 0.61 (0.42-0.87) ^†^ |  |
| **Chemotherapy monotherapy** | | | | | | | | |
| **TAXYNERGY**, 2017 | Rx: Docetaxel Ctrl: Cabazitaxel25 | 60/ 63 | 11/01/2012- 06/01/2014 | 14 | PSA50 response rate | Not reported | Not reported | ADT: 100 ARPI: 44 |
| **Chemotherapy+Immunotherapy** | | | | | | | | |
| **IND 209**, 2018 | Rx: Docetaxel + Reolysin Ctrl: Docetaxel | 80/ 85 | 06/01/2012- 09/01/2015 | NA | LPD rate at 12 weeks | 19.1 vs. 21.1 | Not reported | ADT: 100 ARPI: 54 |
|  |  |  |  |  |  | 1.83 (0.96-3.52) |  |  |
| **Kongsted P et al,**  2017 | Rx: Docetaxel + DCVAC Ctrl: Docetaxel | 40/ 43 | 2011-2015 | 46.3 | Immune response | Not reported | 5.7 vs. 5.5 ^‡^ | ADT: 100 ARPI: 28 |
|  |  |  |  |  |  |  | NA |  |
| **PARPi+ARPi** | | | | | | | | |
| **BRCAAway**, 2024 | Rx1: Abiraterone Rx2: Olaparib Rx3: Abiraterone  + Olaparib | 60/ 61 | 02/01/2017- 03/01/2022 | 18 | PFS | Not reported | Rx3 vs. Rx1: 39 vs. 8.6 ^‡^ Rx3 vs. Rx2: 39 vs. 14 ^‡^ | ADT: 100 Docetaxel: 26 ARPI: 3 |
|  |  |  |  |  |  |  | Rx3 vs. Rx1: 0.33 (0.15-0.72) Rx3 vs. Rx2: 0.37 (0.17-0.84) |  |
| **TKI monotherapy** | | | | | | | | |
| **Monk P et al**, 2018 | Rx: Tivantinib Ctrl: Placebo | 78/ 78 | 01/01/2012- 09/01/2013 | 8.9 | PFS | Not reported | 5.5 vs. 3.7 ^‡^ | ADT: 100 ARPI: 29 |
|  |  |  |  |  |  |  | 0.55 (0.33-0.90) |  |
| **Smith DC et al**, 2013 | Rx: Cabozantinib Ctrl: Placebo | 200/ 170 | 10/01/2009- 02/01/2011 | NA | PFS; ORR at 12 weeks | Not reported | 5.5 vs. 1.4 ^§^ | ADT: 100 ARPI: ^¶^ Docetaxel: ^¶^ |
|  |  |  |  |  |  |  | NA |  |

Abbreviations: ADT: androgen deprivation therapy; OS: overall survival; PFS: progression-free survival; ARPI: androgen-receptor pathway inhibitor; 177Lu: 177Lu-PSMA-617; LPD: lack of disease progression; PARPi: poly(ADP-ribose) polymerase inhibitor; TKI: tyrosine kinase inhibitor; ORR: objective response rate

Note: The color ‘green’ represents a positive trial, ‘red’ represents a negative trial.

^*^ Statistically significant overall survival benefit was observed

^†^ Radiographic progression free survival

^‡^ Composite progression free survival

^§^ Time to disease progression

^¶^ Received but number not specified

Cabazitaxel25: Cabazitaxel 25 mg/m^2^ IV every 3 weeks

Remaining drugs were administered at standard doses.

F) **Trials that reported data for multiple subgroups**

| **Trial** | **Arm** | **Estimated /Actual Accrual** | **Years of enrollment** | **Median follow up - months** | **Primary Endpoint** | **OS HR (95% CI)** | **PFS HR (95% CI)** | **Prior therapy \| %** |
| --- | --- | --- | --- | --- | --- | --- | --- | --- |
| **Chemotherapy monotherapy** | | | | | | | | |
| **Annala M et al**, 2021 | Rx: Cabazitaxel25 Ctrl: Enzalutamide /Abiraterone | 112/ 95 | 11/24/2014- 12/29/2017 | 21.9 | Clinical benefit rate | *Prior ADT:* | | ADT: 100 Docetaxel:  26 (mHSPC) 27 (mCRPC) |
|  |  |  |  |  |  | NA | NA |  |
|  |  |  |  |  |  | 0.57 (0.23-1.42) | 0.72 (0.38-1.37) * |  |
|  |  |  |  |  |  | *Prior ADT+Docetaxel:* | |  |
|  |  |  |  |  |  | NA | NA |  |
|  |  |  |  |  |  | 0.57 (0.26-1.27) | 0.97 (0.53-1.79) * |  |
|  |  |  |  |  |  | *Heterogeneous Prior Therapy:* | |  |
|  |  |  |  |  |  | 37 vs. 15.5 | 5.3 vs. 2.8 ^†^ |  |
|  |  |  |  |  |  | 0.58 (0.32-1.05) | 0.87 (0.56-1.35) |  |
| **Chemotherapy+AKTi** | | | | | | | | |
| **ProCAID**, 2021 | Rx: Docetaxel + Capivasertib Ctrl: Docetaxel | 150/ 150 | 09/10/2015- 01/31/2019 | Rx: 35 Ctrl: 32 | PFS | *Prior ADT+ARPI:* | | ADT: 100 Docetaxel: 26 ARPI: 3 |
|  |  |  |  |  |  | NA | NA |  |
|  |  |  |  |  |  | 0.50 (0.29-0.86) ^‡^ | 0.78 (0.51-1.18) ^†^ |  |
|  |  |  |  |  |  | *Heterogeneous Prior Therapy:* | |  |
|  |  |  |  |  |  | 25.3 vs. 20.3 | 7 vs. 6.7 ^†^ |  |
|  |  |  |  |  |  | 0.70 (0.47-1.05) | 0.92 (0.65-1.31) |  |

Abbreviations: ADT: androgen deprivation therapy; OS: overall survival; PFS: progression-free survival; ARPI: androgen-receptor pathway inhibitor; mHSPC: metastatic hormone sensitive prostate cancer; mCRPC: metastatic castration resistant prostate cancer; AKTi: protein kinase B inhibitor

Note: The color ‘green’ represents a positive trial, ‘red’ represents a negative trial.

* Radiographic progression free survival

^†^ Composite progression free survival

^‡^ Statistically significant overall survival benefit was observed

Cabazitaxel25: Cabazitaxel 25 mg/m^2^ IV every 3 weeks

Remaining drugs were administered at standard doses.

### **Supplement Table 7:** Summary of population characteristics of included phase II trials

| **Trial** | **Arm** | **Age in years \| median (range)** | **ECOG PS** | | | **Gleason Score ≥ 8 \|  N (%)** | **Bone Metastasis \| N (%)** | **Nodal Metastasis \| N (%)** | **Visceral Metastasis \| N (%)** | **Liver Metastasis \| N (%)** | **Lung Metastasis \| N (%)** | **Baseline PSA \|  Median (IQR)** |
| --- | --- | --- | --- | --- | --- | --- | --- | --- | --- | --- | --- | --- |
|  |  |  | **0 \| N (%)** | **1 \| N (%)** | **2 \| N (%)** |  |  |  |  |  |  |  |
| **Prior ADT only** | | | | | | | | | | | | |
| **Antiandrogen+Chemotherapy** | | | | | | | | | | | | |
| **Takahashi M et al**, 2013 | Rx: Antiandrogen + Tegafur-Uracil Ctrl: Antiandrogen | Rx: 76.5  (47-92)  Ctrl: 77  (59-89) | NA | NA | NA | 38 (73) | NA | NA | NA | NA | NA | Rx: 311 (4.7-3454) *   Ctrl: 98.5  (5.8-15168) * |
| **Antifungal monotherapy** | | | | | | | | | | | | |
| **Antonarakis ES et al**, 2013 | Rx: Itraconazole600 Ctrl: Itraconazole200 | Rx: 71  (52-89) Ctrl: 73  (60-81) | 29 (63) | 17 (37) | | NA | 35 (76) | NA | 31 (67) | NA | NA | Rx: 43.5  (2.6–234.5) * Ctrl: 29.2  (7.0–1989.5) * |
| **Antifungal+Bisphosphonate** | | | | | | | | | | | | |
| **Figg WD et al,** 2005 | Rx: Ketoconazole + Alendronate Ctrl: Ketoconazole | Rx: 72  (51–85) Ctrl: 70  (51–79) | 19 (26) | 48 (67) | 5 (7) | NA | NA | NA | NA | NA | NA | Rx: 78.1  (5.5–1562) * Ctrl: 74.4  (1.7–1458) * |
| **Antifungal+Chemotherapy** | | | | | | | | | | | | |
| **Millikan R et al**, 2001 | Rx1: Ketoconazole + Doxorubicin Rx2: Ketoconazole | Rx1: 70.1  (48-85) Rx2: 70.9  (51-82) | NA | NA | NA | NA | 71 (80) | NA | 15 (17) | NA | NA | Rx: 200  (11–2,335) * Ctrl: 98  (5–3255) * |
| **ARPI monotherapy** | | | | | | | | | | | | |
| **TERRAIN**, 2016 | Rx: Enzalutamide Ctrl: Bicalutamide | Rx: 71  (50−96) Ctrl: 71  (48−91) | 276 (74) | 99 (26) | NA | 212 (57) | 175 (47) | NA | NA | NA | NA | Rx: 21  (0.6−5000) * Ctrl: 22  (0.1−4681) * |
| **ARPI+ARPI** | | | | | | | | | | | | |
| **Khalaf DJ et al**, 2019 | Rx1: Abiraterone followed by Enzalutamide Rx2: Enzalutamide followed by Abiraterone | Rx1: 72.9  (51.3–93.3) Rx2: 77.6  (49.3–94.1) | 168 (83) | | NA | NA | 167 (83) | NA | NA | 12 (6) | 17 (8) | Rx1: 35.0  (2.2–2817.0) * Rx2: 37.0  (1.7–1060.0) * |
| **ARPI+Immunotherapy** | | | | | | | | | | | | |
| **STAMP**, 2015 | Rx1: Abiraterone + cSipuleucel-T Rx2: Abiraterone + sSipuleucel-T | Rx1: 69  (55–91) Rx2: 68.5  (49–90) | 54 (78) | 15 (22) | NA | 38 (55) | 56 (81) | NA | NA | NA | NA | Rx1: 36.2  (13.5-87.7) Rx2: 22.4  (9.3-51.8) |
| **STRIDE**, 2023 | Rx1: Enzalutamide + cSipuleucel-T  Rx2: Enzalutamide + sSipuleucel-T | Rx1: 66  (43–86) Rx2: 72  (55–88) | 41 (79) | 11 (21) | NA | 35 (67) | NA | NA | NA | NA | NA | Rx1: 10.9  (5.6-32.1) Rx2: 10.8  (5.3-39.3) |
| **ARPI+TKI** | | | | | | | | | | | | |
| **Dorff TB et al**, 2019 | Rx: Abiraterone  + Dasatinib Ctrl: Abiraterone | Rx: 66.5  (55.7–85.3) Ctrl: 66.4  (55.6–75.9) | 22 (85) | 4 (15) | NA | NA | 12 (46) | 6 (23) | NA | NA | NA | Rx: 19.8  (0.8–184.1) * Ctrl: 22.2  (0.9–1387) * |
| **ASO monotherapy** | | | | | | | | | | | | |
| **Yu EY et al**, 2018 | Rx: Apatorsen Ctrl: Prednisone | Rx: 67  (53-86) Ctrl: 73  (31-89) | 48 (65) | 26 (35) | NA | 32 (43) | NA | NA | NA | NA | NA | Rx: 57  (0.4-1530) * Ctrl: 51.5  (0.1-606.2) * |
| **Tolcher AW et al,** 2002 | Rx: ISIS 3521 Ctrl: ISIS 5132 | Rx: 72  (58–78) Ctrl: 66  (52–81) | 17 (57) | 13 (43) | NA | NA | 26 (87) | 10 (33) | NA | NA | NA | NA |
| **Chemotherapy monotherapy** | | | | | | | | | | | | |
| **Heidenreich A et al**, 2004 | Rx1: L-Doxorubicin25  Rx2: L-Doxorubicin50 | Rx: 69.3  (11.5) ^†^  Ctrl: 67.5  (11.2) ^†^ | NA | NA | NA | NA | NA | NA | NA | NA | NA | Rx1: 685.2  (8–6340) * Rx2: 670.3  (12–5947) * |
| **TIPC**, 2007 | Rx: Docetaxel30 (W) Ctrl: Prednisolone | Rx: 70  (52-81) Ctrl: 72  (54-84) | 54 (40) | 42 (31) | 13  (10) | NA | 96 (72) | NA | NA | NA | NA | Rx: 130  (14–1681) * Ctrl: 163  (14–2551) * |
| **Krainer M et al**, 2007 | Rx: Docetaxel25 (W) Ctrl: Vinorelbine | Rx: 65.5  (54-82) Ctrl: 69  (55-83) | NA | NA | NA | NA | 30 (79) | NA | NA | NA | NA | Rx: 100.5  (21-834) * Ctrl: 53.2  (0-1536) * |
| **Chemotherapy+Antifungal** | | | | | | | | | | | | |
| **Millikan R et al,**  2003 | Rx: Vin/Estra + Keto/Doxorubicin Ctrl: Paclitaxel + Estramustine + Etoposide | Rx: 68  (51-88) Ctrl: 67  (49-83) | 54 (76) | | 17  (24) ^\|\|^ | NA | 45 (63) | 16 (23) | 10 (14) | NA | NA | Rx: 79  (7.6-8500) * Ctrl: 134  (7.5-2500) * |
| **Chemotherapy+ARPI** | | | | | | | | | | | | |
| **CHEIRON**, 2021 | Rx: Docetaxel + Enzalutamide Ctrl: Docetaxel | Rx: 69  (63.4–75.2) ^‡^  Ctrl: 71  (65.8–75.2) ^‡^ | 160 (65) | 78 (32) | 8 (3) | 130 (53) | 201 (82) | 149 (61) | 58 (24) | NA | NA | Rx: 69  (63.4–75.2) Ctrl: 71  (65.8–75.2) |
| **Chemotherapy+ASO** | | | | | | | | | | | | |
| **Chi KN et al,**  2010 | Rx: Docetaxel + Custirsen Ctrl: Docetaxel | Rx: 69  (54-84) Ctrl: 69  (49-87) | 41 (50) | 41 (50) | NA | 48 (59) | NA | NA | NA | NA | NA | Rx: 110  (5.6-1723.4) * Ctrl: 110  (7.9-1968.6) * |
| **Wiechno P et al**, 2014 | Rx: Docetaxel + LY2181308 Ctrl: Docetaxel | Rx: 69.1  (48-86.9) Ctrl: 70  (41.5-84) | 143 (96) | | 6 (4) | NA | NA | NA | NA | NA | NA | NA |
| **Chemotherapy+Bcl-2 inhibitor** | | | | | | | | | | | | |
| **EORTC**, 2009 | Rx: Docetaxel  + Oblimersen Ctrl: Docetaxel | Rx: 64  (54-84) Ctrl: 69  (43-82) | 66 (59) | 42 (38) | 3 (3) | NA | 43 (39) | NA | 15 (14) | NA | NA | Rx: 99  (10–2469) * Ctrl: 86  (7–2766) * |
| **Sonpavde G et al**, 2011 | Rx: Docetaxel + AT-101 Ctrl: Docetaxel | Rx: 69  (46–86) Ctrl: 69.5  (51–88) | 79 (36) | 129 (59) | 12 (5) | NA | 193 (88) | 105 (48) | 48 (22) | NA | NA | Rx: 90.4  (0.6–3631) * Ctrl: 83.3  (0–13056) * |
| **Chemotherapy+Chemotherapy** | | | | | | | | | | | | |
| **Machiels JP et al**, 2008 | Rx: Docetaxel35 (W) + Estramustine Ctrl: Docetaxel35 (W) | Rx: 70  (46-82) Ctrl: 70  (62-86) | 73 (49) | 60 (40) | 16 (11) | 48 (32) | 134 (90) | 76 (51) | 20 (13) | 12 (8) | 8 (5) | Rx: 76  (10-3865) * Ctrl: 65  (10-1344) * |
| **Berry WR et al**, 2004 | Rx: Paclitaxel + Estramustine Ctrl: Paclitaxel | Rx: 71  (53-89) Ctrl: 71  (50-94) | 54 (33) | 93 (57) | 15 (9) | NA | NA | NA | NA | NA | NA | Rx: 136  (46.6-441.0) Ctrl: 137  (54.4-430.0) |
| **Albrecht W et al**, 2004 | Rx: Estramustine + Vinblastine Ctrl: Estramustine | NA | 15 (17) | 57 (63) | 18 (20) | NA | NA | NA | 25 (28) | NA | NA | NA |
| **Cabrespine A et al**, 2006 | Rx: Paclitaxel + Carboplatin Ctrl: Mitoxantrone | Rx: 68  (52–80) Ctrl: 67  (50–76) | NA | NA | NA | NA | 35 (88) | 20 (50) | 6 (15) | NA | NA | Rx: 95.7  (11.3–2328) * Ctrl: 105  (25.6–1146) * |
| **Nelius T et al**, 2006 | Rx1: Docetaxel70 + Estramustine840 Rx2: Docetaxel70 + Estramustine420 | Rx1: 68 Rx2: 68 | 22 (31) | 37 (51) | 13 (18) | NA | 57 (79) | 31 (43) | 6 (8) | NA | NA | Rx1: 114.6  (0.7-4973) * Rx2: 163.2  (8.9-3899) * |
| **Oudard S et al**, 2005 | Rx1: Docetaxel70 + Estramustine Rx2: Docetaxel35  (D2,D9 Q3W) + Estramustine Ctrl: Mitoxantrone | Rx1: 68  (52-91) ^‡^  Rx2: 68  (51-79) ^‡^ Ctrl: 70  (52-85) ^‡^ | 62 (48) | 43 (33) | 22 (17) | 95 (73) | 118 (91) | 40 (31) | NA | NA | NA | Rx1: 71  (1.9-2818) Rx2: 69.5  (0.01-2416) Ctrl: 77.7  (0.41-1840) |
| **Droz JP et al**, 2003 | Rx1: Oxaliplatin + 5-Fluorouracil Rx2: Oxaliplatin | Rx1: 68 (45-77) Rx2: 68  (44-77) | 14 (26) | 24 (44) | 13 (24) | 23 (43) | 48 (89) | 21 (39) | 16 (30) | NA | NA | Rx1: 254  (18–2763) * Rx2: 85  (1–1651) * |
| **Galsky MD et al**, 2005 | Rx1: Ixabepilone + Estramustine Rx2: Ixabepilone | Rx1: 69  (45-87) Rx2: 72  (54-83) | NA | NA | NA | NA | 76 (83) | NA | NA | NA | NA | Rx1: 63.9  (4.7-391.6) * Rx2: 77.1 ( 1.1-2518.7) * |
| **Eymard JC et al**, 2007 | Rx: Docetaxel70 + Estramustine Ctrl: Docetaxel | Rx: 67.4  (46.2–79.8) Ctrl: 69  (57.3–86.3) | 32 (35) | 50 (55) | 9 (10) | NA | 82 (90) | 34 (37) | NA | 12 (13) | 6 (7) | Rx: 127.8  (0.3-1145) * Ctrl: 79.7  (10.5-1585) * |
| **Chemotherapy+Curcuminoid** | | | | | | | | | | | | |
| **Jahanmohan JP et al**, 2021 | Rx: Docetaxel + Curcumin Ctrl: Docetaxel | Rx: 70  (44-87) Ctrl: 69  (60-80) | NA | 25 (50) | 18 (36) | 17 (34) | 35 (70) | 10 (20) | NA | 3 (6) | 6 (12) | Rx: 50.6  (1.7–1587) * Ctrl: 119  (7.0–1882) * |
| **Chemotherapy+Immunotherapy** | | | | | | | | | | | | |
| **Dahut WL et al**, 2004 | Rx: Docetaxel30 (W) ^f^ + Thalidomide Ctrl: Docetaxel30 (W) ^f^ | Rx: 71  (52–83) Ctrl: 67  (43–82) | 16 (22) | 58 (78) | NA | NA | 66 (89) | NA | NA | NA | NA | Rx: 59.9  (2–1777) * Ctrl: 64.4  (0.2–340.8) * |
| **ECOG 3899**, 2010 | Rx1: 13-cis Retinoic acid + Interferon-alpha2b + Paclitaxel Rx2: Vinorelbine + Mitoxantrone + Estramustine | Rx1: 65  (45-80) Rx2: 70  (50-89) | 32 (51) | 28 (44) | 3 (5) | NA | 57 (91) | NA | 14 (22) | 6 (10) | 8 (13) | NA |
| **E1809**, 2015 | Rx: Docetaxel + PVAC-VF Ctrl: Docetaxel | Rx: 63  (56-72) Ctrl: 65  (65-73) | 7 (70) | 2 (20) | NA | 6 (60) | 8 (80) | 6 (60) | 2 (20) | 1 (10) | 1 (10) | NA |
| **de Bono et al**, 2014 | Rx: Docetaxel + Figitumumab Ctrl: Docetaxel | Rx: 68.9  (7.4) ^†^ Ctrl: 67.9  (7.5) ^†^ | 108 (53) | 86 (42) | 3 (1) | NA | 166 (81) | NA | NA | 25 (13) | 27 (13) | Rx: 105  (6.1–3,683) *  Ctrl: 96.4  (6.3–2124) * |
| **Heidenreich A et al,**  2013 | Rx: Docetaxel  + Intetumumab Ctrl: Docetaxel | Rx: 68 (41-83) Ctrl: 68 (46-82) | 65 (50) | 62 (47) | 4 (3) | 47 (36) | NA | NA | NA | NA | NA | Rx: 96.5  (2.4-1434) *  Ctrl: 66.4  (3.6-8081) * |
| **Chemotherapy+Radiopharmaceutical** | | | | | | | | | | | | |
| **Taxium II**, 2017 | Rx: Docetaxel  + Rhenium-188-HEDP Ctrl: Docetaxel | Rx: 70.5  (54.1–84.9) Ctrl: 71.4  (60–85.8) | 30 (34) | 57 (65) | 1 (1) | 27 (31) | NA | 30 (34) | 8 (9) | NA | NA | Rx: 113.5  (5–4200) *  Ctrl: 187  (16–1101) * |
| **Chemotherapy+TKI** | | | | | | | | | | | | |
| **Horti J et al**, 2009 | Rx: Docetaxel + Vandetanib Ctrl: Docetaxel | Rx: 67  (47–81) Ctrl: 67  (43–79) | NA | NA | NA | NA | NA | NA | NA | NA | NA | NA |
| **Chemotherapy+Vascular disrupting agent** | | | | | | | | | | | | |
| **Pili R et al,**  2010 | Rx: Docetaxel + Vadimezan Ctrl: Docetaxel | Rx: 68  (49-87) Ctrl: 67  (48-84) | 28 (39) | 39 (55) | 3 (4) | NA | 20 (28) | NA | NA | NA | NA | Rx: 73.3  (8-19000) *  Ctrl:78.7  (4-1229) * |
| **Chemotherapy+VitD-Analog** | | | | | | | | | | | | |
| **Attia S et al**, 2008 | Rx: Docetaxel35 (W) ^g^ + Doxercalciferol Ctrl: Docetaxel35 (W) ^g^ | Rx: 72  (56–85) Ctrl: 70  (52–82) | 26 (37) | 40 (57) | 3 (4) | NA | 65 (93) | 32 (46) | NA | NA | NA | Rx: 78.9  (10–1400) *  Ctrl: 82.4  (2.7–1622) * |
| **ASCENT**, 2007 | Rx: Docetaxel36 (W) + Calcitriol Ctrl: Docetaxel36 (W) | Rx: 68  (45-87) Ctrl: 70  (47-92) | 126 (50) | 113 (45) | 11 (5) | NA | 113 (45) | 99 (40) | NA | 16 (6) | NA | Rx: 123  (4-4453) *  Ctrl: 91  (5-6288) * |
| **Corticosteroid monotherapy** | | | | | | | | | | | | |
| **Venkitaraman R et al**, 2015 | Rx: Dexamethasone Ctrl: Prednisolone | Rx: 67  (55–82) Ctrl: 67  (51–82) | 39 (52) | 21 (28) | 4 (5) | 34 (45) | 49 (65) | 32 (43) | 5 (7) | NA | NA | Rx: 34  (6.2–5000) Ctrl: 49  (5.9–4000) |
| **Corticosteroid+Immunotherapy** | | | | | | | | | | | | |
| **Yoshimura K et al**, 2016 | Rx: Dexamethasone + PPV Ctrl: Dexamethasone | Rx: 68 (64–73) ^‡^  Ctrl: 67  (63–67) ^‡^ | NA | NA | NA | 52 (71) | 51 (70) | 23 (32) | NA | NA | 4 (5) | Rx: 4.4  (2.2–6.9) Ctrl: 5.3  (3.5–7.4) |
| **Corticosteroid+VEGFi** | | | | | | | | | | | | |
| **Stadler WM et al**, 2004 | Rx: Dexamethasone + SU5416 Ctrl: Dexamethasone | Overall: 70  (47-84) | 19 (56) | 15 (44) | NA | NA | NA | 10 (29) | NA | NA | NA | NA |
| **ERA monotherapy** | | | | | | | | | | | | |
| **James ND et al**, 2009 | Rx1: Zibotentan15  Rx2: Zibotentan10 Ctrl: Placebo | Rx1: 70  (54–84)  Rx2: 70  (53-85) Ctrl: 72  (49–91) | NA | NA | NA | NA | 312 (100) | NA | NA | NA | NA | Rx1: 72  (0.1–3067) *  Rx2: 55  (2.1–3725) *  Ctrl: 64  (5.3–3776) * |
| **Carducci MA et al**, 2003 | Rx1: Atrasentan2.5 Rx2: Atrasentan10 Ctrl: Placebo | Rx1: 71 Rx2: 72 Ctrl: 72 | NA | NA | NA | NA | NA | NA | NA | NA | NA | Rx1: 67.3 Rx2: 86.4 Ctrl: 94.8 |
| **IMiD monotherapy** | | | | | | | | | | | | |
| **Pili R et al**, 2011 | Rx: Tasquinimod Ctrl: Placebo | Rx: 72.3  (49-89) ^§^ Ctrl: 73.2  (48-89) ^§^ | NA | NA | NA | 75 (37) | 136 (68) | 21 (10) | 42 (21) | NA | NA | Rx: 29 Ctrl: 19 |
| **Immunotherapy monotherapy** | | | | | | | | | | | | |
| **PERSEUS**, 2016 | Rx1: Abituzumab750 + SOC Rx2: Abituzumab1500 + SOC Ctrl: SOC | Rx1: 69.5  (54–84) Rx2: 71  (53-88) Ctrl: 71  (46–88) | 98 (54) | 65 (36) | NA | NA | 173 (96) | NA | NA | 1 (0.5) | 2 (1) | NA |
| **Figg WD et al**, 2001 | Rx: ThalidomideHD Ctrl: Thalidomide | Rx: 65  (57–80) Ctrl: 68  (50–83) | 16 (25) | 44 (70) | 3 (5) | NA | 56 (89) | NA | NA | NA | NA | Rx: 365  (636) ^†^ Ctrl: 315  (450) ^†^ |
| **Immunotherapy+Chemotherapy** | | | | | | | | | | | | |
| **Arlen PM et al,**  2006 | Rx: Vaccine + GMCSF + Docetaxel Ctrl: Vaccine + GMCSF | Rx: 66.5  (56-81) Ctrl: 69.5  (51-85) | NA | NA | NA | 19 (68) | 24 (86) | NA | NA | NA | NA | Rx: 129.1  (17.7-675) *  Ctrl: 61.1  (6.3-1385) * |
| **Immunotherapy+GMCSF** | | | | | | | | | | | | |
| **TBC-PRO-002**, 2010 | Rx: PVAC+GMCSF Ctrl: Placebo | Rx: 71.5  (67-79) ^‡^ Ctrl: 79  (72-83) ^‡^ | 83 (68) | 39 (32) | NA | NA | 114 (93) | 64 (52) | NA | NA | NA | Rx: 36  (17-107) Ctrl: 45  (20-85) |
| **Integrin inhibitor monotherapy** | | | | | | | | | | | | |
| **Bradley DA et al,**  2011 | Rx: Cilengitide500 Ctrl: Cilengitide2000 | Rx: 73  (59–84) Ctrl: 67  (52-85) | 31 (71) | 12 (27) | NA | NA | 16 (36) | NA | NA | NA | NA | Rx: 65  (6–870) *  Ctrl: 26  (5–621) * |
| **Matrix Metalloproteinase Inhibitor monotherapy** | | | | | | | | | | | | |
| **Lara Jr PN et al**, 2006 | Rx: BMS-275291 [1200mg] Ctrl: BMS-275291 [2400mg] | Rx: 69.3 Ctrl: 70.2 | NA | NA | NA | NA | 80 (100) | NA | NA | NA | NA | NA |
| **Progestin monotherapy** | | | | | | | | | | | | |
| **Patel SR et al**, 1990 | Rx: Megestrol Acetate Ctrl: Dexamethasone | NA | NA | NA | NA | NA | NA | NA | NA | NA | NA | NA |
| **Somatostatin+Corticosteroid** | | | | | | | | | | | | |
| **Dimopoulos MA et al**, 2004 | Rx: Lanreotide + Dexamethasone Ctrl: Estramustine + Etoposide | Rx: 74  (60-89) ^§^  Ctrl: 73  (59-86) ^§^ | 29 (76) | | 9 (23) ^\|\|^ | NA | 14 (37) | 11 (29) | NA | 2 (5) | 2 (5) | Rx: 103.5  (8.5–4290) *  Ctrl: 56  (0.1–823) * |
| **TKI monotherapy** | | | | | | | | | | | | |
| **Boccardo F et al**, 2008 | Rx: Gefitinib Ctrl: Placebo | Rx: 75.5  (63–89) Ctrl: 74  (55–86) | 70 (85) | 12 (15) | | NA | 70 (85) | 4 (5) | NA | 1 (1) | NA | Rx: 52.1  (5.34–3351.7) *  Ctrl: 44.4  (7.56–579.2) * |
| **TKI+1st gen AA** | | | | | | | | | | | | |
| **Azad AA et al**, 2014 | Rx: Vandetanib + Bicalutamide Ctrl: Bicalutamide | Rx: 69.8 ^†^ Ctrl: 70.4 ^†^ | 32 (82) | 7 (18) | NA | 17 (44) | 28 (72) | 19 (49) | 8 (21) | 1 (3) | 7 (18) | Rx: 54  (4.1-440.7) ^§^  Ctrl: 28.5  (2.5-614.7) ^§^ |
| **Sridhar SS et al**, 2015 | Rx1: Pazopanib + Bicalutamide Rx2: Pazopanib | Overall: 71  (51-85) | ~100% | |  | NA | 5 (22) | 2 (9) | 2 (9) | NA | NA | Overall: 136  (11-1200) * |
| **Prior ADT+ARPI** | | | | | | | | | | | | |
| **ARPI+TKI** | | | | | | | | | | | | |
| **Spetsieris N et al**, 2021 | Rx: Abiraterone + Sunitinib Ctrl: Abiraterone + Dasatinib | Rx: 67  (48–85) Ctrl: 68  (52–80) | NA | NA | NA | 83 (63) | NA | NA | 19 (14) | NA | NA | Rx: 28.5  (0.8 – 1195.6) *  Ctrl: 19.4  (0.8 – 627.8) * |
| **Bipolar Androgen Therapy** | | | | | | | | | | | | |
| **TRANSFORMER**, 2021 | Rx: BAT Ctrl: Enzalutamide | Rx: 71  (45-87) Ctrl: 71  (49-91) | 126 (65) | 65 (33) | 2 (1) | 114 (58) | NA | NA | 114 (58) | NA | NA | Rx: 44.3  (1.1-323.3) ^§^  Ctrl: 50.6  (1.1-559.2) ^§^ |
| **Chemotherapy+ARPI** | | | | | | | | | | | | |
| **ABIDO SOGUG**, 2022 | Rx: Docetaxel + Abiraterone Ctrl: Docetaxel | Rx: 70  (63-74) ^‡^  Ctrl: 68  (61-73) ^‡^ | 39 (41) | 47 (50) | 8 (9) | 46 (49) | 80 (85) | 57 (61) | NA | 8 (9) | NA | Rx: 73  (32-295) Ctrl: 34  (8-86) |
| **Chemotherapy+Chemotherapy** | | | | | | | | | | | | |
| **MDACC Study**, 2019 | Rx: Cabazitaxel25 + Carboplatin Ctrl: Cabazitaxel25 | Rx: 68  (62–73) ^‡^  Ctrl: 66  (61–69) ^‡^ | 44 (27) | 117 (73) | | NA | 147 (92) | 83 (52) | 57 (36) | NA | NA | Rx: 33.8  (11.4–146.3) Ctrl: 23.7  (8.6–81.2) |
| **Prior ADT+Docetaxel** | | | | | | | | | | | | |
| **ASO+Chemotherapy** | | | | | | | | | | | | |
| **P-06c,**  2011 | Rx1: Custirsen + Docetaxel Rx2: Custirsen  + Mitoxantrone | Rx1: 68  (48–80)  Rx2: 61  (49–81) | NA | NA | NA | NA | 41 (98) | 24 (57) | 10 (24) | NA | NA | Rx1: 154  (5–3570) *  Rx2: 116  (20–2776) * |
| **Aurora kinase inhibitor monotherapy** | | | | | | | | | | | | |
| **Meulenbeld HJ et al**, 2012 | Rx1: Danusertib330 Rx2: Danusertib500 | Rx1: 68.3  (48–80) Rx2: 67.9  (50–80) | 34 (42) | 42 (52) | 5 (6) | 67 (83) | 76 (94) | 44 (54) | 51 (63) | 8 (10) | 4 (5) | Rx1: 158.9  (4.1–5314) *  Rx2: 197  (27–2182) * |
| **Chemotherapy monotherapy** | | | | | | | | | | | | |
| **GETUG-P02,**  2015 | Rx1: Etoposide Rx2: Vinorelbine Rx3: Mitoxantrone | Rx1: 68.9  (55–84) ^§^ Rx2: 69.8  (52–85) ^§^ Rx3: 70.2  (54–80) ^§^ | 55 (60) | | 34 (37) | NA | 91 (99) | 36 (39) | 34 (37) | NA | NA | Rx1: 299  (13.5–7351) *  Rx2: 257  (28–3146) *  Rx3: 275  (0.4–1514) * |
| **CAINTA**, 2023 | Rx: Cabazitaxel ID Ctrl: Cabazitaxel25 | Rx: 69.7  (55-77) Ctrl: 70.8  (53-82) | 34 (47) | 36 (49) | 3 (4) | NA | 69 (95) | 40 (55) | 24 (33) | 17 (23) | 7 (10) | Rx: 95.4  (1–2814) * Ctrl: 85.7  (0–1501) * |
| **Rosenberg JE et al**, 2007 | Rx: Ixabepilone Ctrl: Mitoxantrone | Rx: 66.5 (51–87) Ctrl: 69  (52–84) | 30 (37) | 52 (63) | | 51 (62) | NA | NA | NA | NA | NA | Rx: 141  (4–17995) * Ctrl: 113  (7–1587) * |
| **Chemotherapy+Chemotherapy** | | | | | | | | | | | | |
| **RECARDO**, 2018 | Rx: Docetaxel + Carboplatin Ctrl: Docetaxel | Rx: 70.1  (60-84) ^§^ Ctrl: 69.2  (61-85) ^§^ | 32 (43) | 38 (51) | 3 (4) | 54 (72) | 68 (91) | 39 (52) | NA | 9 (12) | 9 (12) | Rx: 49  (5-1450) *  Ctrl: 75  (5-1113) * |
| **Chemotherapy+Immunotherapy** | | | | | | | | | | | | |
| **Fizazi K et al**, 2012 | Rx: Mitoxantrone + Siltuximab Ctrl: Mitoxantrone | Rx: 67  (49–87) Ctrl: 69  (51–79) | 86 (89) | | 11 (11) | NA | NA | NA | NA | NA | NA | Rx: 125.6  (9.3–4896.5) *  Ctrl: 241.8  (9.1–2496) * |
| **Hakenberg OW et al**, 2019 | Rx: Mitoxantrone  + Olaratumab Ctrl: Mitoxantrone | Rx: 68  (51-81) Ctrl: 69  (40-79) | 52 (43) | 62 (51) | 7 (6) | NA | NA | NA | NA | NA | NA | NA |
| **Immunotherapy monotherapy** | | | | | | | | | | | | |
| **Filaci G et al**, 2021 | Rx1: GX301 vaccine (8D) Rx2: GX301 vaccine (4D) Rx3: GX301 vaccine (2D) | Rx1: 68.7  (9.8) ^†^  Rx2: 70.8  (7.6) ^†^  Rx3: 68.3  (8.6) ^†^ | 68 (69) | 30 (31) | NA | NA | 81 (83) | 16 (16) | 16 (16) | NA | NA | NA |
| **TKI monotherapy** | | | | | | | | | | | | |
| **Droz JP et al**, 2014 | Rx: Nintedanib150 Ctrl: Nintedanib250 | Rx: 69.6  (53–85) Ctrl: 68.4  (55–83) | 30 (37) | 41 (51) | 10 (12) | NA | 80 (99) | 32 (40) | 12 (15) | NA | NA | Rx: 118  (20–1246) * Ctrl: 95.5  (18–2079) * |
| **Prior ADT+Docetaxel+ARPI** | | | | | | | | | | | | |
| **ARPI monotherapy** | | | | | | | | | | | | |
| **SAK-08-16**, 2023 | Rx: Darolutamide Ctrl: Placebo | Rx: 71  (56-81) Ctrl: 72  (55-87) | 89 (99) | 1 (1) | NA | 48 (53) | 79 (88) | 46 (51) | 5 (6) | 2 (2) | 3 (3) | NA |
| **Chemotherapy monotherapy** | | | | | | | | | | | | |
| **ConCab**, 2018 | Rx: Cabazitaxel (W) Ctrl: Cabazitaxel25 | Rx: 69  (57-79) Ctrl: 68.5  (50-80) | 52 (52) | 48 (48) | NA | NA | 54 (54) | 31 (31) | 11 (11) | NA | NA | Rx: 165  (1.1-1689) * Ctrl: 129  (12-3435) * |
| **Chemotherapy+AKTi** | | | | | | | | | | | | |
| **RE-AKT**, 2024 | Rx: Enzalutamide  + Capivasertib Ctrl: Enzalutamide | Rx: 72.3  (67.5-77.9) ^‡^  Ctrl: 71.5  (67.7-76.2) ^‡^ | NA | NA | NA | 37 (37) | 93 (93) | 60 (60) | 31 (31) | 15 (15) | 16 (16) | Rx: 144.2  (60-240.3)  Ctrl: 245  (79.3-591) |
| **Radioligand monotherapy** | | | | | | | | | | | | |
| **TheraP**, 2021 | Rx: 177Lu Ctrl: Cabazitaxel20 | Rx: 72.1  (66.9-76.7) ^‡^  Ctrl: 71.8  (66.7-77.3) ^‡^ | 86 (43) | 105 (53) | 8 (4) | 103 (52) | 180 (90) | 99 (50) | 20 (10) | NA | NA | Rx: 93.5  (44 to 219) Ctrl: 110  (64.2 to 245) |
| **Heterogeneous Prior Therapy** | | | | | | | | | | | | |
| **ARPI+Radioligand** | | | | | | | | | | | | |
| **ENZA-p**, 2024 | Rx: Enzalutamide + 177Lu Ctrl: Enzalutamide | Rx: 71  (66–76) ^‡^  Ctrl: 71  (63–76) ^‡^ | NA | NA | NA | NA | NA | NA | 17 (10) | NA | NA | Rx: 39  (13–75) Ctrl: 33  (14–85) |
| **Chemotherapy monotherapy** | | | | | | | | | | | | |
| **TAXYNERGY**, 2017 | Rx: Docetaxel Ctrl: Cabazitaxel25 | Rx: 71  (53–84) Ctrl: 70 (53–81) | 18 (29) | 42 (67) | 3 (5) | 34 (54) | 56 (89) | 32 (51) | 22 (35) | NA | NA | Rx: 105.7  (2.4–1558.0) * Ctrl: 78.0  (4.4–977.9) * |
| **Chemotherapy+Immunotherapy** | | | | | | | | | | | | |
| **IND 209**, 2018 | Rx: Docetaxel  + Reolysin Ctrl: Docetaxel | Rx: 69.1  (50.3–83.7) Ctrl: 68.6  (49.7–86.6) | 26 (31) | 56 (66) | 3 (3) | 47 (55) | NA | NA | 30 (35) | NA | NA | Rx: 189.7  (266.8) ^†^  Ctrl: 257.1  (473.1) ^†^ |
| **Kongsted P et al,**  2017 | Rx: Docetaxel  + DCVAC Ctrl: Docetaxel | Rx: 70  (60-84) Ctrl: 71  (63-80) | 39 (97) | | 1 (3) | 28 (70) | 35 (88) | 5 (13) | 3 (8) | NA | NA | Rx: 84  (11–428) * Ctrl: 141  (21–997) * |
| **PARPi+ARPi** | | | | | | | | | | | | |
| **BRCAAway**, 2024 | Rx1: Abiraterone Rx2: Olaparib Rx3: Abiraterone  + Olaparib | Rx1: 63  (60–69) ^‡^  Rx2: 68  (66–72) ^‡^  Rx3: 69  (62–74) ^‡^ | 41 (67) | 20 (33) | NA | NA | 45 (74) | 31 (51) | 12 (20) | 4 (7) | 9 (15) | Rx1: 14  (3.7–133)  Rx2: 14  (7.7–33) Rx3: 15  (6.3–39) |
| **TKI monotherapy** | | | | | | | | | | | | |
| **Monk P et al**, 2018 | Rx: Tivantinib Ctrl: Placebo | Rx: 67  (43–84) Ctrl: 66.5  (48–85) | 65 (83) | 13 (17) | NA | 41 (53) | 50 (64) | 17 (22) | NA | NA | 5 (6) | Rx: 13.6  (2.3-868) * Ctrl: 26.7  (2.2-579) * |
| **Smith DC et al**, 2013 | Rx: Cabozantinib Ctrl: Placebo | NA | NA | NA | NA | NA | NA | NA | NA | NA | NA | NA |
| **Multiple subgroups (ADT/ADT+ARPI/ADT+Docetaxel/ADT+ARPI+Docetaxel/Heterogeneous prior therapy)** | | | | | | | | | | | | |
| **Chemotherapy monotherapy** | | | | | | | | | | | | |
| **Annala M et al**, 2021 | Rx: Cabazitaxel25 Ctrl: Enzalutamide /Abiraterone | Rx: 68  (59-73) ^‡^  Ctrl: 67.5  (60.3-71) ^‡^ | 89 (94) | | NA | NA | 80 (84) | NA | 40 (42) | 17 (18) | 23 (24) | Rx: 18.7  (9.4-38.5) Ctrl: 39.4  (20.2-107.5) |
| **Chemotherapy+AKTi** | | | | | | | | | | | | |
| **ProCAID**, 2021 | Rx: Docetaxel + Capivasertib Ctrl: Docetaxel | Rx: 69  (64-67) ^‡^  Ctrl: 70  (66-75) ^‡^ | 90 (60) | 60 (40) | NA | 91 (61) | 124 (83) | 69 (46) | 31 (21) | 8 (5) | 14 (9) | Rx: 37.5  (13.6-93.9) Ctrl: 62  (28-151) |

Abbreviations: ADT: androgen deprivation therapy; ARPI: androgen-receptor pathway inhibitor; cSipuleucel-T: concurrent sipuleucel-T; sSipuleucel-T: sequential sipuleucel-T; TKI: tyrosine kinase inhibitor; ASO: antisense oligonucleotide; Bcl-2: B-cell lymphoma 2 protein; PVAC: PROSTVAC (viral vector-based immunotherapy); VitD: vitamin d; VEGFi: vascular endothelial growth factor inhibitor; ERA: endothelin receptor antagonist; IMiD: immunomodulatory drug; SOC: standard of care; GMCSF: granulocyte-macrophage colony-stimulating factor; AA: antiandrogen; BAT: bipolar androgen therapy; PARPi: poly(ADP-ribose) polymerase inhibitor; 177Lu: 177Lu-PSMA-617; AKTi: protein kinase B inhibitor; Vin/Estra: vinblastine/estramustine; keto: ketoconazole

The color ‘green’ represents a positive trial, ‘red’ represents a negative trial.

* Median (range)

^†^ Mean (SD)

^‡^ Median (IQR)

^§^ Mean (range)

^||^ ECOG PS 2-3

Itraconazole600: Itraconazole 600mg/day PO

Itraconazole200: Itraconazole 200mg/day PO

L-Doxorubicin25: Liposomal Doxorubicin (dissolved in 250 mL 5% glucose) 25 mg/m^2^ IV every 2 weeks for 12 consecutive cycles

L-Doxorubicin50: Liposomal Doxorubicin (dissolved in 250 mL 5% glucose) 50 mg/m^2^ IV every 4 weeks for 6 consecutive cycles

Docetaxel30 (W): Docetaxel 30 mg/m^2^ IV every week for 5 weeks, followed by 1 week rest (6-week cycle)

Docetaxel25 (W): Docetaxel 25 mg/m^2^ IV every week for 2 cycles (8-week cycle)

Docetaxel35 (W): Docetaxel 35 mg/m^2^ IV on days 2 and 9 every 3 weeks

Docetaxel70: Docetaxel 70 mg/m^2^ on day 2 every 3 weeks

Estramustine840: Estramustine 840 mg/day on days 1-5

Estramustine420: Estramustine 420 mg/day on days 1-3

Docetaxel35 (D2,D9 Q3W): Docetaxel 35 mg/m^2^ on day 2 and 9 every 3wks

Docetaxel30 (W) ^f^ : Docetaxel 30 mg/m^2^ IV every week for 3 weeks, followed by 1 week rest (4-week cycle)

Docetaxel35 (W) ^g^ : Docetaxel 35 mg/m^2^ IV on days 1, 8, and 15 (4-week cycle)

Docetaxel36 (W): Docetaxel 36 mg/m^2^ IV every week for 3 weeks (4-week cycle)

Zibotentan15: Zibotentan 15 mg PO OD

Zibotentan10: Zibotentan 10 mg PO OD

Atrasentan2.5: Atrasentan 2.5 mg PO OD

Atrasentan10: Atrasentan 10 mg PO OD

Abituzumab750: Abituzumab 750 mg every 3 weeks

Abituzumab1500: Abituzumab 1500 mg every 3 weeks

ThalidomideHD: Thalidomide initial dose of 200 mg/day with increments of 200 mg/day every 2 weeks to a maximum dose of 1200 mg

Cilengitide500: Cilengitide 500 mg IV twice weekly (six-week cycle)

Cilengitide2000: Cilengitide 2000 mg IV twice weekly (six-week cycle)

Cabazitaxel25: Cabazitaxel 25 mg/m^2^ IV every 3 weeks

Danusertib330: Danusertib 330 mg/m^2^ IV on days 1,8 and 15 every 4 weeks

Danusertib500: Danusertib 500 mg/m^2^ IV on days 1 and 15 every 4 weeks

Cabazitaxel ID: Cabazitaxel at an initial dose of 25 mg/m^2^ followed by dose adaptations in cycle 2 and beyond according to a prespecified dosing algorithm considering previous-cycle hematologic toxicity and cabazitaxel AUC (with a target AUC of 0.8–1.2 mg*hour/L)

GX301 vaccine (8D): GX301, 8 doses on days 1, 3, 5, 7, 14, 21, 35 and 63

GX301 vaccine (4D): GX301, 4 doses on days 1, 14, 35 and 63

GX301 vaccine (2D): GX301, 2 doses on days 1 and 63

Nintedanib150: Nintedanib 150 mg PO BID for 6 months

Nintedanib250: Nintedanib 250 mg PO BID for 6 months

Cabazitaxel (W): Cabazitaxel 25 mg/m^2^ IV weekly for 5 weeks (6-week cycle)

Cabazitaxel20: Cabazitaxel 20 mg/m^2^ IV every 3 weeks

Remaining drugs were administered at standard doses.

### **Supplement Table 8:** Prior therapy administered to patients in included phase II trials

| **Trial** | **Arm** | **Prior Therapy in Any Setting** | | | | **Prior Therapy in mHSPC Setting** | | | | **Prior Therapy in mCRPC Setting** | | | |
| --- | --- | --- | --- | --- | --- | --- | --- | --- | --- | --- | --- | --- | --- |
|  |  | **Docetaxel** | **ARPI** | **Abiraterone** | **Enzalutamide** | **Docetaxel** | **ARPI** | **Abiraterone** | **Enzalutamide** | **Docetaxel** | **ARPI** | **Abiraterone** | **Enzalutamide** |
| **Antiandrogen+Chemotherapy** | | | | | | | | | | | | | |
| **Takahashi**  **M et al**, 2013 | Rx: Antiandrogen + Tegafur-Uracil Ctrl: Antiandrogen | NA | NA | NA | NA | NA | NA | NA | NA | NA | NA | NA | NA |
| **Antifungal monotherapy** | | | | | | | | | | | | | |
| **Antonarakis ES et al**, 2013 | Rx: Itraconazole600 Ctrl: Itraconazole200 | NA | NA | NA | NA | NA | NA | NA | NA | NA | NA | NA | NA |
| **Antifungal+Bisphosphonate** | | | | | | | | | | | | | |
| **Figg WD et al,**  2005 | Rx: Ketoconazole + Alendronate Ctrl: Ketoconazole | NA | NA | NA | NA | NA | NA | NA | NA | NA | NA | NA | NA |
| **Antifungal+Chemotherapy** | | | | | | | | | | | | | |
| **Millikan R**  **et al**, 2001 | Rx1: Ketoconazole + Doxorubicin Rx2: Ketoconazole | NA | NA | NA | NA | NA | NA | NA | NA | NA | NA | NA | NA |
| **ARPI monotherapy** | | | | | | | | | | | | | |
| **TERRAIN**, 2016 | Rx: Enzalutamide Ctrl: Bicalutamide | NA | NA | NA | NA | NA | NA | NA | NA | NA | NA | NA | NA |
| **SAK-08-16**, 2023 | Rx: Darolutamide Ctrl: Placebo | 82  (91) | 90  (100) | 62  (69) | 36  (40) | NA | 17 (19) | NA | NA | 82  (91) | 90  (100) | 62  (69) | 36  (40) |
| **ARPI+ARPI** | | | | | | | | | | | | | |
| **Khalaf DJ**  **et al**, 2019 | Rx1: Abiraterone followed by Enzalutamide Rx2: Enzalutamide followed by Abiraterone | 11  (5) | NA | NA | NA | 11  (5) | NA | NA | NA | NA | NA | NA | NA |
| **ARPI+Immunotherapy** | | | | | | | | | | | | | |
| **STAMP**, 2015 | Rx1: Abiraterone + cSipuleucel-T Rx2: Abiraterone + sSipuleucel-T | NA | NA | NA | NA | NA | NA | NA | NA | NA | NA | NA | NA |
| **STRIDE**, 2023 | Rx1: Enzalutamide + cSipuleucel-T  Rx2: Enzalutamide + sSipuleucel-T | NA | NA | NA | NA | NA | NA | NA | NA | NA | NA | NA | NA |
| **ARPI+Radioligand** | | | | | | | | | | | | | |
| **ENZA-p**, 2024 | Rx: Enzalutamide + 177Lu Ctrl: Enzalutamide | 89  (55) | 21  (13) | 21  (13) | NA | 89  (55) | 21 (13) | 21  (13) | NA | NA | NA | NA | NA |
| **ARPI+TKI** | | | | | | | | | | | | | |
| **Dorff TB**  **et al**, 2019 | Rx: Abiraterone  + Dasatinib Ctrl: Abiraterone | NA | NA | NA | NA | NA | NA | NA | NA | NA | NA | NA | NA |
| **Spetsieris**  **N et al**, 2021 | Rx: Abiraterone + Sunitinib Ctrl: Abiraterone + Dasatinib | 33  (25) | 132  (100) | 132  (100) | NA | NA | NA | NA | NA | NA | 132  (100) | 132  (100) | NA |
| **ASO monotherapy** | | | | | | | | | | | | | |
| **Yu EY et al**, 2018 | Rx: Apatorsen Ctrl: Prednisone | NA | NA | NA | NA | NA | NA | NA | NA | NA | NA | NA | NA |
| **Tolcher AW et al,**  2002 | Rx: ISIS 3521 Ctrl: ISIS 5132 | NA | NA | NA | NA | NA | NA | NA | NA | NA | NA | NA | NA |
| **ASO+Chemotherapy** | | | | | | | | | | | | | |
| **P-06c,**  2011 | Rx1: Custirsen + Docetaxel Rx2: Custirsen  + Mitoxantrone | 42  (100) | NA | NA | NA | NA | NA | NA | NA | 42  (100) | NA | NA | NA |
| **Aurora kinase inhibitor monotherapy** | | | | | | | | | | | | | |
| **Meulenbeld HJ et al**, 2012 | Rx1: Danusertib330 Rx2: Danusertib500 | 81  (100) | NA | NA | NA | NA | NA | NA | NA | 81  (100) | NA | NA | NA |
| **Bipolar Androgen Therapy** | | | | | | | | | | | | | |
| **TRANSFORMER**, 2021 | Rx: BAT Ctrl: Enzalutamide | 24  (12) | 195  (100) | 195  (100) | NA | 24  (12) | NA | NA | NA | NA | 195  (100) | 195  (100) | NA |
| **Chemotherapy monotherapy** | | | | | | | | | | | | | |
| **Heidenreich A et al**, 2004 | Rx1: L-Doxorubicin25  Rx2: L-Doxorubicin50 | NA | NA | NA | NA | NA | NA | NA | NA | NA | NA | NA | NA |
| **Annala M et al**, 2021 | Rx: Cabazitaxel25 Ctrl: Enzalutamide /Abiraterone | NA | NA | NA | NA | 25  (26) | NA | NA | NA | 26  (27) | NA | NA | NA |
| **TAXYNERGY**, 2017 | Rx: Docetaxel Ctrl: Cabazitaxel25 | NA | 28  (44) | NA | NA | NA | NA | NA | NA | NA | 28  (44) | NA | NA |
| **TIPC**, 2007 | Rx: Docetaxel30 (W) Ctrl: Prednisolone | NA | NA | NA | NA | NA | NA | NA | NA | NA | NA | NA | NA |
| **GETUG-P02,**  2015 | Rx1: Etoposide Rx2: Vinorelbine Rx3: Mitoxantrone | 92  (100) | NA | NA | NA | NA | NA | NA | NA | 92  (100) | NA | NA | NA |
| **Krainer M et al**, 2007 | Rx: Docetaxel25 (W) Ctrl: Vinorelbine | NA | NA | NA | NA | NA | NA | NA | NA | NA | NA | NA | NA |
| **ConCab**, 2018 | Rx: Cabazitaxel (W) Ctrl: Cabazitaxel25 | 100  (100) | 84  (84) | 77  (77) | 38  (38) | NA | NA | NA | NA | 100  (100) | 84  (84) | 77  (77) | 38  (38) |
| **CAINTA**, 2023 | Rx: Cabazitaxel ID Ctrl: Cabazitaxel25 | 73  (100) | 49  (67) | NA | NA | NA | NA | NA | NA | 73  (100) | 49  (67) | NA | NA |
| **Rosenberg JE et al**, 2007 | Rx: Ixabepilone Ctrl: Mitoxantrone | 82  (100) | NA | NA | NA | NA | NA | NA | NA | 82  (100) | NA | NA | NA |
| **Chemotherapy+AKTi** | | | | | | | | | | | | | |
| **ProCAID**, 2021 | Rx: Docetaxel + Capivasertib Ctrl: Docetaxel | Given but number not specified | 101  (67) | 36  (24) | 53  (35) | Given but number not specified | NA | NA | NA | NA | 101  (67) | 36  (24) | 53  (35) |
| **RE-AKT**, 2024 | Rx: Enzalutamide  + Capivasertib Ctrl: Enzalutamide | 100 (100) | 100 (100) | 100 (100) | NA | NA | NA | NA | NA | NA | NA | NA | NA |
| **Chemotherapy+Antifungal** | | | | | | | | | | | | | |
| **Millikan R et al,**  2003 | Rx: Vin/Estra + Keto/Doxorubicin Ctrl: Paclitaxel + Estramustine + Etoposide | NA | NA | NA | NA | NA | NA | NA | NA | NA | NA | NA | NA |
| **Chemotherapy+ARPI** | | | | | | | | | | | | | |
| **ABIDO SOGUG**, 2022 | Rx: Docetaxel + Abiraterone Ctrl: Docetaxel | NA | 94  (100) | 94  (100) | NA | NA | NA | NA | NA | NA | 94  (100) | 94  (100) | NA |
| **CHEIRON**, 2021 | Rx: Docetaxel + Enzalutamide Ctrl: Docetaxel | NA | NA | NA | NA | NA | NA | NA | NA | NA | NA | NA | NA |
| **Chemotherapy+ASO** | | | | | | | | | | | | | |
| **Chi KN et al,**  2010 | Rx: Docetaxel + Custirsen Ctrl: Docetaxel | NA | NA | NA | NA | NA | NA | NA | NA | NA | NA | NA | NA |
| **Wiechno P et al**, 2014 | Rx: Docetaxel + LY2181308 Ctrl: Docetaxel | NA | NA | NA | NA | NA | NA | NA | NA | NA | NA | NA | NA |
| **Chemotherapy+Bcl-2 inhibitor** | | | | | | | | | | | | | |
| **EORTC**, 2009 | Rx: Docetaxel  + Oblimersen Ctrl: Docetaxel | NA | NA | NA | NA | NA | NA | NA | NA | NA | NA | NA | NA |
| **Sonpavde G et al**, 2011 | Rx: Docetaxel + AT-101 Ctrl: Docetaxel | NA | NA | NA | NA | NA | NA | NA | NA | NA | NA | NA | NA |
| **Chemotherapy+Chemotherapy** | | | | | | | | | | | | | |
| **Machiels JP et al**, 2008 | Rx: Docetaxel35 (W) + Estramustine Ctrl: Docetaxel35 (W) | NA | NA | NA | NA | NA | NA | NA | NA | NA | NA | NA | NA |
| **Berry WR et al**, 2004 | Rx: Paclitaxel + Estramustine Ctrl: Paclitaxel | NA | NA | NA | NA | NA | NA | NA | NA | NA | NA | NA | NA |
| **Albrecht W et al**, 2004 | Rx: Estramustine + Vinblastine Ctrl: Estramustine | NA | NA | NA | NA | NA | NA | NA | NA | NA | NA | NA | NA |
| **Cabrespine A et al**, 2006 | Rx: Paclitaxel + Carboplatin Ctrl: Mitoxantrone | NA | NA | NA | NA | NA | NA | NA | NA | NA | NA | NA | NA |
| **Nelius T et al**, 2006 | Rx1: Docetaxel70 + Estramustine840 Rx2: Docetaxel70 + Estramustine420 | NA | NA | NA | NA | NA | NA | NA | NA | NA | NA | NA | NA |
| **Oudard S et al**, 2005 | Rx1: Docetaxel70 + Estramustine Rx2: Docetaxel35 (D2,D9 Q3W) + Estramustine Ctrl: Mitoxantrone | NA | NA | NA | NA | NA | NA | NA | NA | NA | NA | NA | NA |
| **MDACC Study**, 2019 | Rx: Cabazitaxel25 + Carboplatin Ctrl: Cabazitaxel25 | 46  (29) | 147  (92) | 98  (61) | 49  (31) | NA | NA | NA | NA | 46  (29) | 147  (92) | 98  (61) | 49  (31) |
| **RECARDO**, 2018 | Rx: Docetaxel + Carboplatin Ctrl: Docetaxel | 75  (100) | NA | NA | NA | NA | NA | NA | NA | 75  (100) | NA | NA | NA |
| **Droz JP et al**, 2003 | Rx1: Oxaliplatin + 5-Fluorouracil Rx2: Oxaliplatin | NA | NA | NA | NA | NA | NA | NA | NA | NA | NA | NA | NA |
| **Galsky MD et al**, 2005 | Rx1: Ixabepilone + Estramustine Rx2: Ixabepilone | NA | NA | NA | NA | NA | NA | NA | NA | NA | NA | NA | NA |
| **Eymard JC et al**, 2007 | Rx: Docetaxel70 + Estramustine Ctrl: Docetaxel | NA | NA | NA | NA | NA | NA | NA | NA | NA | NA | NA | NA |
| **Chemotherapy+Curcuminoid** | | | | | | | | | | | | | |
| **Jahanmohan JP et al**, 2021 | Rx: Docetaxel + Curcumin Ctrl: Docetaxel | NA | NA | NA | NA | NA | NA | NA | NA | NA | NA | NA | NA |
| **Chemotherapy+Immunotherapy** | | | | | | | | | | | | | |
| **Dahut WL et al**, 2004 | Rx: Docetaxel30 (W) ^a^ + Thalidomide Ctrl: Docetaxel30 (W) ^a^ | NA | NA | NA | NA | NA | NA | NA | NA | NA | NA | NA | NA |
| **IND 209**, 2018 | Rx: Docetaxel  + Reolysin Ctrl: Docetaxel | NA | 46  (54) | 28  (33) | 18  (21) | NA | NA | NA | NA | NA | 46  (54) | 28  (33) | 18  (21) |
| **Fizazi K et al**, 2012 | Rx: Mitoxantrone + Siltuximab Ctrl: Mitoxantrone | 97  (100) | NA | NA | NA | NA | NA | NA | NA | 97  (100) | NA | NA | NA |
| **ECOG 3899**, 2010 | Rx1: 13-cis Retinoic acid + Interferon-alpha2b + Paclitaxel Rx2: Vinorelbine + Mitoxantrone + Estramustine | NA | NA | NA | NA | NA | NA | NA | NA | NA | NA | NA | NA |
| **Hakenberg OW et al**, 2019 | Rx: Mitoxantrone  + Olaratumab Ctrl: Mitoxantrone | 121  (100) | NA | NA | NA | NA | NA | NA | NA | 121  (100) | NA | NA | NA |
| **E1809**, 2015 | Rx: Docetaxel + PVAC-VF Ctrl: Docetaxel | NA | NA | NA | NA | NA | NA | NA | NA | NA | NA | NA | NA |
| **Kongsted P et al,**  2017 | Rx: Docetaxel  + DCVAC Ctrl: Docetaxel | NA | 11  (28) | NA | NA | NA | NA | NA | NA | NA | 11  (28) | NA | NA |
| **de Bono et al**, 2014 | Rx: Docetaxel + Figitumumab Ctrl: Docetaxel | NA | NA | NA | NA | NA | NA | NA | NA | NA | NA | NA | NA |
| **Heidenreich A et al,**  2013 | Rx: Docetaxel  + Intetumumab Ctrl: Docetaxel | NA | NA | NA | NA | NA | NA | NA | NA | NA | NA | NA | NA |
| **Chemotherapy+Radiopharmaceutical** | | | | | | | | | | | | | |
| **Taxium II**, 2017 | Rx: Docetaxel  + Rhenium-188-HEDP Ctrl: Docetaxel | NA | NA | NA | NA | NA | NA | NA | NA | NA | NA | NA | NA |
| **Chemotherapy+TKI** | | | | | | | | | | | | | |
| **Horti J et al**, 2009 | Rx: Docetaxel + Vandetanib Ctrl: Docetaxel | NA | NA | NA | NA | NA | NA | NA | NA | NA | NA | NA | NA |
| **Chemotherapy+Vascular disrupting agent** | | | | | | | | | | | | | |
| **Pili R et al,**  2010 | Rx: Docetaxel + Vadimezan Ctrl: Docetaxel | NA | NA | NA | NA | NA | NA | NA | NA | NA | NA | NA | NA |
| **Chemotherapy+VitD-Analog** | | | | | | | | | | | | | |
| **Attia S et al**, 2008 | Rx: Docetaxel35 (W) ^b^ + Doxercalciferol Ctrl: Docetaxel35 (W) ^b^ | NA | NA | NA | NA | NA | NA | NA | NA | NA | NA | NA | NA |
| **ASCENT**, 2007 | Rx: Docetaxel36 (W) + Calcitriol Ctrl: Docetaxel36 (W) | NA | NA | NA | NA | NA | NA | NA | NA | NA | NA | NA | NA |
| **Corticosteroid monotherapy** | | | | | | | | | | | | | |
| **Venkitaraman R et al**, 2015 | Rx: Dexamethasone Ctrl: Prednisolone | NA | NA | NA | NA | NA | NA | NA | NA | NA | NA | NA | NA |
| **Corticosteroid+Immunotherapy** | | | | | | | | | | | | | |
| **Yoshimura K et al**, 2016 | Rx: Dexamethasone + PPV Ctrl: Dexamethasone | NA | NA | NA | NA | NA | NA | NA | NA | NA | NA | NA | NA |
| **Corticosteroid+VEGFi** | | | | | | | | | | | | | |
| **Stadler WM et al**, 2004 | Rx: Dexamethasone + SU5416 Ctrl: Dexamethasone | NA | NA | NA | NA | NA | NA | NA | NA | NA | NA | NA | NA |
| **ERA monotherapy** | | | | | | | | | | | | | |
| **James ND et al**, 2009 | Rx1: Zibotentan15  Rx2: Zibotentan10 Ctrl: Placebo | NA | NA | NA | NA | NA | NA | NA | NA | NA | NA | NA | NA |
| **Carducci MA et al**, 2003 | Rx1: Atrasentan2.5 Rx2: Atrasentan10 Ctrl: Placebo | NA | NA | NA | NA | NA | NA | NA | NA | NA | NA | NA | NA |
| **IMiD monotherapy** | | | | | | | | | | | | | |
| **Pili R et al**, 2011 | Rx: Tasquinimod Ctrl: Placebo | NA | NA | NA | NA | NA | NA | NA | NA | NA | NA | NA | NA |
| **Immunotherapy monotherapy** | | | | | | | | | | | | | |
| **PERSEUS**, 2016 | Rx1: Abituzumab750 + SOC Rx2: Abituzumab1500 + SOC Ctrl: SOC | NA | NA | NA | NA | NA | NA | NA | NA | NA | NA | NA | NA |
| **Filaci G et al**, 2021 | Rx1: GX301 vaccine (8D) Rx2: GX301 vaccine (4D) Rx3: GX301 vaccine (2D) | 98  (100) | 9  (9) | NA | NA | NA | NA | NA | NA | 98  (100) | 9  (9) | NA | NA |
| **Figg WD et al**, 2001 | Rx: ThalidomideHD Ctrl: Thalidomide | NA | NA | NA | NA | NA | NA | NA | NA | NA | NA | NA | NA |
| **Immunotherapy+Chemotherapy** | | | | | | | | | | | | | |
| **Arlen PM et al,** 2006 | Rx: Vaccine + GMCSF + Docetaxel Ctrl: Vaccine + GMCSF | NA | NA | NA | NA | NA | NA | NA | NA | NA | NA | NA | NA |
| **Immunotherapy+GMCSF** | | | | | | | | | | | | | |
| **TBC-PRO-002**, 2010 | Rx: PVAC+GMCSF Ctrl: Placebo | NA | NA | NA | NA | NA | NA | NA | NA | NA | NA | NA | NA |
| **Integrin inhibitor monotherapy** | | | | | | | | | | | | | |
| **Bradley DA et al,**  2011 | Rx: Cilengitide500 Ctrl: Cilengitide2000 | NA | NA | NA | NA | NA | NA | NA | NA | NA | NA | NA | NA |
| **Matrix Metalloproteinase Inhibitor monotherapy** | | | | | | | | | | | | | |
| **Lara Jr PN et al**, 2006 | Rx: BMS-275291  [1200mg] Ctrl: BMS-275291 [2400mg] | NA | NA | NA | NA | NA | NA | NA | NA | NA | NA | NA | NA |
| **PARPi+ARPi** | | | | | | | | | | | | | |
| **BRCAAway**, 2024 | Rx1: Abiraterone Rx2: Olaparib Rx3: Abiraterone  + Olaparib | 16 (26) | 2  (3) | NA | NA | 16  (26) | NA | NA | NA | NA | NA | NA | NA |
| **Progestin monotherapy** | | | | | | | | | | | | | |
| **Patel SR et al**, 1990 | Rx: Megestrol Acetate Ctrl: Dexamethasone | NA | NA | NA | NA | NA | NA | NA | NA | NA | NA | NA | NA |
| **Radioligand monotherapy** | | | | | | | | | | | | | |
| **TheraP**, 2021 | Rx: 177Lu Ctrl: Cabazitaxel20 | 200  (100) | 182  (91) | 75  (38) | 137  (69) | NA | NA | NA | NA | 200  (100) | 182  (91) | 75  (38) | 137  (69) |
| **Somatostatin+Corticosteroid** | | | | | | | | | | | | | |
| **Dimopoulos MA et al**, 2004 | Rx: Lanreotide + Dexamethasone Ctrl: Estramustine + Etoposide | NA | NA | NA | NA | NA | NA | NA | NA | NA | NA | NA | NA |
| **TKI monotherapy** | | | | | | | | | | | | | |
| **Droz JP et al**, 2014 | Rx: Nintedanib150 Ctrl: Nintedanib250 | 81  (100) | NA | NA | NA | NA | NA | NA | NA | 81  (100) | NA | NA | NA |
| **Boccardo F et al**, 2008 | Rx: Gefitinib Ctrl: Placebo | NA | NA | NA | NA | NA | NA | NA | NA | NA | NA | NA | NA |
| **Monk P et al**, 2018 | Rx: Tivantinib Ctrl: Placebo | NA | 23  (29) | 23  (29) | NA | NA | NA | NA | NA | NA | 23  (29) | 23  (29) | NA |
| **Smith DC et al**, 2013 | Rx: Cabozantinib Ctrl: Placebo | Given but number not specified | Given but number not specified | NA | NA | NA | NA | NA | NA | Given but number not specified | Given but number not specified | NA | NA |
| **TKI+1st gen AA** | | | | | | | | | | | | | |
| **Azad AA et al**, 2014 | Rx: Vandetanib + Bicalutamide Ctrl: Bicalutamide | NA | NA | NA | NA | NA | NA | NA | NA | NA | NA | NA | NA |
| **Sridhar SS et al**, 2015 | Rx1: Pazopanib + Bicalutamide Rx2: Pazopanib | NA | NA | NA | NA | NA | NA | NA | NA | NA | NA | NA | NA |

Abbreviations: mHSPC: metastatic hormone sensitive prostate cancer; mCRPC: metastatic castration resistant prostate cancer; ADT: androgen deprivation therapy; ARPI: androgen-receptor pathway inhibitor; cSipuleucel-T: concurrent sipuleucel-T; sSipuleucel-T: sequential sipuleucel-T; TKI: tyrosine kinase inhibitor; ASO: antisense oligonucleotide; Bcl-2: B-cell lymphoma 2 protein; PVAC: PROSTVAC (viral vector-based immunotherapy); VitD: vitamin d; VEGFi: vascular endothelial growth factor inhibitor; ERA: endothelin receptor antagonist; IMiD: immunomodulatory drug; SOC: standard of care; GMCSF: granulocyte-macrophage colony-stimulating factor; AA: antiandrogen; BAT: bipolar androgen therapy; PARPi: poly(ADP-ribose) polymerase inhibitor; 177Lu: 177Lu-PSMA-617;; AKTi: protein kinase B inhibitor; Vin/Estra: vinblastine/estramustine; keto: ketoconazole

The color ‘green’ represents a positive trial, ‘red’ represents a negative trial.

Itraconazole600: Itraconazole 600mg/day PO

Itraconazole200: Itraconazole 200mg/day PO

L-Doxorubicin25: Liposomal Doxorubicin (dissolved in 250 mL 5% glucose) 25 mg/m^2^ IV every 2 weeks for 12 consecutive cycles

L-Doxorubicin50: Liposomal Doxorubicin (dissolved in 250 mL 5% glucose) 50 mg/m^2^ IV every 4 weeks for 6 consecutive cycles

Docetaxel30 (W): Docetaxel 30 mg/m^2^ IV every week for 5 weeks, followed by 1 week rest (6-week cycle)

Docetaxel25 (W): Docetaxel 25 mg/m^2^ IV every week for 2 cycles (8-week cycle)

Docetaxel35 (W): Docetaxel 35 mg/m^2^ IV on days 2 and 9 every 3 weeks

Docetaxel70: Docetaxel 70 mg/m^2^ on day 2 every 3 weeks

Estramustine840: Estramustine 840 mg/day on days 1-5

Estramustine420: Estramustine 420 mg/day on days 1-3

Docetaxel35 (D2,D9 Q3W): Docetaxel 35 mg/m^2^ on day 2 and 9 every 3wks

Docetaxel30 (W) ^a^ : Docetaxel 30 mg/m^2^ IV every week for 3 weeks, followed by 1 week rest (4-week cycle)

Docetaxel35 (W) ^b^ : Docetaxel 35 mg/m^2^ IV on days 1, 8, and 15 (4-week cycle)

Docetaxel36 (W): Docetaxel 36 mg/m^2^ IV every week for 3 weeks (4-week cycle)

Zibotentan15: Zibotentan 15 mg PO OD

Zibotentan10: Zibotentan 10 mg PO OD

Atrasentan2.5: Atrasentan 2.5 mg PO OD

Atrasentan10: Atrasentan 10 mg PO OD

Abituzumab750: Abituzumab 750 mg every 3 weeks

Abituzumab1500: Abituzumab 1500 mg every 3 weeks

ThalidomideHD: Thalidomide initial dose of 200 mg/day with increments of 200 mg/day every 2 weeks to a maximum dose of 1200 mg

Cilengitide500: Cilengitide 500 mg IV twice weekly (six-week cycle)

Cilengitide2000: Cilengitide 2000 mg IV twice weekly (six-week cycle)

Cabazitaxel25: Cabazitaxel 25 mg/m^2^ IV every 3 weeks

Danusertib330: Danusertib 330 mg/m^2^ IV on days 1,8 and 15 every 4 weeks

Danusertib500: Danusertib 500 mg/m^2^ IV on days 1 and 15 every 4 weeks

Cabazitaxel ID: Cabazitaxel at an initial dose of 25 mg/m^2^ followed by dose adaptations in cycle 2 and beyond according to a prespecified dosing algorithm considering previous-cycle hematologic toxicity and cabazitaxel AUC (with a target AUC of 0.8–1.2 mg*hour/L)

GX301 vaccine (8D): GX301, 8 doses on days 1, 3, 5, 7, 14, 21, 35 and 63

GX301 vaccine (4D): GX301, 4 doses on days 1, 14, 35 and 63

GX301 vaccine (2D): GX301, 2 doses on days 1 and 63

Nintedanib150: Nintedanib 150 mg PO BID for 6 months

Nintedanib250: Nintedanib 250 mg PO BID for 6 months

Cabazitaxel (W): Cabazitaxel 25 mg/m^2^ IV weekly for 5 weeks (6-week cycle)

Cabazitaxel20: Cabazitaxel 20 mg/m^2^ IV every 3 weeks

Remaining drugs were administered at standard doses.

### **Supplement Table 9:** Matrix for reported overall survival and progression free survival according to prior therapy in included phase II trials

| **Trial** | **Prior ADT** | | **Prior  ADT+ARPI** | | **Prior  ADT+Docetaxel** | | **Prior  ADT+Docetaxel+ARPI** | | **Prior True Triplet Therapy** | | **Heterogeneous Prior Therapy** | |
| --- | --- | --- | --- | --- | --- | --- | --- | --- | --- | --- | --- | --- |
|  | **OS** | **PFS** | **OS** | **PFS** | **OS** | **PFS** | **OS** | **PFS** | **OS** | **PFS** | **OS** | **PFS** |
| **Antiandrogen+Chemotherapy** | | | | | | | | | | | | |
| **Takahashi M et al**, 2013 |  |  |  |  |  |  |  |  |  |  |  |  |
| **Antifungal monotherapy** | | | | | | | | | | | | |
| **Antonarakis ES et al**, 2013 |  | Overall |  |  |  |  |  |  |  |  |  |  |
| **Antifungal+Bisphosphonate** | | | | | | | | | | | | |
| **Figg WD et al,** 2005 | Overall | Overall |  |  |  |  |  |  |  |  |  |  |
| **Antifungal+Chemotherapy** | | | | | | | | | | | | |
| **Millikan R et al**, 2001 | Overall |  |  |  |  |  |  |  |  |  |  |  |
| **ARPI monotherapy** | | | | | | | | | | | | |
| **TERRAIN**, 2016 |  | Overall |  |  |  |  |  |  |  |  |  |  |
| **SAK-08-16**, 2023 |  |  |  |  |  |  | Overall | Overall |  |  |  |  |
| **ARPI+ARPI** | | | | | | | | | | | | |
| **Khalaf DJ et al**, 2019 | Overall | Overall |  |  |  |  |  |  |  |  |  |  |
| **ARPI+Immunotherapy** | | | | | | | | | | | | |
| **STAMP**, 2015 | Overall |  |  |  |  |  |  |  |  |  |  |  |
| **STRIDE**, 2023 | Overall |  |  |  |  |  |  |  |  |  |  |  |
| **ARPI+Radioligand** | | | | | | | | | | | | |
| **ENZA-p**, 2024 |  |  |  |  |  |  |  |  |  |  |  | Overall |
| **ARPI+TKI** | | | | | | | | | | | | |
| **Dorff TB et al**, 2019 | Overall | Overall |  |  |  |  |  |  |  |  |  |  |
| **Spetsieris N et al**, 2021 |  |  | Overall | Overall |  |  |  |  |  |  |  |  |
| **ASO monotherapy** | | | | | | | | | | | | |
| **Yu EY et al**, 2018 |  | Overall |  |  |  |  |  |  |  |  |  |  |
| **Tolcher AW et al**, 2002 |  |  |  |  |  |  |  |  |  |  |  |  |
| **ASO+Chemotherapy** | | | | | | | | | | | | |
| **P-06c**, 2011 |  |  |  |  | Overall | Overall |  |  |  |  |  |  |
| **Aurora kinase inhibitor monotherapy** | | | | | | | | | | | | |
| **Meulenbeld HJ et al**, 2012 |  |  |  |  |  | Overall |  |  |  |  |  |  |
| **Bipolar Androgen Therapy** | | | | | | | | | | | | |
| **TRANSFORMER**, 2021 |  |  | Overall | Overall |  |  |  |  |  |  |  |  |
| **Chemotherapy monotherapy** | | | | | | | | | | | | |
| **Heidenreic A et al**, 2004 |  |  |  |  |  |  |  |  |  |  |  |  |
| **Annala M et al**, 2021 | No Docetaxel  Subgroup | No Docetaxel Subgroup |  |  | Docetaxel Subgroup | Docetaxel Subgroup |  |  |  |  | Overall | Overall |
| **TAXYNERGY**, 2017 |  |  |  |  |  |  |  |  |  |  |  |  |
| **TIPC**, 2007 | Overall | Overall |  |  |  |  |  |  |  |  |  |  |
| **GETUG-P02**, 2015 |  |  |  |  | Overall | Overall |  |  |  |  |  |  |
| **Krainer M et al**, 2007 |  | Overall |  |  |  |  |  |  |  |  |  |  |
| **ConCab**, 2018 |  |  |  |  |  |  | Overall | Overall |  |  |  |  |
| **CAINTA**, 2023 |  |  |  |  | Overall | Overall |  |  |  |  |  |  |
| **Rosenberg JE et al**, 2007 |  |  |  |  | Overall |  |  |  |  |  |  |  |
| **Chemotherapy+AKTi** | | | | | | | | | | | | |
| **ProCAID**, 2021 |  |  | ARPI Subgroup | ARPI Subgroup |  |  |  |  |  |  | Overall | Overall |
| **RE-AKT**, 2024 |  |  |  |  |  |  | Overall | Overall |  |  |  |  |
| **Chemotherapy+Antifungal** | | | | | | | | | | | | |
| **Millikan R et al**, 2003 | Overall |  |  |  |  |  |  |  |  |  |  |  |
| **Chemotherapy+ARPI** | | | | | | | | | | | | |
| **ABIDO SOGUG**, 2022 |  |  | Overall | Overall |  |  |  |  |  |  |  |  |
| **CHEIRON**, 2021 | Overall | Overall |  |  |  |  |  |  |  |  |  |  |
| **Chemotherapy+ASO** | | | | | | | | | | | | |
| **Chi KN et al**, 2010 | Overall | Overall |  |  |  |  |  |  |  |  |  |  |
| **Wiechno P et al**, 2014 | Overall | Overall |  |  |  |  |  |  |  |  |  |  |
| **Chemotherapy+Bcl-2 inhibitor** | | | | | | | | | | | | |
| **EORTC**, 2009 |  | Overall |  |  |  |  |  |  |  |  |  |  |
| **Sonpavde G et al**, 2011 | Overall | Overall |  |  |  |  |  |  |  |  |  |  |
| **Chemotherapy+Chemotherapy** | | | | | | | | | | | | |
| **Machiels JP et al**, 2008 | Overall | Overall |  |  |  |  |  |  |  |  |  |  |
| **Berry WR et al**, 2004 | Overall | Overall |  |  |  |  |  |  |  |  |  |  |
| **Albrecht W et al**, 2004 | Overall |  |  |  |  |  |  |  |  |  |  |  |
| **Cabrespine A et al**, 2006 | Overall |  |  |  |  |  |  |  |  |  |  |  |
| **Nelius T et al**, 2006 | Overall |  |  |  |  |  |  |  |  |  |  |  |
| **Oudard S et al**, 2005 | Overall |  |  |  |  |  |  |  |  |  |  |  |
| **MDACC Study**, 2019 |  |  | Overall | No Docetaxel Subgroup |  |  |  |  |  |  |  |  |
| **RECARDO**, 2018 |  |  |  |  | Overall | Overall |  |  |  |  |  |  |
| **Droz JP et al**, 2003 | Overall | Overall |  |  |  |  |  |  |  |  |  |  |
| **Galsky MD et al**, 2005 |  |  |  |  |  |  |  |  |  |  |  |  |
| **Eymard JC et al**, 2007 | Overall | Overall |  |  |  |  |  |  |  |  |  |  |
| **Chemotherapy+Curcuminoid** | | | | | | | | | | | | |
| **Jahanmohan JP et al**, 2021 | Overall | Overall |  |  |  |  |  |  |  |  |  |  |
| **Chemotherapy+Immunotherapy** | | | | | | | | | | | | |
| **Dahut WL et al**, 2004 | Overall | Overall |  |  |  |  |  |  |  |  |  |  |
| **IND 209**, 2018 |  |  |  |  |  |  |  |  |  |  | Overall |  |
| **Fizazi K et al**, 2012 |  |  |  |  | Overall | Overall |  |  |  |  |  |  |
| **ECOG 3899**, 2010 | Overall | Overall |  |  |  |  |  |  |  |  |  |  |
| **Hakenberg OW et al**, 2019 |  |  |  |  | Overall | Overall |  |  |  |  |  |  |
| **E1809**, 2015 | Overall |  |  |  |  |  |  |  |  |  |  |  |
| **Kongsted P et al**, 2017 |  |  |  |  |  |  |  |  |  |  |  | Overall |
| **de Bono et al**, 2014 |  | Overall |  |  |  |  |  |  |  |  |  |  |
| **Heidenreich A et al**, 2013 | Overall | Overall |  |  |  |  |  |  |  |  |  |  |
| **Chemotherapy+Radiopharmaceutical** | | | | | | | | | | | | |
| **Taxium II**, 2017 | Overall | Overall |  |  |  |  |  |  |  |  |  |  |
| **Chemotherapy+TKI** | | | | | | | | | | | | |
| **Horti J et al**, 2009 |  | Overall |  |  |  |  |  |  |  |  |  |  |
| **Chemotherapy+Vascular disrupting agent** | | | | | | | | | | | | |
| **Pili R et al**, 2010 | Overall | Overall |  |  |  |  |  |  |  |  |  |  |
| **Chemotherapy+VitD-Analog** | | | | | | | | | | | | |
| **Attia S et al**, 2008 | Overall | Overall |  |  |  |  |  |  |  |  |  |  |
| **ASCENT**, 2007 | Overall |  |  |  |  |  |  |  |  |  |  |  |
| **Corticosteroid monotherapy** | | | | | | | | | | | | |
| **Venkitaraman R et al**, 2015 |  |  |  |  |  |  |  |  |  |  |  |  |
| **Corticosteroid+Immunotherapy** | | | | | | | | | | | | |
| **Yoshimura K et al**, 2016 | Overall |  |  |  |  |  |  |  |  |  |  |  |
| **Corticosteroid+VEGFi** | | | | | | | | | | | | |
| **Stadler WM et al**, 2004 |  | Overall |  |  |  |  |  |  |  |  |  |  |
| **ERA monotherapy** | | | | | | | | | | | | |
| **James ND et al**, 2009 | Overall | Overall |  |  |  |  |  |  |  |  |  |  |
| **Carducci MA et al**, 2003 |  | Overall |  |  |  |  |  |  |  |  |  |  |
| **IMiD monotherapy** | | | | | | | | | | | | |
| **Pili R et al**, 2011 | Overall | Overall |  |  |  |  |  |  |  |  |  |  |
| **Immunotherapy monotherapy** | | | | | | | | | | | | |
| **PERSEUS**, 2016 |  | Overall |  |  |  |  |  |  |  |  |  |  |
| **Filaci G et al**, 2021 |  |  |  |  | Overall | Overall |  |  |  |  |  |  |
| **Figg WD et al**, 2001 |  |  |  |  |  |  |  |  |  |  |  |  |
| **Immunotherapy+Chemotherapy** | | | | | | | | | | | | |
| **Arlen PM et al**, 2006 |  | Overall |  |  |  |  |  |  |  |  |  |  |
| **Immunotherapy+GMCSF** | | | | | | | | | | | | |
| **TBC-PRO-002**, 2010 | Overall | Overall |  |  |  |  |  |  |  |  |  |  |
| **Integrin inhibitor monotherapy** | | | | | | | | | | | | |
| **Bradley DA et al**, 2011 |  | Overall |  |  |  |  |  |  |  |  |  |  |
| **Matrix Metalloproteinase Inhibitor monotherapy** | | | | | | | | | | | | |
| **Lara Jr PN et al**, 2006 | Overall |  |  |  |  |  |  |  |  |  |  |  |
| **PARPi+ARPi** | | | | | | | | | | | | |
| **BRCAAway**, 2024 |  |  |  |  |  |  |  |  |  |  |  | Overall |
| **Progestin monotherapy** | | | | | | | | | | | | |
| **Patel SR et al**, 1990 | Overall |  |  |  |  |  |  |  |  |  |  |  |
| **Radioligand monotherapy** | | | | | | | | | | | | |
| **TheraP**, 2021 |  |  |  |  |  |  | Overall | Overall |  |  |  |  |
| **Somatostatin+Corticosteroid** | | | | | | | | | | | | |
| **Dimopoulos MA et al**, 2004 | Overall | Overall |  |  |  |  |  |  |  |  |  |  |
| **TKI monotherapy** | | | | | | | | | | | | |
| **Droz JP et al**, 2014 |  |  |  |  | Overall | Overall |  |  |  |  |  |  |
| **Boccardo F et al**, 2008 | Overall | Overall |  |  |  |  |  |  |  |  |  |  |
| **Monk P et al**, 2018 |  |  |  |  |  |  |  |  |  |  |  | Overall |
| **Smith DC et al**, 2013 |  |  |  |  |  |  |  |  |  |  |  | Overall |
| **TKI+1st gen AA** | | | | | | | | | | | | |
| **Azad AA et al**, 2014 |  |  |  |  |  |  |  |  |  |  |  |  |
| **Sridhar SS et al**, 2015 |  | Overall |  |  |  |  |  |  |  |  |  |  |

Abbreviations: ADT: androgen deprivation therapy; ARPI: androgen-receptor pathway inhibitor; TKI: tyrosine kinase inhibitor; ASO: antisense oligonucleotide; AKTi: protein kinase B inhibitor; Bcl-2: B-cell lymphoma 2 protein; VitD: vitamin d; VEGFi: vascular endothelial growth factor inhibitor; ERA: endothelin receptor antagonist; IMiD: immunomodulatory drug; GMCSF: granulocyte-macrophage colony-stimulating factor; AA: antiandrogen; PARPi: poly(ADP-ribose) polymerase inhibitor; 177Lu: 177Lu-PSMA-617

### **Supplement Table 10:** Summary of population characteristics of included phase III trials

| **Trial** | **Arm** | | **Age in years \| median (range)** | **ECOG PS** | | | **Gleason Score ≥ 8 \|  N (%)** | **Bone Metastasis \| N (%)** | **Nodal Metastasis \| N (%)** | **Visceral Metastasis \| N (%)** | **Liver Metastasis \| N (%)** | **Lung Metastasis \| N (%)** | **Baseline PSA \|  Median (IQR)** |
| --- | --- | --- | --- | --- | --- | --- | --- | --- | --- | --- | --- | --- | --- |
|  |  |  |  | **0 \| N (%)** | **1 \| N (%)** | **2 \| N (%)** |  |  |  |  |  |  |  |
| **Prior ADT only** | | | | | | | | | | | | | |
| **ARPI monotherapy** | | | | | | | | | | | | | |
| **COU-AA-302**, 2013 | Rx: Abiraterone Ctrl: Placebo | | Rx: 71  (44–95) Ctrl: 70  (44–90) | NA | NA | NA | 517 (48) | 884 (82) | NA | NA | NA | NA | Rx: 42.0  (0.0–3927.4) *  Ctrl: 37.7 (0.7–6606.4) * |
| **PREVAIL**, 2014 | Rx: Enzalutamide Ctrl: Placebo | | Rx: 72  (43-93) Ctrl: 71  (42-93) | 1169 (68) | 548 (32) | NA | 847 (49) | 1431 (83) | 871 (51) | 204 (12) | 74 (4) | 139 (8) | Rx: 54.1 (0.1–3182.0) *  Ctrl: 44.2 (0.3–3637.0) * |
| **ELM-PC 4**, 2015 | Rx: TAK-700 Ctrl: Placebo | | Rx: 71  (65–77) ^†^  Ctrl: 72  (66–77) ^†^ | 1046 (67) | 512 (33) | 2 (0.1) | 753 (48) | 1435 (92) | 675 (43) | 277 (18) | 72 (5) | 141 (9) | Rx: 55.8  (19.8–150) Ctrl: 55.3  (18–150) |
| **ARPI+ARPI** | | | | | | | | | | | | | |
| **ACIS**, 2021 | Rx: Abiraterone  + Apalutamide Ctrl: Abiraterone | | Rx: 71  (66–78) ^†^  Ctrl: 71  (65–77) ^†^ | 669 (68) | 313 (32) | NA | 518 (53) | 829 (84) | 465 (48) | 143 (15) | 41 (4) | 103 (11) | Rx: 32.3  (11.5–91.4) Ctrl: 31.2 (12.2-106.5) |
| **AllianceA031201**, 2023 | Rx: Enzalutamide + Abiraterone Ctrl: Enzalutamide | | NA | 763 (58) | 548 (42) | NA | 727 (55) | 1084 (83) | 630 (48) | NA | 56 (4) | 151 (12) | Rx: 24.3 (9.8-69.7) Ctrl: 23.8 (9.0-69.8) |
| **ARPI+PI3K/AKTi** | | | | | | | | | | | | | |
| **IPATential150**, 2021 | Rx: Abiraterone + Ipatasertib Ctrl: Abiraterone | | Rx: 69  (47–93) Ctrl: 70  (44–90) | 822 (75) | 274 (25) | 1 (0.1) | 697 (63) | 927 (84) | 430 (39) | 131 (12) | NA | NA | Rx: 21.3 (0.1–7264) *  Ctrl: 9.3 (0.1–2867.3) * |
| **ARPI+Radiopharmaceutical** | | | | | | | | | | | | | |
| **ERA 223**, 2019 | Rx: Abiraterone + Radium-223 Ctrl: Abiraterone | | Rx: 71  (65–77) ^†^ Ctrl: 71  (66–77) ^†^ | 543 (67) | 258 (32) | NA | 479 (59) | 806 (100) | NA | NA | NA | NA | Rx: 30  (12–92)  Ctrl: 31  (11–77) |
| **Chemotherapy monotherapy** | | | | | | | | | | | | | |
| **CALGB9182**, 1999 | Rx: Mitoxantrone + Hydrocortisone Ctrl: Hydrocortisone | | Rx: 72  (67-75) ^†^  Ctrl: 72  (65-75) ^†^ | NA | NA | NA | NA | 219 (91) | 46 (19) | NA | 29 (12) | 22 (9) | Rx: 150  (52-362) Ctrl:141  (54-416) |
| **FIRSTANA**, 2017 | Rx1: Cabazitaxel20 Rx2: Cabazitaxel25 Ctrl: Docetaxel | | Rx1: 68  (44-90) Rx2: 68.5  (42-85) Ctrl: 69  (41-87) | 1168 (96) | | 48 (4) | NA | 1046 (90) | 626 (54) | 261 (22) | 106 (9) | 155 (13) | Rx1: 76 (0-3289.3) *  Rx2: 80  (0.1-6312.7) * Ctrl: 73.9 (2.4-6862.0) * |
| **Kellokumpu-Lehtinen PL et al**, 2013 | Rx: Docetaxel (2W) Ctrl: Docetaxel | | Rx: 68  (46-85) Ctrl: 69  (45-87) | 114 (33) | 211 (61) | 21 (6) | NA | 302 (88) | 162 (47) | NA | 24 (7) | 18 (5) | Rx: 116  (45-271) Ctrl: 109  (52-246) |
| **Berry W et al**, 2002 | Rx: Mitoxantrone Ctrl: Placebo | | Rx: 70  (49-87) Ctrl: 74  (51-90) | 89 (75) | 29 (24) | 1 (1) | NA | 98 (83) | 21 (18) | NA | 2 (2) | 5 (4) | Rx: 56.7  (3.7-2375) *  Ctrl: 71  (1.1-1233) * |
| **Abratt RP et al**, 2004 | Rx: Vinorelbine + Hydrocortisone Ctrl: Hydrocortisone | | Rx: 70  (48–86) Ctrl: 68  (48–83) | NA | NA | NA | 131 (32) | 184 (45) | NA | NA | NA | NA | Rx: 100  (5–2005) *  Ctrl: 82  (4–4919) * |
| **PRINCE**, 2018 | Rx: Docetaxel (int) Ctrl: Docetaxel | | Rx: 70  (51–86) Ctrl: 69  (54–87) | NA | NA | NA | 77 (49) | NA | NA | NA | NA | NA | Rx: 80  (1.0–7850) *  Ctrl: 64  (1.7–18939) * |
| **TAX327**, 2004 | Rx1: Docetaxel Rx2: Docetaxel30(W) Ctrl: Mitoxantrone | | Rx1: 68  (42-92) Rx2: 69  (36-92) Ctrl: 68  (43-86) | NA | NA | NA | 300 (30) | 914 (91) | NA | 227 (23) | NA | NA | Rx1: 114 Rx2: 108 Ctrl: 123 |
| **Chemotherapy+ASO** | | | | | | | | | | | | | |
| **SYNERGY**, 2017 | Rx: Docetaxel + Custirsen Ctrl: Docetaxel | | Rx: 69  (39-86) Ctrl: 69  (47–88) | NA | NA | NA | 577 (56) | 886 (87) | 623 (61) | 234 (23) | NA | NA | Rx: 86  (0–5471) * Ctrl: 78  (0–9966) * |
| **Chemotherapy+Chemotherapy** | | | | | | | | | | | | | |
| **SWOG-99-16**, 2004 | Rx: Docetaxel60 + Estramustine Ctrl: Mitoxantrone | | Rx: 70  (47-88) Ctrl: 70  (43-87) | 600 (89) | | 74 (11) ^‡^ | NA | 579 (86) | 168 (25) | 123 (18) | 57 (8) | 66 (10) | Rx: 84  (0.1–10820) * Ctrl: 90  (0.1–8378) * |
| **Chemotherapy+DES** | | | | | | | | | | | | | |
| **ECOG 3882** 2003 | Rx: Doxorubicin + DES Ctrl: Doxorubicin | | NA | 85 (57) | | NA | NA | 144 (96) | NA | NA | NA | NA | NA |
| **Chemotherapy+ERA** | | | | | | | | | | | | | |
| **ENTHUSE (M1c)**, 2013 | Rx: Docetaxel + Zibotentan Ctrl: Docetaxel | | Rx: 68  (42–90) Ctrl: 68  (40–86) | 586 (56) | 466 (44) | NA | NA | 1052 (100) | NA | NA | NA | NA | Rx: 83.3 Ctrl: 101 |
| **SWOG S0421**, 2013 | Rx: Docetaxel + Atrasentan Ctrl: Docetaxel | | Rx: 69 (40-92) Ctrl: 69  (43-89) | NA | NA | NA | 547 (55) | 409 (42) | 297 (30) | 195 (20) | NA | NA | Rx: 79  (23.5-228.3) Ctrl: 67.7  (24.6-202.4) |
| **Chemotherapy+IMiD** | | | | | | | | | | | | | |
| **MAINSAIL**, 2015 | Rx: Docetaxel + Lenalidomide Ctrl: Docetaxel | | Rx: 70  (43-89) Ctrl: 70  (47-90) | 509 (48) | 503 (48) | 45 (4) | NA | 326 (31) | NA | NA | NA | NA | Rx: 105  (33.9–306) Ctrl: 84.9  (32.1–272) |
| **Chemotherapy+PDGFRi** | | | | | | | | | | | | | |
| **Mathew P et al**, 2007 | Rx: Docetaxel30 (W) + Imatinib Ctrl: Docetaxel30 (W) | | NA | 109 (94) | | 7 (6) | NA | 116 (100) | NA | NA | NA | NA | NA |
| **Chemotherapy+TKI** | | | | | | | | | | | | | |
| **READY**, 2013 | Rx: Docetaxel + Dasatinib Ctrl: Docetaxel | | Rx: 69  (45–92) Ctrl: 68  (40–90) | 737 (48) | 700 (46) | 85 (6) | NA | 593 (39) | NA | NA | NA | NA | Rx: 77  (0–6818) *  Ctrl: 87  (0–8318) * |
| **Chemotherapy+VEGFi** | | | | | | | | | | | | | |
| **CALGB 90401**, 2012 | Rx: Docetaxel + Bevacizumab Ctrl: Docetaxel | | Rx: 68.8  (63-74.4) ^†^ Ctrl: 69.3  (62.4-75.6) ^†^ | 587 (56) | 414 (39) | 47 (5) | NA | 898 (86) | 446 (42) | 163 (16) | 74 (7) | 111 (11) | Rx: 88  (31-237) Ctrl: 82  (31-239) |
| **VENICE**, 2013 | Rx: Docetaxel + Aflibercept Ctrl: Docetaxel | | Rx: 68  (43–88) Ctrl: 68  (40–87) | 568 (46) | 602 (49) | 54 (4) | 573 (47) | 1085 (89) | 664 (54) | 347 (28) | NA | NA | Rx: 82.6  (0–6138) *  Ctrl: 92.9  (0–3821) * |
| **Chemotherapy+VitD-Analog** | | | | | | | | | | | | | |
| **ASCENT2**, 2011 | Rx: Docetaxel + Calcitriol Ctrl: Docetaxel | | Rx: 70.4 Ctrl: 70.9 | 436 (46) | 463 (49) | 54 (6) | NA | 834 (88) | 342 (36) | NA | 55 (6) | 46 (5) | Rx: 73.1  (0.02-3942) *  Ctrl: 64  (0.04-11952) * |
| **ERA monotherapy** | | | | | | | | | | | | | |
| **ENTHUSE (PF)**, 2012 | Rx: Zibotentan Ctrl: Placebo | | Rx: 71  (46-90) Ctrl: 71  (46-95) | 452 (76) | 142 (24) | NA | NA | 592 (99.5) | NA | NA | NA | NA | Rx: 52.9  (0.7-1860) *  Ctrl: 52.6  (0.1-5172) * |
| **Carducci MA et al**, 2007 | Rx: Atrasentan Ctrl: Placebo | | Rx: 73  (45–93) Ctrl: 72  (45–92) | NA | NA | NA | NA | NA | NA | NA | NA | NA | Rx: 69.8  (1.7–5784) *  Ctrl: 79.6  (2.2–5424.8) * |
| **IMiD monotherapy** | | | | | | | | | | | | | |
| **Sternberg C et al**, 2016 | Rx: Tasquinimod Ctrl: Placebo | | Rx: 71  (43-92) Ctrl: 71  (48-92) | NA | NA | NA | 588 (47) | 1233 (99) | 476 (38) | 237 (19) | NA | NA | Rx: 54.3  (0.6-8710.7) *  Ctrl: 50.1  (0.2-5679.5) * |
| **Immunotherapy monotherapy** | | | | | | | | | | | | | |
| **CA184-095**, 2017 | Rx: Ipilimumab Ctrl: Placebo | | Rx: 70  (44-91) Ctrl: 69  (42-92) | 450 (75) | 151 (25) | 1 (0.2) | 283 (47) | 471 (79) | NA | NA | NA | NA | Rx: 41.2  (0.05-4956) *  Ctrl: 49.5  (0.01-9297) * |
| **D9901**, 2006 | Rx: Sipuleucel-T  Ctrl: Placebo | | Rx: 73  (47-85) Ctrl: 71  (50-86) | 99 (78) | 28 (22) | NA | 52 (41) | 118 (93) | NA | NA | NA | NA | Rx: 46  (3.5-3621) *  Ctrl: 47.9  (7.9-2799) * |
| **D9902A**, 2009 | Rx: Sipuleucel-T  Ctrl: Placebo | | Rx: 70  (51-84) Ctrl: 71  (57-87) | 74 (76) | NA | NA | 36 (37) | 98 (86) | NA | NA | NA | NA | Rx: 61.3  (8.0-936.5) *  Ctrl: 44  (8.2-1342.5) * |
| **Immunotherapy+GMCSF** | | | | | | | | | | | | | |
| **PROSPECT**, 2019 | Rx1: PVAC + GMCSF Rx2: PVAC Ctrl: Placebo | | NA | 962 (74) | 330 (25) | 3 (0.2) | NA | 969 (75) | 203 (16) | 109 (8) | NA | NA | NA |
| **PARPi+ARPI** | | | | | | | | | | | | | |
| **TALAPRO-2**, 2023 | Rx: Talazoparib + Enzalutamide Ctrl: Enzalutamide | | Rx: 71  (66–76) ^†^  Ctrl: 71  (65–76) ^†^ | 530 (66) | 275 (34) | NA | 564 (70) | 691 (86) | 314 (39) | 134 (17) | 28 (4) | 106 (13) | Rx: 18.2  (6.9–59.4) Ctrl: 16.2  (6.4–53.4) |
| **Phenylurea monotherapy** | | | | | | | | | | | | | |
| **Small EJ et al**, 2000 | Rx: Suramin Ctrl: Placebo | | Rx: 68  (43-86) Ctrl: 68  (39-87) | NA | NA | NA | NA | 306 (67) | NA | NA | NA | NA | Rx: 162  (6-4200) *  Ctrl: 186  (6-7898) * |
| **Prior ADT+ARPI** | | | | | | | | | | | | | |
| **ARPI+ARPI** | | | | | | | | | | | | | |
| **PLATO**, 2018 | Rx: Enzalutamide + Abiraterone Ctrl: Abiraterone | | Rx: 72  (67-77) ^†^  Ctrl: 71  (65-77) ^†^ | 163 (65) | 88 (35) | NA | 138 (55) | 225 (90) | 108 (43) | 13 (5) | NA | NA | Rx: 14.4 (5.2-34.2) Ctrl: 11  (4.9-48.2) |
| **ARPI+Immunotherapy** | | | | | | | | | | | | | |
| **IMbassador250**, 2022 | Rx: Enzalutamide + Atezolizumab Ctrl: Enzalutamide | | Rx: 70  (51–91) Ctrl: 70  (40–92) | NA | NA | NA | NA | 680 (90) | 225 (30) | 269 (35) | 85 (11) | NA | NA |
| **Chemotherapy+ARPI** | | | | | | | | | | | | | |
| **PRESIDE**, 2022 | Rx: Docetaxel  + Enzalutamide Ctrl: Docetaxel | | Rx: 71.5  (65–75) ^†^  Ctrl: 69  (65–74) ^†^ | 128 (47) | 131 (48) | 12 (4) | 153 (56) | 104 (39) | NA | NA | NA | NA | Rx: 36.9  (13.6-80.8) Ctrl: 28.1  (13.7-92.3) |
| **Chemotherapy+Immunotherapy** | | | | | | | | | | | | | |
| **KEYNOTE-921**, 2025 | Rx: Docetaxel  + Pembrolizumab Ctrl: Docetaxel | | Rx:71  (43-89)  Ctrl: 71  (47-92) | 584 (57) | 439 (43) | NA | 672 (65) | 507 (49) | NA | NA | 67 (7) | NA | Rx: 30.4  (0.1-3586) *  Ctrl: 28.7  (0.1-5000) * |
| **PARPi monotherapy** | | | | | | | | | | | | | |
| **TRITON-3**, 2023 | Rx: Rucaparib Ctrl: Enzalutamide /Abiraterone /Docetaxel | | Rx: 70  (45–90) Ctrl: 71  (47–92) | 200 (49) | 205 (51) | NA | 269 (66) | 349 (86) | 178 (44) | 120 (30) | 34 (8) | NA | Rx: 26.9  (0.1–1247) *  Ctrl: 28.8  (0–1039) * |
| **PROfound**, 2020 | Rx: Olaparib Ctrl: Enzalutamide /Abiraterone | | Rx: 69 (47–91) Ctrl: 69  (49–87) | 186 (48) | 183 (47) | 17 (4) | 278 (72) | 124 (32) | NA | 112 (29) | NA | NA | Rx: 68.2  (24.1-294.4) Ctrl: 106.5  (37.2-326.6) |
| **Radioligand monotherapy** | | | | | | | | | | | | | |
| **PSMAfore**, 2024 | Rx: 177Lu Ctrl: Enzalutamide /Abiraterone | | Rx: 71  (65-77) ^†^ Ctrl: 72  (67-77) ^†^ | 262 (56) | 200 (43) | NA | 247 (53) | 408 (87) | 149 (32) | 64 (14) | 20 (4) | 44 (9) | Rx: 18.4  (7.0–51.7)  Ctrl: 14.9  (5.9–38.3) |
|  | | **Prior ADT+Docetaxel** | | | | | | | | | | | |
| **ARPI monotherapy** | | | | | | | | | | | | | |
| **AFFIRM**, 2012 | Rx: Enzalutamide Ctrl: Placebo | | Rx: 69  (41-92) Ctrl: 69  (49-89) | 1097 (91) | | 102 (9) | 559 (47) | 1099 (92) | 661 (55) | 307 (26) | 126 (11) | 181 (15) | Rx: 107.7  (0.2-11794.1) *  Ctrl: 128.3  (0.0-19000) * |
| **COU-AA-301**, 2011 | Rx: Abiraterone Ctrl: Placebo | | Rx: 69  (42-95) Ctrl: 69  (39-90) | 1068 (90) | | 127 (11) | 545 (46) | 1066 (89) | 525 (44) | 335 (28) | 120 (10) | 148 (12) | Rx: 128.8 (0.4–9253) *  Ctrl: 137.7 (0.6–10114) * |
| **ELM-PC 5**, 2015 | Rx: TAK-700 Ctrl: Placebo | | Rx: 69.5  (43-89) Ctrl: 70  (48-87) | 454 (41) | 560 (51) | 92 (8) | 543 (49) | 1039 (94) | 515 (47) | 296 (27) | 108 (10) | 129 (12) | Rx: 122.5  (0-8456) *  Ctrl: 134  (1-19009) * |
| **SAKK 08/11**, 2016 | Rx: TAK-700 Ctrl: Placebo | | Rx: 70  (61–74) ^†^  Ctrl: 70.5  (66-76.5) ^†^ | 25 (53) | 20 (43) | 2 (4) | 29 (62) | 37 (79) | 27 (57) | NA | 3 (6) | 5 (11) | NA |
| **Chemotherapy monotherapy** | | | | | | | | | | | | | |
| **PROSELICA**, 2017 | Rx: Cabazitaxel20 Ctrl: Cabazitaxel25 | | Rx: 68.2  (7.2) ^§^  Ctrl: 68.4  (7.8) ^§^ | 1079 (90) | | 121 (10) | NA | 1128 (95) | 593 (49) | NA | 184 (15) | 189 (16) | Rx: 159.5 Ctrl: 170.9 |
| **TROPIC**, 2010 | Rx: Cabazitaxel25 Ctrl: Mitoxantrone | | Rx: 68  (62-73) ^†^  Ctrl: 67  (61-72) ^†^ | 694 (92) | | NA | NA | 631 (84) | NA | 188 (25) | NA | NA | Rx: 143.9  (51.1-416.0) Ctrl: 127.5  (44.0-419.0) |
| **Chemotherapy+ASO** | | | | | | | | | | | | | |
| **AFFINITY**, 2017 | Rx: Cabazitaxel25  + Custirsen Ctrl: Cabazitaxel25 | | Rx: 68  (46–86) ^†^ Ctrl: 68  (47–88) ^†^ | NA | NA | NA | NA | 589 (93) | 346 (55) | 223 (35) | NA | NA | NA |
| **TKI monotherapy** | | | | | | | | | | | | | |
| **SUN 1120**, 2013 | Rx: Sunitinib Placebo | | Rx: 69 (39-90) Ctrl: 68  (47-86) | 437 (50) | 436 (50) | NA | 425 (49) | NA | NA | NA | NA | NA | NA |
|  | | **Prior ADT+Docetaxel+ARPI** | | | | | | | | | | | |
| **Chemotherapy monotherapy** | | | | | | | | | | | | | |
| **CARD**, 2019 | Rx: Cabazitaxel25 Ctrl: Enzalutamide /Abiraterone | | Rx: 70  (46–85) Ctrl: 71  (45–88) | 242 (95) | | 13 (5) | 154 (60) | 215 (86) | 14 (5) | 46 (18) | 29 (11) | 17 (7) | Rx: 62  (1.1–15000) *  Ctrl: 60.5  (1.5–2868) * |
| **PARPi+Immunotherapy** | | | | | | | | | | | | | |
| **KEYLYNK-010**, 2023 | Rx: Olaparib + Pembrolizumab Ctrl: Enzalutamide /Abiraterone | | Rx: 71  (40-89) Ctrl: 69  (49-84) | 394 (50) | 397 (50) | 2 (0.3) | 551 (69) | 333 (42) | NA | NA | 84 (11) | NA | Rx: 52.9  (0.1--5000) *  Ctrl: 42.6  (0.1-4007) * |
| **Radioligand therapy+Standard of Care** | | | | | | | | | | | | | |
| **VISION**, 2021 | Rx: 177Lu + SOC Ctrl: SOC | | Rx: 70  (48–94) Ctrl: 71.5  (40–89) | 768 (92) | | 63 (8) | 494 (59) | 760 (91) | 415 (50) | 178 (21) | 101 (12) | 77 (9) | Rx: 77.5  (0–6988) *  Ctrl: 74.6  (0–8995) * |
| **TKI monotherapy** | | | | | | | | | | | | | |
| **COMET-1**, 2016 | Rx: Cabozantinib Ctrl: Prednisone | | Rx: 69.5  (35-87) Ctrl: 69  (43-89) | 908 (88) | | 119 (12) | 460 (45) | 1027 (100) | 453 (44) | 191 (19) | 126 (12) | 98 (10) | NA |
|  | | **Heterogeneous Prior Therapy** | | | | | | | | | | | |
| **Chemotherapy+Immunotherapy** | | | | | | | | | | | | | |
| **VIABLE**, 2022 | Rx: Docetaxel + DCVAC Ctrl: Docetaxel | | Rx: 68  (46-89) Ctrl: 69  (46-89) | 716 (61) | 450 (38) | 16 (1) | 654 (55) | 1062 (91) | 588 (50) | 158 (13) | 80 (7) | 119 (10) | Rx: 46.4  (0.0–75000) *  Ctrl: 54.0  (0.1–5000.0) * |
|  | | **Multiple subgroups (ADT/ADT+ARPI/ADT+Docetaxel/ADT+ARPI+Docetaxel/Heterogeneous prior therapy)** | | | | | | | | | | | |
| **Chemotherapy monotherapy** | | | | | | | | | | | | | |
| **SPARC**, 2009 | Rx: Satraplatin Ctrl: Placebo | | Rx: 70 (42-88) Ctrl: 68  (45-95) | 837 (89) | | 101 (11) | NA | NA | NA | NA | NA | NA | NA |
| **Immunotherapy monotherapy** | | | | | | | | | | | | | |
| **IMPACT**, 2010 | Rx: Sipuleucel-T  Ctrl: Placebo | | Rx: 72  (49–91) Ctrl: 70  (40–89) | 419 (82) | NA | NA | 126 (25) | 473 (92) | NA | NA | NA | NA | Rx: 51.7 Ctrl: 47.2 |
| **PARPi+ARPI** | | | | | | | | | | | | | |
| **PROpel**, 2023 | Rx: Olaparib + Abiraterone Ctrl: Abiraterone | | Rx: 69  (63–74) ^†^ Ctrl: 70  (65–76) ^†^ | 558 (70) | 236 (30) | NA | 523 (66) | 688 (86) | 252 (32) | 115 (14) | 33 (4) | 82 (10) | Rx: 17.9  (6.09–67.0) Ctrl: 16.8  (6.3–53.3) |
| **MAGNITUDE**, 2023 | Rx: Niraparib + Abiraterone Ctrl: Abiraterone | | Rx: 69  (45-100) Ctrl: 69  (43-88) | 276 (65) | 147 (35) | NA | 286 (68) | 353 (83) | 208 (49) | 90 (21) | 31 (7) | 45 (11) | Rx: 21.4  (0-4826.5) *  Ctrl: 17.4  (0.1-4400) * |
| **Radiopharmaceutical monotherapy** | | | | | | | | | | | | | |
| **ALSYMPCA**, 2013 | Rx: Radium-223 Ctrl: Placebo | | Rx: 71  (49-90) Ctrl: 71  (44-94) | 243 (26) | 558 (61) | 118 (13) | NA | 921 (100) | NA | NA | NA | NA | Rx: 146  (3.8-6026) *  Ctrl: 173  (1.5-14500) * |

Abbreviations: PSA: prostate-specific antigen; ECOG PS: eastern cooperative oncology group performance status; ARPI: androgen-receptor pathway inhibitor; PI3K/AKTi: phosphatidylinositol 3-kinase and protein kinase B inhibitor; PDGFR: platelet-derived growth factor receptor; DES: diethylstilbestrol diphosphate; PVAC: PROSTVAC (viral vector-based immunotherapy); GMCSF: granulocyte-macrophage colony-stimulating factor; ASO: antisense oligonucleotide; ERA: endothelin receptor antagonist; IMiD: immunomodulatory drug; TKI: tyrosine kinase inhibitor; VitD: vitamin d; PARPi: poly(ADP-ribose) polymerase inhibitor; 177Lu: 177Lu-PSMA-617; SOC: standard of care

Note: PLATO and CARD were phase IV trials. All remaining trials were phase III. The color ‘green’ represents a positive trial, ‘red’ represents a negative trial.

* Median (range)

^†^ Median (IQR)

^‡^ ECOG PS 2-3

^§^ Mean (SD)

Cabazitaxel20: Cabazitaxel 20 mg/m^2^ IV on day 1 of every 3-week cycle

Cabazitaxel25: Cabazitaxel 25 mg/m^2^ IV on day 1 of every 3-week cycle

Docetaxel (2W): Docetaxel 75 mg/m^2^ IV on days 1 and 15 of a 4-week cycle

Docetaxel (int): Docetaxel 35 mg/m^2^ IV on days 1, 8, 15, repeat cycle at day 29

Docetaxel30 (W): Docetaxel 30 mg/m^2^ IV on days 1, 8, 15, 22 and 29 of a 6-week cycle

Remaining drugs were administered at standard doses.

### **Supplement Table 11:** Prior therapy administered to patients in included phase III trials

| **Trial** | **Arm** | **Prior Therapy in Any Setting** | | | | **Prior Therapy in mHSPC Setting** | | | | **Prior Therapy in mCRPC Setting** | | | |
| --- | --- | --- | --- | --- | --- | --- | --- | --- | --- | --- | --- | --- | --- |
|  |  | **Docetaxel** | **ARPI** | **Abiraterone** | **Enzalutamide** | **Docetaxel** | **ARPI** | **Abiraterone** | **Enzalutamide** | **Docetaxel** | **ARPI** | **Abiraterone** | **Enzalutamide** |
| **ARPI monotherapy** | | | | | | | | | | | | | |
| **AFFIRM**, 2012 | Rx: Enzalutamide Ctrl: Placebo | 1199 (100) | NA | NA | NA | NA | NA | NA | NA | 1199 (100) | NA | NA | NA |
| **COU-AA-301**, 2011 | Rx: Abiraterone Ctrl: Placebo | 1195 (100) | NA | NA | NA | NA | NA | NA | NA | 1195 (100) | NA | NA | NA |
| **COU-AA-302**, 2013 | Rx: Abiraterone Ctrl: Placebo | NA | NA | NA | NA | NA | NA | NA | NA | NA | NA | NA | NA |
| **PREVAIL**, 2014 | Rx: Enzalutamide Ctrl: Placebo | NA | NA | NA | NA | NA | NA | NA | NA | NA | NA | NA | NA |
| **ELM-PC 4**, 2015 | Rx: TAK-700 Ctrl: Placebo | NA | NA | NA | NA | NA | NA | NA | NA | NA | NA | NA | NA |
| **ELM-PC 5**, 2015 | Rx: TAK-700 Ctrl: Placebo | 1099 (100) | NA | NA | NA | NA | NA | NA | NA | 1099 (100) | NA | NA | NA |
| **SAKK 08/11**, 2016 | Rx: TAK-700 Ctrl: Placebo | 47  (100) | NA | NA | NA | NA | NA | NA | NA | 47  (100) | NA | NA |  |
| **ARPI+ARPI** | | | | | | | | | | | | | |
| **ACIS**, 2021 | Rx: Abiraterone  + Apalutamide Ctrl: Abiraterone | NA | NA | NA | NA | NA | NA | NA | NA | NA | NA | NA | NA |
| **AllianceA031201**, 2023 | Rx: Enzalutamide + Abiraterone Ctrl: Enzalutamide | NA | NA | NA | NA | NA | NA | NA | NA | NA | NA | NA | NA |
| **PLATO**, 2018 | Rx: Enzalutamide + Abiraterone Ctrl: Abiraterone | NA | 251 (100) | NA | 251  (100) | NA | NA | NA | NA | NA | 251 (100) | NA | 251  (100) |
| **ARPI+Immunotherapy** | | | | | | | | | | | | | |
| **IMbassador250**, 2022 | Rx: Enzalutamide + Atezolizumab Ctrl: Enzalutamide | 362  (48) | 759 (100) | NA | NA | NA | NA | NA | NA | 362  (48) | 759 (100) | NA | NA |
| **ARPI+PI3K/AKTi** | | | | | | | | | | | | | |
| **IPATential150**, 2021 | Rx: Abiraterone + Ipatasertib Ctrl: Abiraterone | 197  (18) | NA | NA | NA | 197  (18) | NA | NA | NA | NA | NA | NA | NA |
| **ARPI+Radiopharmaceutical** | | | | | | | | | | | | | |
| **ERA 223**, 2019 | Rx: Abiraterone + Radium-223 Ctrl: Abiraterone | 15  (2) | 53  (7) | NA | 53  (7) | NA | NA | NA | NA | NA | NA | NA | NA |
| **Chemotherapy monotherapy** | | | | | | | | | | | | | |
| **CALGB9182**, 1999 | Rx: Mitoxantrone + Hydrocortisone Ctrl: Hydrocortisone | NA | NA | NA | NA | NA | NA | NA | NA | NA | NA | NA | NA |
| **FIRSTANA**, 2017 | Rx1: Cabazitaxel20 Rx2: Cabazitaxel25 Ctrl: Docetaxel | NA | 23  (2) | 13  (1) | 10  (1) | NA | NA | NA | NA | NA | NA | NA | NA |
| **Kellokumpu-Lehtinen PL et al**, 2013 | Rx: Docetaxel (2W) Ctrl: Docetaxel | NA | NA | NA | NA | NA | NA | NA | NA | NA | NA | NA | NA |
| **Berry W et al**, 2002 | Rx: Mitoxantrone Ctrl: Placebo | NA | NA | NA | NA | NA | NA | NA | NA | NA | NA | NA | NA |
| **Abratt RP et al**, 2004 | Rx: Vinorelbine + Hydrocortisone Ctrl: Hydrocortisone | NA | NA | NA | NA | NA | NA | NA | NA | NA | NA | NA | NA |
| **PRINCE**, 2018 | Rx: Docetaxel (int) Ctrl: Docetaxel | NA | NA | NA | NA | NA | NA | NA | NA | NA | NA | NA | NA |
| **PROSELICA**, 2017 | Rx: Cabazitaxel20 Ctrl: Cabazitaxel25 | 1200 (100) | 323  (27) | 291  (24) | 32  (3) | NA | NA | NA | NA | 1200 (100) | 323 (27) | 291  (24) | 32  (3) |
| **SPARC**, 2009 | Rx: Satraplatin Ctrl: Placebo | 488  (51) | NA | NA | NA | NA | NA | NA | NA | 488  (51) | NA | NA | NA |
| **TAX327**, 2004 | Rx1: Docetaxel Rx2: Docetaxel30 (W) Ctrl: Mitoxantrone | NA | NA | NA | NA | NA | NA | NA | NA | NA | NA | NA | NA |
| **TROPIC**, 2010 | Rx: Cabazitaxel25 Ctrl: Mitoxantrone | 755  (100) | NA | NA | NA | NA | NA | NA | NA | 755  (100) | NA | NA | NA |
| **CARD**, 2019 | Rx: Cabazitaxel25 Ctrl: Enzalutamide /Abiraterone | 255  (100) | 254 (99.6) | 123  (48) | 131  (51) | NA | NA | NA | NA | NA | NA | NA | NA |
| **Chemotherapy+ARPI** | | | | | | | | | | | | | |
| **PRESIDE**, 2022 | Rx: Docetaxel  + Enzalutamide Ctrl: Docetaxel | NA | 271 (100) | NA | 271  (100) | NA | NA | NA | NA | NA | 271 (100) | NA | 271  (100) |
| **Chemotherapy+ASO** | | | | | | | | | | | | | |
| **AFFINITY**, 2017 | Rx: Cabazitaxel25  + Custirsen Ctrl: Cabazitaxel25 | 635  (100) | 373  (59) | NA | NA | NA | NA | NA | NA | 635  (100) | 373 (59) | NA | NA |
| **SYNERGY**, 2017 | Rx: Docetaxel + Custirsen Ctrl: Docetaxel | NA | 33  (3) | 23  (2) | 10  (1) | NA | NA | NA | NA | NA | NA | NA | NA |
| **Chemotherapy+Chemotherapy** | | | | | | | | | | | | | |
| **SWOG-99-16**, 2004 | Rx: Docetaxel60 + Estramustine Ctrl: Mitoxantrone | NA | NA | NA | NA | NA | NA | NA | NA | NA | NA | NA | NA |
| **Chemotherapy+DES** | | | | | | | | | | | | | |
| **ECOG 3882** 2003 | Rx: Doxorubicin + DES Ctrl: Doxorubicin | NA | NA | NA | NA | NA | NA | NA | NA | NA | NA | NA | NA |
| **Chemotherapy+ERA** | | | | | | | | | | | | | |
| **ENTHUSE (M1c)**, 2013 | Rx: Docetaxel + Zibotentan Ctrl: Docetaxel | NA | NA | NA | NA | NA | NA | NA | NA | NA | NA | NA | NA |
| **SWOG S0421**, 2013 | Rx: Docetaxel + Atrasentan Ctrl: Docetaxel | NA | NA | NA | NA | NA | NA | NA | NA | NA | NA | NA | NA |
| **Chemotherapy+IMiD** | | | | | | | | | | | | | |
| **MAINSAIL**, 2015 | Rx: Docetaxel + Lenalidomide Ctrl: Docetaxel | NA | NA | NA | NA | NA | NA | NA | NA | NA | NA | NA | NA |
| **Chemotherapy+Immunotherapy** | | | | | | | | | | | | | |
| **VIABLE**, 2022 | Rx: Docetaxel + DCVAC Ctrl: Docetaxel | NA | 365  (31) | 290  (25) | 197  (17) | NA | NA | NA | NA | NA | NA | NA | NA |
| **KEYNOTE-921**, 2025 | Rx: Docetaxel  + Pembrolizumab Ctrl: Docetaxel | 128  (12) | 1015 (99) | 555  (54) | 463  (45) | 128 (12) | NA | NA | NA | NA | NA | NA | NA |
| **Chemotherapy+PDGFRi** | | | | | | | | | | | | | |
| **Mathew P et al**, 2007 | Rx: Docetaxel30 (W) + Imatinib Ctrl: Docetaxel30 (W) | NA | NA | NA | NA | NA | NA | NA | NA | NA | NA | NA | NA |
| **Chemotherapy+TKI** | | | | | | | | | | | | | |
| **READY**, 2013 | Rx: Docetaxel + Dasatinib Ctrl: Docetaxel | NA | NA | NA | NA | NA | NA | NA | NA | NA | NA | NA | NA |
| **Chemotherapy+VEGFi** | | | | | | | | | | | | | |
| **CALGB 90401**, 2012 | Rx: Docetaxel + Bevacizumab Ctrl: Docetaxel | NA | NA | NA | NA | NA | NA | NA | NA | NA | NA | NA | NA |
| **VENICE**, 2013 | Rx: Docetaxel+ Aflibercept Ctrl: Docetaxel | NA | NA | NA | NA | NA | NA | NA | NA | NA | NA | NA | NA |
| **Chemotherapy+VitD-Analog** | | | | | | | | | | | | | |
| **ASCENT2**, 2011 | Rx: Docetaxel + Calcitriol Ctrl: Docetaxel | NA | NA | NA | NA | NA | NA | NA | NA | NA | NA | NA | NA |
| **ERA monotherapy** | | | | | | | | | | | | | |
| **ENTHUSE (PF)**, 2012 | Rx: Zibotentan Ctrl: Placebo | NA | NA | NA | NA | NA | NA | NA | NA | NA | NA | NA | NA |
| **Carducci MA et al**, 2007 | Rx: Atrasentan Ctrl: Placebo | NA | NA | NA | NA | NA | NA | NA | NA | NA | NA | NA | NA |
| **IMiD monotherapy** | | | | | | | | | | | | | |
| **Sternberg C et al**, 2016 | Rx: Tasquinimod Ctrl: Placebo | NA | 113  (9) | NA | NA | NA | NA | NA | NA | NA | 113 (9) | NA | NA |
| **Immunotherapy monotherapy** | | | | | | | | | | | | | |
| **CA184-095**, 2017 | Rx: Ipilimumab Ctrl: Placebo | NA | NA | NA | NA | NA | NA | NA | NA | NA | NA | NA | NA |
| **D9901**, 2006 | Rx: Sipuleucel-T  Ctrl: Placebo | NA | NA | NA | NA | NA | NA | NA | NA | NA | NA | NA | NA |
| **IMPACT**, 2010 | Rx: Sipuleucel-T  Ctrl: Placebo | 74  (14) | NA | NA | NA | NA | NA | NA | NA | NA | NA | NA | NA |
| **D9902A**, 2009 | Rx: Sipuleucel-T  Ctrl: Placebo | NA | NA | NA | NA | NA | NA | NA | NA | NA | NA | NA | NA |
| **Immunotherapy+GMCSF** | | | | | | | | | | | | | |
| **PROSPECT**, 2019 | Rx1: PVAC + GMCSF Rx2: PVAC Ctrl: Placebo | NA | NA | NA | NA | NA | NA | NA | NA | NA | NA | NA | NA |
| **PARPi monotherapy** | | | | | | | | | | | | | |
| **PROfound**, 2020 | Rx: Olaparib Ctrl: Enzalutamide /Abiraterone | 250  (65) | 387 (100) | 228  (59) | 233  (60) | NA | NA | NA | NA | NA | NA | NA | NA |
| **TRITON-3**, 2023 | Rx: Rucaparib Ctrl: Enzalutamide /Abiraterone /Docetaxel | 91  (22) | 405 (100) | 230  (57) | 180  (44) | 91  (22) | NA | NA | NA | NA | NA | NA | NA |
| **PARPi+ARPI** | | | | | | | | | | | | | |
| **MAGNITUDE**, 2023 | Rx: Niraparib + Abiraterone Ctrl: Abiraterone | 85  (20) | 111 (26) | 98  (23) | NA | 85  (20) | NA | NA | NA | NA | 98 (23) | 98  (23) | NA |
| **PROpel**, 2023 | Rx: Olaparib + Abiraterone Ctrl: Abiraterone | 195  (24) | 1 (0.1) | NA | NA | 179  (22) | 1 (0.1) | NA | NA | NA | NA | NA | NA |
| **TALAPRO-2**, 2023 | Rx: Talazoparib + Enzalutamide Ctrl: Enzalutamide | 179  (22) | 46  (6) | 46  (6) | NA | 179  (22) | 46  (6) | 46  (6) | NA | NA | NA | NA | NA |
| **PARPi+Immunotherapy** | | | | | | | | | | | | | |
| **KEYLYNK-010**, 2023 | Rx: Olaparib + Pembrolizumab Ctrl: Enzalutamide /Abiraterone | 777  (98) | 793 (100) | 433  (55) | 361  (46) | NA | NA | Given but number not specified | NA | NA | NA | Given but number not specified | 361  (46) |
| **Phenylurea monotherapy** | | | | | | | | | | | | | |
| **Small EJ et al**, 2000 | Rx: Suramin Ctrl: Placebo | NA | NA | NA | NA | NA | NA | NA | NA | NA | NA | NA | NA |
| **Radioligand monotherapy** | | | | | | | | | | | | | |
| **PSMAfore**, 2024 | Rx: 177Lu Ctrl: Enzalutamide /Abiraterone | NA | 468 (100) | 249  (53) | 177  (38) | NA | 88 (19) | NA | NA | NA | 380 (81) | NA | NA |
| **Radiopharmaceutical monotherapy** | | | | | | | | | | | | | |
| **ALSYMPCA**, 2013 | Rx: Radium-223 Ctrl: Placebo | 526  (57) | NA | NA | NA | NA | NA | NA | NA | 526  (57) | NA | NA | NA |
| **Radioligand therapy+Standard of Care** | | | | | | | | | | | | | |
| **VISION**, 2021 | Rx: 177Lu + SOC Ctrl: SOC | 807  (97) | 831 (100) | 617  (74) | 601  (72) | NA | NA | NA | NA | NA | NA | NA | NA |
| **TKI monotherapy** | | | | | | | | | | | | | |
| **COMET-1**, 2016 | Rx: Cabozantinib Ctrl: Prednisone | 1028 (100) | NA | 946  (92) | 259  (25) | NA | NA | NA | NA | 1028 (100) | NA | 946  (92) | 259  (25) |
| **SUN 1120**, 2013 | Rx: Sunitinib Placebo | 873  (100) | NA | NA | NA | NA | NA | NA | NA | 873  (100) | NA | NA | NA |

Abbreviations: mHSPC: metastatic hormone sensitive prostate cancer; mCRPC: metastatic castration resistant prostate cancer; ARPI: androgen-receptor pathway inhibitor; PI3K/AKTi: phosphatidylinositol 3-kinase and protein kinase B inhibitor; PDGFR: platelet-derived growth factor receptor; DES: diethylstilbestrol diphosphate; PVAC: PROSTVAC (viral vector-based immunotherapy); GMCSF: granulocyte-macrophage colony-stimulating factor; ASO: antisense oligonucleotide; ERA: endothelin receptor antagonist; IMiD: immunomodulatory drug; TKI: tyrosine kinase inhibitor; VitD: vitamin d; PARPi: poly(ADP-ribose) polymerase inhibitor; 177Lu: 177Lu-PSMA-617; SOC: standard of care

Note: PLATO and CARD were phase IV trials. All remaining trials were phase III. The color ‘green’ represents a positive trial, ‘red’ represents a negative trial.

Cabazitaxel20: Cabazitaxel 20 mg/m^2^ IV on day 1 of every 3-week cycle

Cabazitaxel25: Cabazitaxel 25 mg/m^2^ IV on day 1 of every 3-week cycle

Docetaxel (2W): Docetaxel 75 mg/m^2^ IV on days 1 and 15 of a 4-week cycle

Docetaxel (int): Docetaxel 35 mg/m^2^ IV on days 1, 8, 15, repeat cycle at day 29

Docetaxel30 (W): Docetaxel 30 mg/m^2^ IV on days 1, 8, 15, 22 and 29 of a 6-week cycle

Remaining drugs were administered at standard doses.

### **Supplement Table 12:** Matrix for reported overall survival and progression free survival according to prior therapy in included phase III trials

| **Trial** | **Prior ADT** | | **Prior  ADT+ARPI** | | **Prior  ADT+Docetaxel** | | **Prior  ADT+Docetaxel+ARPI** | | **Prior True Triplet Therapy** | | **Heterogeneous Prior Therapy** | |
| --- | --- | --- | --- | --- | --- | --- | --- | --- | --- | --- | --- | --- |
|  | **OS** | **PFS** | **OS** | **PFS** | **OS** | **PFS** | **OS** | **PFS** | **OS** | **PFS** | **OS** | **PFS** |
| **ARPI monotherapy** | | | | | | | | | | | | |
| **AFFIRM**, 2012 |  |  |  |  | Overall | Overall |  |  |  |  |  |  |
| **COU-AA-301**, 2011 |  |  |  |  | Overall | Overall |  |  |  |  |  |  |
| **COU-AA-302**, 2013 | Overall | Overall |  |  |  |  |  |  |  |  |  |  |
| **PREVAIL**, 2014 | Overall | Overall |  |  |  |  |  |  |  |  |  |  |
| **ELM-PC 4**, 2015 | Overall | Overall |  |  |  |  |  |  |  |  |  |  |
| **ELM-PC 5**, 2015 |  |  |  |  | Overall | Overall |  |  |  |  |  |  |
| **SAKK 08/11**, 2016 |  |  |  |  |  | Overall |  |  |  |  |  |  |
| **ARPI+ARPI** | | | | | | | | | | | | |
| **ACIS**, 2021 | Overall | Overall |  |  |  |  |  |  |  |  |  |  |
| **AllianceA031201**, 2023 | Overall | Overall |  |  |  |  |  |  |  |  |  |  |
| **PLATO**, 2018 |  |  |  | Overall |  |  |  |  |  |  |  |  |
| **ARPI+Immunotherapy** | | | | | | | | | | | | |
| **IMbassador250**, 2022 |  |  | Overall | No Taxane Subgroup |  |  |  |  |  |  |  |  |
| **ARPI+PI3K/AKTi** | | | | | | | | | | | | |
| **IPATential150**, 2021 | Overall | No Docetaxel  Subgroup |  |  |  |  |  |  |  |  |  |  |
| **ARPI+Radiopharmaceutical** | | | | | | | | | | | | |
| **ERA 223**, 2019 | Overall | Overall |  |  |  |  |  |  |  |  |  |  |
| **Chemotherapy monotherapy** | | | | | | | | | | | | |
| **CALGB9182**, 1999 | Overall | Overall |  |  |  |  |  |  |  |  |  |  |
| **FIRSTANA**, 2017 | Overall | Overall |  |  |  |  |  |  |  |  |  |  |
| **Kellokumpu-Lehtinen PL et al**, 2013 | Overall | Overall |  |  |  |  |  |  |  |  |  |  |
| **Berry W et al**, 2002 | Overall |  |  |  |  |  |  |  |  |  |  |  |
| **Abratt RP et al**, 2004 | Overall | Overall |  |  |  |  |  |  |  |  |  |  |
| **PRINCE**, 2018 | Overall | Overall |  |  |  |  |  |  |  |  |  |  |
| **PROSELICA**, 2017 |  |  |  |  | No ARPI Subgroup | Overall |  |  |  |  |  |  |
| **SPARC**, 2009 | No Docetaxel  Subgroup | No Docetaxel  Subgroup |  |  | Docetaxel  Subgroup | Docetaxel  Subgroup |  |  |  |  | Overall | Overall |
| **TAX327**, 2004 | Overall |  |  |  |  |  |  |  |  |  |  |  |
| **TROPIC**, 2010 |  |  |  |  | Overall | Overall |  |  |  |  |  |  |
| **CARD**, 2019 |  |  |  |  |  |  | Overall | Overall |  |  |  |  |
| **Chemotherapy+ARPI** | | | | | | | | | | | | |
| **PRESIDE**, 2022 |  |  |  | Overall |  |  |  |  |  |  |  |  |
| **Chemotherapy+ASO** | | | | | | | | | | | | |
| **AFFINITY**, 2017 |  |  |  |  | Overall |  |  |  |  |  |  |  |
| **SYNERGY**, 2017 | Overall |  |  |  |  |  |  |  |  |  |  |  |
| **Chemotherapy+Chemotherapy** | | | | | | | | | | | | |
| **SWOG-99-16**, 2004 | Overall | Overall |  |  |  |  |  |  |  |  |  |  |
| **Chemotherapy+DES** | | | | | | | | | | | | |
| **ECOG 3882** 2003 | Overall |  |  |  |  |  |  |  |  |  |  |  |
| **Chemotherapy+ERA** | | | | | | | | | | | | |
| **ENTHUSE (M1c)**, 2013 | Overall | Overall |  |  |  |  |  |  |  |  |  |  |
| **SWOG S0421**, 2013 | Overall | Overall |  |  |  |  |  |  |  |  |  |  |
| **Chemotherapy+IMiD** | | | | | | | | | | | | |
| **MAINSAIL**, 2015 | Overall | Overall |  |  |  |  |  |  |  |  |  |  |
| **Chemotherapy+Immunotherapy** | | | | | | | | | | | | |
| **VIABLE**, 2022 |  |  |  |  |  |  |  |  |  |  | Overall | Overall |
| **KEYNOTE-921**, 2025 |  |  | Overall | Overall |  |  |  |  |  |  |  |  |
| **Chemotherapy+PDGFRi** | | | | | | | | | | | | |
| **Mathew P et al**, 2007 | Overall | Overall |  |  |  |  |  |  |  |  |  |  |
| **Chemotherapy+TKI** | | | | | | | | | | | | |
| **READY**, 2013 | Overall | Overall |  |  |  |  |  |  |  |  |  |  |
| **Chemotherapy+VEGFi** | | | | | | | | | | | | |
| **CALGB 90401**, 2012 | Overall | Overall |  |  |  |  |  |  |  |  |  |  |
| **VENICE**, 2013 | Overall | Overall |  |  |  |  |  |  |  |  |  |  |
| **Chemotherapy+VitD-Analog** | | | | | | | | | | | | |
| **ASCENT2**, 2011 | Overall |  |  |  |  |  |  |  |  |  |  |  |
| **ERA monotherapy** | | | | | | | | | | | | |
| **ENTHUSE (PF)**, 2012 | Overall | Overall |  |  |  |  |  |  |  |  |  |  |
| **Carducci MA et al**, 2007 | Overall | Overall |  |  |  |  |  |  |  |  |  |  |
| **IMiD monotherapy** | | | | | | | | | | | | |
| **Sternberg C et al**, 2016 | Overall | Overall |  |  |  |  |  |  |  |  |  |  |
| **Immunotherapy monotherapy** | | | | | | | | | | | | |
| **CA184-095**, 2017 | Overall | Overall |  |  |  |  |  |  |  |  |  |  |
| **D9901**, 2006 | Overall | Overall |  |  |  |  |  |  |  |  |  |  |
| **IMPACT**, 2010 | No Docetaxel  Subgroup | Overall |  |  | Docetaxel  Subgroup |  |  |  |  |  |  |  |
| **D9902A**, 2009 | Overall | Overall |  |  |  |  |  |  |  |  |  |  |
| **Immunotherapy+GMCSF** | | | | | | | | | | | | |
| **PROSPECT**, 2019 | Overall |  |  |  |  |  |  |  |  |  |  |  |
| **PARPi monotherapy** | | | | | | | | | | | | |
| **PROfound**, 2020 |  |  | No Taxane  Subgroup | No Taxane  Subgroup |  |  |  |  |  |  |  |  |
| **TRITON-3**, 2023 |  |  | Overall | Overall |  |  |  |  |  |  |  |  |
| **PARPi+ARPI** | | | | | | | | | | | | |
| **MAGNITUDE**, 2023 |  |  |  | ARPI  Subgroup |  | Docetaxel  Subgroup |  |  |  |  | Overall | Overall |
| **PROpel**, 2023 | No Docetaxel  Subgroup | No Docetaxel  Subgroup |  |  | Docetaxel  Subgroup | Docetaxel  Subgroup |  |  |  |  |  |  |
| **TALAPRO-2**, 2023 | Overall | No ARPI/Taxane Subgroup |  |  |  |  |  |  |  |  |  |  |
| **PARPi+Immunotherapy** | | | | | | | | | | | | |
| **KEYLYNK-010**, 2023 |  |  |  |  |  |  | Overall | Overall |  |  |  |  |
| **Phenylurea monotherapy** | | | | | | | | | | | | |
| **Small EJ et al**, 2000 | Overall |  |  |  |  |  |  |  |  |  |  |  |
| **Radioligand monotherapy** | | | | | | | | | | | | |
| **PSMAfore**, 2024 |  |  | Overall | Overall |  |  |  |  |  |  |  |  |
| **Radiopharmaceutical monotherapy** | | | | | | | | | | | | |
| **ALSYMPCA**, 2013 | No Docetaxel  Subgroup |  |  |  | Docetaxel  Subgroup |  |  |  |  |  | Overall | Overall |
| **Radioligand therapy + Standard of Care** | | | | | | | | | | | | |
| **VISION**, 2021 |  |  |  |  |  |  | Overall | Overall |  |  |  |  |
| **TKI monotherapy** | | | | | | | | | | | | |
| **COMET-1**, 2016 |  |  |  |  |  |  | Overall | Overall |  |  |  |  |
| **SUN 1120**, 2013 |  |  |  |  | Overall | Overall |  |  |  |  |  |  |

Abbreviations: ADT: androgen deprivation therapy; ARPI: androgen-receptor pathway inhibitor; PI3K/AKTi: phosphatidylinositol 3-kinase and protein kinase B inhibitor; ASO: antisense oligonucleotide; DES: diethylstilbestrol diphosphate; ERA: endothelin receptor antagonist; IMiD: immunomodulatory drug; PDGFR: platelet-derived growth factor receptor; TKI: tyrosine kinase inhibitor; VEGFi: vascular endothelial growth factor inhibitor; VitD: vitamin d; GMCSF: granulocyte-macrophage colony-stimulating factor; PARPi: poly(ADP-ribose) polymerase inhibitor;

### **Supplement Table 13:** Outcomes reporting matrix for included phase III trials

| **Trial** | **Overall survival** | **Progression-free survival** | | | | **Radiographic Objective Response Rate** | **PSA 50% Response** |
| --- | --- | --- | --- | --- | --- | --- | --- |
|  |  | **rPFS** | **cPFS** | **TTP** | **TTF** |  |  |
| **Prior ADT only** | | | | | | | |
| **ARPI monotherapy** | | | | | | | |
| **COU-AA-302**, 2013 |  |  |  |  |  |  |  |
| **PREVAIL**, 2014 |  |  |  |  |  |  |  |
| **ELM-PC 4**, 2015 |  |  |  |  |  |  |  |
| **ARPI+ARPI** | | | | | | | |
| **ACIS**, 2021 |  |  |  |  |  |  |  |
| **AllianceA031201**, 2023 |  |  |  |  |  |  |  |
| **ARPI+PI3K/AKTi** | | | | | | | |
| **IPATential150**, 2021 |  |  |  |  |  |  |  |
| **ARPI+Radiopharmaceutical** | | | | | | | |
| **ERA 223**, 2019 |  |  |  |  |  |  |  |
| **Chemotherapy monotherapy** | | | | | | | |
| **CALGB9182**, 1999 |  |  |  |  |  |  |  |
| **FIRSTANA**, 2017 |  |  |  |  |  |  |  |
| **Kellokumpu-Lehtinen PL et al**, 2013 |  |  |  |  |  |  |  |
| **Berry W et al**, 2002 |  |  |  |  |  |  |  |
| **Abratt RP et al**, 2004 |  |  |  |  |  |  |  |
| **PRINCE**, 2018 |  |  |  |  |  |  |  |
| **TAX327**, 2004 |  |  |  |  |  |  |  |
| **Chemotherapy+ASO** | | | | | | | |
| **SYNERGY**, 2017 |  |  |  |  |  |  |  |
| **Chemotherapy+Chemotherapy** | | | | | | | |
| **SWOG-99-16**, 2004 |  |  |  |  |  |  |  |
| **Chemotherapy+DES** | | | | | | | |
| **ECOG 3882** 2003 |  |  |  |  |  |  |  |
| **Chemotherapy+ERA** | | | | | | | |
| **ENTHUSE (M1c)**, 2013 |  |  |  |  |  |  |  |
| **SWOG S0421**, 2013 |  |  |  |  |  |  |  |
| **Chemotherapy+IMiD** | | | | | | | |
| **MAINSAIL**, 2015 |  |  |  |  |  |  |  |
| **Chemotherapy+PDGFRi** | | | | | | | |
| **Mathew P et al**, 2007 |  |  |  |  |  |  |  |
| **Chemotherapy+TKI** | | | | | | | |
| **READY**, 2013 |  |  |  |  |  |  |  |
| **Chemotherapy+VEGFi** | | | | | | | |
| **CALGB 90401**, 2012 |  |  |  |  |  |  |  |
| **VENICE**, 2013 |  |  |  |  |  |  |  |
| **Chemotherapy+VitD-Analog** | | | | | | | |
| **ASCENT2**, 2011 |  |  |  |  |  |  |  |
| **ERA monotherapy** | | | | | | | |
| **ENTHUSE (PF)**, 2012 |  |  |  |  |  |  |  |
| **Carducci MA et al**, 2007 |  |  |  |  |  |  |  |
| **IMiD monotherapy** | | | | | | | |
| **Sternberg C et al**, 2016 |  |  |  |  |  |  |  |
| **Immunotherapy monotherapy** | | | | | | | |
| **CA184-095**, 2017 |  |  |  |  |  |  |  |
| **D9901**, 2006 |  |  |  |  |  |  |  |
| **D9902A**, 2009 |  |  |  |  |  |  |  |
| **Immunotherapy+GMCSF** | | | | | | | |
| **PROSPECT**, 2019 |  |  |  |  |  |  |  |
| **PARPi+ARPI** | | | | | | | |
| **PROpel**, 2023 |  |  |  |  |  |  |  |
| **TALAPRO-2**, 2023 |  |  |  |  |  |  |  |
| **Phenylurea monotherapy** | | | | | | | |
| **Small EJ et al**, 2000 |  |  |  |  |  |  |  |
| **Prior ADT+ARPI** | | | | | | | |
| **ARPI+ARPI** | | | | | | | |
| **PLATO**, 2018 |  |  |  |  |  |  |  |
| **ARPI+Immunotherapy** | | | | | | | |
| **IMbassador250**, 2022 |  |  |  |  |  |  |  |
| **Chemotherapy+ARPI** | | | | | | | |
| **PRESIDE**, 2022 |  |  |  |  |  |  |  |
| **Chemotherapy+Immunotherapy** | | | | | | | |
| **KEYNOTE-921**, 2025 |  |  |  |  |  |  |  |
| **PARPi monotherapy** | | | | | | | |
| **TRITON-3**, 2023 |  |  |  |  |  |  |  |
| **PROfound**, 2020 |  |  |  |  |  |  |  |
| **Radioligand monotherapy** | | | | | | | |
| **PSMAfore**, 2024 |  |  |  |  |  |  |  |
| **Prior ADT+Docetaxel** | | | | | | | |
| **ARPI monotherapy** | | | | | | | |
| **AFFIRM**, 2012 |  |  |  |  |  |  |  |
| **COU-AA-301**, 2011 |  |  |  |  |  |  |  |
| **ELM-PC 5**, 2015 |  |  |  |  |  |  |  |
| **SAKK 08/11**, 2016 |  |  |  |  |  |  |  |
| **Chemotherapy monotherapy** | | | | | | | |
| **PROSELICA**, 2017 |  |  |  |  |  |  |  |
| **TROPIC**, 2010 |  |  |  |  |  |  |  |
| **Chemotherapy+ASO** | | | | | | | |
| **AFFINITY**, 2017 |  |  |  |  |  |  |  |
| **TKI monotherapy** | | | | | | | |
| **SUN 1120**, 2013 |  |  |  |  |  |  |  |
| **Prior ADT+Docetaxel+ARPI** | | | | | | | |
| **Chemotherapy monotherapy** | | | | | | | |
| **CARD**, 2019 |  |  |  |  |  |  |  |
| **PARPi+Immunotherapy** | | | | | | | |
| **KEYLYNK-010**, 2023 |  |  |  |  |  |  |  |
| **Radioligand therapy+Standard of Care** | | | | | | | |
| **VISION**, 2021 |  |  |  |  |  |  |  |
| **TKI monotherapy** | | | | | | | |
| **COMET-1**, 2016 |  |  |  |  |  |  |  |
| **Heterogeneous Prior Therapy** | | | | | | | |
| **Chemotherapy+Immunotherapy** | | | | | | | |
| **VIABLE**, 2022 |  |  |  |  |  |  |  |
| **Multiple subgroups (ADT/ADT+ARPI/ADT+Docetaxel/ADT+ARPI+Docetaxel/Heterogeneous prior therapy)** | | | | | | | |
| **Chemotherapy monotherapy** | | | | | | | |
| **SPARC**, 2009 |  |  |  |  |  |  |  |
| **Immunotherapy monotherapy** | | | | | | | |
| **IMPACT**, 2010 |  |  |  |  |  |  |  |
| **PARPi+ARPI** | | | | | | | |
| **MAGNITUDE**, 2023 |  |  |  |  |  |  |  |
| **Radiopharmaceutical monotherapy** | | | | | | | |
| **ALSYMPCA**, 2013 |  |  |  |  |  |  |  |

Abbreviations: rPFS: radiographic progression free survival; cPFS: composite progression free survival; TTP; time to disease progression; ARPI: androgen-receptor pathway inhibitor; PI3K/AKTi: phosphatidylinositol 3-kinase and protein kinase B inhibitor; PDGFR: platelet-derived growth factor receptor; DES: diethylstilbestrol diphosphate;GMCSF: granulocyte-macrophage colony-stimulating factor; ASO: antisense oligonucleotide; ERA: endothelin receptor antagonist; IMiD: immunomodulatory drug; TKI: tyrosine kinase inhibitor; VitD: vitamin d; PARPi: poly(ADP-ribose) polymerase inhibitor;

Note: PLATO and CARD were phase IV trials. All remaining trials were phase III.

The color ‘green’ represents the outcome being reported in the respective study, ‘red’ represents lack of outcome reporting.

### **Supplement Table 14:** Outcomes reporting matrix for included phase II trials

| **Trial** | **Overall survival** | **Progression-free survival** | | | | **Radiographic Objective Response Rate** | **PSA 50% Response** |
| --- | --- | --- | --- | --- | --- | --- | --- |
|  |  | **rPFS** | **cPFS** | **TTP** | **TTF** |  |  |
| **Prior ADT only** | | | | | | | |
| **Antiandrogen+Chemotherapy** | | | | | | | |
| **Takahashi M et al**, 2013 |  |  |  |  |  |  |  |
| **Antifungal monotherapy** | | | | | | | |
| **Antonarakis ES et al**, 2013 |  |  |  |  |  |  |  |
| **Antifungal+Bisphosphonate** | | | | | | | |
| **Figg WD et al,** 2005 |  |  |  |  |  |  |  |
| **Antifungal+Chemotherapy** | | | | | | | |
| **Millikan R et al**, 2001 |  |  |  |  |  |  |  |
| **ARPI monotherapy** | | | | | | | |
| **TERRAIN**, 2016 |  |  |  |  |  |  |  |
| **ARPI+ARPI** | | | | | | | |
| **Khalaf DJ et al**, 2019 |  |  |  |  |  |  |  |
| **ARPI+Immunotherapy** | | | | | | | |
| **STAMP**, 2015 |  |  |  |  |  |  |  |
| **STRIDE**, 2023 |  |  |  |  |  |  |  |
| **ARPI+TKI** | | | | | | | |
| **Dorff TB et al**, 2019 |  |  |  |  |  |  |  |
| **ASO monotherapy** | | | | | | | |
| **Yu EY et al**, 2018 |  |  |  |  |  |  |  |
| **Tolcher AW et al**, 2002 |  |  |  |  |  |  |  |
| **Chemotherapy monotherapy** | | | | | | | |
| **Heidenreich A et al**, 2004 |  |  |  |  |  |  |  |
| **TIPC**, 2007 |  |  |  |  |  |  |  |
| **Krainer M et al**, 2007 |  |  |  |  |  |  |  |
| **Chemotherapy+Antifungal** | | | | | | | |
| **Millikan R et al**, 2003 |  |  |  |  |  |  |  |
| **Chemotherapy+ARPI** | | | | | | | |
| **CHEIRON**, 2021 |  |  |  |  |  |  |  |
| **Chemotherapy+ASO** | | | | | | | |
| **Chi KN et al**, 2010 |  |  |  |  |  |  |  |
| **Wiechno P et al**, 2014 |  |  |  |  |  |  |  |
| **Chemotherapy+Bcl-2 inhibitor** | | | | | | | |
| **EORTC**, 2009 |  |  |  |  |  |  |  |
| **Sonpavde G et al**, 2011 |  |  |  |  |  |  |  |
| **Chemotherapy+Chemotherapy** | | | | | | | |
| **Machiels JP et al**, 2008 |  |  |  |  |  |  |  |
| **Berry WR et al**, 2004 |  |  |  |  |  |  |  |
| **Albrecht W et al**, 2004 |  |  |  |  |  |  |  |
| **Cabrespine A et al**, 2006 |  |  |  |  |  |  |  |
| **Nelius T et al**, 2006 |  |  |  |  |  |  |  |
| **Oudard S et al**, 2005 |  |  |  |  |  |  |  |
| **Droz JP et al**, 2003 |  |  |  |  |  |  |  |
| **Galsky MD et al**, 2005 |  |  |  |  |  |  |  |
| **Eymard JC et al**, 2007 |  |  |  |  |  |  |  |
| **Chemotherapy+Curcuminoid** | | | | | | | |
| **Jahanmohan JP et al**, 2021 |  |  |  |  |  |  |  |
| **Chemotherapy+Immunotherapy** | | | | | | | |
| **Dahut WL et al**, 2004 |  |  |  |  |  |  |  |
| **ECOG 3899**, 2010 |  |  |  |  |  |  |  |
| **E1809**, 2015 |  |  |  |  |  |  |  |
| **de Bono et al**, 2014 |  |  |  |  |  |  |  |
| **Heidenreich A et al**, 2013 |  |  |  |  |  |  |  |
| **Chemotherapy+Radiopharmaceutical** | | | | | | | |
| **Taxium II**, 2017 |  |  |  |  |  |  |  |
| **Chemotherapy+TKI** | | | | | | | |
| **Horti J et al**, 2009 |  |  |  |  |  |  |  |
| **Chemotherapy+Vascular disrupting agent** | | | | | | | |
| **Pili R et al**, 2010 |  |  |  |  |  |  |  |
| **Chemotherapy+VitD-Analog** | | | | | | | |
| **Attia S et al**, 2008 |  |  |  |  |  |  |  |
| **ASCENT**, 2007 |  |  |  |  |  |  |  |
| **Corticosteroid monotherapy** | | | | | | | |
| **Venkitaraman R et al**, 2015 |  |  |  |  |  |  |  |
| **Corticosteroid+Immunotherapy** | | | | | | | |
| **Yoshimura K et al**, 2016 |  |  |  |  |  |  |  |
| **Corticosteroid+VEGFi** | | | | | | | |
| **Stadler WM et al**, 2004 |  |  |  |  |  |  |  |
| **ERA monotherapy** | | | | | | | |
| **James ND et al**, 2009 |  |  |  |  |  |  |  |
| **Carducci MA et al**, 2003 |  |  |  |  |  |  |  |
| **IMiD monotherapy** | | | | | | | |
| **Pili R et al**, 2011 |  |  |  |  |  |  |  |
| **Immunotherapy monotherapy** | | | | | | | |
| **PERSEUS**, 2016 |  |  |  |  |  |  |  |
| **Figg WD et al**, 2001 |  |  |  |  |  |  |  |
| **Immunotherapy+Chemotherapy** | | | | | | | |
| **Arlen PM et al**, 2006 |  |  |  |  |  |  |  |
| **Immunotherapy+GMCSF** | | | | | | | |
| **TBC-PRO-002**, 2010 |  |  |  |  |  |  |  |
| **Integrin inhibitor monotherapy** | | | | | | | |
| **Bradley DA et al**, 2011 |  |  |  |  |  |  |  |
| **Matrix Metalloproteinase Inhibitor monotherapy** | | | | | | | |
| **Lara Jr PN et al**, 2006 |  |  |  |  |  |  |  |
| **Progestin monotherapy** | | | | | | | |
| **Patel SR et al**, 1990 |  |  |  |  |  |  |  |
| **Somatostatin+Corticosteroid** | | | | | | | |
| **Dimopoulos MA et al**, 2004 |  |  |  |  |  |  |  |
| **TKI monotherapy** | | | | | | | |
| **Boccardo F et al**, 2008 |  |  |  |  |  |  |  |
| **TKI+1st gen AA** | | | | | | | |
| **Azad AA et al**, 2014 |  |  |  |  |  |  |  |
| **Sridhar SS et al**, 2015 |  |  |  |  |  |  |  |
| **Prior ADT+ARPI** | | | | | | | |
| **ARPI+TKI** | | | | | | | |
| **Spetsieris N et al**, 2021 |  |  |  |  |  |  |  |
| **Bipolar Androgen Therapy** | | | | | | | |
| **TRANSFORMER**, 2021 |  |  |  |  |  |  |  |
| **Chemotherapy+ARPI** | | | | | | | |
| **ABIDO SOGUG**, 2022 |  |  |  |  |  |  |  |
| **Chemotherapy+Chemotherapy** | | | | | | | |
| **MDACC Study**, 2019 |  |  |  |  |  |  |  |
| **Prior ADT+Docetaxel** | | | | | | | |
| **ASO+Chemotherapy** | | | | | | | |
| **P-06c**, 2011 |  |  |  |  |  |  |  |
| **Aurora kinase inhibitor monotherapy** | | | | | | | |
| **Meulenbeld HJ et al**, 2012 |  |  |  |  |  |  |  |
| **Chemotherapy monotherapy** | | | | | | | |
| **GETUG-P02**, 2015 |  |  |  |  |  |  |  |
| **CAINTA**, 2023 |  |  |  |  |  |  |  |
| **Rosenberg JE et al**, 2007 |  |  |  |  |  |  |  |
| **Chemotherapy+Chemotherapy** | | | | | | | |
| **RECARDO**, 2018 |  |  |  |  |  |  |  |
| **Chemotherapy+Immunotherapy** | | | | | | | |
| **Fizazi K et al**, 2012 |  |  |  |  |  |  |  |
| **Hakenberg OW et al**, 2019 |  |  |  |  |  |  |  |
| **Immunotherapy monotherapy** | | | | | | | |
| **Filaci G et al**, 2021 |  |  |  |  |  |  |  |
| **TKI monotherapy** | | | | | | | |
| **Droz JP et al**, 2014 |  |  |  |  |  |  |  |
| **Prior ADT+Docetaxel+ARPI** | | | | | | | |
| **ARPI monotherapy** | | | | | | | |
| **SAK-08-16**, 2023 |  |  |  |  |  |  |  |
| **Chemotherapy monotherapy** | | | | | | | |
| **ConCab**, 2018 |  |  |  |  |  |  |  |
| **Chemotherapy+AKTi** | | | | | | | |
| **RE-AKT**, 2024 |  |  |  |  |  |  |  |
| **Radioligand monotherapy** | | | | | | | |
| **TheraP**, 2021 |  |  |  |  |  |  |  |
| **Heterogeneous Prior Therapy** | | | | | | | |
| **ARPI+Radioligand** | | | | | | | |
| **ENZA-p**, 2024 |  |  |  |  |  |  |  |
| **Chemotherapy monotherapy** | | | | | | | |
| **TAXYNERGY**, 2017 |  |  |  |  |  |  |  |
| **Chemotherapy+Immunotherapy** | | | | | | | |
| **IND 209**, 2018 |  |  |  |  |  |  |  |
| **Kongsted P et al**, 2017 |  |  |  |  |  |  |  |
| **PARPi+ARPi** | | | | | | | |
| **BRCAAway**, 2024 |  |  |  |  |  |  |  |
| **TKI monotherapy** | | | | | | | |
| **Monk P et al**, 2018 |  |  |  |  |  |  |  |
| **Smith DC et al**, 2013 |  |  |  |  |  |  |  |
| **Multiple subgroups (ADT/ADT+ARPI/ADT+Docetaxel/ADT+ARPI+Docetaxel/Heterogeneous prior therapy)** | | | | | | | |
| **Chemotherapy monotherapy** | | | | | | | |
| **Annala M et al**, 2021 |  |  |  |  |  |  |  |
| **Chemotherapy+AKTi** | | | | | | | |
| **ProCAID**, 2021 |  |  |  |  |  |  |  |

Abbreviations: rPFS: radiographic progression free survival; cPFS: composite progression free survival; TTP; time to disease progression; PSA: prostate-specific antigen; ARPI: androgen-receptor pathway inhibitor; TKI: tyrosine kinase inhibitor; ASO: antisense oligonucleotide; BAT: bipolar androgen therapy; AKTi: protein kinase B inhibitor; Bcl-2: B-cell lymphoma 2 protein; VitD: vitamin d; VEGFi: vascular endothelial growth factor inhibitor; ERA: endothelin receptor antagonist; IMiD: immunomodulatory drug; GMCSF: granulocyte-macrophage colony-stimulating factor; PARPi: poly(ADP-ribose) polymerase inhibitor; AA: antiandrogen; SOC: standard of care

The color ‘green’ represents the outcome being reported in the respective study, ‘red’ represents lack of outcome reporting.

### **Supplement Table 15:** Overall survival subgroups reported across all included phase II and phase III trials

| **Subgroup Reported** | **Phase III** | **Phase II** | **Total** |
| --- | --- | --- | --- |
| Age | 24 | 3 | 27 |
| ECOG-PS | 23 | 4 | 27 |
| Baseline LDH, U/L | 17 | 4 | 21 |
| Baseline PSA , ng/ml | 14 | 4 | 18 |
| Geographic Area | 16 | 0 | 16 |
| Baseline ALP | 10 | 4 | 14 |
| Visceral disease at entry | 12 | 2 | 14 |
| Race | 9 | 1 | 10 |
| Gleason Score | 7 | 2 | 9 |
| Baseline BPI level | 8 | 0 | 8 |
| Measurable Disease | 8 | 0 | 8 |
| Bisphosphonates | 6 | 1 | 7 |
| Hemoglobin | 4 | 3 | 7 |
| Bone metastases | 7 | 0 | 7 |
| No. of bone lesions | 5 | 1 | 6 |
| Type of Disease progression | 6 | 0 | 6 |
| Bone-only Metastasis | 4 | 1 | 5 |
| Pain at baseline | 5 | 0 | 5 |
| Prior Docetaxel Use | 4 | 1 | 5 |
| Number of previous chemotherapies | 4 | 0 | 4 |
| Liver Metastasis | 4 | 0 | 4 |
| Prior NHT treatment | 3 | 1 | 4 |
| Prior Opioid use | 3 | 0 | 3 |
| Baseline Albumin | 1 | 1 | 2 |
| BRCA mutational status | 2 | 0 | 2 |
| HLA-A2 | 1 | 1 | 2 |
| Node only at baseline | 2 | 0 | 2 |
| Previous taxane-containing regimen | 2 | 0 | 2 |
| Prior radical prostatectomy | 1 | 1 | 2 |
| Prior radiotherapy | 1 | 1 | 2 |
| Soft tissue metastases | 1 | 1 | 2 |
| Tumor progression at baseline | 2 | 0 | 2 |
| PD-L1 status | 2 | 0 | 2 |
| 1R gene alteration status | 1 | 0 | 1 |
| AR amplification | 0 | 1 | 1 |
| ARPi part of planned standard care | 1 | 0 | 1 |
| Average pain score | 1 | 0 | 1 |
| BAP | 1 | 0 | 1 |
| CRP | 0 | 1 | 1 |
| ctDNA fraction | 0 | 1 | 1 |
| DNA repair defect | 0 | 1 | 1 |
| Docetaxel exposure time | 1 | 0 | 1 |
| Duration of Prior Abiraterone | 0 | 1 | 1 |
| Ethnicity | 0 | 1 | 1 |
| Extent of disease | 1 | 0 | 1 |
| FACT-P | 1 | 0 | 1 |
| Germline status | 1 | 0 | 1 |
| Halabi median (m) | 0 | 1 | 1 |
| Karnofsky status | 1 | 0 | 1 |
| Neutrophil to Leukocyte Ratio | 1 | 0 | 1 |
| New lesion at baseline | 1 | 0 | 1 |
| No. of bone scan > 0 | 0 | 1 | 1 |
| Nodal metastasis | 1 | 0 | 1 |
| Bone and non-bone | 0 | 1 | 1 |
| Non-bone only | 0 | 1 | 1 |
| Number of previous hormonal treatments | 1 | 0 | 1 |
| Patients with Baseline Corticosteroid Use | 1 | 0 | 1 |
| PD-L1 status at 1% cutoff | 1 | 0 | 1 |
| PD-L1 status at 5% cutoff | 1 | 0 | 1 |
| PI3K defect | 0 | 1 | 1 |
| Post baseline abiraterone | 1 | 0 | 1 |
| Postbaseline docetaxel | 1 | 0 | 1 |
| Present Pain Intensity Score | 1 | 0 | 1 |
| Previous docetaxel usage | 1 | 0 | 1 |
| Previous primary therapy | 1 | 0 | 1 |
| Primary Gleason Grade | 1 | 0 | 1 |
| Prior Cabazitaxel | 1 | 0 | 1 |
| Prior Castration | 1 | 0 | 1 |
| Prior Chemotherapy use | 1 | 0 | 1 |
| Prior Complete Androgen blockade | 1 | 0 | 1 |
| Prior orchiectomy | 1 | 0 | 1 |
| Prior radical prostatectomy or radiotherapy | 1 | 0 | 1 |
| Progression <3 months after docetaxel | 1 | 0 | 1 |
| Progression ≥3 months after docetaxel | 1 | 0 | 1 |
| Progression during docetaxel treatment | 1 | 0 | 1 |
| Prostatic acid phosphatase | 1 | 0 | 1 |
| PSA Doubling Time (months) | 0 | 1 | 1 |
| Reason for discontinuation of docetaxel | 1 | 0 | 1 |
| Rising PSA at baseline | 1 | 0 | 1 |
| sRAGE | 0 | 1 | 1 |
| Stage | 1 | 0 | 1 |
| Testosterone | 1 | 0 | 1 |
| TGFb1 | 0 | 1 | 1 |
| Time from diagnosis | 1 | 0 | 1 |
| Total Docetaxel Dose | 1 | 0 | 1 |
| Total nontarget lesions regroup | 0 | 1 | 1 |
| Total target lesions regroup | 0 | 1 | 1 |
| TP53 defect | 0 | 1 | 1 |
| Treatment of abiraterone acetate plus prednisone started | 1 | 0 | 1 |
| TSP1 | 0 | 1 | 1 |
| Urinary N-telopeptide level | 1 | 0 | 1 |
| Use of G-CSF | 1 | 0 | 1 |
| VEGF-A | 0 | 1 | 1 |
| VEGF-C | 0 | 1 | 1 |
| Weight | 1 | 0 | 1 |

Abbreviations: ECOG-PS: eastern cooperative oncology group performance status; LDH: lactate dehydrogenase; PSA: prostate specific antigen; ALP: alkaline phosphatase; BPI: brief pain inventory; AR: androgen receptor; ARPI: androgen receptor pathway inhibitor; BAP: bone alkaline phosphatase; CRP: c-reactive protein; sRAGE : soluble receptor for advanced glycation end products; TGFb: transforming growth factor beta; G-CSF: granulocyte colony stimulating factor; TSP1: thrombospondin-1; VEGF: vascular endothelial growth factor

### **Supplement Table 16:** Progression free survival subgroups reported across all included phase II and phase III trials

| **Subgroup Reported** | **Phase III** | **Phase II** | **Total** |
| --- | --- | --- | --- |
| Age | 18 | 4 | 22 |
| ECOG-PS | 16 | 4 | 20 |
| Baseline PSA, ng/mL | 13 | 5 | 18 |
| Baseline LDH, U/L | 10 | 4 | 14 |
| Geographic Area | 12 | 2 | 14 |
| Visceral disease | 9 | 3 | 11 |
| Gleason Score | 7 | 3 | 10 |
| Baseline ALP | 5 | 4 | 9 |
| Bone metastasis at baseline | 6 | 3 | 9 |
| Disease Location | 6 | 3 | 9 |
| Bone-only Metastasis | 5 | 3 | 8 |
| Race | 8 | 0 | 8 |
| Disease progression | 7 | 0 | 7 |
| Prior ARPI treatment | 4 | 1 | 5 |
| Baseline BPI level | 4 | 0 | 4 |
| Hemoglobin | 4 | 0 | 4 |
| Measurable Disease | 4 | 0 | 4 |
| BRCA mutational status | 3 | 0 | 3 |
| No. of bone lesions at baseline | 3 | 0 | 3 |
| Previous taxane-based therapy | 3 | 0 | 3 |
| Liver Metastasis | 3 | 0 | 3 |
| Baseline Opioid use | 0 | 2 | 2 |
| HRR gene alteration status | 2 | 0 | 2 |
| Karnofsky status | 1 | 1 | 2 |
| Prior Docetaxel Use | 1 | 1 | 2 |
| PD-L1 status | 2 | 0 | 2 |
| Albumin at baseline | 0 | 1 | 1 |
| AR amplification | 0 | 1 | 1 |
| ARPi part of planned standard care | 1 | 0 | 1 |
| Baseline bisphosphonate or denosumab use | 0 | 1 | 1 |
| Baseline Visual Analog Score | 1 | 0 | 1 |
| ctDNA fraction | 0 | 1 | 1 |
| DNA repair defect | 0 | 1 | 1 |
| Duration of first ADT | 1 | 0 | 1 |
| Gene | 1 | 0 | 1 |
| Germline status | 1 | 0 | 1 |
| LHRHa or orchiectomy | 0 | 1 | 1 |
| M1 disease at diagnosis | 1 | 0 | 1 |
| Neutrophil count | 1 | 0 | 1 |
| Neutrophil to lymphocyte ratio | 1 | 0 | 1 |
| Number of previous chemotherapies | 1 | 0 | 1 |
| Patients with Baseline Corticosteroid Use | 1 | 0 | 1 |
| PI3K defect | 0 | 1 | 1 |
| Present Pain Intensity Score (PPI) | 1 | 0 | 1 |
| Previous antiandrogen therapy | 0 | 1 | 1 |
| Previous ARPI setting | 1 | 0 | 1 |
| Previous bisphosphonate use | 1 | 0 | 1 |
| Previous primary therapy | 1 | 0 | 1 |
| Previous therapy with curative intent for localized disease | 1 | 0 | 1 |
| Prior Prostatectomy | 0 | 1 | 1 |
| Prior Radiotherapy | 0 | 1 | 1 |
| Prior Taxane or NHT | 1 | 0 | 1 |
| PSA Doubling Time (months) | 0 | 1 | 1 |
| PTEN loss by immunohistochemistry | 1 | 0 | 1 |
| PTEN loss by next-generation sequencing | 1 | 0 | 1 |
| Stage at Diagnosis | 1 | 0 | 1 |
| Stratification factor | 0 | 1 | 1 |
| Symptomatology | 1 | 0 | 1 |
| Testosterone | 1 | 0 | 1 |
| Time from initiation of Androgen-signaling targeted inhibitor to progression | 1 | 0 | 1 |
| Timing of Androgen-signaling targeted inhibitor | 1 | 0 | 1 |
| TP53 defect | 0 | 1 | 1 |
| Tumor progression | 1 | 0 | 1 |

Abbreviations: ECOG-PS: eastern cooperative oncology group performance status; LDH: lactate dehydrogenase; PSA: prostate specific antigen; ALP: alkaline phosphatase; BPI: brief pain inventory; AR: androgen receptor; ARPI: androgen receptor pathway inhibitor; PI3K: phosphoinositide 3-kinase; LHRHa: luteinizing hormone-releasing hormone agonist

### **Supplement Table 17:** Overall survival and progression free survival reported in included phase III trials

| **Trial** | **Arm** | **Overall Survival (OS)** | | **Progression Free Survival (PFS)** | |
| --- | --- | --- | --- | --- | --- |
|  |  | **Median OS (months)** | **HR (95% CI)** | **Median PFS (months)** | **HR (95% CI)** |
| **ARPI monotherapy** | | | | | |
| **AFFIRM**, 2012 | Rx: Enzalutamide Ctrl: Placebo | 18.4 vs. 13.6 | 0.63 (0.53-0.75) * | 8.3 vs. 2.9 ^†^ | 0.40 (0.35-0.47) |
| **COU-AA-301**, 2011 | Rx: Abiraterone Ctrl: Placebo | 15.8 vs. 11.2 | 0.74 (0.64-0.86) * | 5.6 vs. 3.6 ^†^ | 0.66 (0.58-0.76) |
| **COU-AA-302**, 2013 | Rx: Abiraterone Ctrl: Placebo | 34.7 vs. 30.3 | 0.81 (0.70-0.93) * | 16.5 vs. 8.2 ^†^ | 0.52 (0.45-0.61) |
| **PREVAIL**, 2014 | Rx: Enzalutamide Ctrl: Placebo | 35.3 vs. 31.3 | 0.77 (0.67-0.88) * | 20 vs 5.4 ^†^ | 0.32 (0.28-0.36) |
| **ELM-PC 4**, 2015 | Rx: TAK-700 Ctrl: Placebo | 31.4 vs. 29.5 | 0.92 (0.79-1.08) | 13.8 vs. 8.7 ^†^ | 0.71 (0.63-0.80) |
| **ELM-PC 5**, 2015 | Rx: TAK-700 Ctrl: Placebo | 17 vs. 15.2 | 0.89 (0.74-1.06) | 8.3 vs. 5.7 ^†^ | 0.76 (0.65-0.89) |
| **SAKK 08/11**, 2016 | Rx: TAK-700 Ctrl: Placebo | Not reported | | 8.5 vs. 2.8 ^†^ | 0.42 (0.20-0.91) |
| **ARPI+ARPI** | | | | | |
| **ACIS**, 2021 | Rx: Abiraterone + Apalutamide Ctrl: Abiraterone | 36.2 vs. 33.7 | 0.95 (0.81-1.11) | 22 vs. 19.2 ^†^ | 0.86 (0.72-1.04) |
| **AllianceA031201**, 2023 | Rx: Enzalutamide + Abiraterone Ctrl: Enzalutamide | 34.2 vs. 32.7 | 0.89 (0.78-1.01) | 24.3 vs. 21.3 ^†^ | 0.86 (0.76-0.97) |
| **PLATO**, 2018 | Rx: Enzalutamide + Abiraterone Ctrl: Abiraterone | Not reported | | 5.7 vs. 5.6 ^‡^ | 0.83 (0.61-1.12) |
| **ARPI+Immunotherapy** | | | | | |
| **IMbassador250**, 2022 | Rx: Enzalutamide + Atezolizumab Ctrl: Enzalutamide | 15.2 vs. 16.6 | 1.12 (0.91-1.37) | 4.2 vs. 4.1 ^†^ | 0.90 (0.75-1.07) |
| **ARPI+PI3K/AKTi** | | | | | |
| **IPATential150**, 2021 | Rx: Abiraterone + Ipatasertib Ctrl: Abiraterone | NR vs. NR | 0.93 (0.73-1.18) | 19.2 vs. 16.6 ^†^ | 0.84 (0.71-0.99) |
| **ARPI+Radiopharmaceutical** | | | | | |
| **ERA 223**, 2019 | Rx: Abiraterone + Radium-223 Ctrl: Abiraterone | 30.7 vs. 33.3 | 1.19 (0.95-1.50) | 11.2 vs.12.4 ^†^ | 1.15 (0.96-1.38) |
| **Chemotherapy monotherapy** | | | | | |
| **CALGB9182**, 1999 | Rx: Mitoxantrone + Hydrocortisone Ctrl: Hydrocortisone | 12.3 vs. 12.6 | NA | 3.7 vs. 2.3 ^§^ | NA |
| **FIRSTANA**, 2017 | Rx1: Cabazitaxel20 Rx2: Cabazitaxel25 Ctrl: Docetaxel | Rx1 vs. Ctrl:  24.5 vs. 24.3 Rx2 vs. Ctrl:  25.2 vs. 24.3 | Rx1 vs. Ctrl:  1.01 (0.85-1.20) Rx2 vs. Ctrl:  0.97 (0.82-1.16) | Rx1 vs. Ctrl:  13.4 vs. 12.1 ^†^ Rx2 vs. Ctrl:   13.1 vs. 12.1 ^†^ | Rx1 vs. Ctrl:  0.92 (0.75-1.12) Rx2 vs. Ctrl:  0.96 (0.79-1.17) |
| **Kellokumpu-Lehtinen PL et al**, 2013 | Rx: Docetaxel (2W) Ctrl: Docetaxel | 19.5 vs. 17 | 1.40 (1.10-1.80) | 15.8 vs. 14.6 ^§^ | 1.30 (1.00-1.60) |
| **Berry W et al**, 2002 | Rx: Mitoxantrone Ctrl: Placebo | 23 vs. 19 | NA | Not reported | |
| **Abratt RP et al**, 2004 | Rx: Vinorelbine + Hydrocortisone Ctrl: Hydrocortisone | 14.7 vs. 15.2 | NA | 3.7 vs. 2.8 ^‡^ | NA |
| **PRINCE**, 2018 | Rx: Docetaxel (int) Ctrl: Docetaxel | 18.3 vs. 19.3 | 1.14 (0.75-1.72) | 10 vs. 5.4 ^‡^ | 0.69 (0.43-1.05) |
| **PROSELICA**, 2017 | Rx: Cabazitaxel20 Ctrl: Cabazitaxel25 | 13.4 vs. 14.5 | 1.02 (0.92-1.18) | 2.9 vs 3.5 ^‡^ | 1.10 (0.97-1.24) |
| **SPARC**, 2009 | Rx: Satraplatin Ctrl: Placebo | 14.1 vs. 14.1 | 0.98 (0.84-1.15) | 2.6 vs. 2.2 ^‡^ | 0.67 (0.57-0.77) |
| **TAX327**, 2004 | Rx1: Docetaxel Rx2: Docetaxel30 (W) Ctrl: Mitoxantrone | Rx1 vs. Ctrl: 19.2 vs. 16.3 Rx2 vs. Ctrl: 17.8 vs. 16.3 | Rx1 vs. Ctrl: 0.79 (0.67-0.93) * Rx2 vs. Ctrl: 0.87 (0.74-1.02) | Not reported | |
| **TROPIC**, 2010 | Rx: Cabazitaxel25 Ctrl: Mitoxantrone | 15.1 vs. 12.7 | 0.70 (0.59-0.83) * | 2.8 vs. 1.4 ^‡^ | 0.74 (0.64-0.86) |
| **CARD**, 2019 | Rx: Cabazitaxel25 Ctrl: Enzalutamide /Abiraterone | 13.6 vs. 11 | 0.64 (0.46-0.89) * | 8 vs. 3.7 ^†^ | 0.54 (0.40-0.73) |
| **Chemotherapy+ARPI** | | | | | |
| **PRESIDE**, 2022 | Rx: Docetaxel  + Enzalutamide Ctrl: Docetaxel | Not reported | | 9.5 vs. 8.3 ^‡^ | 0.72 (0.53-0.96) |
| **Chemotherapy+ASO** | | | | | |
| **AFFINITY**, 2017 | Rx: Cabazitaxel25  + Custirsen Ctrl: Cabazitaxel25 | 14.1 vs. 13.4 | 0.95 (0.80-1.12) | Not reported | |
| **SYNERGY**, 2017 | Rx: Docetaxel + Custirsen Ctrl: Docetaxel | 23.4 vs. 22 | 0.93 (0.79-1.10) | Not reported | |
| **Chemotherapy+Chemotherapy** | | | | | |
| **SWOG-99-16**, 2004 | Rx: Docetaxel60 + Estramustine Ctrl: Mitoxantrone | 17.5 vs. 15.6 | 0.80 (0.67-0.97) * | 6.3 vs. 3.2 ^‡^ | NA |
| **Chemotherapy+DES** | | | | | |
| **ECOG 3882** 2003 | Rx: Doxorubicin + DES Ctrl: Doxorubicin | 8.5 vs. 7.7 | NA | Not reported | |
| **Chemotherapy+ERA** | | | | | |
| **ENTHUSE (M1c)**, 2013 | Rx: Docetaxel + Zibotentan Ctrl: Docetaxel | 20 vs. 19.2 | 1.00 (0.84-1.18) | NA | 1.00 (0.87-1.14) ^‡^ |
| **SWOG S0421**, 2013 | Rx: Docetaxel + Atrasentan Ctrl: Docetaxel | 17.8 vs. 17.6 | 1.04 (0.90-1.19) | 9.2 vs. 9.1 ^†^ | 1.02 (0.89-1.16) |
| **Chemotherapy+IMiD** | | | | | |
| **MAINSAIL**, 2015 | Rx: Docetaxel + Lenalidomide Ctrl: Docetaxel | 17.7 vs. NR | 1.53 (1.17-2.00) | 10.4 vs. 10.6 ^†^ | 1.32 (1.05-1.66) |
| **Chemotherapy+Immunotherapy** | | | | | |
| **VIABLE**, 2022 | Rx: Docetaxel + DCVAC Ctrl: Docetaxel | 23.9 vs. 24.3 | 1.04 (0.90-1.21) | 11.1 vs. 11.1 ^†^ | 0.99 (0.86-1.14) |
| **KEYNOTE-921**, 2025 | Rx: Docetaxel  + Pembrolizumab Ctrl: Docetaxel | 19.6 vs. 19 | 0.92 (0.78-1.09) | 8.6 vs. 8.3 ^†^ | 0.85 (0.71-1.01) |
| **Chemotherapy+PDGFRi** | | | | | |
| **Mathew P et al**, 2007 | Rx: Docetaxel30 (W) + Imatinib Ctrl: Docetaxel30 (W) | 20.9 vs. NR | 1.67 (0.77-3.64) | 4.2 vs. 4.2 ^‡^ | NA |
| **Chemotherapy+TKI** | | | | | |
| **READY**, 2013 | Rx: Docetaxel + Dasatinib Ctrl: Docetaxel | 21.5 vs. 21.2 | 0.99 (0.87-1.13) | 11.8 vs. 11.1 ^‡^ | 0.92 (0.82-1.05) |
| **Chemotherapy+VEGFi** | | | | | |
| **CALGB 90401**, 2012 | Rx: Docetaxel + Bevacizumab Ctrl: Docetaxel | 22.6 vs. 21.5 | 0.91 (0.70-1.05) | 9.9 vs. 7.5 ^†^ | 0.80 (0.71-0.91) |
| **VENICE**, 2013 | Rx: Docetaxel + Aflibercept Ctrl: Docetaxel | 22.1 vs. 21.2 | 0.94 (0.82-1.08) | 6.9 vs. 6.2 ^‡^ | NA |
| **Chemotherapy+VitD-Analog** | | | | | |
| **ASCENT2**, 2011 | Rx: Docetaxel + Calcitriol Ctrl: Docetaxel | 17.8 vs. 20.2 | NA | Not reported | |
| **ERA monotherapy** | | | | | |
| **ENTHUSE (PF)**, 2012 | Rx: Zibotentan Ctrl: Placebo | 24.5 vs. 22.5 | 0.87 (0.69-1.10) | 6.2 vs. 6.5 ^‡^ | 1.01 (0.85-1.21) |
| **Carducci MA et al**, 2007 | Rx: Atrasentan Ctrl: Placebo | 20.5 vs. 20.3 | 0.97 (0.81-1.17) | NA | 0.89 (0.76-1.04) ^§^ |
| **IMiD monotherapy** | | | | | |
| **Sternberg C et al**, 2016 | Rx: Tasquinimod Ctrl: Placebo | 21.3 vs. 24 | 1.01 (0.94-1.28) | 7.0 vs. 4.4 ^†^ | 0.64 (0.54-0.75) |
| **Immunotherapy monotherapy** | | | | | |
| **CA184-095**, 2017 | Rx: Ipilimumab Ctrl: Placebo | 28.7 vs. 29.7 | 1.11 (0.88-1.39) | 5.6 vs. 3.8 ^‡^ | 0.67 (0.55-0.81) |
| **D9901**, 2006 | Rx: Sipuleucel-T  Ctrl: Placebo | 25.9 vs. 21.4 | 0.58 (0.39-0.88) * | 11.7 vs. 9.1 ^§^ | 0.69 (0.47-1.01) |
| **IMPACT**, 2010 | Rx: Sipuleucel-T  Ctrl: Placebo | 25.8 vs. 21.7 | 0.78 (0.61-0.98) * | 3.7 vs. 3.6 ^§^ | 0.95 (0.77-1.17) |
| **D9902A**, 2009 | Rx: Sipuleucel-T  Ctrl: Placebo | 19 vs. 15.7 | 0.79 (0.48-1.28) | 10.9 vs. 9.9 ^§^ | 0.92 (0.59-1.45) |
| **Immunotherapy+GMCSF** | | | | | |
| **PROSPECT**, 2019 | Rx1: PVAC + GMCSF Rx2: PVAC Ctrl: Placebo | Rx1 vs. Ctrl: 33.2 vs. 34.3 Rx2 vs. Ctrl: 34.4 vs. 34.3 | Rx1 vs. Ctrl: 1.02 (0.86-1.22) Rx2 vs. Ctrl: 1.01 (0.84-1.20) | Not reported | |
| **PARPi monotherapy** | | | | | |
| **PROfound**, 2020 | Rx: Olaparib Ctrl: Enzalutamide /Abiraterone | 17.3 vs. 14 | 0.79 (0.61-1.03) | 5.8 vs. 3.5 ^†^ | 0.49 (0.38-0.63) |
| **TRITON-3**, 2023 | Rx: Rucaparib Ctrl: Enzalutamide /Abiraterone /Docetaxel | 23.6 vs. 20.9 | 0.94 (0.72-1.23) | 10.2 vs. 6.4 ^†^ | 0.61 (0.47-0.80) |
| **PARPi+ARPI** | | | | | |
| **MAGNITUDE**, 2023 | Rx: Niraparib + Abiraterone Ctrl: Abiraterone | NA | 0.70 (0.49-0.99) * | 16.7 vs. 13.7 ^†^ | 0.76 (0.60-0.97) |
| **PROpel**, 2023 | Rx: Olaparib + Abiraterone Ctrl: Abiraterone | 42.1 vs. 34.7 | 0.81 (0.67-1.00) | 27.6 vs. 16.4 ^†^ | 0.61 (0.49-0.74) |
| **TALAPRO-2**, 2023 | Rx: Talazoparib + Enzalutamide Ctrl: Enzalutamide | 45.8 vs. 37 | 0.80 (0.66-0.96) * | 33.1 vs. 19.5 ^†^ | 0.67 (0.55-0.81) |
| **PARPi+Immunotherapy** | | | | | |
| **KEYLYNK-010**, 2023 | Rx: Olaparib + Pembrolizumab Ctrl: Enzalutamide /Abiraterone | 15.8 vs. 14.6 | 0.94 (0.77-1.14) | 4.6 vs. 4.2 ^†^ | 0.96 (0.79-1.16) |
| **Phenylurea monotherapy** | | | | | |
| **Small EJ et al**, 2000 | Rx: Suramin Ctrl: Placebo | 9.4 vs. 9.2 | NA | Not reported | |
| **Radioligand monotherapy** | | | | | |
| **PSMAfore**, 2024 | Rx: 177Lu Ctrl: Enzalutamide /Abiraterone | 23.7 vs. 23.9 | 0.98 (0.75-1.28) | 11.6 vs. 5.6 ^†^ | 0.49 (0.39-0.61) |
| **Radioligand therapy+Standard of Care** | | | | | |
| **VISION**, 2021 | Rx: 177Lu + SOC Ctrl: SOC | 15.3 vs. 11.3 | 0.62 (0.52-0.74) * | 8.7 vs. 3.4 ^†^ | 0.40 (0.29-0.57) |
| **Radiopharmaceutical monotherapy** | | | | | |
| **ALSYMPCA**, 2013 | Rx: Radium-223 Ctrl: Placebo | 14.9 vs. 11.3 | 0.70 (0.58-0.83) * | Not reported | |
| **TKI monotherapy** | | | | | |
| **COMET-1**, 2016 | Rx: Cabozantinib Ctrl: Prednisone | 11 vs. 9.8 | 0.90 (0.76-1.06) | 5.6 vs. 2.8 ^†^ | 0.48 (0.40-0.57) |
| **SUN 1120**, 2013 | Rx: Sunitinib Placebo | 13.1 vs. 11.8 | 0.91 (0.76-1.10) | 5.6 vs. 4.1 ^†^ | 0.73 (0.59-0.89) |

Abbreviations: ARPI: androgen-receptor pathway inhibitor; PI3K/AKTi: phosphatidylinositol 3-kinase and protein kinase B inhibitor; PDGFR: platelet-derived growth factor receptor; DES: diethylstilbestrol diphosphate; PVAC: PROSTVAC (viral vector-based immunotherapy); GMCSF: granulocyte-macrophage colony-stimulating factor; ASO: antisense oligonucleotide; ERA: endothelin receptor antagonist; IMiD: immunomodulatory drug; TKI: tyrosine kinase inhibitor; VitD: vitamin d; PARPi: poly(ADP-ribose) polymerase inhibitor; 177Lu: 177Lu-PSMA-617; SOC: standard of care

Note: PLATO and CARD were phase IV trials. All remaining trials were phase III. The color ‘green’ represents a positive trial, ‘red’ represents a negative trial.

* Statistically significant overall survival benefit was observed

^†^ Radiographic progression-free survival

^‡^ Composite progression-free survival

^§^ Time to disease progression

Cabazitaxel20: Cabazitaxel 20 mg/m^2^ IV on day 1 of every 3-week cycle

Cabazitaxel25: Cabazitaxel 25 mg/m^2^ IV on day 1 of every 3-week cycle

Docetaxel (2W): Docetaxel 75 mg/m^2^ IV on days 1 and 15 of a 4-week cycle

Docetaxel (int): Docetaxel 35 mg/m^2^ IV on days 1, 8, 15, repeat cycle at day 29

Docetaxel30 (W): Docetaxel 30 mg/m^2^ IV on days 1, 8, 15, 22 and 29 of a 6-week cycle

Remaining drugs were administered at standard doses.

### **Supplement Table 18:** Overall survival and progression free survival reported in included phase II trials

| **Trial** | **Arm** | **Overall Survival (OS)** | | **Progression Free Survival (PFS)** | |
| --- | --- | --- | --- | --- | --- |
|  |  | **Median OS (months)** | **HR (95% CI)** | **Median PFS (months)** | **HR (95% CI)** |
| **Antiandrogen+Chemotherapy** | | | | | |
| **Takahashi M et al**, 2013 | Rx: Antiandrogen + Tegafur-Uracil Ctrl: Antiandrogen | Not reported | | Not reported | |
| **Antifungal monotherapy** | | | | | |
| **Antonarakis ES et al**, 2013 | Rx: Itraconazole600 Ctrl: Itraconazole200 | Not reported | | 8.3 vs. 2.7 * | NA |
| **Antifungal+Bisphosphonate** | | | | | |
| **Figg WD et al,**  2005 | Rx: Ketoconazole + Alendronate Ctrl: Ketoconazole | 19 vs. NR | NA | 4.6 vs. 3.8 * | NA |
| **Antifungal+Chemotherapy** | | | | | |
| **Millikan R et al**, 2001 | Rx1: Ketoconazole + Doxorubicin Rx2: Ketoconazole | 12.5 vs. 12.5 | NA | Not reported | |
| **ARPI monotherapy** | | | | | |
| **TERRAIN**, 2016 | Rx: Enzalutamide Ctrl: Bicalutamide | Not reported | | 15.7 vs. 5.8 * | 0.44 (0.34-0.57) |
| **SAK-08-16**, 2023 | Rx: Darolutamide Ctrl: Placebo | 24 vs. 21.3 | 0.62 (0.30-1.26) | 5.5 vs. 4.5 ^†^ | 0.54 (0.32-0.91) |
| **ARPI+ARPI** | | | | | |
| **Khalaf DJ et al**, 2019 | Rx1: Abiraterone followed by Enzalutamide Rx2: Enzalutamide followed by Abiraterone | 28.8 vs. 24.7 | 0.79 (0.54-1.16) | 7.9 vs. 7.3 ^‡^ | 0.95 (0.70-1.29) |
| **ARPI+Immunotherapy** | | | | | |
| **STAMP**, 2015 | Rx1: Abiraterone + cSipuleucel-T Rx2: Abiraterone + sSipuleucel-T | 30 vs. 34.2 | 1.0 (0.51-1.98) | Not reported | |
| **STRIDE**, 2023 | Rx1: Enzalutamide + cSipuleucel-T  Rx2: Enzalutamide + sSipuleucel-T | 34.7 vs. 32.5 | 1.41 (0.68-2.92) | Not reported | |
| **ARPI+Radioligand** | | | | | |
| **ENZA-p**, 2024 | Rx: Enzalutamide + 177Lu Ctrl: Enzalutamide | 34 vs. 26 | 0.55 (0.36-0.84) ^\|\|^ | NA | 0.61 (0.42-0.87) ^†^ |
| **ARPI+TKI** | | | | | |
| **Dorff TB et al**, 2019 | Rx: Abiraterone  + Dasatinib Ctrl: Abiraterone | 41.2 vs. 26.9 | NA | 15.7 vs. 9 ^†^ | NA |
| **Spetsieris N et al**, 2021 | Rx: Abiraterone + Sunitinib Ctrl: Abiraterone + Dasatinib | 22.9 vs. 20.8 | 1.04 (0.72-1.49) | 5.5 vs. 5.7 ^§^ | 0.85 (0.59-1.22) |
| **ASO monotherapy** | | | | | |
| **Yu EY et al**, 2018 | Rx: Apatorsen Ctrl: Prednisone | Not reported | | 17.9 vs. 13.9 | NA |
| **Tolcher AW et al,**  2002 | Rx: ISIS 3521 Ctrl: ISIS 5132 | Not reported | | Not reported | |
| **ASO+Chemotherapy** | | | | | |
| **P-06c,**  2011 | Rx1: Custirsen + Docetaxel Rx2: Custirsen  + Mitoxantrone | 15.8 vs. 11.5 | NA | 7.2 vs. 3.4 | NA |
| **Aurora kinase inhibitor monotherapy** | | | | | |
| **Meulenbeld HJ et al**, 2012 | Rx1: Danusertib330 Rx2: Danusertib500 | Not reported | | 2.8 vs. 2.8 | NA |
| **Bipolar Androgen Therapy** | | | | | |
| **TRANSFORMER**, 2021 | Rx: BAT Ctrl: Enzalutamide | 32.9 vs. 29 | 0.95 (0.66-1.39) | 6.1 vs. 8.3 ^†^ | 1.24 (0.87-1.77) |
| **Chemotherapy monotherapy** | | | | | |
| **Heidenreich A et al**, 2004 | Rx1: L-Doxorubicin25  Rx2: L-Doxorubicin50 | Not reported | | Not reported | |
| **Annala M et al**, 2021 | Rx: Cabazitaxel25 Ctrl: Enzalutamide /Abiraterone | 37 vs. 15.5 | 0.58 (0.32-1.05) | 5.3 vs. 2.8 * | 0.87 (0.56-1.35) |
| **TAXYNERGY**, 2017 | Rx: Docetaxel Ctrl: Cabazitaxel25 | Not reported | | Not reported | |
| **TIPC**, 2007 | Rx: Docetaxel30 (W) Ctrl: Prednisolone | 27 vs. 18 | NA | 11 vs. 4 * | NA |
| **GETUG-P02,**  2015 | Rx1: Etoposide Rx2: Vinorelbine Rx3: Mitoxantrone | 8.4 vs. 12.6 vs. 10.6 | NA | 2 vs. 4.1 vs. 3.9 ^†^ | NA |
| **Krainer M et al**, 2007 | Rx: Docetaxel25 (W) Ctrl: Vinorelbine | Not reported | | 14.5 vs. 4.4 ^‡^ | NA |
| **ConCab**, 2018 | Rx: Cabazitaxel (W) Ctrl: Cabazitaxel25 | 15.6 vs. 14.6 | NA | 6.4 vs. 6 * | 0.73 (0.47-1.13) |
| **CAINTA**, 2023 | Rx: Cabazitaxel ID Ctrl: Cabazitaxel25 | 16.2 vs. 7.3 | NA | 9.5 vs. 4.4 ^†^ | NA |
| **Rosenberg JE et al**, 2007 | Rx: Ixabepilone Ctrl: Mitoxantrone | 10.4 vs. 9.8 | NA | Not reported | |
| **Chemotherapy+AKTi** | | | | | |
| **ProCAID**, 2021 | Rx: Docetaxel + Capivasertib Ctrl: Docetaxel | 25.3 vs. 20.3 | 0.70 (0.47-1.05) | 7 vs. 6.7 * | 0.92 (0.65-1.31) |
| **RE-AKT**, 2024 | Rx: Enzalutamide  + Capivasertib Ctrl: Enzalutamide | 13.9 vs. 11 | 0.76 (0.51-1.15) | 5.6 vs. 3.5 ^†^ | 0.78 (0.47-1.30) |
| **Chemotherapy+Antifungal** | | | | | |
| **Millikan R et al,**  2003 | Rx: Vin/Estra + Keto/Doxorubicin Ctrl: Paclitaxel + Estramustine + Etoposide | 23.4 vs. 16.9 | NA | Not reported | |
| **Chemotherapy+ARPI** | | | | | |
| **ABIDO SOGUG**, 2022 | Rx: Docetaxel + Abiraterone Ctrl: Docetaxel | 17.4 vs. 16.9 | NA | 8.8 vs. 9.8 ^‡^ | NA |
| **CHEIRON**, 2021 | Rx: Docetaxel + Enzalutamide Ctrl: Docetaxel | 28.7 vs. 30.3 | 1.11 (0.79-1.56) | 12.8 vs. 9.6 ^†^ | 0.62 (0.44-0.85) |
| **Chemotherapy+ASO** | | | | | |
| **Chi KN et al,**  2010 | Rx: Docetaxel + Custirsen Ctrl: Docetaxel | 23.8 vs. 16.9 | 0.61 (0.36-1.02) | 7.3 vs. 6.1 * | 0.86 (0.54-1.38) |
| **Wiechno P et al**, 2014 | Rx: Docetaxel + LY2181308 Ctrl: Docetaxel | 27 vs. 29 | NA | 8.6 vs. 9 * | NA |
| **Chemotherapy+Bcl-2 inhibitor** | | | | | |
| **EORTC**, 2009 | Rx: Docetaxel  + Oblimersen Ctrl: Docetaxel | Not reported | | 4.2 vs. 6.3 ^‡^ | NA |
| **Sonpavde G et al**, 2011 | Rx: Docetaxel + AT-101 Ctrl: Docetaxel | 18.1 vs. 17.8 | 1.07 (0.72-1.55) | 11 vs. 10.3 * | 0.88 (0.63-1.22) |
| **Chemotherapy+Chemotherapy** | | | | | |
| **Machiels JP et al**, 2008 | Rx: Docetaxel35 (W) + Estramustine Ctrl: Docetaxel35 (W) | 19.3 vs. 21 | NA | 6.3 vs. 6.6 * | NA |
| **Berry WR et al**, 2004 | Rx: Paclitaxel + Estramustine Ctrl: Paclitaxel | 16.1 vs. 13.1 | NA | 5.5 vs. 4.3 ^‡^ | NA |
| **Albrecht W et al**, 2004 | Rx: Estramustine + Vinblastine Ctrl: Estramustine | 10.1 vs. 11.7 | NA | Not reported | |
| **Cabrespine A et al**, 2006 | Rx: Paclitaxel + Carboplatin Ctrl: Mitoxantrone | 14.5 vs. 11.1 | NA | Not reported | |
| **Nelius T et al**, 2006 | Rx1: Docetaxel70 + Estramustine840 Rx2: Docetaxel70 + Estramustine420 | 21 vs. 22 | NA | Not reported | |
| **Oudard S et al**, 2005 | Rx1: Docetaxel70 + Estramustine Rx2: Docetaxel35  (D2,D9 Q3W) + Estramustine Ctrl: Mitoxantrone | Rx1 vs Rx2: 18.6 vs. 18.4 Rx1 vs Ctrl:  18.6 vs. 13.4 Rx2 vs Ctrl:  18.4 vs. 13.4 | Rx1 vs Rx2: 1.43 (0.89-2.31) Rx1 vs Ctrl:  1.08 (0.66-1.76) Rx2 vs Ctrl:  0.75 (0.46-1.21) | Not reported | |
| **MDACC Study**, 2019 | Rx: Cabazitaxel25 + Carboplatin Ctrl: Cabazitaxel25 | 18.5 vs. 17.3 | 0.89 (0.63-1.25) | 7.3 vs. 4.5 * | 0.69 (0.50-0.95) |
| **RECARDO**, 2018 | Rx: Docetaxel + Carboplatin Ctrl: Docetaxel | 18.9 vs. 18.5 | NA | 11.7 vs. 12.7 * | NA |
| **Droz JP et al**, 2003 | Rx1: Oxaliplatin + 5-Fluorouracil Rx2: Oxaliplatin | 11.4 vs. 9.4 | NA | 3.4 vs. 2.6 ^‡^ | NA |
| **Galsky MD et al**, 2005 | Rx1: Ixabepilone + Estramustine Rx2: Ixabepilone | Not reported | | Not reported | |
| **Eymard JC et al**, 2007 | Rx: Docetaxel70 + Estramustine Ctrl: Docetaxel | 19.3 vs. 17.8 | NA | 5.7 vs. 2.9 ^‡^ | NA |
| **Chemotherapy+Curcuminoid** | | | | | |
| **Jahanmohan JP et al**, 2021 | Rx: Docetaxel + Curcumin Ctrl: Docetaxel | 15.8 vs. 19.8 | NA | 3.7 vs. 5.3 * | NA |
| **Chemotherapy+Immunotherapy** | | | | | |
| **Dahut WL et al**, 2004 | Rx: Docetaxel30 (W) ^e^ + Thalidomide Ctrl: Docetaxel30 (W) ^e^ | 28.9 vs. 14.7 | NA | 5.9 vs. 3.7 * | NA |
| **IND 209**, 2018 | Rx: Docetaxel  + Reolysin Ctrl: Docetaxel | 19.1 vs. 21.1 | 1.83 (0.96-3.52) | Not reported | |
| **Fizazi K et al**, 2012 | Rx: Mitoxantrone + Siltuximab Ctrl: Mitoxantrone | 10.2 vs. 13 | 1.45 (0.79-2.68) | 3.2 vs. 7.5 * | 1.72 (1.01-2.93) |
| **ECOG 3899**, 2010 | Rx1: 13-cis Retinoic acid + Interferon-alpha2b + Paclitaxel Rx2: Vinorelbine + Mitoxantrone + Estramustine | 13.9 vs. 19.4 | NA | 2.5 vs. 5.9 * | NA |
| **Hakenberg OW et al**, 2019 | Rx: Mitoxantrone  + Olaratumab Ctrl: Mitoxantrone | 14.2 vs. 12.8 | 1.08 (0.72-1.61) | 2.3 vs. 2.4 * | 1.29 (0.87-1.90) |
| **E1809**, 2015 | Rx: Docetaxel + PVAC-VF Ctrl: Docetaxel | 20.8 vs. NR | NA | Not reported | |
| **Kongsted P et al,**  2017 | Rx: Docetaxel  + DCVAC Ctrl: Docetaxel | Not reported | | 5.7 vs. 5.5 * | NA |
| **de Bono et al**, 2014 | Rx: Docetaxel + Figitumumab Ctrl: Docetaxel | Not reported | | 4.9 vs. 7.9 * | 1.44 (1.06-1.96) |
| **Heidenreich A et al,**  2013 | Rx: Docetaxel  + Intetumumab Ctrl: Docetaxel | 17.2 vs. 20.6 | 1.47 (0.85-2.52) | 7.6 vs. 11 * | 1.73 (1.11-2.69) |
| **Chemotherapy+Radiopharmaceutical** | | | | | |
| **Taxium II**, 2017 | Rx: Docetaxel  + Rhenium-188-HEDP Ctrl: Docetaxel | 23.7 vs. 21 | NA | 9.8 vs. 8.6 * | NA |
| **Chemotherapy+TKI** | | | | | |
| **Horti J et al**, 2009 | Rx: Docetaxel + Vandetanib Ctrl: Docetaxel | Not reported | | 7.7 vs. 9.8 ^‡^ | NA |
| **Chemotherapy+Vascular disrupting agent** | | | | | |
| **Pili R et al,**  2010 | Rx: Docetaxel + Vadimezan Ctrl: Docetaxel | 17 vs. 17.2 | 0.80 (0.46-1.39) | 8.7 vs. 8.4 ^‡^ | 0.81 (0.39-1.70) |
| **Chemotherapy+VitD-Analog** | | | | | |
| **Attia S et al**, 2008 | Rx: Docetaxel35 (W) ^f^ + Doxercalciferol Ctrl: Docetaxel35 (W) ^f^ | 17.8 vs. 16.4 | NA | 6.2 vs. 6.2 * | NA |
| **ASCENT**, 2007 | Rx: Docetaxel36 (W) + Calcitriol Ctrl: Docetaxel36 (W) | NR vs. 16.4 | 0.67 (0.45-0.97) ^\|\|^ | Not reported | |
| **Corticosteroid monotherapy** | | | | | |
| **Venkitaraman R et al**, 2015 | Rx: Dexamethasone Ctrl: Prednisolone | Not reported | | Not reported | |
| **Corticosteroid+Immunotherapy** | | | | | |
| **Yoshimura K et al**, 2016 | Rx: Dexamethasone + PPV Ctrl: Dexamethasone | 73.9 vs. 34.9 | 0.41 (0.21-0.83) ^\|\|^ | Not reported | |
| **Corticosteroid+VEGFi** | | | | | |
| **Stadler WM et al**, 2004 | Rx: Dexamethasone + SU5416 Ctrl: Dexamethasone | Not reported | | 2.3 vs. 1 ^‡^ | NA |
| **ERA monotherapy** | | | | | |
| **James ND et al**, 2009 | Rx1: Zibotentan15  Rx2: Zibotentan10 Ctrl: Placebo | Rx1 vs. Ctrl: 23.9 vs. 19.9 Rx2 vs. Ctrl: 23.5 vs. 19.9 | Rx1 vs. Ctrl: 0.76 (0.61-0.94) ^\|\|^ Rx2 vs. Ctrl: 0.83 (0.67-1.02) | Rx1 vs. Ctrl: 3.8 vs. 3.7 ^‡^ Rx2 vs. Ctrl: 4.6 vs. 3.7 ^‡^ | Rx1 vs. Ctrl: 0.86 (0.72-1.04) Rx2 vs. Ctrl: 1.06 (0.89-1.27) |
| **Carducci MA et al**, 2003 | Rx1: Atrasentan2.5 Rx2: Atrasentan10 Ctrl: Placebo | Not reported | | Rx1 vs. Ctrl: 5.9 vs. 4.5 ^‡^ Rx2 vs. Ctrl: 6 vs. 4.5 ^‡^ | NA |
| **IMiD monotherapy** | | | | | |
| **Pili R et al**, 2011 | Rx: Tasquinimod Ctrl: Placebo | 33.4 vs. 30.4 | 0.87 (0.59-1.29) | 8.8 vs. 4.4 ^†^ | 0.54 (0.36-0.82) |
| **Immunotherapy monotherapy** | | | | | |
| **PERSEUS**, 2016 | Rx1: Abituzumab750 + SOC Rx2: Abituzumab1500 + SOC Ctrl: SOC | Not reported | | Rx1 vs. Ctrl: 3.4 vs. 3.3 Rx2 vs. Ctrl: 4.3 vs. 3.3 | Rx1 vs. Ctrl: 0.89 (0.57-1.39) Rx2 vs. Ctrl: 0.81 (0.52-1.26) |
| **Filaci G et al**, 2021 | Rx1: GX301 vaccine (8D) Rx2: GX301 vaccine (4D) Rx3: GX301 vaccine (2D) | 22.9 vs. NR vs. 21.5 | NA | 4.9 vs. 5.7 vs. 4.2 | NA |
| **Figg WD et al**, 2001 | Rx: ThalidomideHD Ctrl: Thalidomide | Not reported | | Not reported | |
| **Immunotherapy+Chemotherapy** | | | | | |
| **Arlen PM et al,**  2006 | Rx: Vaccine + GMCSF + Docetaxel Ctrl: Vaccine + GMCSF | Not reported | | 3.2 vs. 1.8 * | NA |
| **Immunotherapy+GMCSF** | | | | | |
| **TBC-PRO-002**, 2010 | Rx: PVAC+GMCSF Ctrl: Placebo | 25.1 vs. 16.6 | 0.56 (0.37-0.85) ^\|\|^ | 3.8 vs. 3.7 * | 0.88 (0.57-1.38) |
| **Integrin inhibitor monotherapy** | | | | | |
| **Bradley DA et al,**  2011 | Rx: Cilengitide500 Ctrl: Cilengitide2000 | Not reported | | 2.7 vs. 2.8 ^‡^ | NA |
| **Matrix Metalloproteinase Inhibitor monotherapy** | | | | | |
| **Lara Jr PN et al**, 2006 | Rx: BMS-275291  [1200mg] Ctrl: BMS-275291 [2400mg] | NR vs. 21 | NA | Not reported | |
| **PARPi+ARPi** | | | | | |
| **BRCAAway**, 2024 | Rx1: Abiraterone Rx2: Olaparib Rx3: Abiraterone+Olaparib | Not reported | | Rx3 vs. Rx1: 39 vs. 8.6 * Rx3 vs. Rx2: 39 vs. 14 * | Rx3 vs. Rx1: 0.33 (0.15-0.72) Rx3 vs. Rx2: 0.37 (0.17-0.84) |
| **Progestin monotherapy** | | | | | |
| **Patel SR et al**, 1990 | Rx: Megestrol Acetate Ctrl: Dexamethasone | 8.8 vs. 8.1 | NA | Not reported | |
| **Radioligand monotherapy** | | | | | |
| **TheraP**, 2021 | Rx: 177Lu Ctrl: Cabazitaxel20 | 16.4 vs. 19.4 | 0.97 (0.70-1.35) | NA | 0.64 (0.46-0.88) ^b^ |
| **Somatostatin+Corticosteroid** | | | | | |
| **Dimopoulos MA et al**, 2004 | Rx: Lanreotide + Dexamethasone Ctrl: Estramustine + Etoposide | 18 vs. 18.8 | NA | 4 vs. 6 | NA |
| **TKI monotherapy** | | | | | |
| **Droz JP et al**, 2014 | Rx: Nintedanib150 Ctrl: Nintedanib250 | 8.2 vs. NR | NA | 2.4 vs. 2.5 * | NA |
| **Boccardo F et al**, 2008 | Rx: Gefitinib Ctrl: Placebo | 26.5 vs. 20.5 | 0.69 (0.39-1.23) | 4 vs. 4.5 * | 0.74 (0.46-1.18) |
| **Monk P et al**, 2018 | Rx: Tivantinib Ctrl: Placebo | Not reported | | 5.5 vs. 3.7 * | 0.55 (0.33-0.90) |
| **Smith DC et al**, 2013 | Rx: Cabozantinib Ctrl: Placebo | Not reported | | 5.5 vs. 1.4 ^†^ | NA |
| **TKI+1st gen AA** | | | | | |
| **Azad AA et al**, 2014 | Rx: Vandetanib + Bicalutamide Ctrl: Bicalutamide | Not reported | | Not reported | |
| **Sridhar SS et al**, 2015 | Rx1: Pazopanib + Bicalutamide Rx2: Pazopanib | Not reported | | 7.3 vs. 7.3 * | NA |

Abbreviations: ARPI: androgen-receptor pathway inhibitor; cSipuleucel-T: concurrent sipuleucel-T; sSipuleucel-T: sequential sipuleucel-T; 177Lu: 177Lu-PSMA-617; TKI: tyrosine kinase inhibitor; ASO: antisense oligonucleotide; BAT: bipolar androgen therapy; AKTi: protein kinase B inhibitor; Bcl-2: B-cell lymphoma 2 protein; VitD: vitamin d; VEGFi: vascular endothelial growth factor inhibitor; ERA: endothelin receptor antagonist; IMiD: immunomodulatory drug; GMCSF: granulocyte-macrophage colony-stimulating factor; PARPi: poly(ADP-ribose) polymerase inhibitor; PVAC: PROSTVAC (viral vector-based immunotherapy); AA: antiandrogen; SOC: standard of care; Vin/Estra: vinblastine/estramustine; keto: ketoconazole

The color ‘green’ represents a positive trial, ‘red’ represents a negative trial.

* Composite progression free survival

^†^ Radiographic progression free survival

^‡^ Time to disease progression

^§^ Time to treatment failure

^||^ Statistically significant overall survival benefit was observed

Itraconazole600: Itraconazole 600mg/day PO

Itraconazole200: Itraconazole 200mg/day PO

Danusertib330: Danusertib 330 mg/m^2^ IV on days 1,8 and 15 every 4 weeks

Danusertib500: Danusertib 500 mg/m^2^ IV on days 1 and 15 every 4 weeks

L-Doxorubicin25: Liposomal Doxorubicin (dissolved in 250 mL 5% glucose) 25 mg/m^2^ IV every 2 weeks for 12 consecutive cycles

L-Doxorubicin50: Liposomal Doxorubicin (dissolved in 250 mL 5% glucose) 50 mg/m^2^ IV every 4 weeks for 6 consecutive cycles

Cabazitaxel25: Cabazitaxel 25 mg/m^2^ IV every 3 weeks

Docetaxel30 (W): Docetaxel 30 mg/m^2^ IV every week for 5 weeks, followed by 1 week rest (6-week cycle)

Docetaxel30 (W) ^e^ : Docetaxel 30 mg/m^2^ IV every week for 3 weeks, followed by 1 week rest (4-week cycle)

Docetaxel25 (W): Docetaxel 25 mg/m^2^ IV every week for 2 cycles (8-week cycle)

Cabazitaxel (W): Cabazitaxel 25 mg/m^2^ IV weekly for 5 weeks (6-week cycle)

Cabazitaxel ID: Cabazitaxel at an initial dose of 25 mg/m^2^ followed by dose adaptations in cycle 2 and beyond according to a prespecified dosing algorithm considering previous-cycle hematologic toxicity and cabazitaxel AUC (with a target AUC of 0.8–1.2 mg*hour/L)

Docetaxel35 (W): Docetaxel 35 mg/m^2^ IV on days 2 and 9 every 3 weeks

Docetaxel35 (W) ^f^ : Docetaxel 35 mg/m^2^ IV on days 1, 8, and 15 (4-week cycle)

Docetaxel36 (W): Docetaxel 36 mg/m^2^ IV every week for 3 weeks (4-week cycle)

Docetaxel70: Docetaxel 70 mg/m^2^ on day 2 every 3 weeks

Estramustine840: Estramustine 840 mg/day on days 1-5

Estramustine420: Estramustine 420 mg/day on days 1-3

Docetaxel35 (D2,D9 Q3W): Docetaxel 35 mg/m^2^ on day 2 and 9 every 3wks

Zibotentan15: Zibotentan 15 mg PO OD

Zibotentan10: Zibotentan 10 mg PO OD

Atrasentan2.5: Atrasentan 2.5 mg PO OD

Atrasentan10: Atrasentan 10 mg PO OD

Abituzumab750: Abituzumab 750 mg every 3 weeks

Abituzumab1500: Abituzumab 1500 mg every 3 weeks

GX301 vaccine (8D): GX301, 8 doses on days 1, 3, 5, 7, 14, 21, 35 and 63

GX301 vaccine (4D): GX301, 4 doses on days 1, 14, 35 and 63

GX301 vaccine (2D): GX301, 2 doses on days 1 and 63

ThalidomideHD: Thalidomide initial dose of 200 mg/day with increments of 200 mg/day every 2 weeks to a maximum dose of 1200 mg

Cilengitide500: Cilengitide 500 mg IV twice weekly (six-week cycle)

Cilengitide2000: Cilengitide 2000 mg IV twice weekly (six-week cycle)

Cabazitaxel20: Cabazitaxel 20 mg/m^2^ IV every 3 weeks

### **Supplement Table 19:** Summary of findings with certainty of evidence for phase III trials in which patients received prior ADT only

A) **ARPI monotherapy**

*Abiraterone+Prednisone vs. Placebo*

| **Outcome** | **Relative effect (95% CI)** | **Absolute risk estimates** | | | **Certainty of Evidence** | **Trial Used** |
| --- | --- | --- | --- | --- | --- | --- |
|  |  | Risk with control | Risk with intervention | Risk Difference (95% CI) |  |  |
| Overall Survival | **HR 0.81** (0.70-0.93) | 714 per 1000 | 637 per 1000 | **77 fewer per 1000** (130 fewer-26 fewer) | High | COU-AA-302 |
| Progression-free Survival | **HR 0.52** (0.45-0.61) | 649 per 1000 | 420 per 1000 | **229 fewer per 1000** (273 fewer-177 fewer) | High | COU-AA-302 |

*Enzalutamide vs. Placebo*

| **Outcome** | **Relative effect (95% CI)** | **Absolute risk estimates** | | | **Certainty of Evidence** | **Trial Used** |
| --- | --- | --- | --- | --- | --- | --- |
|  |  | Risk with control | Risk with intervention | Risk Difference (95% CI) |  |  |
| Overall Survival | **HR 0.77** (0.67-0.88) | 422 per 1000 | 344 per 1000 | **78 fewer per 1000** (115 fewer-39 fewer) | High | PREVAIL |
| Progression-free survival | **HR 0.32** (0.28-0.36) | 401 per 1000 | 151 per 1000 | **250 fewer per 1000** (267 fewer-232 fewer) | High | PREVAIL |

*TAK-700 vs. Placebo*

| **Outcome** | **Relative effect (95% CI)** | **Absolute risk estimates** | | | **Certainty of Evidence** | **Trial Used** |
| --- | --- | --- | --- | --- | --- | --- |
|  |  | Risk with control | Risk with intervention | Risk Difference (95% CI) |  |  |
| Overall Survival | **HR 0.92**  (0.79-1.08) | 407 per 1000 | 382 per 1000 | **25 fewer per 1000** (69 fewer-24 more) | Low * | ELM-PC 4 |
| Progression-free Survival | **HR 0.71**  (0.63-0.80) | 720  per 1000 | 595 per 1000 | **125 fewer per 1000** (168 fewer-81 fewer) | High | ELM-PC 4 |

* Very serious imprecision due to wide confidence interval and null effect indicating both clinical benefit and harm

B) **ARPI+ARPI**

*Abiraterone+Apalutamide vs. Abiraterone alone*

| **Outcome** | **Relative effect (95% CI)** | **Absolute risk estimates** | | | **Certainty of Evidence** | **Trial Used** |
| --- | --- | --- | --- | --- | --- | --- |
|  |  | Risk with control | Risk with intervention | Risk Difference (95% CI) |  |  |
| Overall Survival | **HR 0.95** (0.81-1.11) | 686 per 1000 | 667 per 1000 | **19 fewer per 1000** (77 fewer-38 more) | Low * | ACIS |
| Progression-free Survival | **HR 0.86** (0.72-1.04) | 496 per 1000 | 445 per 1000 | **51 fewer per 1000** (107 fewer-14 more) | Low * | ACIS |

* Very serious imprecision due to wide confidence interval and null effect indicating both clinical benefit and harm

*Enzalutamide+Abiraterone vs. Enzalutamide alone*

| **Outcome** | **Relative effect (95% CI)** | **Absolute risk estimates** | | | **Certainty of Evidence** | **Trial Used** |
| --- | --- | --- | --- | --- | --- | --- |
|  |  | Risk with control | Risk with intervention | Risk Difference (95% CI) |  |  |
| Overall Survival | **HR 0.89** (0.78-1.01) | 740 per 1000 | 698 per 1000 | **42 fewer per 1000** (90 fewer-3 more) | Low * | AllianceA031201 |
| Progression-free Survival | **HR 0.86** (0.76-0.97) | 800 per 1000 | 749 per 1000 | **51 fewer per 1000** (94 fewer-10 fewer) | High | AllianceA031201 |

* Very serious imprecision due to wide confidence interval and null effect indicating both clinical benefit and harm

C) **ARPI+PI3K/AKTi**

*Abiraterone+Ipatasertib vs. Abiraterone alone*

| **Outcome** | **Relative effect (95% CI)** | **Absolute risk estimates** | | | **Certainty of Evidence** | **Trial Used** |
| --- | --- | --- | --- | --- | --- | --- |
|  |  | Risk with control | Risk with intervention | Risk Difference (95% CI) |  |  |
| Overall Survival | **HR 0.93** (0.73-1.18) | 258 per 1000 | 242 per 1000 | **16 fewer per 1000** (62 fewer-39 more) | Very Low *^,†^ | IPATential150 |
| Progression-free Survival | **HR 0.82** (0.68-0.99) | 496 per 1000 | 430 per 1000 | **66 fewer per 1000** (124 fewer-3 fewer) | High | IPATential150 |

Note: PFS subgroup data for no prior taxane chemotherapy was reported in the trial and used here.

* Very serious imprecision due to wide confidence interval and null effect indicating both clinical benefit and harm

^†^ Serious indirectness due to receipt of prior docetaxel in some patients

D) **ARPI+Radiopharmaceutical**

*Abiraterone+Radium-223 vs. Abiraterone alone*

| **Outcome** | **Relative effect (95% CI)** | **Absolute risk estimates** | | | **Certainty of Evidence** | **Trial Used** |
| --- | --- | --- | --- | --- | --- | --- |
|  |  | Risk with control | Risk with intervention | Risk Difference (95% CI) |  |  |
| Overall Survival | **HR 1.19** (0.95-1.50) | 348 per 1000 | 399 per 1000 | **51 more per 1000** (14 fewer-126 more) | Very Low *^,†^ | ERA 223 |
| Progression-free Survival | **HR 1.15** (0.96-1.38) | 553 per 1000 | 604 per 1000 | **51 more per 1000** (15 fewer-118 more) | Very Low *^,†^ | ERA 223 |

* Very serious imprecision due to wide confidence interval and null effect indicating both clinical benefit and harm

^†^ Serious indirectness due to receipt of prior docetaxel and ARPI in some patients

E) **Chemotherapy monotherapy**

*Cabazitaxel25 vs. Docetaxel*

| **Outcome** | **Relative effect (95% CI)** | **Absolute risk estimates** | | | **Certainty of Evidence** | **Trial Used** |
| --- | --- | --- | --- | --- | --- | --- |
|  |  | Risk with control | Risk with intervention | Risk Difference (95% CI) |  |  |
| Overall Survival | **HR 0.97** (0.82-1.16) | 714 per 1000 | 703 per 1000 | **11 fewer per 1000** (72 fewer-52 more) | Moderate * | FIRSTANA |
| Progression-free survival | **HR 0.96** (0.79-1.17) | 545 per 1000 | 530 per 1000 | **15 fewer per 1000** (82 fewer-57 more) | Moderate * | FIRSTANA |

* Serious imprecision due to null effect indicating both clinical benefit and harm

*Cabazitaxel20 vs. Docetaxel*

| **Outcome** | **Relative effect (95% CI)** | **Absolute risk estimates** | | | **Certainty of Evidence** | **Trial Used** |
| --- | --- | --- | --- | --- | --- | --- |
|  |  | Risk with control | Risk with intervention | Risk Difference (95% CI) |  |  |
| Overall Survival | **HR 1.01** (0.85-1.20) | 714 per 1000 | 718 per 1000 | **4 more per 1000** (59 fewer-63 more) | Moderate * | FIRSTANA |
| Progression-free survival | **HR 0.92** (0.75-1.12) | 545 per 1000 | 515 per 1000 | **30 fewer per 1000** (99 fewer-41 more) | Moderate * | FIRSTANA |

* Serious imprecision due to null effect indicating both clinical benefit and harm

*Docetaxel (2W) vs. Docetaxel (standard)*

| **Outcome** | **Relative effect (95% CI)** | **Absolute risk estimates** | | | **Certainty of Evidence** | **Trial Used** |
| --- | --- | --- | --- | --- | --- | --- |
|  |  | Risk with control | Risk with intervention | Risk Difference (95% CI) |  |  |
| Overall Survival | **HR 1.40** (1.10-1.80) | 850 per 1000 | 930 per 1000 | **80 more per 1000** (26 more-117 more) | High | Kellokumpu-Lehtinen PL et al |
| Time to disease progression | **HR 1.30** (1.00-1.60) | 545 per 1000 | 641 per 1000 | **96 more per 1000** (0 more-171 more) | Low * | Kellokumpu-Lehtinen PL et al |

Note: Progression-free survival was not reported

* Very serious imprecision due to wide confidence interval and null effect indicating both clinical benefit and harm

*Docetaxel (intermittent) vs. Docetaxel (standard)*

| **Outcome** | **Relative effect (95% CI)** | **Absolute risk estimates** | | | **Certainty of Evidence** | **Trial Used** |
| --- | --- | --- | --- | --- | --- | --- |
|  |  | Risk with control | Risk with intervention | Risk Difference (95% CI) |  |  |
| Overall Survival | **HR 1.14** (0.75-1.72) | 846 per 1000 | 881 per 1000 | **35 more per 1000** (92 fewer-114 more) | Low * | PRINCE |
| Progression-free Survival | **HR 0.69** (0.43-1.05) | 969 per 1000 | 909 per 1000 | **60 fewer per 1000** (194 fewer-5 more) | Low * | PRINCE |

* Very serious imprecision due to wide confidence interval and null effect indicating both clinical benefit and harm

*Docetaxel vs. Mitoxantrone*

| **Outcome** | **Relative effect (95% CI)** | **Absolute risk estimates** | | | **Certainty of Evidence** | **Trial Used** |
| --- | --- | --- | --- | --- | --- | --- |
|  |  | Risk with control | Risk with intervention | Risk Difference (95% CI) |  |  |
| Overall Survival | **HR 0.79** (0.67-0.93) | 790 per 1000 | 709 per 1000 | **81 fewer per 1000** (141 fewer-24 fewer) | High | TAX327 |

Note: Progression-free survival was not reported

*Docetaxel30(W) vs. Mitoxantrone*

| **Outcome** | **Relative effect (95% CI)** | **Absolute risk estimates** | | | **Certainty of Evidence** | **Trial Used** |
| --- | --- | --- | --- | --- | --- | --- |
|  |  | Risk with control | Risk with intervention | Risk Difference (95% CI) |  |  |
| Overall Survival | **HR 0.87** (0.74-1.02) | 881 per 1000 | 843 per 1000 | **38 fewer per 1000** (88 fewer-5 more) | Low * | TAX327 |

Note: Progression-free survival was not reported

* Very serious imprecision due to wide confidence interval and null effect indicating both clinical benefit and harm

F) **Chemotherapy+ASO**

*Docetaxel+Custirsen vs. Placebo*

| **Outcome** | **Relative effect (95% CI)** | **Absolute risk estimates** | | | **Certainty of Evidence** | **Trial Used** |
| --- | --- | --- | --- | --- | --- | --- |
|  |  | Risk with control | Risk with intervention | Risk Difference (95% CI) |  |  |
| Overall Survival | **HR 0.93** (0.79-1.10) | 504 per 1000 | 479 per 1000 | **25 fewer per 1000** (79 fewer-34 more) | Very Low *^,†^ | SYNERGY |

Note: Progression-free survival was not reported

* Very serious imprecision due to wide confidence interval and null effect indicating both clinical benefit and harm

^†^ Serious indirectness due to receipt of prior ARPI in some patients

G) **Chemotherapy+Chemotherapy**

*Docetaxel60+Estramustine vs. Mitoxantrone*

| **Outcome** | **Relative effect (95% CI)** | **Absolute risk estimates** | | | **Certainty of Evidence** | **Trial Used** |
| --- | --- | --- | --- | --- | --- | --- |
|  |  | Risk with control | Risk with intervention | Risk Difference (95% CI) |  |  |
| Overall Survival | **HR 0.80** (0.67-0.97) | 699 per 1000 | 617 per 1000 | **82 fewer per 1000** (146 fewer-11 fewer) | High | SWOG-  99-16 |

Note: Progression-free survival was not reported

H) **Chemotherapy+ERA**

*Docetaxel+Zibotentan vs. Docetaxel*

| **Outcome** | **Relative effect (95% CI)** | **Absolute risk estimates** | | | **Certainty of Evidence** | **Trial Used** |
| --- | --- | --- | --- | --- | --- | --- |
|  |  | Risk with control | Risk with intervention | Risk Difference (95% CI) |  |  |
| Overall Survival | **HR 1.00** (0.84-1.18) | 530 per 1000 | 530 per 1000 | **0 fewer per 1000** (60 fewer-60 more) | Low * | ENTHUSE (M1c) |
| Progression-free Survival | **HR 1.00** (0.87-1.14) | 820 per 1000 | 820 per 1000 | **0 fewer per 1000** (45 fewer-38 more) | Low * | ENTHUSE (M1c) |

* Very serious imprecision due to wide confidence interval and null effect indicating both clinical benefit and harm

*Docetaxel+Atrasentan vs. Docetaxel*

| **Outcome** | **Relative effect (95% CI)** | **Absolute risk estimates** | | | **Certainty of Evidence** | **Trial Used** |
| --- | --- | --- | --- | --- | --- | --- |
|  |  | Risk with control | Risk with intervention | Risk Difference (95% CI) |  |  |
| Overall Survival | **HR 1.04** (0.90-1.19) | 774 per 1000 | 787 per 1000 | **13 more per 1000** (36 fewer-56 more) | Low * | SWOG S0421 |
| Progression-free Survival | **HR 1.02**  (0.89-1.16) | 893 per 1000 | 898 per 1000 | **5 more per 1000** (30 fewer-32 more) | Low * | SWOG S0421 |

* Very serious imprecision due to wide confidence interval and null effect indicating both clinical benefit and harm

I) **Chemotherapy+IMiD**

*Docetaxel+Lenalidomide vs. Docetaxel*

| **Outcome** | **Relative effect (95% CI)** | **Absolute risk estimates** | | | **Certainty of Evidence** | **Trial Used** |
| --- | --- | --- | --- | --- | --- | --- |
|  |  | Risk with control | Risk with intervention | Risk Difference (95% CI) |  |  |
| Overall Survival | **HR 1.53** (1.17-2.00) | 175 per 1000 | 255 per 1000 | **80 more per 1000** (27 more-144 more) | High | MAINSAIL |
| Progression-free Survival | **HR 1.32**  (1.05-1.66) | 546 per 1000 | 647 per 1000 | **101 more per 1000** (18 more-184 more) | High | MAINSAIL |

J) **Chemotherapy+PDGFRi**

*Docetaxel(30W)+Imatinib vs. Docetaxel30(W)*

| **Outcome** | **Relative effect (95% CI)** | **Absolute risk estimates** | | | **Certainty of Evidence** | **Trial Used** |
| --- | --- | --- | --- | --- | --- | --- |
|  |  | Risk with control | Risk with intervention | Risk Difference (95% CI) |  |  |
| Overall Survival | **HR 1.67**  (0.77-3.64) | 254 per 1000 | 387 per 1000 | **133 more per 1000** (52 fewer-402 more) | Low * | Mathew P  et al |

Note: Progression-free survival was not reported

* Very serious imprecision due to wide confidence interval and null effect indicating both clinical benefit and harm

K) **Chemotherapy+TKI**

*Docetaxel+Dasatinib vs. Docetaxel*

| **Outcome** | **Relative effect (95% CI)** | **Absolute risk estimates** | | | **Certainty of Evidence** | **Trial Used** |
| --- | --- | --- | --- | --- | --- | --- |
|  |  | Risk with control | Risk with intervention | Risk Difference (95% CI) |  |  |
| Overall Survival | **HR 0.99**  (0.87-1.13) | 608 per 1000 | 604 per 1000 | **4 fewer per 1000** (51 fewer-45 more) | Low * | READY |
| Progression-free Survival | **HR 0.92**  (0.82-1.05) | 678 per 1000 | 647 per 1000 | **31 fewer per 1000** (73 fewer-18 more) | Low * | READY |

* Very serious imprecision due to wide confidence interval and null effect indicating both clinical benefit and harm

L) **Chemotherapy+VEGFi**

*Docetaxel+Bevacizumab vs. Docetaxel*

| **Outcome** | **Relative effect (95% CI)** | **Absolute risk estimates** | | | **Certainty of Evidence** | **Trial Used** |
| --- | --- | --- | --- | --- | --- | --- |
|  |  | Risk with control | Risk with intervention | Risk Difference (95% CI) |  |  |
| Overall Survival | **HR 0.91**  (0.70-1.05) | 850 per 1000 | 822 per 1000 | **28 fewer per 1000** (115 fewer-14 more) | Low * | CALGB 90401 |
| Progression-free Survival | **HR 0.80**  (0.71-0.91) | 890 per 1000 | 829 per 1000 | **61 fewer per 1000** (99 fewer-24 fewer) | High | CALGB 90401 |

* Very serious imprecision due to wide confidence interval and null effect indicating both clinical benefit and harm

*Docetaxel+Aflibercept vs. Docetaxel*

| **Outcome** | **Relative effect (95% CI)** | **Absolute risk estimates** | | | **Certainty of Evidence** | **Trial Used** |
| --- | --- | --- | --- | --- | --- | --- |
|  |  | Risk with control | Risk with intervention | Risk Difference (95% CI) |  |  |
| Overall Survival | **HR 0.94**  (0.82-1.08) | 727 per 1000 | 705 per 1000 | **22 fewer per 1000** (72 fewer-27 more) | Low * | VENICE |

Note: Progression-free survival was not reported

* Very serious imprecision due to wide confidence interval and null effect indicating both clinical benefit and harm

M) **ERA monotherapy**

*Zibotentan vs. Placebo*

| **Outcome** | **Relative effect (95% CI)** | **Absolute risk estimates** | | | **Certainty of Evidence** | **Trial Used** |
| --- | --- | --- | --- | --- | --- | --- |
|  |  | Risk with control | Risk with intervention | Risk Difference (95% CI) |  |  |
| Overall Survival | **HR 0.87**  (0.69-1.10) | 498 per 1000 | 451 per 1000 | **47 fewer per 1000** (120 fewer-33 more) | Low * | ENTHUSE (PF) |
| Progression-free Survival | **HR 1.01**  (0.85-1.21) | 870 per 1000 | 873 per 1000 | **3 more per 1000** (47 fewer-45 fewer) | Low * | ENTHUSE (PF) |

* Very serious imprecision due to wide confidence interval and null effect indicating both clinical benefit and harm

*Atrasentan vs. Placebo*

| **Outcome** | **Relative effect (95% CI)** | **Absolute risk estimates** | | | **Certainty of Evidence** | **Trial Used** |
| --- | --- | --- | --- | --- | --- | --- |
|  |  | Risk with control | Risk with intervention | Risk Difference (95% CI) |  |  |
| Overall Survival | **HR 0.97**  (0.81-1.17) | 713 per 1000 | 702 per 1000 | **11 fewer per 1000** (77 fewer-55 more) | Low * | Carducci MA et al |
| Time to disease progression | **HR 0.89**  (0.76-1.04) | 776 per 1000 | 736 per 1000 | **40 fewer per 1000** (97 fewer-13 more) | Low * | Carducci MA et al |

Note: Progression-free survival was not reported

* Very serious imprecision due to wide confidence interval and null effect indicating both clinical benefit and harm

N) **IMiD monotherapy**

*Tasquinimod vs. Placebo*

| **Outcome** | **Relative effect (95% CI)** | **Absolute risk estimates** | | | **Certainty of Evidence** | **Trial Used** |
| --- | --- | --- | --- | --- | --- | --- |
|  |  | Risk with control | Risk with intervention | Risk Difference (95% CI) |  |  |
| Overall Survival | **HR 1.01**  (0.94-1.28) | 576 per 1000 | 580 per 1000 | **4 more per 1000** (22 fewer-91 more) | Very Low *^,†^ | Sternberg C et al |
| Progression-free Survival | **HR 0.64**  (0.54-0.75) | 625 per 1000 | 466 per 1000 | **159 fewer per 1000** (214 fewer-104 fewer) | Moderate ^†^ | Sternberg C et al |

* Very serious imprecision due to wide confidence interval and null effect indicating both clinical benefit and harm

^†^ Serious indirectness due to receipt of prior ARPI in some patients

O) **Immunotherapy monotherapy**

*Ipilimumab vs. Placebo*

| **Outcome** | **Relative effect (95% CI)** | **Absolute risk estimates** | | | **Certainty of Evidence** | **Trial Used** |
| --- | --- | --- | --- | --- | --- | --- |
|  |  | Risk with control | Risk with intervention | Risk Difference (95% CI) |  |  |
| Overall Survival | **HR 1.11**  (0.88-1.39) | 624 per 1000 | 662 per 1000 | **38 more per 1000** (47 fewer-119 more) | Low * | CA184-095 |
| Progression-free Survival | **HR 0.67**  (0.55-0.81) | 870 per 1000 | 745 per 1000 | **125 fewer per 1000** (196 fewer-62 fewer) | High | CA184-095 |

* Very serious imprecision due to wide confidence interval and null effect indicating both clinical benefit and harm

*Sipuleucel-T*

| **Outcome** | **Relative effect (95% CI)** | **Absolute risk estimates** | | | **Certainty of Evidence** | **Trial Used** |
| --- | --- | --- | --- | --- | --- | --- |
|  |  | Risk with control | Risk with intervention | Risk Difference (95% CI) |  |  |
| Overall survival | **HR 0.73** (0.61-0.98) | 659 per 1000 | 544 per 1000 | **115 fewer per 1000** (178 fewer-43 fewer) | Moderate * | IMPACT, D9902A, D9901 |
| Time to disease progression | **HR 0.88** (0.73-1.06) | 776 per 1000 | 732 per 1000 | **44 fewer per 1000** (111 fewer-19 more) | Very Low *^,†^ | IMPACT, D9902A, D9901 |

Note: Progression-free survival was not reported

* Serious indirectness due to differences in receipt of prior chemotherapy in the included trials

^†^ Very serious imprecision due to wide confidence interval and null effect indicating both clinical benefit and harm

P) **Immunotherapy+GMCSF**

*PVAC+GMCSF vs. Placebo*

| **Outcome** | **Relative effect (95% CI)** | **Absolute risk estimates** | | | **Certainty of Evidence** | **Trial Used** |
| --- | --- | --- | --- | --- | --- | --- |
|  |  | Risk with control | Risk with intervention | Risk Difference (95% CI) |  |  |
| Overall Survival | **HR 1.02**  (0.86-1.22) | 714 per 1000 | 721 per 1000 | **7 more per 1000** (55 fewer-69 more) | Low * | PROSPECT |

Note: Progression-free survival was not reported

* Very serious imprecision due to wide confidence interval and null effect indicating both clinical benefit and harm

*PVAC vs. Placebo*

| **Outcome** | **Relative effect (95% CI)** | **Absolute risk estimates** | | | **Certainty of Evidence** | **Trial Used** |
| --- | --- | --- | --- | --- | --- | --- |
|  |  | Risk with control | Risk with intervention | Risk Difference (95% CI) |  |  |
| Overall Survival | **HR 1.01**  (0.84-1.20) | 714 per 1000 | 718 per 1000 | **4 more per 1000** (63 fewer-63 more) | Low * | PROSPECT |

Note: Progression-free survival was not reported

* Very serious imprecision due to wide confidence interval and null effect indicating both clinical benefit and harm

Q) **PARPi+ARPI**

*Talazoparib+Enzalutamide*

i) **Overall Population**

| **Outcome** | **Relative effect (95% CI)** | **Absolute risk estimates** | | | **Certainty of Evidence** | **Trial Used** |
| --- | --- | --- | --- | --- | --- | --- |
|  |  | Risk with control | Risk with intervention | Risk Difference (95% CI) |  |  |
| Overall Survival | **HR 0.80** (0.66-0.96) | 603 per 1000 | 522 per 1000 | **81 fewer per 1000** (147 fewer-15 fewer) | Moderate * | TALAPRO-2 |
| Progression-free Survival | **HR 0.68** (0.53-0.88) | 450 per 1000 | 334 per 1000 | **116 fewer per 1000** (178 fewer-41 fewer) | Moderate * | TALAPRO-2 |

Note: PFS subgroup data for only those patients who received prior ADT alone was reported in the trial and used here

* Serious indirectness due to differences in the trial designs and receipt of prior therapy

ii) **Patients with *HRR alterations***

| **Outcome** | **Relative effect (95% CI)** | **Absolute risk estimates** | | | **Certainty of Evidence** | **Trial Used** |
| --- | --- | --- | --- | --- | --- | --- |
|  |  | Risk with control | Risk with intervention | Risk Difference (95% CI) |  |  |
| Overall Survival | **HR 0.62**  (0.48-0.81) | 633 per 1000 | 463 per 1000 | **170 fewer per 1000** (251 fewer-77 fewer) | Moderate * | TALAPRO-2 |
| Progression-free Survival | **HR 0.47**  (0.36-0.61) | 523 per 1000 | 294 per 1000 | **229 fewer per 1000** (289 fewer-160 fewer) | Moderate * | TALAPRO-2 |

* Serious indirectness due to differences in the trial designs and receipt of prior therapy

iii) **Patients with *BRCA1/2* alterations**

| **Outcome** | **Relative effect (95% CI)** | **Absolute risk estimates** | | | **Certainty of Evidence** | **Trial Used** |
| --- | --- | --- | --- | --- | --- | --- |
|  |  | Risk with control | Risk with intervention | Risk Difference (95% CI) |  |  |
| Overall Survival | **HR 0.50** (0.32-0.78) | 320 per 1000 | 175 per 1000 | **145 fewer per 1000** (204 fewer-60 fewer) | Moderate * | TALAPRO-2 |
| Progression-free Survival | **HR 0.20** (0.11-0.36) | 643 per 1000 | 186 per 1000 | **457 fewer per 1000** (536 fewer-333 fewer) | Moderate * | TALAPRO-2 |

* Serious indirectness due to differences in the trial designs and receipt of prior therapy

iv) **Patients with *HRR* alterations other than *BRCA1/2***

| **Outcome** | **Relative effect (95% CI)** | **Absolute risk estimates** | | | **Certainty of Evidence** | **Trial Used** |
| --- | --- | --- | --- | --- | --- | --- |
|  |  | Risk with control | Risk with intervention | Risk Difference (95% CI) |  |  |
| Overall Survival | **HR 0.73** (0.52-1.02) | 320 per 1000 | 245 per 1000 | **75 fewer per 1000** (138 fewer-5 more) | Very Low *^,†^ | TALAPRO-2 |
| Progression-free Survival | **HR 0.71** (0.52-0.96) | 520 per 1000 | 406 per 1000 | **114 fewer per 1000** (203 fewer-14 fewer) | Low * | TALAPRO-2 |

Note: Data for progression-free survival was used from the post-hoc analysis of the TALAPRO-2 trial recently published by Shore ND et al (https://doi.org/10.1200/JCO.2024.42.4_suppl.136)

* Very serious indirectness due to heterogenous population in *BRCA* negative subgroup and receipt of prior therapy

^†^ Very serious imprecision due to wide confidence interval and null effect indicating both clinical benefit and harm

v) **Patients with no *HRR alterations***

| **Outcome** | **Relative effect (95% CI)** | **Absolute risk estimates** | | | **Certainty of Evidence** | **Trial Used** |
| --- | --- | --- | --- | --- | --- | --- |
|  |  | Risk with control | Risk with intervention | Risk Difference (95% CI) |  |  |
| Overall Survival | **HR 0.88**  (0.71-1.08) | 450 per 1000 | 409 per 1000 | **41 fewer per 1000** (104 fewer-26 more) | Very Low *^,†^ | TALAPRO-2 |
| Progression-free Survival | **HR 0.70** (0.54-0.89) | 445 per 1000 | 338 per 1000 | **107 fewer per 1000** (173 fewer-37 fewer) | Moderate * | TALAPRO-2 |

* Serious indirectness due to differences in the trial designs and receipt of prior therapy

^†^ Very serious imprecision due to wide confidence interval and null effect indicating both clinical benefit and harm

### **Supplement Table 20:** Summary of findings with certainty of evidence for phase III trials in which patients received prior ADT + ARPI

A) **ARPI+ARPI**

*Enzalutamide+Abiraterone vs. Abiraterone*

| **Outcome** | **Relative effect (95% CI)** | **Absolute risk estimates** | | | **Certainty of Evidence** | **Trial Used** |
| --- | --- | --- | --- | --- | --- | --- |
|  |  | Risk with control | Risk with intervention | Risk Difference (95% CI) |  |  |
| Progression-free survival | **HR 0.83** (0.61-1.12) | 736 per 1000 | 669 per 1000 | **67 fewer per 1000** (180 fewer-39 more) | Low * | PLATO |

Note: Overall survival was not reported

* Very serious imprecision due to wide confidence interval and null effect indicating both clinical benefit and harm

B) **ARPI+Immunotherapy**

*Enzalutamide+Atezolizumab vs. Enzalutamide*

| **Outcome** | **Relative effect (95% CI)** | **Absolute risk estimates** | | | **Certainty of Evidence** | **Trial Used** |
| --- | --- | --- | --- | --- | --- | --- |
|  |  | Risk with control | Risk with intervention | Risk Difference (95% CI) |  |  |
| Overall Survival | **HR 1.58**  (1.13-2.20) | 345 per 1000 | 488 per 1000 | **143 more per 1000** (35 more-261 more) | High | IMbassador250 |
| Progression-free Survival | **HR 0.98**  (0.75-1.27) | 643  per 1000 | 636 per 1000 | **7 fewer per 1000** (105 fewer-87 more) | Low * | IMbassador250 |

Note: OS and PFS subgroup data for patients with no prior taxane therapy was reported in the trial and used here

* Very serious imprecision due to wide confidence interval and null effect indicating both clinical benefit and harm

C) **Chemotherapy+ARPI**

*Docetaxel+Enzalutamide vs. Docetaxel*

| **Outcome** | **Relative effect (95% CI)** | **Absolute risk estimates** | | | **Certainty of Evidence** | **Trial Used** |
| --- | --- | --- | --- | --- | --- | --- |
|  |  | Risk with control | Risk with intervention | Risk Difference (95% CI) |  |  |
| Progression-free survival | **HR 0.72** (0.53-0.96) | 756 per 1000 | 638 per 1000 | **118 fewer per 1000** (229 fewer-14 fewer) | High | PRESIDE |

D) **Chemotherapy+Immunotherapy**

*Docetaxel+Pembrolizumab vs. Docetaxel*

| **Outcome** | **Relative effect (95% CI)** | **Absolute risk estimates** | | | **Certainty of Evidence** | **Trial Used** |
| --- | --- | --- | --- | --- | --- | --- |
|  |  | Risk with control | Risk with intervention | Risk Difference (95% CI) |  |  |
| Overall survival | **HR 0.92**  (0.78-1.09) | 559 per 1000 | 529  per 1000 | **30 fewer per 1000** (87 fewer-31 more) | Very Low *^,†^ | KEYNOTE-921 |
| Progression-free survival | **HR 0.85**  (0.71-1.01) | 522 per 1000 | 466 per 1000 | **56 fewer per 1000** (114 fewer-4 more) | Very Low *^,†^ | KEYNOTE-921 |

* Serious indirectness due to receipt of prior docetaxel in some patients

^†^ Very serious imprecision due to wide confidence interval and null effect indicating both clinical benefit and harm

E) **PARPi monotherapy**
i) **Overall Population**

*Olaparib vs. Enzalutamide/Abiraterone*

| **Outcome** | **Relative effect (95% CI)** | **Absolute risk estimates** | | | **Certainty of Evidence** | **Trial Used** |
| --- | --- | --- | --- | --- | --- | --- |
|  |  | Risk with control | Risk with intervention | Risk Difference (95% CI) |  |  |
| Overall survival | **HR 0.79**  (0.61-1.03) | 670 per 1000 | 583  per 1000 | **87 fewer per 1000** (179 fewer-11 more) | Very Low *^,†^ | PROfound |
| Progression-free survival | **HR 0.49**  (0.38-0.63) | 670 per 1000 | 419 per 1000 | **251 fewer per 1000** (326 fewer-167 fewer) | Moderate * | PROfound |

* Serious indirectness due to receipt of prior docetaxel in some patients

^†^ Very serious imprecision due to wide confidence interval and null effect indicating both clinical benefit and harm

| **Outcome** | **Relative effect (95% CI)** | **Absolute risk estimates** | | | **Certainty of Evidence** | **Trial Used** |
| --- | --- | --- | --- | --- | --- | --- |
|  |  | Risk with control | Risk with intervention | Risk Difference (95% CI) |  |  |
| Overall survival | **HR 1.12**  (0.69-1.85) | 510 per 1000 | 550 per 1000 | **40 more per 1000** (121 fewer-223 more) | Low * | PROfound |
| Progression-free survival | **HR 0.77** (0.50-1.22) | 733 per 1000 | 638 per 1000 | **95 fewer per 1000** (250 fewer-67 more) | Low * | PROfound |

Note: OS and PFS subgroup data for no prior taxane use was reported and used here

* Very serious imprecision due to wide confidence interval and null effect indicating both clinical benefit and harm

*Rucaparib vs. Enzalutamide/Abiraterone/Docetaxel*

| **Outcome** | **Relative effect (95% CI)** | **Absolute risk estimates** | | | **Certainty of Evidence** | **Trial Used** |
| --- | --- | --- | --- | --- | --- | --- |
|  |  | Risk with control | Risk with intervention | Risk Difference (95% CI) |  |  |
| Overall survival | **HR 0.94** (0.72-1.23) | 670 per 1000 | 647 per 1000 | **23 fewer per 1000** (120 fewer-74 more) | Very Low *^,†^ | TRITON-3 |
| Progression-free survival | **HR 0.61** (0.47-0.80) | 696 per 1000 | 516 per 1000 | **180 fewer per 1000** (267 fewer-82 fewer) | Moderate * | TRITON-3 |

* Serious indirectness due to receipt of prior docetaxel in some patients

^†^ Very serious imprecision due to wide confidence interval and null effect indicating both clinical benefit and harm

*Rucaparib vs. Enzalutamide/Abiraterone*

| **Outcome** | **Relative effect (95% CI)** | **Absolute risk estimates** | | | **Certainty of Evidence** | **Trial Used** |
| --- | --- | --- | --- | --- | --- | --- |
|  |  | Risk with control | Risk with intervention | Risk Difference (95% CI) |  |  |
| Overall survival | **HR 0.38** (0.25-0.58) | 680 per 1000 | 351 per 1000 | **329 fewer per 1000** (432 fewer-196 fewer) | Moderate * | TRITON-3 |
| Progression-free survival | **HR 0.47** (0.34-0.66) | 730 per 1000 | 460 per 1000 | **270 fewer per 1000** (371 fewer-151 fewer) | Moderate * | TRITON-3 |

Note: Overall survival data was only reported for the population with BRCA alterations

* Serious indirectness due to receipt of prior docetaxel in some patients

*Rucaparib vs. Docetaxel*

| **Outcome** | **Relative effect (95% CI)** | **Absolute risk estimates** | | | **Certainty of Evidence** | **Trial Used** |
| --- | --- | --- | --- | --- | --- | --- |
|  |  | Risk with control | Risk with intervention | Risk Difference (95% CI) |  |  |
| Overall survival | **HR 0.53** (0.37-0.77) | 680 per 1000 | 453 per 1000 | **227 fewer per 1000** (336 fewer-96 fewer) | Moderate * | TRITON-3 |
| Progression-free survival | **HR 0.64** (0.46-0.88) | 670 per 1000 | 508 per 1000 | **162 fewer per 1000** (271 fewer-47 fewer) | Moderate * | TRITON-3 |

Note: Overall survival data was only reported for the population with BRCA alterations

* Serious indirectness due to receipt of prior docetaxel in some patients

ii) **Patients with *HRR* alterations**

*Olaparib vs. Enzalutamide/Abiraterone*

Refer to Section (i) PROfound data. All patients in PROfound had *HRR* alterations.

*Rucaparib vs. Enzalutamide/Abiraterone/Docetaxel*

Refer to Section (i) for TRITON-3 data. All patients in TRITON-3 had *HRR* alterations.

iii) **Patients with *BRCA1/2* alterations**

*Olaparib vs. Enzalutamide/Abiraterone*

| **Outcome** | **Relative effect (95% CI)** | **Absolute risk estimates** | | | **Certainty of Evidence** | **Trial Used** |
| --- | --- | --- | --- | --- | --- | --- |
|  |  | Risk with control | Risk with intervention | Risk Difference (95% CI) |  |  |
| Overall survival | **HR 0.63** (0.42-0.95) | 707 per 1000 | 539 per 1000 | **168 fewer per 1000** (304 fewer-19 fewer) | Moderate * | PROfound |
| Progression-free survival | **HR 0.22** (0.15-0.32) | 879 per 1000 | 372 per 1000 | **507 fewer per 1000** (607 fewer-388 fewer) | Moderate * | PROfound |

* Serious indirectness due to receipt of prior docetaxel in some patients

*Rucaparib vs. Enzalutamide/Abiraterone/Docetaxel*

| **Outcome** | **Relative effect (95% CI)** | **Absolute risk estimates** | | | **Certainty of Evidence** | **Trial Used** |
| --- | --- | --- | --- | --- | --- | --- |
|  |  | Risk with control | Risk with intervention | Risk Difference (95% CI) |  |  |
| Overall survival | **HR 0.81** (0.58-1.12) | 564 per 1000 | 490 per 1000 | **74 fewer per 1000** (182 fewer-41 more) | Very Low *^,†^ | TRITON-3 |
| Progression-free survival | **HR 0.50** (0.36-0.69) | 663 per 1000 | 420 per 1000 | **243 fewer per 1000** (339 fewer-135 fewer) | Moderate * | TRITON-3 |

* Serious indirectness due to receipt of prior docetaxel in some patients

^†^ Very serious imprecision due to wide confidence interval and null effect indicating both clinical benefit and harm

iv) **Patients with *HRR* alterations other than *BRCA1/2***

*Olaparib vs. Enzalutamide/Abiraterone*

| **Outcome** | **Relative effect (95% CI)** | **Absolute risk estimates** | | | **Certainty of Evidence** | **Trial Used** |
| --- | --- | --- | --- | --- | --- | --- |
|  |  | Risk with control | Risk with intervention | Risk Difference (95% CI) |  |  |
| Overall survival | **HR 0.79**  (0.61-1.03) | 672 per 1000 | 585 per 1000 | **87 fewer per 1000** (179 fewer-11 more) | Low *^,†^ | PROfound |
| Progression-free survival | **HR 0.49** (0.38-0.63) | 550 per 1000 | 324 per 1000 | **226 fewer per 1000** (288 fewer-155 fewer) | Moderate ^†^ | PROfound |

Note: Data from Cohort A+B of the PROfound trial was used for overall survival and progression free survival. The following report was used: https://doi.org/10.1056/NEJMoa2022485

* Serious imprecision due to null effect indicating both clinical benefit and harm

^†^ Serious indirectness due to receipt of prior docetaxel in some patients

| **Outcome** | **Relative effect (95% CI)** | **Absolute risk estimates** | | | **Certainty of Evidence** | **Trial Used** |
| --- | --- | --- | --- | --- | --- | --- |
|  |  | Risk with control | Risk with intervention | Risk Difference (95% CI) |  |  |
| Overall survival | **HR 1.03** (0.70-1.50) | 620 per 1000 | 631 per 1000 | **11 more per 1000** (128 fewer-146 more) | Very Low *^,†^ | PROfound |
| Progression-free survival | **HR 0.92** (0.64-1.32) | 550 per 1000 | 520 per 1000 | **30 fewer per 1000** (150 fewer-101 more) | Very Low *^,†^ | PROfound |

Note: Data from non *BRCA1/2* HRR genes were pooled using a fixed-effect meta-analysis model from the PROfound trial.

* Serious indirectness due to receipt of prior docetaxel in some patients

^†^ Very serious imprecision due to wide confidence interval and null effect indicating both clinical benefit and harm

*Rucaparib vs. Enzalutamide/Abiraterone/Docetaxel*

| **Outcome** | **Relative effect (95% CI)** | **Absolute risk estimates** | | | **Certainty of Evidence** | | **Trial Used** |
| --- | --- | --- | --- | --- | --- | --- | --- |
|  |  | Risk with control | Risk with intervention | Risk Difference (95% CI) |  |  |  |
| Overall survival | **HR 1.20**  (0.74-1.95) | 620 per 1000 | 687 per 1000 | **67 more per 1000** (109 fewer-228 more) | Very Low *^,†^ | TRITON-3 | |
| Progression-free survival | **HR 0.95** (0.59-1.52) | 735 per 1000 | 717 per 1000 | **18 fewer per 1000** (192 fewer-132 more) | Very Low *^,†^ | | TRITON-3 |

Note: Overall survival was not reported

* Serious indirectness due to receipt of prior docetaxel in some patients

^†^ Very serious imprecision due to wide confidence interval and null effect indicating both clinical benefit and harm

v) **Radioligand monotherapy**

*177LuPSMA 617 vs. Enzalutamide/Abiraterone*

| **Outcome** | **Relative effect (95% CI)** | **Absolute risk estimates** | | | **Certainty of Evidence** | **Trial Used** |
| --- | --- | --- | --- | --- | --- | --- |
|  |  | Risk with control | Risk with intervention | Risk Difference (95% CI) |  |  |
| Overall Survival | **HR 0.98**  (0.75-1.28) | 479 per 1000 | 472 per 1000 | **7 fewer per 1000** (92 fewer-87 more) | Low * | PSMAfore |
| Progression-free Survival | **HR 0.49**  (0.39-0.61) | 769  per 1000 | 512 per 1000 | **257 fewer per 1000** (334 fewer-178 fewer) | High | PSMAfore |

* Very serious imprecision due to wide confidence interval and null effect indicating both clinical benefit and harm

### **Supplement Table 21:** Summary of findings with certainty of evidence for phase III trials in which patients received prior ADT + Docetaxel

A) **ARPI monotherapy**

*Abiraterone + Prednisone vs. Placebo*

| **Outcome** | **Relative effect (95% CI)** | **Absolute risk estimates** | | | **Certainty of Evidence** | **Trial Used** |
| --- | --- | --- | --- | --- | --- | --- |
|  |  | Risk with control | Risk with intervention | Risk Difference (95% CI) |  |  |
| Overall Survival | **HR 0.74** (0.64-0.86) | 550 per 1000 | 446 per 1000 | **104 fewer per 1000** (150 fewer-53 fewer) | High | COU-AA-301 |
| Progression-free Survival | **HR 0.66** (0.58-0.76) | 648 per 1000 | 498 per 1000 | **150 fewer per 1000** (194 fewer-100 fewer) | High | COU-AA-301 |

*Enzalutamide vs. Placebo*

| **Outcome** | **Relative effect (95% CI)** | **Absolute risk estimates** | | | **Certainty of Evidence** | **Trial Used** |
| --- | --- | --- | --- | --- | --- | --- |
|  |  | Risk with control | Risk with intervention | Risk Difference (95% CI) |  |  |
| Overall Survival | **HR 0.63** (0.53-0.75) | 530 per 1000 | 379 per 1000 | **151 fewer per 1000** (200 fewer-98 fewer) | High | AFFIRM |
| Progression-free survival | **HR 0.40** (0.35-0.47) | 649 per 1000 | 342 per 1000 | **307 fewer per 1000** (342 fewer -260 fewer) | High | AFFIRM |

*TAK-700 (400mg) vs. Placebo*

| **Outcome** | **Relative effect (95% CI)** | **Absolute risk estimates** | | | **Certainty of Evidence** | **Trial Used** |
| --- | --- | --- | --- | --- | --- | --- |
|  |  | Risk with control | Risk with intervention | Risk Difference (95% CI) |  |  |
| Overall Survival | **HR 0.89**  (0.74-1.06) | 499 per 1000 | 459 per 1000 | **40 fewer per 1000** (99 fewer-20 more) | Low * | ELM-PC5 |
| Progression-free survival | **HR 0.76** (0.65-0.89) | 718 per 1000 | 618 per 1000 | **100 fewer per 1000** (157 fewer -42 fewer) | High | ELM-PC5 |

* Very serious imprecision due to wide confidence interval and null effect indicating both clinical benefit and harm

*TAK-700 (300mg) vs. Placebo*

| **Outcome** | **Relative effect (95% CI)** | **Absolute risk estimates** | | | **Certainty of Evidence** | **Trial Used** |
| --- | --- | --- | --- | --- | --- | --- |
|  |  | Risk with control | Risk with intervention | Risk Difference (95% CI) |  |  |
| Progression-free survival | **HR 0.42** (0.20-0.91) | 500 per 1000 | 253 per 1000 | **247 fewer per 1000** (371 fewer-32 fewer) | Moderate * | SAKK 08/11 |

* Serious indirectness present as TAK-700 was administered as maintenance therapy after patients were stable on first-line docetaxel

B) **Chemotherapy monotherapy**

*Cabazitaxel25 vs. Mitoxantrone*

| **Outcome** | **Relative effect (95% CI)** | **Absolute risk estimates** | | | **Certainty of Evidence** | **Trial Used** |
| --- | --- | --- | --- | --- | --- | --- |
|  |  | Risk with control | Risk with intervention | Risk Difference (95% CI) |  |  |
| Overall Survival | **HR 0.70** (0.59-0.83) | 740 per 1000 | 611 per 1000 | **129 fewer per 1000** (192 fewer-67 fewer) | High | TROPIC |
| Progression-free survival | **HR 0.74** (0.64-0.86) | 915 per 1000 | 839 per 1000 | **76 fewer per 1000** (121 fewer -35 fewer) | High | TROPIC |

*Cabazitaxel20 vs. Cabazitaxel25*

| **Outcome** | **Relative effect (95% CI)** | **Absolute risk estimates** | | | **Certainty of Evidence** | **Trial Used** |
| --- | --- | --- | --- | --- | --- | --- |
|  |  | Risk with control | Risk with intervention | Risk Difference (95% CI) |  |  |
| Overall Survival | **HR 1.02**  (0.92-1.18) | 620 per 1000 | 627 per 1000 | **7 more per 1000** (31 fewer-61 more) | Low * | PROSELICA |
| Progression-free survival | **HR 1.10** (0.97-1.24) | 885 per 1000 | 907 per 1000 | **22 more per 1000** (8 fewer -47 more) | Very Low *^,†^ | PROSELICA |

Note: OS subgroup data for patients with no prior ARPI was reported in the trial and used here

* Very serious imprecision due to wide confidence interval and null effect indicating both clinical benefit and harm

^†^ Serious indirectness due to receipt of prior ARPI in some patients

C) **Chemotherapy+ASO**

*Cabazitaxel25+Custirsen vs. Cabazitaxel25*

| **Outcome** | **Relative effect (95% CI)** | **Absolute risk estimates** | | | **Certainty of Evidence** | **Trial Used** |
| --- | --- | --- | --- | --- | --- | --- |
|  |  | Risk with control | Risk with intervention | Risk Difference (95% CI) |  |  |
| Overall Survival | **HR 0.95**  (0.80-1.12) | 840 per 1000 | 825 per 1000 | **15 fewer per 1000** (71 fewer-32 more) | Very Low *^,†^ | AFFINITY |

* Very serious imprecision due to wide confidence interval and null effect indicating both clinical benefit and harm

^†^ Serious indirectness due to receipt of prior ARPI in some patients

D) **TKI monotherapy**

*Sunitinib vs. Placebo*

| **Outcome** | **Relative effect (95% CI)** | **Absolute risk estimates** | | | **Certainty of Evidence** | **Trial Used** |
| --- | --- | --- | --- | --- | --- | --- |
|  |  | Risk with control | Risk with intervention | Risk Difference (95% CI) |  |  |
| Overall Survival | **HR 0.91**  (0.76-1.10) | 713 per 1000 | 679 per 1000 | **34 fewer per 1000** (100 fewer-34 more) | Low * | SUN 1120 |
| Progression-free survival | **HR 0.73** (0.59-0.89) | 519 per 1000 | 414 per 1000 | **105 fewer per 1000** (168 fewer -40 fewer) | High | SUN 1120 |

* Very serious imprecision due to wide confidence interval and null effect indicating both clinical benefit and harm

### **Supplement Table 22:** Summary of findings with certainty of evidence for phase III trials in which patients received prior ADT + ARPI + Docetaxel

A) **Chemotherapy monotherapy**

*Cabazitaxel25 vs. Enzalutamide/Abiraterone*

| **Outcome** | **Relative effect (95% CI)** | **Absolute risk estimates** | | | **Certainty of Evidence** | **Trial Used** |
| --- | --- | --- | --- | --- | --- | --- |
|  |  | Risk with control | Risk with intervention | Risk Difference (95% CI) |  |  |
| Overall survival | **HR 0.64** (0.46-0.89) | 659 per 1000 | 498 per 1000 | **161 fewer per 1000** (269 fewer-43 fewer) | Moderate * | CARD |
| Progression-Free survival | **HR 0.54** (0.40-0.73) | 802 per 1000 | 583 per 1000 | **219 fewer per 1000** (325 fewer-109 fewer) | Moderate * | CARD |

* Serious indirectness due to lack of patients receiving “true” triplet therapy in hormone sensitive setting

B) **PARPi+Immunotherapy**
i) **Overall Population**

*Olaparib+Pembrolizumab vs. Enzalutamide/Abiraterone*

| **Outcome** | **Relative effect (95% CI)** | **Absolute risk estimates** | | | **Certainty of Evidence** | **Trial Used** |
| --- | --- | --- | --- | --- | --- | --- |
|  |  | Risk with control | Risk with intervention | Risk Difference (95% CI) |  |  |
| Overall survival | **HR 0.94**  (0.77-1.14) | 564 per 1000 | 542 per 1000 | **22 fewer per 1000** (92 fewer-48 more) | Very Low *^,†^ | KEYLYNK-010 |
| Progression-free survival | **HR 0.96** (0.79-1.16) | 564 per 1000 | 549 per 1000 | **15 fewer per 1000** (83 fewer-54 more) | Very Low *^,†^ | KEYLYNK-010 |

* Serious indirectness due to lack of patients receiving “true” triplet therapy in hormone sensitive setting

^†^ Very serious imprecision due to wide confidence interval and null effect indicating both clinical benefit and harm

ii) **Patients with *HRR* alterations**

*Olaparib+Pembrolizumab vs. Enzalutamide/Abiraterone*

| **Outcome** | **Relative effect (95% CI)** | **Absolute risk estimates** | | | **Certainty of Evidence** | **Trial Used** |
| --- | --- | --- | --- | --- | --- | --- |
|  |  | Risk with control | Risk with intervention | Risk Difference (95% CI) |  |  |
| Overall survival | **HR 0.88**  (0.59-1.33) | 576 per 1000 | 530 per 1000 | **46 fewer per 1000** (179 fewer-105 more) | Very Low *^,†^ | KEYLYNK-010 |
| Progression-free survival | **HR 0.65** (0.45-0.94) | 627 per 1000 | 473 per 1000 | **154 fewer per 1000** (269 fewer-23 fewer) | Moderate * | KEYLYNK-010 |

* Serious indirectness due to lack of patients receiving “true” triplet therapy in hormone sensitive setting

^†^ Very serious imprecision due to wide confidence interval and null effect indicating both clinical benefit and harm

iii) **Patients with *BRCA 1/2* alterations**

*Olaparib+Pembrolizumab vs. Enzalutamide/Abiraterone*

| **Outcome** | **Relative effect (95% CI)** | **Absolute risk estimates** | | | **Certainty of Evidence** | **Trial Used** |
| --- | --- | --- | --- | --- | --- | --- |
|  |  | Risk with control | Risk with intervention | Risk Difference (95% CI) |  |  |
| Overall survival | **HR 0.52**  (0.27-0.99) | 625 per 1000 | 400 per 1000 | **225 fewer per 1000** (392 fewer-4 fewer) | Moderate * | KEYLYNK-010 |
| Progression-free survival | **HR 0.45** (0.24-0.84) | 625 per 1000 | 357 per 1000 | **268 fewer per 1000** (415 fewer-64 fewer) | Moderate * | KEYLYNK-010 |

* Serious indirectness due to lack of patients receiving “true” triplet therapy in hormone sensitive setting

iv) **Patients with *HRR* alteration*s* other than *BRCA 1/2***

*Olaparib+Pembrolizumab vs. Enzalutamide/Abiraterone*

| **Outcome** | **Relative effect (95% CI)** | **Absolute risk estimates** | | | **Certainty of Evidence** | **Trial Used** |
| --- | --- | --- | --- | --- | --- | --- |
|  |  | Risk with control | Risk with intervention | Risk Difference (95% CI) |  |  |
| Overall survival | **HR 1.12**  (0.91-1.38) | 495 per 1000 | 535 per 1000 | **40 more per 1000** (32 fewer-115 more) | Very Low *^,†^ | KEYLYNK-010 |
| Progression-free survival | **HR 1.00**  (0.80-1.24) | 577 per 1000 | 577 per 1000 | **0 fewer per 1000** (79 fewer-79 more) | Very Low *^,†^ | KEYLYNK-010 |

* Serious indirectness due to lack of patients receiving “true” triplet therapy in hormone sensitive setting

^†^ Very serious imprecision due to wide confidence interval and null effect indicating both clinical benefit and harm

v) **Patients with *no HRR* alterations**

*Olaparib+Pembrolizumab vs. Enzalutamide/Abiraterone*

| **Outcome** | **Relative effect (95% CI)** | **Absolute risk estimates** | | | **Certainty of Evidence** | **Trial Used** |
| --- | --- | --- | --- | --- | --- | --- |
|  |  | Risk with control | Risk with intervention | Risk Difference (95% CI) |  |  |
| Overall survival | **HR 0.97**  (0.76-1.23) | 584 per 1000 | 573 per 1000 | **11 fewer per 1000** (97 fewer-76 more) | Very Low *^,†^ | KEYLYNK-010 |
| Progression-free survival | **HR 1.21**  (0.96-1.53) | 468 per 1000 | 534 per 1000 | **66 more per 1000** (14 fewer-151 more) | Very Low *^,†^ | KEYLYNK-010 |

* Serious indirectness due to lack of patients receiving “true” triplet therapy in hormone sensitive setting

^†^ Very serious imprecision due to wide confidence interval and null effect indicating both clinical benefit and harm

C) **Radioligand therapy + Standard of Care**

*177LuPSMA 617+Standard of Care vs. Standard of Care alone*

| **Outcome** | **Relative effect (95% CI)** | **Absolute risk estimates** | | | **Certainty of Evidence** | **Trial Used** |
| --- | --- | --- | --- | --- | --- | --- |
|  |  | Risk with control | Risk with intervention | Risk Difference (95% CI) |  |  |
| Overall survival | **HR 0.62** (0.52-0.74) | 668 per 1000 | 495 per 1000 | **173 fewer per 1000** (232 fewer-110 fewer) | Moderate * | VISION |
| Progression-free survival | **HR 0.40** (0.29-0.57) | 474 per 1000 | 227 per 1000 | **247 fewer per 1000** (304 fewer-167 fewer) | Moderate * | VISION |

* Serious indirectness due to lack of patients receiving “true” triplet therapy in hormone sensitive setting

D) **TKI monotherapy**

*Cabozantinib vs. Prednisone*

| **Outcome** | **Relative effect (95% CI)** | **Absolute risk estimates** | | | **Certainty of Evidence** | **Trial Used** |
| --- | --- | --- | --- | --- | --- | --- |
|  |  | Risk with control | Risk with intervention | Risk Difference (95% CI) |  |  |
| Overall survival | **HR 0.90**  (0.76-1.06) | 610 per 1000 | 571 per 1000 | **39 fewer per 1000** (99 fewer-21 more) | Very Low *^,†^ | COMET-1 |
| Progression-free survival | **HR 0.48** (0.40-0.57) | 636 per 1000 | 384 per 1000 | **252 fewer per 1000** (303 fewer-198 fewer) | Moderate * | COMET-1 |

* Serious indirectness due to lack of patients receiving “true” triplet therapy in hormone sensitive setting

^†^ Very serious imprecision due to wide confidence interval and null effect indicating both clinical benefit and harm

### **Supplement Table 23:** Summary of findings with certainty of evidence for phase III trials in which patients received heterogeneous prior therapy

A) **Chemotherapy+Immunotherapy**

*Docetaxel+DCVAC vs. Docetaxel*

| **Outcome** | **Relative effect (95% CI)** | **Absolute risk estimates** | | | **Certainty of Evidence** | **Trial Used** |
| --- | --- | --- | --- | --- | --- | --- |
|  |  | Risk with control | Risk with intervention | Risk Difference (95% CI) |  |  |
| Overall survival | **HR 1.04**  (0.90-1.21) | 643 per 1000 | 657 per 1000 | **14 more per 1000** (39 fewer-69 more) | Low * | VIABLE |
| Progression-free survival | **HR 0.99** (0.86-1.14) | 547 per 1000 | 543 per 1000 | **4 fewer per 1000** (53 fewer-48 more) | Low * | VIABLE |

* Very serious imprecision due to wide confidence interval and null effect indicating both clinical benefit and harm

### **Supplement Table 24:** Summary of findings with certainty of evidence for phase III trials that reported data for multiple subgroups

A) **Chemotherapy monotherapy**

*Satraplatin vs. Placebo*

i) Patients who received prior ADT alone

| **Outcome** | **Relative effect (95% CI)** | **Absolute risk estimates** | | | **Certainty of Evidence** | **Trial Used** |
| --- | --- | --- | --- | --- | --- | --- |
|  |  | Risk with control | Risk with intervention | Risk Difference (95% CI) |  |  |
| Overall Survival | **HR 1.03** (0.82-1.29) | 713 per 1000 | 724 per 1000 | **11 more per 1000** (72 fewer-87 more) | Low * | SPARC |

Note: Progression-free survival was not reported

* Very serious imprecision due to wide confidence interval and null effect indicating both clinical benefit and harm

ii) Patients who received prior ADT and Docetaxel

| **Outcome** | **Relative effect (95% CI)** | **Absolute risk estimates** | | | **Certainty of Evidence** | **Trial Used** |
| --- | --- | --- | --- | --- | --- | --- |
|  |  | Risk with control | Risk with intervention | Risk Difference (95% CI) |  |  |
| Overall Survival | **HR 0.91**  (0.72-1.14) | 530 per 1000 | 497 per 1000 | **33 fewer per 1000** (111 fewer-47 more) | Low * | SPARC |

Note: Progression-free survival was not reported

* Very serious imprecision due to wide confidence interval and null effect indicating both clinical benefit and harm

iii) Patients who received heterogeneous prior therapy

| **Outcome** | **Relative effect (95% CI)** | **Absolute risk estimates** | | | **Certainty of Evidence** | **Trial Used** |
| --- | --- | --- | --- | --- | --- | --- |
|  |  | Risk with control | Risk with intervention | Risk Difference (95% CI) |  |  |
| Overall survival | **HR 0.98**  (0.84-1.15) | 759 per 1000 | 752 per 1000 | **7 fewer per 1000** (62 fewer-46 more) | Very Low *^,†^ | SPARC |
| Progression-free survival | **HR 0.67** (0.57-0.77) | 870 per 1000 | 745 per 1000 | **125 fewer per 1000** (183 fewer-78 fewer) | Moderate * | SPARC |

* Serious indirectness due to receipt of heterogeneous prior therapy

^†^ Very serious imprecision due to wide confidence interval and null effect indicating both clinical benefit and harm

B) **Immunotherapy monotherapy**

*Sipuleucel-T vs. Placebo*

i) Patients who received prior ADT alone

| **Outcome** | **Relative effect (95% CI)** | **Absolute risk estimates** | | | **Certainty of Evidence** | **Trial Used** |
| --- | --- | --- | --- | --- | --- | --- |
|  |  | Risk with control | Risk with intervention | Risk Difference (95% CI) |  |  |
| Overall survival | **HR 0.79**  (0.59-1.03) | 545 per 1000 | 463 per 1000 | **82 fewer per 1000** (173 fewer-11 more) | Low * | IMPACT |
| Time to disease progression | **HR 0.95** (0.77-1.17) | 622 per 1000 | 603 per 1000 | **19 fewer per 1000** (95 fewer-58 more) | Low * | IMPACT |

Note: Progression-free survival was not reported

* Very serious imprecision due to wide confidence interval and null effect indicating both clinical benefit and harm

ii) Patients who received prior ADT and Docetaxel

| **Outcome** | **Relative effect (95% CI)** | **Absolute risk estimates** | | | **Certainty of Evidence** | **Trial Used** |
| --- | --- | --- | --- | --- | --- | --- |
|  |  | Risk with control | Risk with intervention | Risk Difference (95% CI) |  |  |
| Overall Survival | **HR 0.68**  (0.35-1.24) | 635 per 1000 | 496 per 1000 | **139 fewer per 1000** (338 fewer-78 more) | Low * | IMPACT |

Note: Progression-free survival was not reported

* Very serious imprecision due to wide confidence interval and null effect indicating both clinical benefit and harm

C) **PARPi+ARPI**

*Niraparib+Abiraterone vs. Abiraterone*

i) **Patients with *HRR* alterations**
i) Patients who received prior ADT and ARPI

| **Outcome** | **Relative effect (95% CI)** | **Absolute risk estimates** | | | **Certainty of Evidence** | **Trial Used** |
| --- | --- | --- | --- | --- | --- | --- |
|  |  | Risk with control | Risk with intervention | Risk Difference (95% CI) |  |  |
| Progression-free survival | **HR 0.83**  (0.48-1.42) | 590 per 1000 | 523 per 1000 | **67 fewer per 1000** (242 fewer-128 more) | Very Low *^,†^ | MAGNITUDE |

Note: Overall survival was not reported

* Serious indirectness due to differences within the trial design

^†^ Very serious imprecision due to wide confidence interval and null effect indicating both clinical benefit and harm

ii) Patients who received prior ADT and Docetaxel

| **Outcome** | **Relative effect (95% CI)** | **Absolute risk estimates** | | | **Certainty of Evidence** | **Trial Used** |
| --- | --- | --- | --- | --- | --- | --- |
|  |  | Risk with control | Risk with intervention | Risk Difference (95% CI) |  |  |
| Progression-free survival | **HR 0.89**  (0.48-1.66) | 512 per 1000 | 472 per 1000 | **40 fewer per 1000** (221 fewer-184 more) | Very Low *^,†^ | MAGNITUDE |

Note: Overall survival was not reported

* Serious indirectness due to differences within the trial design

^†^ Very serious imprecision due to wide confidence interval and null effect indicating both clinical benefit and harm

iii) Patients who received heterogeneous prior therapy

| **Outcome** | **Relative effect (95% CI)** | **Absolute risk estimates** | | | **Certainty of Evidence** | **Trial Used** |
| --- | --- | --- | --- | --- | --- | --- |
|  |  | Risk with control | Risk with intervention | Risk Difference (95% CI) |  |  |
| Overall survival | **HR 0.70**  (0.49-0.99) | 420 per 1000 | 317 per 1000 | **103 fewer per 1000** (186 fewer-3 fewer) | Moderate * | MAGNITUDE |
| Progression-free survival | **HR 0.76** (0.60-0.97) | 550 per 1000 | 455 per 1000 | **95 fewer per 1000** (169 fewer-11 fewer) | Moderate * | MAGNITUDE |

* Serious indirectness due to differences in the trial designs and receipt of prior therapy

ii) **Patients with *BRCA1/2* alterations**

| **Outcome** | **Relative effect (95% CI)** | **Absolute risk estimates** | | | **Certainty of Evidence** | **Trial Used** |
| --- | --- | --- | --- | --- | --- | --- |
|  |  | Risk with control | Risk with intervention | Risk Difference (95% CI) |  |  |
| Overall survival | **HR 0.54**  (0.33-0.90) | 658 per 1000 | 440 per 1000 | **218 fewer per 1000** (360 fewer-39 fewer) | Moderate * | MAGNITUDE |
| Progression-free survival | **HR 0.55** (0.39-0.78) | 570 per 1000 | 371 per 1000 | **199 fewer per 1000** (290 fewer-88 fewer) | Moderate * | MAGNITUDE |

* Serious indirectness due to differences in the trial designs and receipt of prior therapy

iii) **Patients with *HRR* alterations other than *BRCA 1/2***

| **Outcome** | **Relative effect (95% CI)** | **Absolute risk estimates** | | | **Certainty of Evidence** | **Trial Used** |
| --- | --- | --- | --- | --- | --- | --- |
|  |  | Risk with control | Risk with intervention | Risk Difference (95% CI) |  |  |
| Progression-free survival | **HR 0.99** (0.68-1.45) | 540 per 1000 | 536 per 1000 | **4 fewer per 1000** (130 fewer-136 more) | Very Low *^,†^ | MAGNITUDE |

Note: Overall survival was not reported

* Serious indirectness due to differences in the trial designs and receipt of prior therapy

^†^ Very serious imprecision due to wide confidence interval and null effect indicating both clinical benefit and harm

iv) **Patients with no *HRR* alterations**

| **Outcome** | **Relative effect (95% CI)** | **Absolute risk estimates** | | | **Certainty of Evidence** | **Trial Used** |
| --- | --- | --- | --- | --- | --- | --- |
|  |  | Risk with control | Risk with intervention | Risk Difference (95% CI) |  |  |
| Progression-free survival | **HR 1.03**  (0.63-1.67) | 498 per 1000 | 508 per 1000 | **10 more per 1000** (146 fewer-186 more) | Very Low *^,†^ | MAGNITUDE |

Note: Overall survival was not reported.

* Serious indirectness due to differences in the trial designs and receipt of prior therapy

^†^ Very serious imprecision due to wide confidence interval and null effect indicating both clinical benefit and harm

*Olaparib+Abiraterone*

i) **Overall population irrespective of HRR alteration status stratified according to receipt of prior therapy**
i) Patients who received prior ADT alone

| **Outcome** | **Relative effect (95% CI)** | **Absolute risk estimates** | | | **Certainty of Evidence** | **Trial Used** |
| --- | --- | --- | --- | --- | --- | --- |
|  |  | Risk with control | Risk with intervention | Risk Difference (95% CI) |  |  |
| Overall Survival | **HR 0.85** (0.67-1.07) | 480 per 1000 | 426 per 1000 | **54 fewer per 1000** (125 fewer-23 more) | Very Low *^,†^ | PROpel |
| Progression-free Survival | **HR 0.62** (0.49-0.79) | 545 per 1000 | 386 per 1000 | **159 fewer per 1000** (225 fewer-82 fewer) | Moderate * | PROpel |

* Serious indirectness due to differences within the trial design

^†^ Very serious imprecision due to wide confidence interval and null effect indicating both clinical benefit and harm

ii) Patients who received prior ADT and Docetaxel

| **Outcome** | **Relative effect (95% CI)** | **Absolute risk estimates** | | | **Certainty of Evidence** | **Trial Used** |
| --- | --- | --- | --- | --- | --- | --- |
|  |  | Risk with control | Risk with intervention | Risk Difference (95% CI) |  |  |
| Overall Survival | **HR 0.76** (0.52-1.11) | 630 per 1000 | 530 per 1000 | **100 fewer per 1000** (226 fewer-38 more) | Very Low *^,†^ | PROpel |
| Progression-free Survival | **HR 0.66** (0.44-0.98) | 564 per 1000 | 422 per 1000 | **142 fewer per 1000** (258 fewer-7 fewer) | Moderate * | PROpel |

* Serious indirectness due to differences within the trial design

^†^ Very serious imprecision due to wide confidence interval and null effect indicating both clinical benefit and harm

ii) **Patients with *HRR alterations***

*Olaparib+Abiraterone*

| **Outcome** | **Relative effect (95% CI)** | **Absolute risk estimates** | | | **Certainty of Evidence** | **Trial Used** |
| --- | --- | --- | --- | --- | --- | --- |
|  |  | Risk with control | Risk with intervention | Risk Difference (95% CI) |  |  |
| Overall Survival | **HR 0.66** (0.45-0.95) | 600 per 1000 | 454 per 1000 | **146 fewer per 1000** (262 fewer-19 fewer) | Moderate * | PROpel |
| Progression-free Survival | **HR 0.45** (0.31-0.65) | 680 per 1000 | 401 per 1000 | **279 fewer per 1000** (382 fewer-157 fewer) | Moderate * | PROpel |

* Serious indirectness due to differences in the trial designs and receipt of prior therapy

iii) **Patients with *BRCA1/2* alterations**

*Olaparib+Abiraterone*

| **Outcome** | **Relative effect (95% CI)** | **Absolute risk estimates** | | | **Certainty of Evidence** | **Trial Used** |
| --- | --- | --- | --- | --- | --- | --- |
|  |  | Risk with control | Risk with intervention | Risk Difference (95% CI) |  |  |
| Overall Survival | **HR 0.29** (0.14-0.56) | 658 per 1000 | 267 per 1000 | **391 fewer per 1000** (519 fewer-206 fewer) | Moderate * | PROpel |
| Progression-free Survival | **HR 0.18** (0.09-0.34) | 737 per 1000 | 214 per 1000 | **523 fewer per 1000** (624 fewer-372 fewer) | Moderate * | PROpel |

* Serious indirectness due to differences in the trial designs and receipt of prior therapy

iv) **Patients with *HRR* alterations other than *BRCA1/2***

*Olaparib+Abiraterone*

| **Outcome** | **Relative effect (95% CI)** | **Absolute risk estimates** | | | **Certainty of Evidence** | **Trial Used** |
| --- | --- | --- | --- | --- | --- | --- |
|  |  | Risk with control | Risk with intervention | Risk Difference (95% CI) |  |  |
| Overall Survival | **HR 0.91** (0.73-1.13) | 503 per 1000 | 471 per 1000 | **32 fewer per 1000** (103 fewer-43 more) | Very Low *^,†^ | PROpel |
| Progression-free Survival | **HR 0.72** (0.58-0.90) | 554 per 1000 | 441 per 1000 | **113 fewer per 1000** (180 fewer-38 fewer) | Moderate * | PROpel |

* Serious indirectness due to differences in the trial designs and receipt of prior therapy

^†^ Very serious imprecision due to wide confidence interval and null effect indicating both clinical benefit and harm

v) **Patients with no *HRR alterations***

*Olaparib+Abiraterone*

| **Outcome** | **Relative effect (95% CI)** | **Absolute risk estimates** | | | **Certainty of Evidence** | **Trial Used** |
| --- | --- | --- | --- | --- | --- | --- |
|  |  | Risk with control | Risk with intervention | Risk Difference (95% CI) |  |  |
| Overall Survival | **HR 0.89** (0.70-1.14) | 480 per 1000 | 441  per 1000 | **39 fewer per 1000** (113 fewer-45 more) | Very Low *^,†^ | PROpel |
| Progression-free Survival | **HR 0.72** (0.56-0.93) | 498 per 1000 | 391 per 1000 | **107 fewer per 1000** (178 fewer-25 fewer) | Moderate * | PROpel |

* Serious indirectness due to differences in the trial designs and receipt of prior therapy

^†^ Very serious imprecision due to wide confidence interval and null effect indicating both clinical benefit and harm

D) **Radiopharmaceutical monotherapy**

*Radium-223 vs. Placebo*

i) Patients who received prior ADT alone

| **Outcome** | **Relative effect (95% CI)** | **Absolute risk estimates** | | | **Certainty of Evidence** | **Trial Used** |
| --- | --- | --- | --- | --- | --- | --- |
|  |  | Risk with control | Risk with intervention | Risk Difference (95% CI) |  |  |
| Overall survival | **HR 0.69** (0.52-0.92) | 635 per 1000 | 501 per 1000 | **134 fewer per 1000** (227 fewer-31 fewer) | High | ALSYMPCA |

Note: Progression-free survival was not reported

ii) Patients who received prior ADT and Docetaxel

| **Outcome** | **Relative effect (95% CI)** | **Absolute risk estimates** | | | **Certainty of Evidence** | **Trial Used** |
| --- | --- | --- | --- | --- | --- | --- |
|  |  | Risk with control | Risk with intervention | Risk Difference (95% CI) |  |  |
| Overall Survival | **HR 0.70**  (0.56-0.88) | 529 per 1000 | 410 per 1000 | **119 fewer per 1000** (185 fewer-45 fewer) | High | ALSYMPCA |

Note: Progression-free survival was not reported

iii) Patients who received heterogeneous prior therapy

| **Outcome** | **Relative effect (95% CI)** | **Absolute risk estimates** | | | **Certainty of Evidence** | **Trial Used** |
| --- | --- | --- | --- | --- | --- | --- |
|  |  | Risk with control | Risk with intervention | Risk Difference (95% CI) |  |  |
| Overall survival | **HR 0.70** (0.58-0.83) | 635 per 1000 | 506 per 1000 | **129 fewer per 1000** (192 fewer-68 fewer) | Moderate * | ALSYMPCA |

Note: Progression-free survival was not reported

* Serious indirectness due to receipt of heterogeneous prior therapy

### **Supplement Table 25:** Summary of efficacy outcomes according to each gene alteration in trials assessing PARPi combination therapy

| **Gene** | **PROpel** | | | **TALAPRO-2 *** | | | **MAGNITUDE** | | | **Meta Analysis** | | | | |
| --- | --- | --- | --- | --- | --- | --- | --- | --- | --- | --- | --- | --- | --- | --- |
|  | **N** | **OS  HR (95% CI)** | **PFS HR (95% CI)** | **N** | **OS  HR (95% CI)** | **PFS HR (95% CI)** | **N** | **OS ^†^ HR (95% CI)** | **PFS HR (95% CI)** | | **OS** | | **PFS** | |
|  |  |  |  |  |  |  |  |  |  |  | **HR (95% CI)** | ***p*-value** | **HR (95% CI)** | ***p*-value** |
| ***HRR+*** | 111 vs 115 | NR vs. 28.5 | 28.8 vs. 13.8 | 200 vs. 199 | 45.1 vs. 31.1 | 30.7 vs. 12.3 | 212 vs. 211 | NA | 16.7 vs. 13.7 | | 0.65 (0.54-0.78) | <0.001 | 0.55 (0.39-0.79) | 0.001 |
|  |  | 0.66 (0.45-0.95) | 0.45 (0.31-0.65) |  | 0.62 (0.48-0.81) | 0.47 (0.36-0.61) |  | 0.70 (0.49-0.99) | 0.76 (0.60-0.97) | |  |  |  |  |
| ***BRCA+*** | 47 vs. 38 | NR vs. 23 | NR vs. 8.4 | 71 vs. 84 | NA | NA | 113 vs. 112 | NA | 19.5 vs. 10.9 | | 0.46 (0.34-0.63) | <0.001 | 0.28 (0.13-0.62) | 0.002 |
|  |  | 0.29 (0.14-0.56) | 0.18 (0.09-0.34) |  | 0.50 (0.32-0.78) | 0.20 (0.11-0.36) |  | 0.54 (0.33-0.90) | 0.55 (0.39-0.78) | |  |  |  |  |
| *BRCA1* | 6 vs. 3 | NA | NA | 8 vs. 9 | NA | 20 vs. 11.7 | 12 vs. 4 | NA | NA | | NA | NA | 0.12 (0.02-0.73) | 0.009 |
|  |  | 0.17 (0.02-1.59) | 0.07 (0.01-1.38) |  |  | 0.17 (0.02-1.51) |  |  |  |  |  |  |  |  |
| *BRCA2* | 30 vs. 28 | NR vs. 23.6 | NR vs. 8.4 | 55 vs. 60 | NA | NR vs. 11 | 86 vs. 88 | NA | NA | | NA | NA | 0.19 (0.11-0.33) | <0.001 |
|  |  | 0.20 (0.07-0.48) | 0.20 (0.08-0.44) |  |  | 0.19 (0.10-0.38) |  |  |  |  |  |  |  |  |
| ***BRCA-*** | 343 vs 350 | 39.6 vs. 38 | 27.6 vs. 16.6 | 127 vs. 113 | NA | 33.1 vs. 22.1 | 99 vs. 99 | NA | 14.8 vs. 16.4 | | 0.85 (0.69-1.04) | 0.11 | 0.76 (0.64-0.91) | 0.003 |
|  |  | 0.91 (0.73-1.13) | 0.72 (0.58-0.90) |  | 0.73 (0.52-1.02) | 0.71 (0.52-0.96) ^‡^ |  |  | 0.99 (0.68-1.45) | |  |  |  |  |
| *PALB2* | 3 vs. 4 | NA | NA | 6 vs. 5 | NA | NR vs. 8.6 | 8 vs. 4 | NA | NA | | 0.33 (0.10-1.16) | 0.084 | 0.53 (0.21-1.32) | 0.17 |
|  |  | 0.17 (0.02-1.59) | 0.44 (0.05-4.25) |  |  | 0.56 (0.12-2.51) |  | 0.46 (0.10-2.08) | 0.54 (0.14-2.03) | |  |  |  |  |
| *CDK12* | 19 vs. 21 | NR vs. 33.7 | NR vs. 16.6 | 28 vs. 30 | NA | 21.9 vs. 13.8 | 11 vs. 16 | NA | NA | | 0.80 (0.36-1.78) | 0.6 | 0.58 (0.35-0.95) | 0.029 |
|  |  | 0.57 (0.24-1.27) | 0.51 (0.20-1.18) |  |  | 0.49 (0.23-1.02) |  | 1.30 (0.45-3.81) | 0.89 (0.34-2.36) | |  |  |  |  |
| *ATM* | 21 vs. 28 | NR vs. 31.9 | NR vs. 19.9 | 35 vs. 22 | NA | NR vs. 27.7 | 43 vs. 42 | NA | NA | | 0.97 (0.57-1.67) | 0.9 | 0.93 (0.57-1.53) | 0.8 |
|  |  | 0.79 (0.33-1.77) | 0.55 (0.20-1.38) |  |  | 0.76 (0.30-1.94) |  | 1.13 (0.56-2.30) | 1.26 (0.74-2.13) | |  |  |  |  |
| *CHEK2* | 7 vs. 12 | NA | NA | 24 vs. 24 | NA | 22.1 vs. NR | 18 vs. 20 | NA | NA | | 0.81 (0.37-1.75) | 0.6 | 0.92 (0.53-1.61) | 0.8 |
|  |  | 1.14 (0.32-4.04) | 1.06 (0.35-3.25) |  |  | 0.90 (0.34-2.39) |  | 0.66 (0.25-1.75) | 0.87 (0.37-2.01) | |  |  |  |  |
| ***HRR-*** | 279 vs. 273 | 42.1 vs. 38.9 | 27.6 vs. 19.1 | 154 vs. 160 | NA | NR vs 22.5 | 117 vs 116 | NA | 12 vs. NR | | 0.88 (0.75-1.04) | 0.13 | 0.74 (0.63-0.88) | <0.001 |
|  |  | 0.89 (0.70-1.14) | 0.72 (0.56-0.93) |  | 0.88 (0.71-1.08) | 0.70 (0.54-0.89) |  |  | 1.03 (0.63-1.67) | |  |  |  |  |

* Data from TALAPRO-2 cohort 2 was used

^†^ Data from MAGNITUDE trial adjusted for subsequent therapy and cross-over was used for overall survival

^‡^ Data from TALAPRO-2 cohort 1 was used

### **Supplement Table 26:** Subsequent therapy in included phase III trials after patients received protocol treatment

| **Trial** | **Arm** | **Subsequent Therapy** | | | | |
| --- | --- | --- | --- | --- | --- | --- |
|  |  | **ARPI** | **PARPi** | **Chemotherapy** | **Immunotherapy** | **Radiopharmaceuticals** |
| **Prior ADT only** | | | | | | |
| **ARPI monotherapy** | | | | | | |
| **COU-AA-302**, 2013 | Rx: Abiraterone Ctrl: Placebo | 80 (7) | NA | 591 (54) | 51 (5) | NA |
| **PREVAIL**, 2014 | Rx: Enzalutamide Ctrl: Placebo | 943 (55) | NA | 1090 (63) | 28 (2) | 38 (2) |
| **ELM-PC 4**, 2015 | Rx: TAK-700 Ctrl: Placebo | 359 (23) | NA | 585 (38) | 25 (2) | NA |
| **ARPI+ARPI** | | | | | | |
| **ACIS**, 2021 | Rx: Abiraterone  + Apalutamide Ctrl: Abiraterone | 117 (12) | NA | 389 (40) | 7 (<1) | 51 (5) |
| **AllianceA031201**, 2023 | Rx: Enzalutamide + Abiraterone Ctrl: Enzalutamide | 404 (31) | 27 (2) | 665 (51) | 71 (5) | 274 (21) |
| **ARPI+PI3K/AKTi** | | | | | | |
| **IPATential150**, 2021 | Rx: Abiraterone + Ipatasertib Ctrl: Abiraterone | NA | NA | NA | NA | NA |
| **ARPI+Radiopharmaceutical** | | | | | | |
| **ERA 223**, 2019 | Rx: Abiraterone + Radium-223 Ctrl: Abiraterone | 203 (25) | NA | 325 (40) | NA | 25 (3) |
| **Chemotherapy monotherapy** | | | | | | |
| **CALGB9182**, 1999 | Rx: Mitoxantrone + Hydrocortisone Ctrl: Hydrocortisone | NA | NA | NA | NA | NA |
| **FIRSTANA**, 2017 | Rx1: Cabazitaxel20 Rx2: Cabazitaxel25 Ctrl: Docetaxel | 691 (59) | NA | 544 (47) | NA | NA |
| **Kellokumpu-Lehtinen PL et al**, 2013 | Rx: Docetaxel (2W) Ctrl: Docetaxel | NA | NA | NA | NA | NA |
| **Berry W et al**, 2002 | Rx: Mitoxantrone Ctrl: Placebo | NA | NA | NA | NA | NA |
| **Abratt RP et al**, 2004 | Rx: Vinorelbine + Hydrocortisone Ctrl: Hydrocortisone | NA | NA | NA | NA | NA |
| **PRINCE**, 2018 | Rx: Docetaxel (int) Ctrl: Docetaxel | NA | NA | 41 (26) | 1 (<1) | 11 (7) |
| **TAX327**, 2004 | Rx1: Docetaxel Rx2: Docetaxel30(W) Ctrl: Mitoxantrone | NA | NA | NA | NA | NA |
| **Chemotherapy+ASO** | | | | | | |
| **SYNERGY**, 2017 | Rx: Docetaxel + Custirsen Ctrl: Docetaxel | 744 (73) | NA | 369 (36) | NA | 257 (25) |
| **Chemotherapy+Chemotherapy** | | | | | | |
| **SWOG-99-16**, 2004 | Rx: Docetaxel60 + Estramustine Ctrl: Mitoxantrone | NA | NA | NA | NA | NA |
| **Chemotherapy+DES** | | | | | | |
| **ECOG 3882** 2003 | Rx: Doxorubicin + DES Ctrl: Doxorubicin | NA | NA | NA | NA | NA |
| **Chemotherapy+ERA** | | | | | | |
| **ENTHUSE (M1c)**, 2013 | Rx: Docetaxel + Zibotentan Ctrl: Docetaxel | NA | NA | NA | NA | NA |
| **SWOG S0421**, 2013 | Rx: Docetaxel + Atrasentan Ctrl: Docetaxel | NA | NA | NA | NA | NA |
| **Chemotherapy+IMiD** | | | | | | |
| **MAINSAIL**, 2015 | Rx: Docetaxel + Lenalidomide Ctrl: Docetaxel | NA | NA | NA | NA | NA |
| **Chemotherapy+PDGFRi** | | | | | | |
| **Mathew P et al**, 2007 | Rx: Docetaxel30(W) + Imatinib Ctrl: Docetaxel30(W) | NA | NA | NA | NA | NA |
| **Chemotherapy+TKI** | | | | | | |
| **READY**, 2013 | Rx: Docetaxel + Dasatinib Ctrl: Docetaxel | NA | NA | NA | NA | NA |
| **Chemotherapy+VEGFi** | | | | | | |
| **CALGB 90401**, 2012 | Rx: Docetaxel + Bevacizumab Ctrl: Docetaxel | NA | NA | NA | NA | NA |
| **VENICE**, 2013 | Rx: Docetaxel+ Aflibercept Ctrl: Docetaxel | NA | NA | NA | NA | NA |
| **Chemotherapy+VitD-Analog** | | | | | | |
| **ASCENT2**, 2011 | Rx: Docetaxel + Calcitriol Ctrl: Docetaxel | NA | NA | NA | NA | NA |
| **ERA monotherapy** | | | | | | |
| **ENTHUSE (PF)**, 2012 | Rx: Zibotentan Ctrl: Placebo | NA | NA | NA | NA | NA |
| **Carducci MA et al**, 2007 | Rx: Atrasentan Ctrl: Placebo | NA | NA | NA | NA | NA |
| **IMiD monotherapy** | | | | | | |
| **Sternberg C et al**, 2016 | Rx: Tasquinimod Ctrl: Placebo | NA | NA | NA | NA | NA |
| **Immunotherapy monotherapy** | | | | | | |
| **CA184-095**, 2017 | Rx: Ipilimumab Ctrl: Placebo | 70 (12) | NA | 136 (23) | 18 (3) | NA |
| **D9901**, 2006 | Rx: Sipuleucel-T  Ctrl: Placebo | NA | NA | NA | NA | NA |
| **D9902A**, 2009 | Rx: Sipuleucel-T  Ctrl: Placebo | NA | NA | NA | NA | NA |
| **Immunotherapy+GMCSF** | | | | | | |
| **PROSPECT**, 2019 | Rx1: PVAC + GMCSF Rx2: PVAC Ctrl: Placebo | 1065 (83) |  | 682 (53) | 69 (5) | 132 (10) |
| **PARPi+ARPI** | | | | | | |
| **PROpel**, 2023 | Rx: Olaparib + Abiraterone Ctrl: Abiraterone | 129 (16) | 7 (1) | 366 (46) | 49 (6) | 32 (4) |
| **TALAPRO-2**, 2023 | Rx: Talazoparib + Enzalutamide Ctrl: Enzalutamide | 81 (10) | 14 (2) | 243 (30) | 1 (<1) | 40 (5) |
| **Phenylurea monotherapy** | | | | | | |
| **Small EJ et al**, 2000 | Rx: Suramin Ctrl: Placebo | NA | NA | NA | NA | NA |
| **Prior ADT+ARPI** | | | | | | |
| **ARPI+ARPI** | | | | | | |
| **PLATO**, 2018 | Rx: Enzalutamide + Abiraterone Ctrl: Abiraterone | NA | NA | NA | NA | NA |
| **ARPI+Immunotherapy** | | | | | | |
| **IMbassador250**, 2022 | Rx: Enzalutamide + Atezolizumab Ctrl: Enzalutamide | NA | NA | NA | NA | NA |
| **Chemotherapy+ARPI** | | | | | | |
| **PRESIDE**, 2022 | Rx: Docetaxel  + Enzalutamide Ctrl: Docetaxel | NA | NA | NA | NA | NA |
| **KEYNOTE-921**, 2025 | Rx: Docetaxel  + Pembrolizumab Ctrl: Docetaxel | 296 (29) | NA | 485 (47) | NA | 76 (7) |
| **PARPi monotherapy** | | | | | | |
| **TRITON-3**, 2023 | Rx: Rucaparib Ctrl: Enzalutamide /Abiraterone /Docetaxel | NA | 225 (58) | 143 (35) | NA | NA |
| **PROfound**, 2020 | Rx: Olaparib Ctrl: Enzalutamide /Abiraterone | 52 (13) | 89 (22) | 48 (12) | 15 (4) | 5 (1) |
| **Radioligand monotherapy** | | | | | | |
| **PSMAfore**, 2024 | Rx: 177Lu Ctrl: Enzalutamide /Abiraterone | NA | 8 (2) | 209 (45) | NA | 18 (4) |
| **Prior ADT+Docetaxel** | | | | | | |
| **ARPI monotherapy** | | | | | | |
| **AFFIRM**, 2012 | Rx: Enzalutamide Ctrl: Placebo | 264 (22) | NA | 323 (27) | NA | NA |
| **COU-AA-301**, 2011 | Rx: Abiraterone Ctrl: Placebo | NA | NA | NA | NA | NA |
| **ELM-PC 5**, 2015 | Rx: TAK-700 Ctrl: Placebo | 272 (25) | NA | 245 (22) | NA | NA |
| **SAKK 08/11**, 2016 | Rx: TAK-700 Ctrl: Placebo | 31 (66) |  | 34 (72) | NA | 7 (15) |
| **Chemotherapy monotherapy** | | | | | | |
| **PROSELICA**, 2017 | Rx: Cabazitaxel20 Ctrl: Cabazitaxel25 | NA | NA | NA | NA | NA |
| **TROPIC**, 2010 | Rx: Cabazitaxel25 Ctrl: Mitoxantrone | NA | NA | NA | NA | NA |
| **Chemotherapy+ASO** | | | | | | |
| **AFFINITY**, 2017 | Rx: Cabazitaxel25  + Custirsen Ctrl: Cabazitaxel25 | 279 (44) | NA | 220 (35) | NA | 143 (23) |
| **TKI monotherapy** | | | | | | |
| **SUN 1120**, 2013 | Rx: Sunitinib Placebo | NA | NA | NA | NA | NA |
| **Prior ADT+Docetaxel+ARPI** | | | | | | |
| **Chemotherapy monotherapy** | | | | | | |
| **CARD**, 2019 | Rx: Cabazitaxel25 Ctrl: Enzalutamide /Abiraterone | 32 (13) | NA | 53 (21) | NA | 9 (4) |
| **PARPi+Immunotherapy** | | | | | | |
| **KEYLYNK-010**, 2023 | Rx: Olaparib + Pembrolizumab Ctrl: Enzalutamide /Abiraterone | 96 (12) | NA | 417 (53) | NA | 61 (8) |
| **Radioligand therapy+Standard of Care** | | | | | | |
| **VISION**, 2021 | Rx: 177Lu + SOC Ctrl: SOC | 49 (6) | NA | 227 (27) | 38 (5) | 39 (5) |
| **TKI monotherapy** | | | | | | |
| **COMET-1**, 2016 | Rx: Cabozantinib Ctrl: Prednisone | 275 (27) | NA | 220 (21) | NA | 61 (6) |
| **Heterogeneous Prior Therapy** | | | | | | |
| **Chemotherapy+Immunotherapy** | | | | | | |
| **VIABLE**, 2022 | Rx: Docetaxel + DCVAC Ctrl: Docetaxel | NA | NA | NA | NA | NA |
| **Multiple subgroups (ADT/ADT+ARPI/ADT+Docetaxel/ADT+ARPI+Docetaxel/Heterogeneous prior therapy)** | | | | | | |
| **Chemotherapy monotherapy** | | | | | | |
| **SPARC**, 2009 | Rx: Satraplatin Ctrl: Placebo | NA | NA | NA | NA | NA |
| **Immunotherapy monotherapy** | | | | | | |
| **IMPACT**, 2010 | Rx: Sipuleucel-T  Ctrl: Placebo | NA | NA | NA | NA | NA |
| **PARPi+ARPI** | | | | | | |
| **MAGNITUDE**, 2023 | Rx: Niraparib + Abiraterone Ctrl: Abiraterone | 26 (11) | 25 (6) | 92 (41) | 1 (<1) | 6 (3) |
| **Radiopharmaceutical monotherapy** | | | | | | |
| **ALSYMPCA**, 2013 | Rx: Radium-223 Ctrl: Placebo | NA | NA | NA | NA | NA |

Abbreviations: ARPI: androgen-receptor pathway inhibitor; PI3K/AKTi: phosphatidylinositol 3-kinase and protein kinase B inhibitor; PDGFR: platelet-derived growth factor receptor; DES: diethylstilbestrol diphosphate; PVAC: PROSTVAC (viral vector-based immunotherapy); GMCSF: granulocyte-macrophage colony-stimulating factor; ASO: antisense oligonucleotide; ERA: endothelin receptor antagonist; IMiD: immunomodulatory drug; TKI: tyrosine kinase inhibitor; VitD: vitamin d; PARPi: poly(ADP-ribose) polymerase inhibitor; 177Lu: 177Lu-PSMA-617; SOC: standard of care

Note: PLATO and CARD were phase IV trials. All remaining trials were phase III. The color ‘green’ represents a positive trial, ‘red’ represents a negative trial.

Cabazitaxel20: Cabazitaxel 20 mg/m^2^ IV on day 1 of every 3-week cycle

Cabazitaxel25: Cabazitaxel 25 mg/m^2^ IV on day 1 of every 3-week cycle

Docetaxel (2W): Docetaxel 75 mg/m^2^ IV on days 1 and 15 of a 4-week cycle

Docetaxel (int): Docetaxel 35 mg/m^2^ IV on days 1, 8, 15, repeat cycle at day 29

Docetaxel30 (W): Docetaxel 30 mg/m^2^ IV on days 1, 8, 15, 22 and 29 of a 6-week cycle

Remaining drugs were administered at standard doses.

### **Supplement Table 27:** Summary of efficacy outcomes according to each gene alteration in trials assessing PARPi monotherapy

| **Gene** | **TRITON-3** | | | **PROfound** | | |
| --- | --- | --- | --- | --- | --- | --- |
|  | **N** | **OS  HR (95% CI)** | **PFS HR (95% CI)** | **N** | **OS  HR (95% CI)** | **PFS HR (95% CI)** |
| ***HRR+*** | 270 vs. 135 | 23.6 vs. 20.9 | 10.2 vs. 6.4 | 256 vs. 131 | 17.3 vs. 14 | 5.8 vs. 3.5 |
|  |  | 0.94 (0.72-1.23) | 0.61 (0.47-0.80) |  | 0.79 (0.61-1.03) | 0.49 (0.38-0.63) |
| ***BRCA+*** | 201 vs. 101 | 24.3 vs. 20.8 | 11.2 vs. 6.4 | 102 vs. 58 | 20.1 vs. 14.4 | 9.8 vs. 3.0 |
|  |  | 0.81 (0.58-1.12) | 0.50 (0.36-0.69) |  | 0.63 (0.42-0.95) | 0.22 (0.15-0.32) |
| *BRCA1* | 29 vs. 15 | NA | NA | 8 vs. 5 | NA | NA |
|  |  |  | 0.97 (0.47-2.00) |  | 0.42 (0.12-1.53) | 0.41 (0.13-1.39) |
| *BRCA2* | 172 vs. 86 | NA | NA | 81 vs. 47 | NA | NA |
|  |  |  | 0.45 (0.32-0.63) |  | 0.59 (0.37-0.95) | 0.21 (0.13-0.32) |
| ***BRCA-*** | 69 vs. 34 | 21.1 vs. 21.7 | 8.1 vs. 6.8 | 167 vs. 79 | NA | NA |
|  |  | 1.20 (0.74-1.95) | 0.95 (0.59-1.52) |  | 1.03 (0.70-1.50) | 0.92 (0.64-1.32) |
| *PALB2* | *PALB2* patients not included | | | 3 vs. 1 | NA | NA |
| *CDK12* | *CDK12* patients not included | | | 61 vs. 28 | 14.1 vs. 11.5 | 5.1 vs. 2.2 |
|  |  |  |  |  | 0.97 (0.57-1.71) | 0.74 (0.44-1.31) |
| *ATM* | 69 vs. 34 | 21.1 vs. 21.7 | 8.1 vs. 6.8 | 62 vs. 24 | 18.0 vs. 15.6 | 5.4 vs. 4.7 |
|  |  | 1.20 (0.74-1.95) | 0.95 (0.59-1.52) |  | 0.93 (0.53-1.75) | 1.04 (0.61-1.87) |
| *CHEK2* | *CHEK2* patients not included | | | 7 vs. 5 | 16.6 vs. 17.1 | 5.6 vs. 3.4 |
|  |  |  |  |  | 0.87 (0.19-4.44) | 0.87 (0.23-4.13) |
| ***HRR-*** | *HRR-* patients not included | | | *HRR-* patients not included | | |

Note: Meta-analysis pooling data from TRITON3 and PROfound was not done due to heterogenous population in terms of receipt of prior therapy and different control arms across these trials

### **Supplement Table 28:** Proportion of patients that received docetaxel as subsequent therapy in mHSPC trials assessing ARPI + ADT

| **Trial** | **Arm** | **Receipt of subsequent therapy \| N (%)** | |
| --- | --- | --- | --- |
|  |  | **Any subsequent therapy** | **Docetaxel** |
| **STAMPEDE Arm G** | Abiraterone + ADT | 131 (53) | 115 (46) |
|  | ADT | 310 (58) | 200 (37) |
| **LATITUDE** | Abiraterone + ADT | 125 (21) | 106 (34) |
|  | ADT | 246 (41) | 187 (40) |
| **ENZAMET** | Enzalutamide + ADT | 112 (67) | 45 (27) |
|  | NSAA + ADT | 271 (85) | 69 (22) |
| **ARCHES** | Enzalutamide + ADT | 131 (23) | 48 (8) |
|  | ADT | 221 (38) | 71 (12) |
| **TITAN** | Apalutamide + ADT | 87 (51) | 29 (17) |
|  | ADT | 190 (70) | 67 (25) |
| **ARANOTE** | Darolutamide + ADT | 66 (33) | 46 (23) |
|  | ADT | 68 (43) | 46 (29) |

Abbreviation: ADT: androgen deprivation therapy; mHSPC: metastatic hormone sensitive prostate cancer; NSAA: nonsteroidal antiandrogen

### **Supplement Table 29:** Ongoing trials clinical trials for mCRPC

| **Trial identifier (name)** | **Phase** | **Treatment** | **Control** | **Estimated/**  **Actual**  **enrollment** | **Primary endpoint** | **Status** | **Prior Therapy** |
| --- | --- | --- | --- | --- | --- | --- | --- |
| **AR degrader+ARPI** | | | | | | | |
| **NCT05067140** | 1/2 | ARV-766 + Abiraterone | ARV-766 | 220/NA | DLT; Toxicity; Safety; PSA response | Recruiting | Progressed on/after ARPI; ≤ 2 prior chemotherapy regimens permitted |
| **ARPI monotherapy** | | | | | | | |
| **NCT03851640** | 3 | Deutenzalutamide | Placebo | 255/NA | OS | Recruiting | Prior ARPI and docetaxel use permitted |
| **ARPI+Immunotherapy** | | | | | | | |
| **NCT03834493 (MK-3475-641 /KEYNOTE-641)** | 3 | Enzalutamide + Pembrolizumab | Enzalutamide | 1200/1244 | OS; rPFS | Active,  not recruiting | Prior ARPI use permitted |
| **ARPI+Radiopharmaceutical** | | | | | | | |
| **NCT02194842 (PEACE III)** | 3 | Radium-223 + Enzalutamide | Enzalutamide | NA/446 | rPFS | Active,  not recruiting | Prior docetaxel and ARPI use permitted in mHSPC stage but not in mCRPC |
| **CDK4/6 inhibitor+Immunotherapy** | | | | | | | |
| **NCT04751929** | 2 | Abemaciclib + Atezolizumab | Abemaciclib | 75/NA | 6-mo PFS rate; Overall response rate; DLT; Safety | Active,  not recruiting | Progressed on/after ARPI; Prior Docetaxel use permitted |
| **Chemotherapy+AKTI** | | | | | | | |
| **NCT05348577 (CAPItello280)** | 3 | Docetaxel + Capivasertib | Docetaxel | 790/1017 | OS | Active,  not recruiting | Progressed on ARPI; Prior Docetaxel use permitted in mHSPC stage but not in mCRPC |
| **Chemotherapy+ARPI** | | | | | | | |
| **NCT05762536 (DAROTAXEL)** | 2 | Docetaxel/Cabazitaxel + Darolutamide | Docetaxel /Cabazitaxel | 245/NA | PFS | Recruiting | Progressed on/after ARPI; Prior Docetaxel use permitted in mHSPC stage but not in mCRPC |
| **NCT05627752** | 2/3 | Docetaxel + Enzalutamide | Docetaxel | 120/NA | PFS | Recruiting | Progressed on/after Abiraterone in mHSPC; Prior docetaxel/enzalutamide not permitted |
| **Chemotherapy+Radiopharmaceutical** | | | | | | | |
| **NCT03574571** | 3 | Docetaxel + Radium | Docetaxel | 738/NA | OS | Recruiting | Prior Docetaxel and ARPI use permitted in mHSPC stage but not in mCRPC |
| **CYP11A1 inhibitor monotherapy** | | | | | | | |
| **NCT06136624 (OMAHA1)** | 3 | Opevesostat | Abiraterone /Enzalutamide | 1200/NA | OS; rPFS | Recruiting | Progressed on/after ≤2 taxanes and 1 ARPI |
| **NCT06136650 (OMAHA2a)** | 3 | Opevesostat | Abiraterone /Enzalutamide | 1500/NA | OS; rPFS | Recruiting | Progressed on/after ARPI |
| **PARPI+Chemotherapy** | | | | | | | |
| **NCT03442556 (PLATI-PARP)** | 2 | Induction: Docetaxel + Carboplatin Maintenance: Rucaparib camsylate | None | 20/18 | rPFS | Active,  not recruiting | Prior Sipuleucel-T, ARPI, taxane therapy permitted |
| **PARPi+ARPI** | | | | | | | |
| **NCT04691804** | 3 | Fuzulaparib  + Abiraterone | Abiraterone | 804/NA | rPFS | Recruiting | No prior therapy in mCRPC |
| **Radiopharmaceutical monotherapy** | | | | | | | |
| **NCT06402331 (AlphaBreak)** | 2/3 | Rx1: FPI-2265 (50 kBq/kg) Rx2: FPI-2265 (75 kBq/kg) Rx3: FPI-2265 (100 kBq/kg) | NA | 60/NA | Safety; PSA50 response | Recruiting | Prior treatment with Lu177-PSMA therapy mandatory |
| **Radiopharmaceutical+DNA-PK inhibitor+Immunotherapy** | | | | | | | |
| **NCT04071236** | 1/2 | Rx1: Radium-223 Rx2: Radium-223 + Nedisertib Rx3: Radium-223 + Nedisertib + Avelumab | NA | 90/NA | rPFS | Recruiting | Progressed on/after ARPI or taxane therapy |
| **Radioligand+Radiopharmaceutical** | | | | | | | |
| **NCT05383079 (AlphaBet)** | 1/2 | 177Lu-PSMA + Radium-223 | NA | 36/NA | DLT; MTD; RP2D; PSA50 response rate | Recruiting | Progressed on/after ARPI |
| **Radioligand+Immunotherapy** | | | | | | | |
| **NCT05766371** | 2 | 177Lu-PSMA + Pembrolizumab | None | 48/NA | 1-yr rPFS | Recruiting | Progressed on/after ARPI or taxane therapy |
| **NCT03093428** | 2 | Radium-223 + Pembrolizumab | Radium-223 | 45/45 | Number of participants with increased immune cell infiltration | Active,  not recruiting | Progressed on any therapy excluding Radium-223 and anti-PD1/PD-L1/2 therapy |
| **Radiopharmaceutical+Immunotherapy+ARPI** | | | | | | | |
| **NCT04946370** | 1/2 | 225Ac-J591 + Pembrolizumab + ARPI | Pembrolizumab + ARPI | 76/NA | Composite response rate | Recruiting | Progressed on/after ARPI |
| **Radioligand+Immunotherapy+Immunotherapy** | | | | | | | |
| **NCT05150236 (EVOLUTION /ANZUP2001)** | 2 | 177Lu-PSMA + Ipilimumab + Nivolumab | 177Lu-PSMA | 100/93 | 1-yr PSA-PFS | Active,  not recruiting | Progressed on/after ARPI; Prior Docetaxel use permitted |
| **Radioligand-based immunotherapy** | | | | | | | |
| **NCT04876651** | 3 | 177Lu-DOTA-rosopatamb + Standard of care | Standard of care | 392/NA | rPFS | Recruiting | Progressed on/after ARPI or taxane therapy |
| **TKI+Immunotherapy** | | | | | | | |
| **NCT04446117 (CONTACT-02)** | 3 | Cabozantinib + Atezolizumab | Abiraterone /Enzalutamide | 580/507 | OS; rPFS | Active,  not recruiting | Progressed on/after ARPI |
| **NCT05168618 (AtezoCab)** | 2 | Cabozantinib + Atezolizumab | None | 33/NA | 24-wk disease control rate | Recruiting | Progressed on/after ARPI |

Abbreviations: OS: overall survival; PFS: progression-free survival; rPFS: radiographic progression free survival; DLT: dose limiting toxicity; MTD: maximum tolerated dose; RP2D: recommended phase 2 dose; ARPI: androgen receptor pathway inhibitor; mHSPC: metastatic hormone sensitive prostate cancer; mCRPC: metastatic castration resistant prostate cancer; AR: androgen receptor; ARPI: androgen receptor pathway inhibitor; AKT inhibitor: protein kinase B inhibitor; DNA-PK inhibitor: DNA-dependent protein kinase inhibitor; PARPi: poly(ADP-ribose) polymerase inhibitor; TKI: tyrosine kinase inhibitor

**Supplement Methods 1:** PRISMA checklist

| **Section and Topic** | **Item #** | **Checklist item** | **Reported on Page/Figure #** |
| --- | --- | --- | --- |
| **TITLE** | | |  |
| Title | 1 | Identify the report as a literature review. | 1 |
| **ABSTRACT** | | |  |
| Abstract | 2 | Provide a structured summary including, as applicable: background; objectives; data sources; study eligibility criteria, participants, and interventions; study appraisal and synthesis methods; results; limitations; conclusions and implications of key findings.  See the [PRISMA 2020 for Abstracts checklist](http://www.prisma-statement.org/Extensions/Abstracts.aspx) for the complete list. | 3 |
| **INTRODUCTION** | | |  |
| Rationale | 3 | Describe the rationale for the review in the context of existing knowledge, i.e., what is already known about your topic. | 5 |
| Objectives | 4 | Provide an explicit statement of the objective(s) or question(s) the review addresses with reference to participants, interventions, comparisons, outcomes, and study design (PICOS). | 5-6 |
| **METHODS** | | |  |
| Eligibility criteria | 5 | Specify the inclusion and exclusion criteria for the review and how studies were grouped for the syntheses with study characteristics (e.g., PICOS, length of follow-up) and report characteristics (e.g., years considered, language, publication status) used as criteria for eligibility, giving rationale. | 7 |
| Information sources | 6 | Specify all databases, registers, websites, organisations, reference lists and other sources searched or consulted to identify studies. Specify the date when each source was last searched or consulted. | 7 |
| Search strategy | 7 | Present the full search strategies for all databases, registers and websites, including any filters and limits used. | Supplement |
| Selection process | 8 | State the process for selecting studies (i.e., screening, eligibility).  Specify the methods used to decide whether a study met the inclusion criteria of the review, including how many reviewers screened each record and each report retrieved, whether they worked independently, and if applicable, details of automation tools used in the process. | 7 |
| Study risk of bias assessment | 11 | Specify the methods used to assess risk of bias in the included studies, including details of the tool(s) used, how many reviewers assessed each study and whether they worked independently, and if applicable, details of automation tools used in the process. | 7-8 |
| **RESULTS** | | |  |
| Study selection | 16a | Describe the results of the search and selection process, from the number of records identified in the search to the number of studies included in the review, ideally using a flow diagram. | 10, Figure 1 |
|  | 16b | Cite studies that might appear to meet the inclusion criteria, but which were excluded, and explain why they were excluded. | - |
| Study characteristics | 17 | Cite each included study and present its characteristics (e.g., study size, PICOS, follow-up period). | Tables1-6, Supplement |
| Risk of bias in studies | 18 | Present assessments of risk of bias for each included study. | Supplement |
| Results of individual studies | 19 | For all outcomes, present, for each study: (a) summary statistics for each group (where appropriate) and (b) an effect estimate and its precision (e.g. confidence/credible interval), ideally using structured tables or plots. | Tables1-6, Supplement |
| **DISCUSSION** | | |  |
| Discussion | 23a | Provide a general interpretation of the results in the context of other evidence. | 18-22 |
|  | 23b | Discuss any limitations of the evidence included in the review. | 23 |
|  | 23c | Discuss any limitations of the review processes used. | 23 |
|  | 23d | Discuss implications of the results for practice, policy, and future research. | 19-22 |
| **OTHER INFORMATION** | | |  |
| Registration and protocol | 24a | Provide registration information for the review, including register name and registration number, or state that the review was not registered. | - |
|  | 24b | Indicate where the review protocol can be accessed, or state that a protocol was not prepared. | - |
|  | 24c | Describe and explain any amendments to information provided at registration or in the protocol. | - |
| Support | 25 | Describe sources of financial or non-financial support for the review, and the role of the funders or sponsors in the review. | 24 |
| Competing interests | 26 | Declare any competing interests of review authors. | - |
| Availability of data, code, and other materials | 27 | Report which of the following are publicly available and where they can be found: template data collection forms; data extracted from included studies; data used for all analyses; analytic code; any other materials used in the review. | - |

**Supplement Methods 2:** Detailed search strategy

*Search Strategy*

Database(s): Embase 1974 to 2021 June 16, Ovid MEDLINE(R) and Epub Ahead of Print, In-Process, In-Data-Review & Other Non-Indexed Citations and Daily 1946 to June 16, 2021

| # | Searches | Results |
| --- | --- | --- |
| 1 | exp *Prostatic Neoplasms/ | 283331 |
| 2 | exp Clinical Trial/ | 2498364 |
| 3 | exp Meta-Analysis/ | 353101 |
| 4 | 1 and (2 or 3) | 27693 |
| 5 | exp animals/ not exp humans/ | 9641145 |
| 6 | 4 not 5 | 27634 |
| 7 | limit 6 to (letter or editorial or erratum or note or addresses or autobiography or bibliography or biography or blogs or comment or dictionary or directory or interactive tutorial or interview or lectures or legal cases or legislation or news or newspaper article or overall or patient education handout or periodical index or portraits or published erratum or video-audio media or webcasts) [Limit not valid in Embase, Ovid MEDLINE(R), Ovid MEDLINE(R) Daily Update, Ovid MEDLINE(R) PubMed not MEDLINE, Ovid MEDLINE(R) In-Process, Ovid MEDLINE(R) Publisher; records were retained] | 1039 |
| 8 | 6 not 7 | 26595 |

### **Supplement Results:**

Baseline characteristics

A total of 71 trials (50%) reported race and ethnicity and all included 29,883 White/Caucasian patients, 56 (79%) included 1,340 African American/Black patients, 38 (54%) included 2,214 Asian patients, 6 (8%) included 9 Native Hawaiian/Pacific Islander patients, and 11 trials (15%) included 48 American Indian/Alaskan Native patients. Distribution of inclusion by race/ethnicity is provided in **Supplement Tables 3-5.**

Heterogeneity in included trials

*Heterogeneity in reporting of outcomes*

Among phase III/IV trials, OS was reported by 57 (97%) and PFS was reported by 49 (83%) trials; radiographic PFS was reported in 30 (61%) trials, composite PFS in 17 (35%), TTP in only nine (18%), both composite- and radiographic PFS in four (8%), while only one (2%) trial reported both radiographic PFS, and TTP. Outcome reporting matrix is outlined in **Supplement Table 13.**

Among phase II trials, OS was reported by 57 (68%) phase II trials and PFS was reported by 61 (73%) trials; radiographic PFS was reported in 12 (14%), composite PFS in 34 (40%), TTP in 14 (17%), while time to treatment failure was reported in only 1 (1%) trial. Outcome reporting matrix is outlined in **Supplement Table 14.**

*Heterogeneity in reporting of subgroups*

Heterogeneity in reporting of subgroups by different outcomes is outlined in **Supplement Tables 15-16**.
